# Multivariable model of postoperative delirium in cardiac surgery patients: proteomic and demographic contributions

**DOI:** 10.1101/2023.05.30.23289741

**Authors:** MCB Gonçalves, T Khera, HH Otu, S Narayanan, ST Dillon, A Shanker, X Gu, Y Jung, LH Ngo, ER Marcantonio, TA Libermann, B Subramaniam

## Abstract

**Background:** Delirium following cardiac surgery is common, morbid, and costly, but may be prevented with risk stratification and targeted intervention. Preoperative protein signatures may identify patients at increased risk for worse postoperative outcomes, including delirium. In this study, we aimed to identify plasma protein biomarkers and develop a predictive model for postoperative delirium in older patients undergoing cardiac surgery, while also uncovering possible pathophysiological mechanisms.

**Methods:** SOMAscan analysis of 1,305 proteins in the plasma from 57 older adults undergoing cardiac surgery requiring cardiopulmonary bypass was conducted to define delirium-specific protein signatures at baseline (PREOP) and postoperative day 2 (POD2). Selected proteins were validated in 115 patients using the ELLA multiplex immunoassay platform. Proteins were combined with clinical and demographic variables to build multivariable models that estimate the risk of postoperative delirium and bring light to the underlying pathophysiology.

**Results:** A total of 115 and 85 proteins from SOMAscan analyses were found altered in delirious patients at PREOP and POD2, respectively (p<0.05). Using four criteria including associations with surgery, delirium, and biological plausibility, 12 biomarker candidates (Tukey’s fold change (|tFC|)>1.4, Benjamini-Hochberg (BH)-p<0.01) were selected for ELLA multiplex validation. Eight proteins were significantly altered at PREOP, and seven proteins at POD2 (p<0.05), in patients who developed postoperative delirium compared to non-delirious patients. Statistical analyses of model fit resulted in the selection of a combination of age, sex, and three proteins (angiopoietin-2 (ANGPT2); C-C motif chemokine 5 (CCL5); and metalloproteinase inhibitor 1 (TIMP1); AUC=0.829) as the best performing predictive model for delirium at PREOP. The delirium-associated proteins identified as biomarker candidates are involved with inflammation, glial dysfunction, vascularization, and hemostasis, highlighting the multifactorial pathophysiology of delirium.

**Conclusion:** Our study proposes a model of postoperative delirium that includes a combination of older age, female sex, and altered levels of three proteins. Our results support the identification of patients at higher risk of developing postoperative delirium after cardiac surgery and provide insights on the underlying pathophysiology. ClinicalTrials.gov (NCT02546765).

## INTRODUCTION

Delirium, characterized by a sudden and fluctuating decline in attention and cognitive function, affects up to 50% of older adults undergoing cardiac surgery^1^. Postoperative delirium has been linked to both short- and long-term cognitive impairment, persisting for several weeks to as long as 7.5 years after the surgery^2^. Furthermore, delirium is associated with postoperative functional decline and increased mortality^1–4^. It may also prolong ICU and hospital stay, increase institutional discharge, and results in significantly higher cumulative costs of care (with an additional $44,291 per patient per year) compared to non-delirious surgical patients^5^.

The development of delirium is multifactorial. Predisposing factors include age, cognitive impairment, and previous medical history^6^, while precipitating factors encompass surgery-related stressors (e.g. type and duration of surgical procedures, anesthetic medications, intraoperative hypotension, and hypoxia), as well as postoperative pain and intensive care management. To identify and assess the overall risk of neurocognitive complications after surgery, researchers have proposed clinical risk prediction scales and models^7–13^. However, these efforts have often focused on populations other than cardiac surgical patients^14^. Despite significant advances over the past decade^11,15,16^, identifying risk factors specifically for delirium in cardiac surgery patients remains challenging^17^.

The emergence and ongoing development of high throughput “omics” technologies and systems biology offer promising tools for biomarker research, particularly in predicting complex clinical conditions such as delirium^16,18^. In our study, we conducted a comprehensive proteomic profiling and screened biomarker candidates to predict postoperative delirium in surgical cardiac patients. By combining relevant demographic variables with peripheral protein levels obtained from participants of the DexAcet trial^19^, we present a multicomponent model for delirium risk in older patients undergoing cardiac surgery.

## METHODS

### Study design and population

The DexAcet trial, a prospective, randomized, placebo-controlled, triple blinded, factorial design trial, was carried out as a pilot study at a single tertiary care center, from September 2015 through April 2018, with long-term neurocognitive follow-ups completed in April 2019^20^. The study was approved by the Committee on Clinical Investigations Institutional Review Board (CCI-IRB Protocol # 2019 P00075) at Beth Israel Deaconess Medical Center (BIDMC; Boston, MA) and registered at ClinicalTrials.gov (NCT02546765). The details of the protocol and results were previously published^19,21^. Briefly, the DexAcet study enrolled 120 patients aged 60 years and above undergoing cardiac surgery requiring cardiopulmonary bypass (CPB). Patients were randomly assigned into four arms (1:1:1:1) to receive either intravenous (I.V.) acetaminophen along with propofol or dexmedetomidine, or a placebo along with propofol or dexmedetomidine. A sensitivity analysis was performed to evaluate the effect of study drugs on proteomics outcome. All patients approached for participation in the study provided informed consent.

The flow chart for the current study is summarized in **Supplemental Figure S1**.

### Medical record review

The medical record was reviewed daily by trained research staff and was the primary source for patient demographics, details of the intraoperative and postoperative course, and discharge status.

### Delirium and cognitive assessments

Baseline and discharge cognitive assessments were performed in all subjects using the Montreal cognitive assessment (MoCA) as previously described^22^. Delirium was assessed on each postoperative day using a brief cognitive battery to evaluate attention, orientation, and memory, in addition to the Delirium Symptom Interview^23^ to probe for delirium-related symptoms. Delirium diagnosis was determined using the Confusion Assessment Method diagnostic algorithm^24^. For further details, see Subramaniam et al., 2019^19^.

### Biological assessments

Blood samples were collected before surgery (PREOP) and at postoperative day 2 (POD2). Once collected, samples were fractionated into components (red cells, buffy coat, and plasma) by centrifugation at 1100 x *g* for 10 minutes at room temperature. Aliquots of plasma and buffy coat were securely stored in locked –80 °C freezers and later transferred to the lab for further proteomic analyses, ensuring cold chain maintenance. A total of 115 participants out of the 120 enrolled in the DexAcet trial completed the blood work at the two proposed timepoints, and therefore were included in the current proteomic study.

### SOMAscan assay

PREOP and POD2 samples from the subgroup who received I.V. propofol for sedation (57 participants) were selected for the proteomic screening. Plasma samples (50 µL) from the 57 patients, five pooled plasma controls, and one buffer control were analyzed using the SOMAscan Assay Kit for human plasma 1.3k, measuring expression of 1,305 proteins. The assay was performed at the BIDMC Genomics, Proteomics, Bioinformatics, and Systems Biology Center according to the manufacturer’s standard protocol (SomaLogic; Boulder, CO). Several hybridization controls further controlled sample-to-sample variability. Each protein concentration was transformed into a corresponding SOMAmer concentration, then quantified using a custom DNA microarray (Agilent; Santa Clara, CA) read-out, which reports the data as relative fluorescence units (RFU). Data quality control, normalization, and calibration were done according to the manufacturer’s protocol as previously described^25^.

### Selection of biomarker candidates

Twelve proteins were selected as potential biomarkers according to the following criteria:

1. Significant differential protein expression observed from the SOMAscan proteomic scanning, as a result of:

a. Delirium, at either PREOP or POD2, with a defined p-value cut-off of p<0.05 for Delirium vs. Non-delirium (D vs. ND); and
b. Surgery, with Benjamini-Hochberg (BH)-corrected p-value and Tukey’s fold change (tFC) cut-offs for POD2 vs. PREOP defined at BH-p<0.01, |tFC|>1.4; and/or
2. Literature review of proteins for their association with inflammation, cognitive decline, Alzheimer’s disease (AD), delirium, and other perioperative neurocognitive disorders (NCDs);
3. Biological plausibility; and
4. Current availability of antibody pairs to be analyzed by a 4-plex-based ELLA immunoassay.

### ELLA immunoassay

Plasma levels of the twelve biomarker candidates selected from the SOMAscan screening were measured in all 115 subjects of the DexAcet trial^19^ using the fully automated immunoassay platform ELLA (ProteinSimple/Biotechne; San Jose, CA), and custom 4-plex immunoassays optimized for ELLA, following the manufacturer’s protocols. Inter-assay coefficients of variation (CVs) of triplicate measures were generally <5%. If a CV was >10%, the assay was repeated. Assay personnel were blinded to case and control status.

### Systems biology analyses

Functional category, canonical pathway, interactive network, upstream regulators, and regulator effect analyses of all altered proteins associated with delirium at PREOP or POD2 was performed using the Ingenuity Pathway Analysis (IPA) software tool (QIAGEN; Redwood City, CA). We also performed network analysis by applying the Search Tool for the Retrieval of Interacting Genes/Proteins (STRING) database version 11.5.

### Statistical Analyses

#### SOMAscan proteome analysis and identification of biomarker candidates

Paired t-tests were applied to the SOMAscan proteome data to identify proteins significantly differentially expressed between POD2 and PREOP with a BH-corrected paired t-test p-value less than 0.01. FC was calculated as the ratio of the one-step Tukey’s bi-weight average of signal values in the two groups (POD2 and PREOP). Sample and feature clustering was done using principal component analysis (PCA) and hierarchical clustering with average linkage using Pearson’s correlation as the distance metric. Support Vector Machines (SVM) based on PCA results was used for sample classification using linear, polynomial, and Gaussian kernels. The same set of analyses was performed to define the dysregulated proteins in D vs. ND cases at PREOP and POD2. However, due to the subtler differences for delirium and imbalanced sample sets, the threshold for significance was raised to a nominal p-value lower than 0.05.

#### Statistical analysis of ELLA immunoassay data

Normality assumptions of protein concentrations and clinical outcomes were assessed via Shapiro-Wilk test prior to any statistical data comparison. Protein levels (pg/mL) are shown as median and interquartile ranges. Comparisons of mean level of a protein between D vs. ND within each time point were performed either using unpaired t-test or Mann-Whitney according to data distribution. The difference in surgical effect for D vs. ND was assessed by testing the interaction term between delirium status (yes/no) and the protein concentration in a general linear model.

#### Development of a multivariable model

The best model fit took into consideration optimal baseline demographics, clinical traits, and protein biomarkers. First, due to the limited number of delirium cases (21), we reduced the model to 5 predictors to ensure statistical robustness. These predictors included age, sex, and MoCA as baseline factors. Then, we used a subset model selection approach to explore different combinations of up to three proteins alongside the base model. Model performance was assessed using the c-statistic (AUC) to gauge its ability to distinguish delirium from no-delirium cases, and the Akaike Information Criteria (AIC) to measure model goodness-of-fit. Our strategy involved starting with the highest AUC model and subsequently selecting the one with the lowest AIC, addressing multiple testing challenges in model selection.

##### Sensitivity Analysis

To ensure the treatment arms did not influence our selection of the predictive model, we re-ran the above analyses stratified by treatment arm (IV acetaminophen vs. placebo; dexmedetomidine vs. propofol). All analyses were done in SAS 9.4, Cary NC.

## RESULTS

### Study population

The median age of 115 patients in this study, was 69 years (IQR, 63-76), and majority were male (97/115, 84.3%), and white (106/115 [92.2%]). Participants had median baseline MoCA scores of 23 [IQR, 21-26]. Twenty-one patients (18.3%) developed postoperative delirium. Delirium patients were older (median age 75 [IQR, 69-77] vs. 69 [IQR, 63-73] years, respectively; p=0.047) and more likely to be female (38.9% vs. 14.4%, p=0.03) (**Table 1**).

**Table 1.**
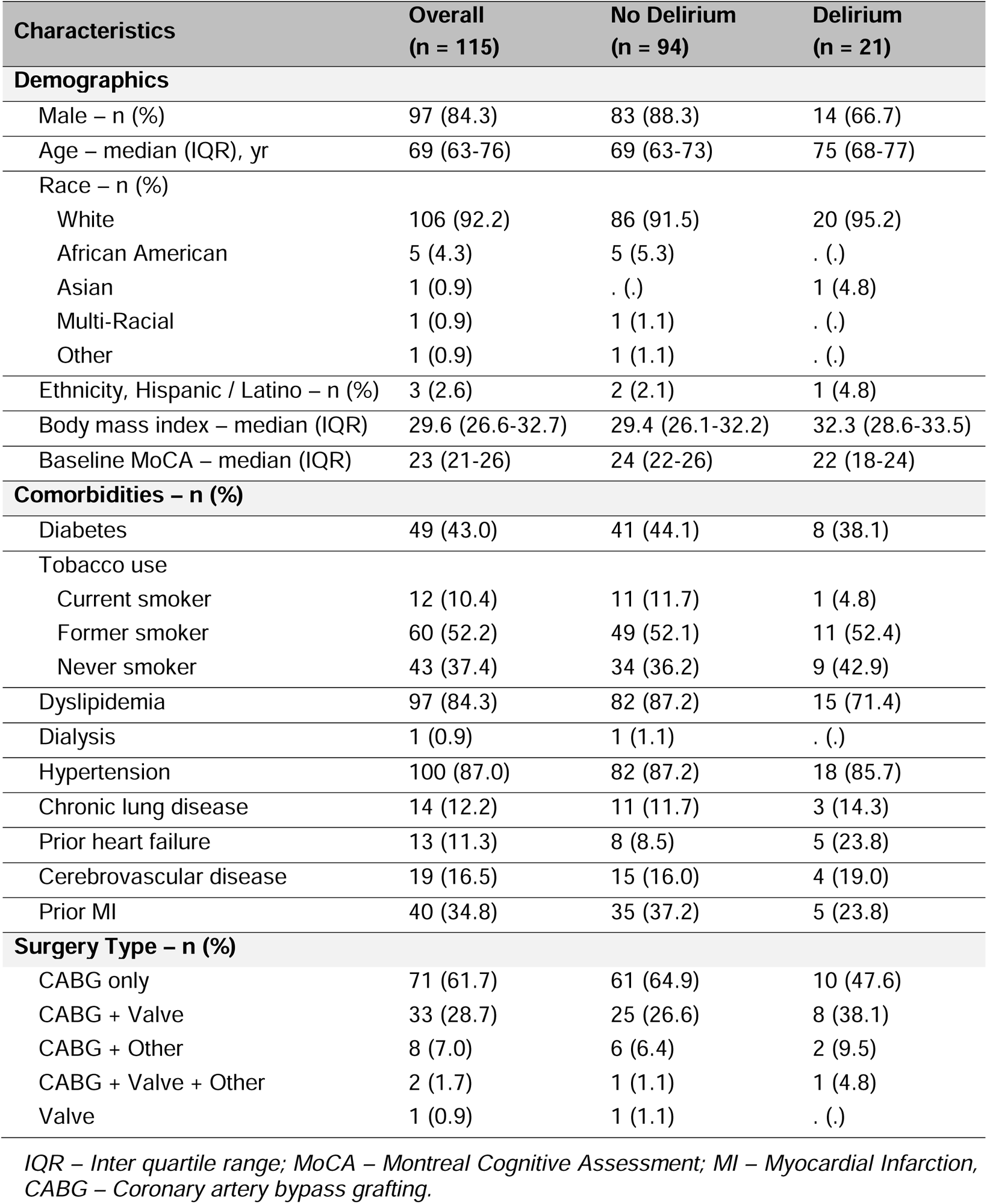
Study population characteristics.

| **Characteristics** | **Overall**<br>**(n = 115)** | **No Delirium**<br>**(n = 94)** | **Delirium**<br>**(n = 21)** |
| --- | --- | --- | --- |
| **Demographics** |  |  |  |
| Male – n (%) | 97 (84.3) | 83 (88.3) | 14 (66.7) |
| Age – median (IQR), yr | 69 (63-76) | 69 (63-73) | 75 (68-77) |
| Race – n (%) |  |  |  |
| White | 106 (92.2) | 86 (91.5) | 20 (95.2) |
| African American | 5 (4.3) | 5 (5.3) | . (.) |
| Asian | 1 (0.9) | . (.) | 1 (4.8) |
| Multi-Racial | 1 (0.9) | 1 (1.1) | . (.) |
| Other | 1 (0.9) | 1 (1.1) | . (.) |
| Ethnicity, Hispanic / Latino – n (%) | 3 (2.6) | 2 (2.1) | 1 (4.8) |
| Body mass index – median (IQR) | 29.6 (26.6-32.7) | 29.4 (26.1-32.2) | 32.3 (28.6-33.5) |
| Baseline MoCA – median (IQR) | 23 (21-26) | 24 (22-26) | 22 (18-24) |
| **Comorbidities – n (%)** |  |  |  |
| Diabetes | 49 (43.0) | 41 (44.1) | 8 (38.1) |
| Tobacco use |  |  |  |
| Current smoker | 12 (10.4) | 11 (11.7) | 1 (4.8) |
| Former smoker | 60 (52.2) | 49 (52.1) | 11 (52.4) |
| Never smoker | 43 (37.4) | 34 (36.2) | 9 (42.9) |
| Dyslipidemia | 97 (84.3) | 82 (87.2) | 15 (71.4) |
| Dialysis | 1 (0.9) | 1 (1.1) | . (.) |
| Hypertension | 100 (87.0) | 82 (87.2) | 18 (85.7) |
| Chronic lung disease | 14 (12.2) | 11 (11.7) | 3 (14.3) |
| Prior heart failure | 13 (11.3) | 8 (8.5) | 5 (23.8) |
| Cerebrovascular disease | 19 (16.5) | 15 (16.0) | 4 (19.0) |
| Prior MI | 40 (34.8) | 35 (37.2) | 5 (23.8) |
| **Surgery Type – n (%)** |  |  |  |
| CABG only | 71 (61.7) | 61 (64.9) | 10 (47.6) |
| CABG + Valve | 33 (28.7) | 25 (26.6) | 8 (38.1) |
| CABG + Other | 8 (7.0) | 6 (6.4) | 2 (9.5) |
| CABG + Valve + Other | 2 (1.7) | 1 (1.1) | 1 (4.8) |
| Valve | 1 (0.9) | 1 (1.1) | . (.) |

*IQR – Inter quartile range; MoCA – Montreal Cognitive Assessment; MI – Myocardial Infarction, CABG – Coronary artery bypass grafting.*

### SOMAscan-proteomic profiling

SOMAscan analysis was performed on plasma samples of 57 participants, of which 12 (21.05%) experienced delirium (**Supplemental Table S1**). Our analysis of 1,305 plasma proteins revealed significant changes in 666 analytes following cardiac surgery at POD2 compared to PREOP (BH-p<0.01) (**Supplemental Table S2**). Plasma from PREOP and POD2 was clearly distinguished by PCA-SVM and hierarchical clustering (**Figure 1A and 1B**). Differential expression of 115 and 85 proteins was observed at PREOP and POD2 respectively comparing delirious and non-delirious individuals (p<0.05) (**Supplemental table S3**); PCA-SVM analysis separated the two groups along principal components 1 and 2 at both timepoints (**Figure 1C and 1D**).

**Figure 1.**
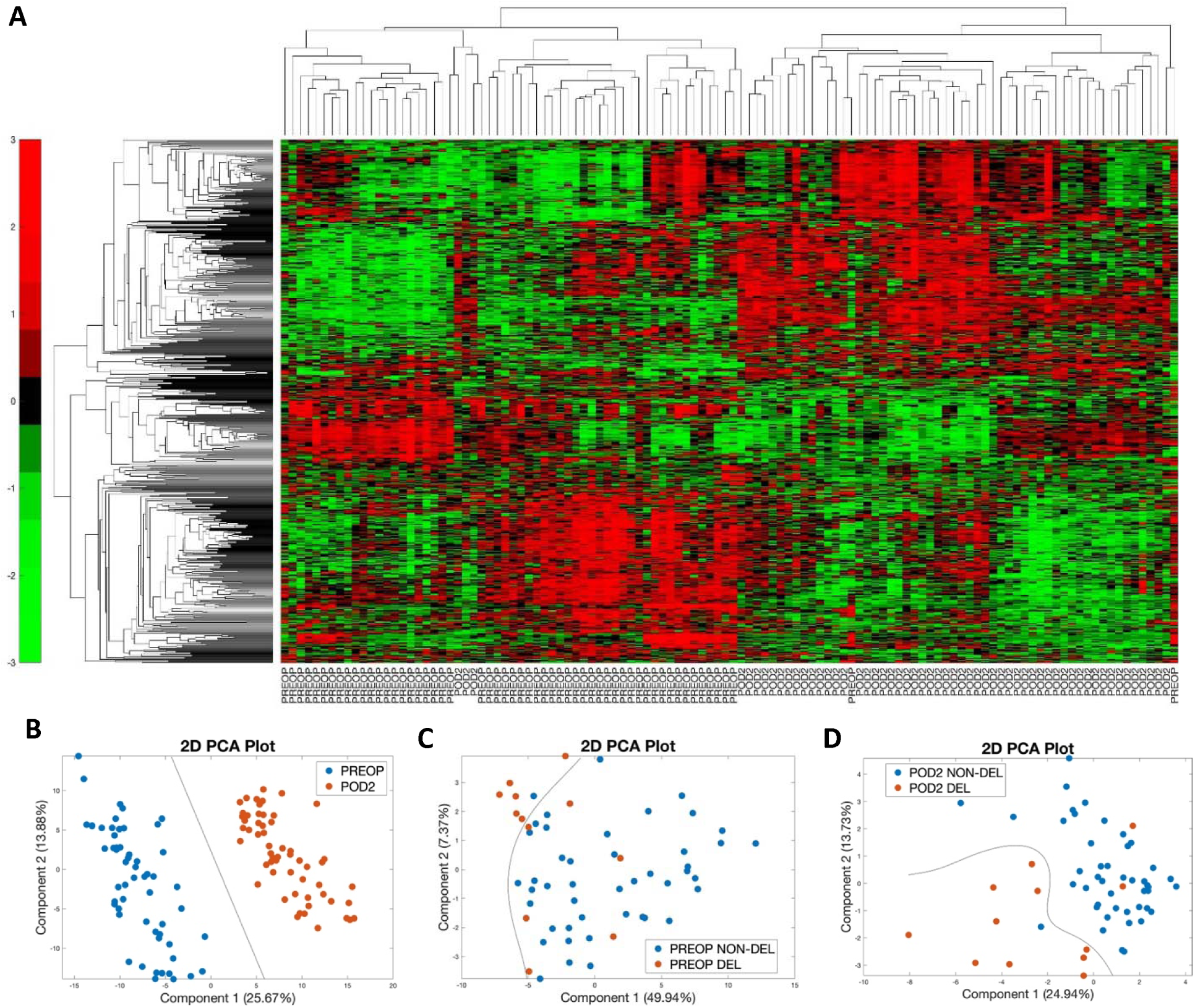
Differential and qualitative analyses of SOMAscan-based global proteome screening of 57 cardiac surgery patients requiring cardiopulmonary bypass. **(A)** The heatmap for unsupervised hierarchical clustering of 666 differentially expressed proteins among patients shows differences between POD2 and PREOP (BH-p<0.01). **(B)** Principal component analysis of all 57 patients based on the 666 differentially expressed proteins projected in the 2D plot shows a clear segregation of features. Component 1 splits the samples by surgery states (POD2 vs. PREOP). (**C**) Principal component analysis of all 57 patients based on the 115 differentially expressed proteins at PREOP projected in the 2D plot shows a clear segregation of features. Component 1 splits the samples by delirium effect (DEL vs NON-DEL). (**D**) Principal component analysis of all 57 patients based on the 85 differentially expressed proteins at POD2 projected in the 2D plot shows a clear segregation of features. Component 1 splits the samples by delirium effect (DEL vs NON-DEL).

### Selection of delirium biomarker candidates

From the 666 differentially expressed surgery-associated analytes (BH-p<0.01), 225 had a |tFC|>1.4 (**Supplemental Table S2**). Based on the specific criteria outlined in Methods, 11 final targets were selected as delirium biomarker candidates: platelet-derived growth factor subunit B (PDGFB), tumor necrosis factor receptor superfamily member 1A (TNFRSF1A/TNF-sR1A/TNFR1/CD120a), interleukin-6 (IL6), C-C motif chemokine 5 (CCL5/RANTES), insulin-like growth factor-binding protein 2 (IGFBP2), metalloproteinase inhibitor 1 (TIMP1), angiopoietin-2 (ANGPT2), chitinase-3-like protein 1 (CHI3L1/YKL-40), hepcidin (HAMP), thrombospondin-1 (THBS1), and lipocalin-2 (LCN2). Additionally, neurofilament light chain (NfL) (not screened by SOMAscan) was selected for being a known marker of neuronal injury with reported association to delirium^26^. These 12 proteins are overall moderately correlated at both PREOP and POD2 (**Supplemental Table S4a** and **S4b**, respectively).

### Immunovalidation of protein levels and the association with delirium

The biomarker candidates were validated by orthogonal immunoassay ELLA in all 115 patients of the initial cohort. All 12 proteins were significantly dysregulated (p<0.01) after surgery (9 proteins increased, 3 proteins decreased at POD2 vs PREOP), correlating well with the SOMAscan data. Eight of the twelve ELLA-measured proteins were significantly associated with delirium at PREOP: increased IGFBP2, ANGPT2, IL6, TIMP1, CHI3L1, NFL, TNFRSF1A; decreased PDGFB (D vs. ND; p<0.05; **Figure 2**). Similarly, seven proteins were associated with delirium at POD2: increased IGFBP2, LCN2, IL6, TIMP1, NFL, TNFRSF1A; decreased CCL5 (D vs. ND; p<0.05; POD2; **Figure 2**). No statistically significant differences were observed at PREOP or at POD2 for HAMP (p=0.842; p=0.115) and THBS1 (p=0.126; p=0.066) protein levels between Delirium vs. Non-delirium groups. Our sensitivity analysis stratifying by the treatment arms of the DexAcet trial demonstrated overall minimal effect of drug treatment on protein levels (data not shown). Analyses of changes in protein concentrations between PREOP and POD2 stratified by delirium status (D vs ND) (**Figure 2**) showed greater increase in LCN2 (p=0.01) and TNFRSF1A (p=0.02), and lower increase in HAMP (p=0.03). Overall, the magnitude of surgical changes was similar for all other biomarker candidates in both Delirium and Non-delirium groups (DiD; ΔΔ Perioperative, **Figure 2**).

**Figure 2.**
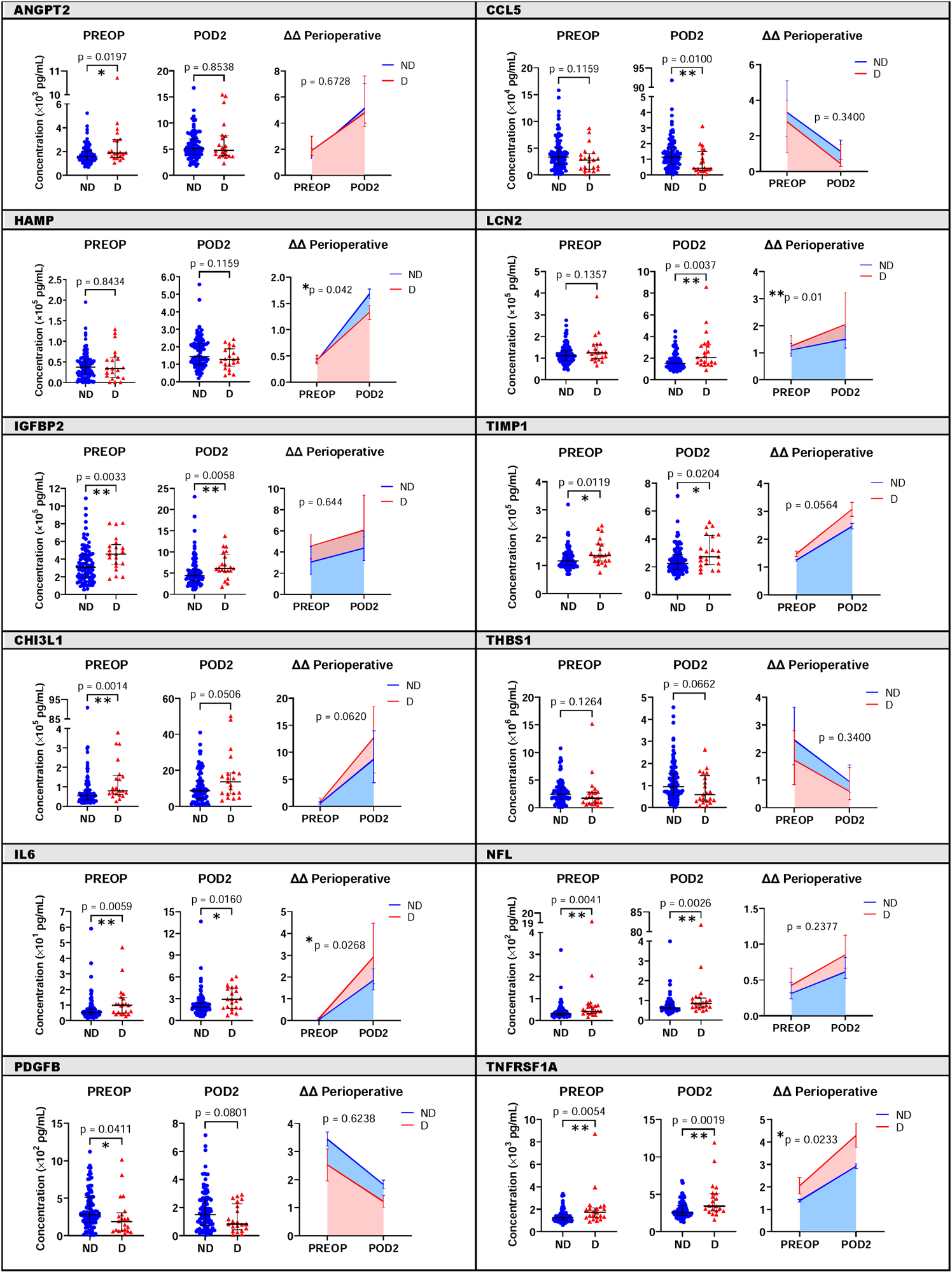
Quantitative validation of biomarker candidates by ELLA 4-multiplex immunoassay according to delirium and surgery status, and the difference in changes in protein levels between delirious and non-delirious patients during the perioperative period. Protein levels (pg/mL) of the twelve selected biomarker candidates were analyzed according to delirium experience among all the 115 patients undergoing cardiac surgery. Scatter plots represent median and interquartile range of plasma protein concentration; p-values correspond to the difference of protein levels between delirium and no delirium outcomes at each timepoint separately (PREOP and POD2). Difference-in-differences (ΔΔ) regression analysis represents the relative protein changes during the perioperative period and are represented by the shading plots; p-values correspond to the difference between protein levels variation during surgery according to delirium status. ANGPT2 = angiopoietin-2; CCL5 = C-C motif chemokine 5; CHI3L1 = chitinase-3-like protein 1; HAMP = hepcidin; IGFBP2 = insulin-like growth factor-binding protein 2; IL6 = interleukin-6; LCN2 = lipocalin 2; NFL = neurofilament light chain; PDGFB = platelet-derived growth factor subunit B; POD2 = postoperative day 2 timepoint; PREOP = preoperative baseline timepoint; THBS1 = thrombospondin-1; TIMP1 = metalloproteinase inhibitor 1; TNFRSF1A = tumor necrosis factor receptor superfamily member 1A. (*p < 0.05; **p < 0.01).

### Development of a statistical predictive model

Multivariable analyses to determine a predictive risk model were performed by including demographic characteristics and up to five proteins proven dysregulated due to surgery and delirium development. The best PREOP model based on AIC and AUC was achieved when combining age, sex, and preoperative altered levels of TIMP1, ANGPT2, and CCL5 (AIC=101.68, AUC=0.829; **Table 2a**). Characteristics of the tested multivariable models linked to delirium risk are summarized in **Table 2b**.

**Table 2a.** Multivariable prediction models of delirium including demographic features combined with a group of preoperatively dysregulated proteins.

| Predictors | AIC | AUC |
| --- | --- | --- |
| **Age, Sex** | 107.62 | .709 |
| TIMP1 ANGPT2 NFL PDGFB | 101.15 | .797 |
| age sex ANGPT2 NFL TNFRSF1A | 100.42 | .797 |
| **age sex TIMP1 ANGPT2 CCL5** | **101.68** | **.829** |
| age sex TIMP1 CCL5 NFL | 101.82 | .814 |
| age sex ANGPT2 IL6 NFL | 101.93 | .800 |
| age sex TIMP1 NFL PDGFB | 102.25 | .825 |
| age sex TIMP1 CCL5 | 102.61 | .807 |
| age sex TIMP1 CCL5 IL6 | 102.87 | .804 |
| age sex TIMP1 ANGPT2 PDGFB | 103.03 | .824 |
| age sex TIMP1 IL6 PDGFB | 103.52 | .820 |
| age sex TIMP1 PDGFB | 103.80 | .820 |
| age sex IGFBP2 TIMP1 CCL5 | 103.98 | .822 |
| age sex IGFBP2 TIMP1 PDGFB | 105.34 | .832 |
| age sex TIMP1 PDGFB TNFRSF1A | 105.49 | .824 |

**Table 2b.** Summary of results for the best prediction model.

| Predictor | Unit for Odds Ratio | Odds Ratio | 95% CI |
| --- | --- | --- | --- |
| Age | 1 year | 1.04 | 0.97 - 1.11 |
| Sex | Female | 3.36 | 0.97 - 11.66 |
| TIMP1 | 1 SD | 1.63 | 1.00 - 2.66 |
| ANGPT2 | 1 SD | 1.57 | 0.86 - 2.88 |
| CCL5 | 1 SD | 0.54 | 0.27 - 1.05 |

ANGPT2; angiopoietin-2; CCL5: C-C motif chemokine 5; TIMP1: metalloproteinase inhibitor 1.

### Systems Biology Analyses

To investigate the systemic functional biology linked to the observed dysregulation on proteins, the IPA analyses revealed that activation of the immune system and inflammation were the main pathways associated with delirium proteins both pre- and postoperatively **(Supplemental Figure S2)** Besides inflammation, vascularization and angiogenesis were identified as significant biological functions linked to the delirium-associated proteins at PREOP and POD2, respectively (**Supplemental Figure S3**). Possible upstream regulators that strongly connect the proteins included in our multivariable predictive model are angiotensinogen (AGT) for dysregulated PREOP proteins and hepatocyte growth factor (HGF) for dysregulated POD2 proteins (**Supplemental Figure S4**). Network and cluster analysis using STRING resulted in 58 of the 115 PREOP delirium-associated proteins forming distinct interacting protein clusters enriched in pathways associated with inflammation, chemokine signaling, extracellular matrix, vascular function, ephrin signaling, TGFβ signaling, adipokines, and fatty acid transport (**Figure 3**). TIMP1, CCL5, and TNFRSF1A occupy key focus hubs in this analysis.

**Figure 3.**
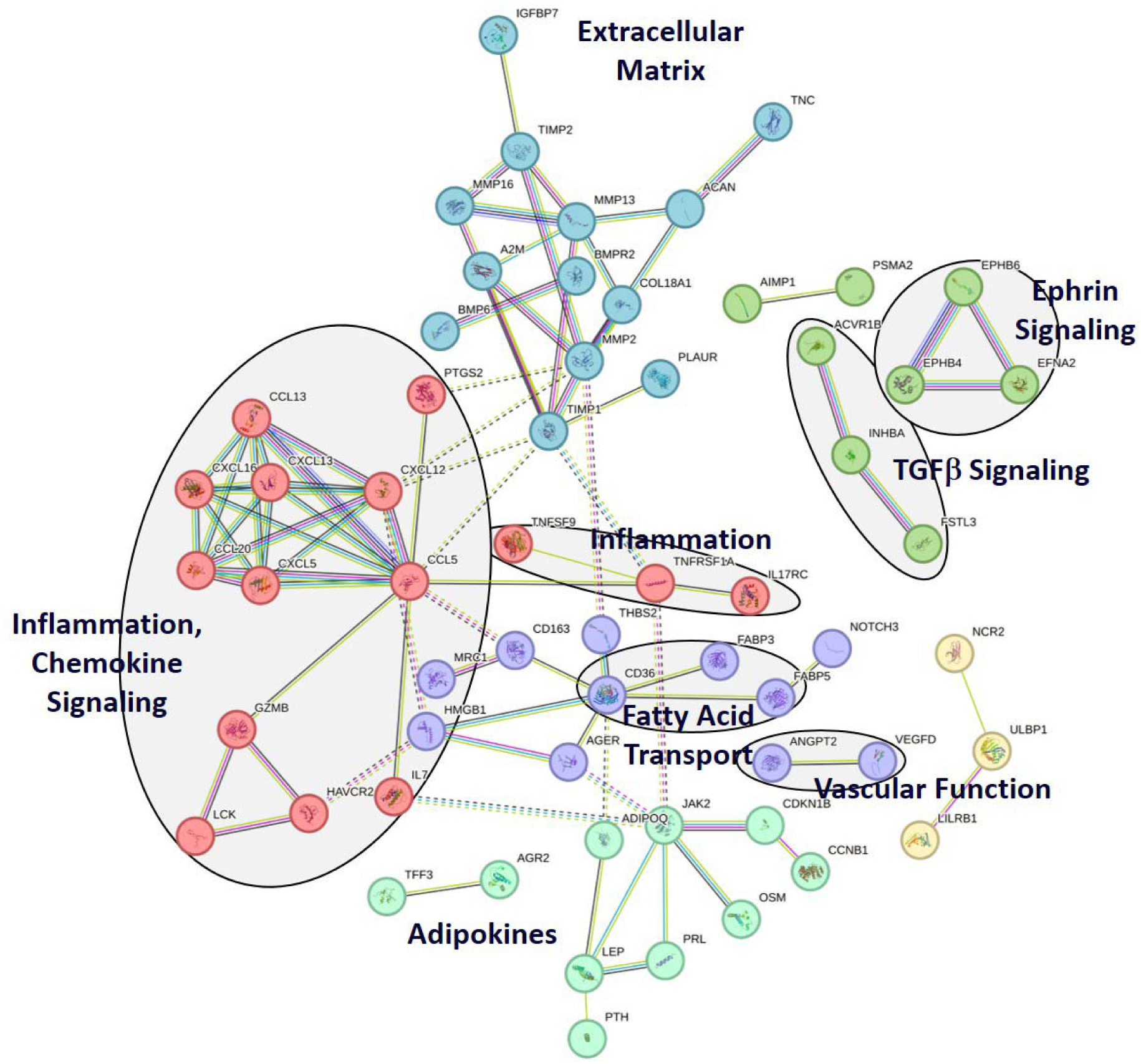
STRING analysis of the 115 significantly altered PREOP proteins. Functional pathways identified using the STRING analysis program. Eight functional groups are identified: inflammation and chemokine signaling, extracellular matrix, inflammation, ephrin signaling, TGFβ signaling, adipokines, vascular function, and fatty acid transport.

## DISCUSSION

In this study, we performed a high throughput semi-targeted proteomics screening using the SOMAscan platform to identify potential plasma biomarkers for postoperative delirium in patients undergoing cardiac surgery. Applying orthogonal immunoassays to further validate the aptamer-based proteomics for delirium biomarker candidates, we derived a 5-variable logistic regression model including age, sex, and the preoperative level of 3 proteins to predict postoperative delirium.

From the demographic perspective, we found that age is associated with increased risk of delirium, as previously shown^27^. However, even though males represent the larger portion of patients undergoing cardiac surgery, our data indicate that females are more likely to experience delirium, differing from the current literature reporting male sex as an inconsistent risk factor for delirium in both cardiac and non-cardiac surgeries^28–30^. Here we highlight that the coronary bypass procedure is often technically more challenging in females compared to males. Thus, while females get bypass surgery less frequently, the surgery itself is considered more difficult, which might lead to more delirium due to an increased exposure to precipitating factors such as prolonged pump time and number/diameter of vessels bypassed.

Our analyses show that about half of the 1305 screened proteins on SOMAScan are significantly dysregulated by surgery. This corroborates recent findings regarding the impact of surgical trauma on specific protein targets linked to neurocognitive impairment, brain injury and delirium^31–34^ in both cardiac and non-cardiac patients^35^. The systemic inflammation resulting from the surgical trauma is believed to induce neuroinflammation and affect brain function and cognition, especially in older and frail patients^36^. In this context, neuroinflammation is evidenced by postoperative complement system activation, induction of the coagulation cascade, increased blood-brain barrier (BBB) permeability followed by the infiltration of immune cells, and activation of glial cells^36^.

Although the search for protein biomarkers of delirium is growing, there is no consensus around one single biomarker. This indicates that a panel of proteins would be of better power and relevance on discriminating delirious from non-delirious patients^16^. Tau protein, NfL and more frequently IL-6 have been proposed as strong biomarker candidates due to their well-known association with cognitive decline and the shared characteristics between perioperative disorders, neurodegenerative diseases, and neuronal injury^26^. The stress-immune hypothesis for delirium has also been supported by evidence showing increased levels of cortisol, CRP, TNF-α, IL-2, IL-6, and IL-8 in delirious surgical patients^32,34,37–42^.

Here we show that a novel group of proteins identified by our multivariable predictive model (TIMP1, ANGPT2 and CCL5) point to altered immune function and inflammation response as well as dysregulation in neovascularization (especially angiogenesis), coagulation, and overall hemostasis as risk factors for delirium development. Interestingly, previously reported biomarker candidates reviewed by Wiredu et al. (2023) and recently reported by Dillon et al. (2023) modulate similar pathways and biological functions, being immune regulation and hemostasis within the most common biological functions associated with postoperative delirium^16,43^. Thus, our data encourage that identifying shared biological functions and pathways linked to delirium might be a useful strategy to identify biomarkers and therapeutic targets.

## STRENGHTS AND LIMITATIONS

It is important to note a few limitations of our study. The DexAcet trial was a multifactorial randomized controlled trial focusing on the protective effect of acetaminophen on postoperative delirium, thus not a pure observational study. Patients who received acetaminophen may not have developed delirium after surgery. Our sensitivity analysis found no significant variations in protein levels across the randomized arms of the original trial, although the low incidence of delirium in our study population could have been related to the interventions administered, and our current results should be verified in observational studies.

The SOMAscan proteomics screened all participants (n=57) who were administered propofol as the sedative. It is common practice to observe BH-corrected values in proteomic data analyses. However, the low incidence of delirium in our study resulted in an imbalanced dataset, hence the observed effect size based on delirium outcome is subtle and BH-adjusted significance is not apparent. To circumvent this limitation, we used a nominal p-value cutoff approach to analyze proteins that differ based on the outcome of delirium. Orthogonal immunovalidation applied to the entire cohort was chosen to prevent the incorporation of false-positive results. In addition, our predictive model includes individual biomarkers that are not statistically significant, which may be due to our limited sample size. Additional independent validation in cohorts with larger and more diverse populations is required and encouraged.

The most recent SOMAscan version 4.1, which measures 7,000 proteins (compared to version 3.2 used in our analysis that measured 1305 proteins), could identify other delirium-associated markers missed by our current approach. Technical constraints limited our selection of twelve biomarker candidates to proteins that could be confirmed by the ELLA platform. Thus, additional comprehensive and detailed pathway analysis and immunoassay validation could improve our delirium predictive model and provide additional pathophysiological insights. Thus, **Table 2** data may be of interest for further studies.

## CONCLUSION

Surgeries cause systemic reactions that translate to the brain, resulting in delirium. Our study shows that a combination of older age, female sex and altered preoperative levels of three proteins stratify patients at higher risk of developing delirium after cardiac surgery. If verified, this prediction model could lead to targeted delirium risk reduction measures for cardiac surgery patients with a specific focus on regulating immune response, hemostasis and vascularization.

## Data Availability

All data produced in the present study are available upon reasonable request to the authors.

## Acknowledgments

The authors acknowledge all clinical and research staff that contributed to the study procedures.

