## Supplemental material for "Multivariable model of postoperative delirium in cardiac surgery patients: proteomic and demographic contributions"

**SUPPLEMENTAL MATERIAL FOR PUBLICATION**

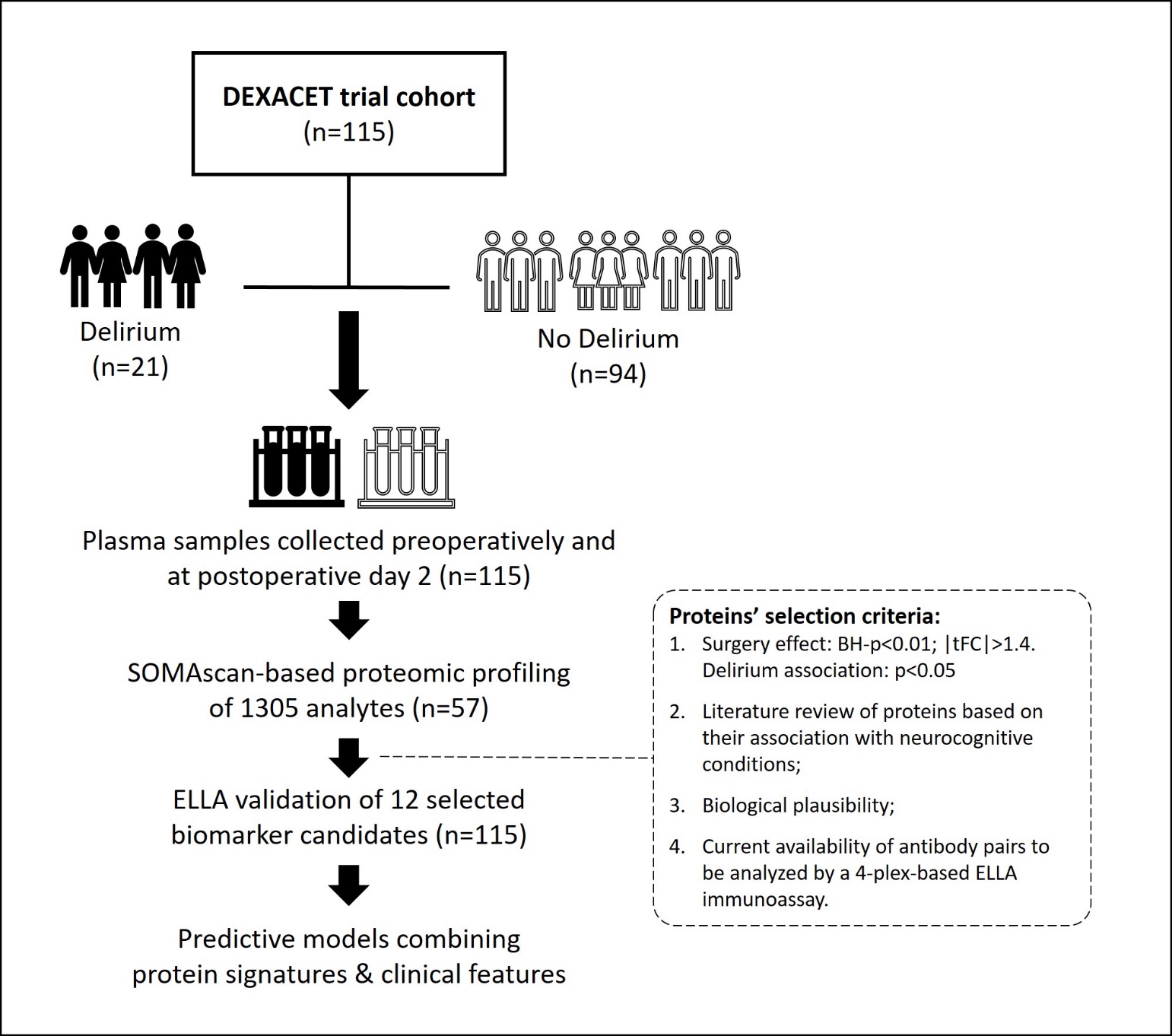

**Figure S1. Study flowchart.** A total of 115 patients of the DexAcet clinical trial were included in this study to identify protein signatures for postoperative delirium in patients undergoing cardiac surgery requiring cardiopulmonary bypass. BH: Benjamini-Hochberg; tFC: Fold-change of Tukey’s biweight robust averages.

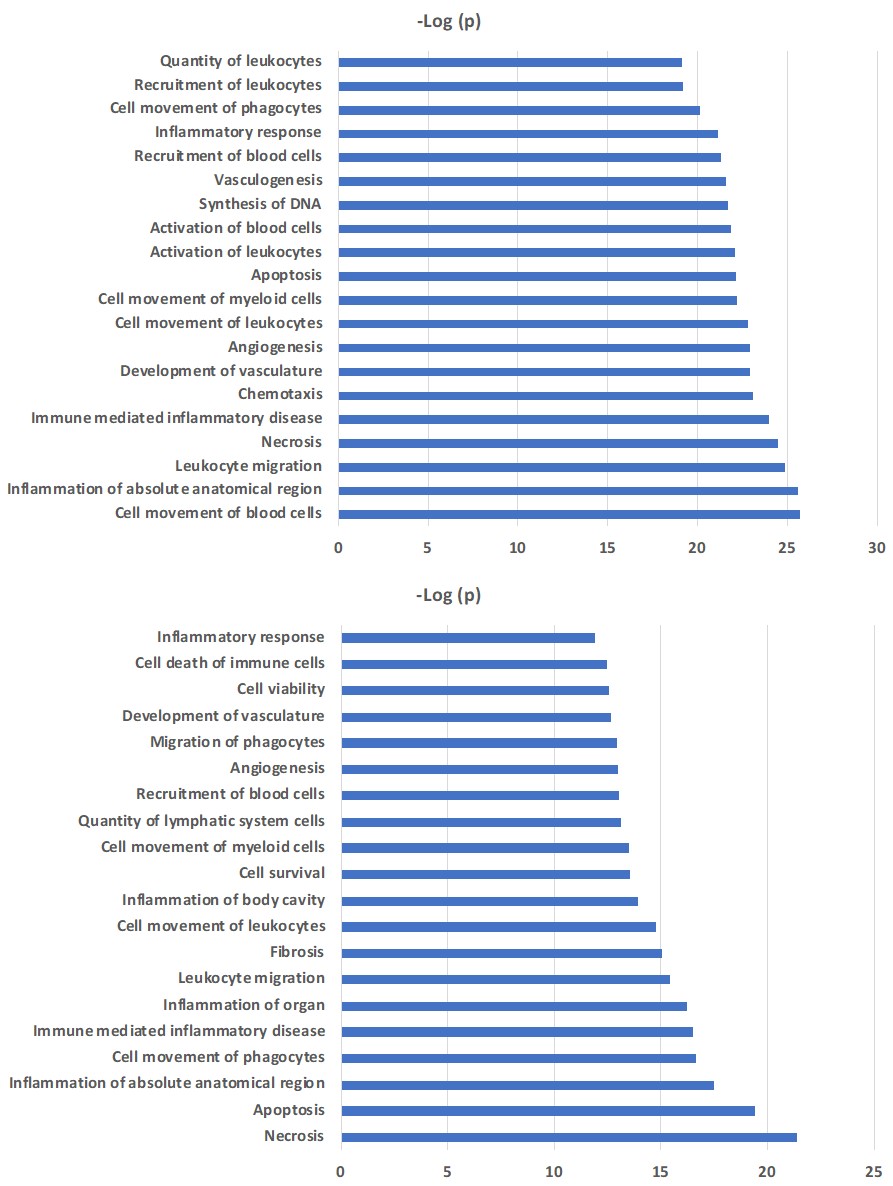

**A**

**B**

**Figure S2. Functional pathways of delirium-associated proteins.** Biological functional pathways that are significantly enriched by the 115 and 85 delirium-associated proteins (**A**) preoperatively and (**B**) at postoperative day 2, respectively, were determined using Ingenuity Pathway Analysis (IPA). Level of significance: p < 10^−5^.

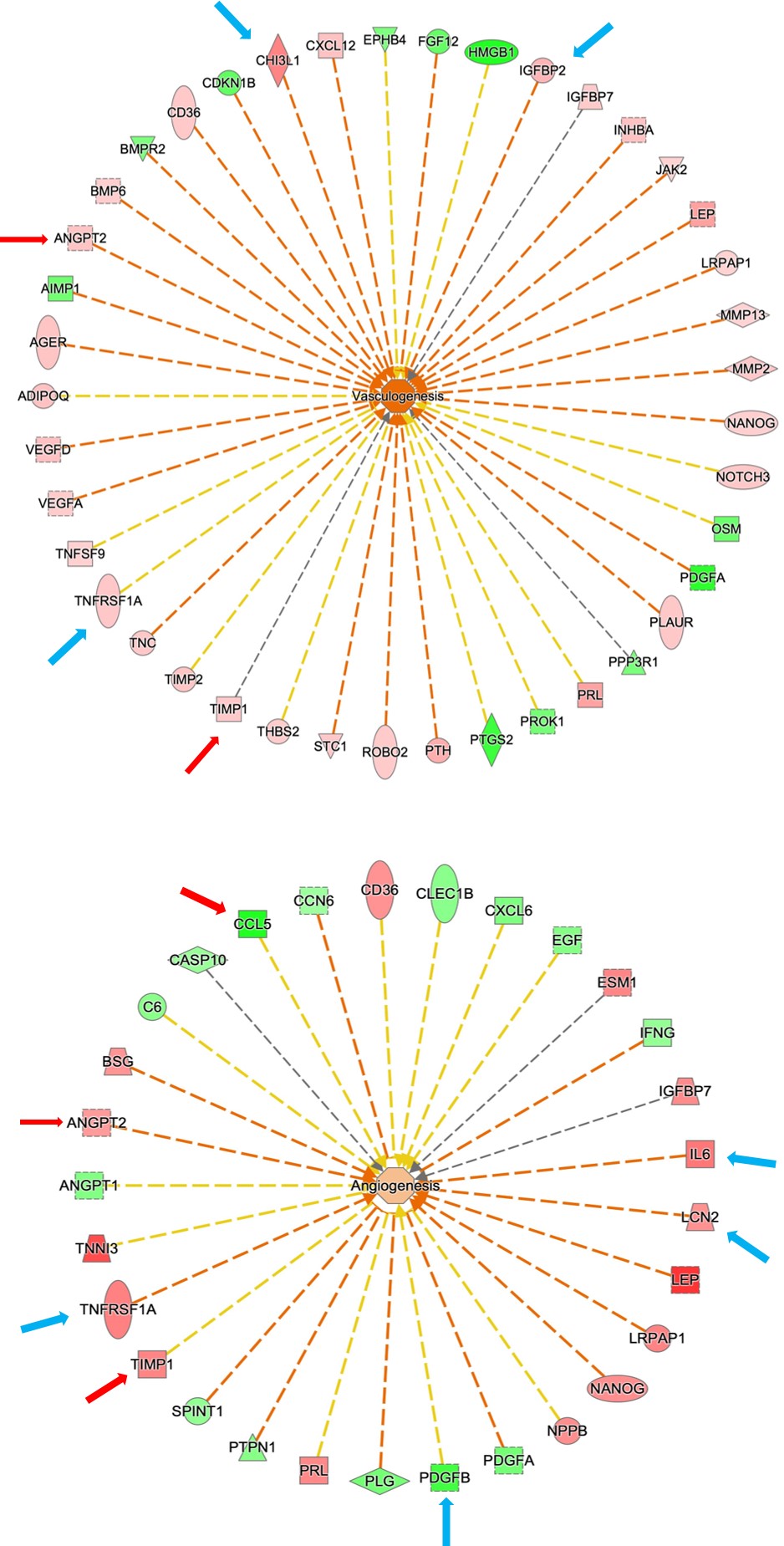

**B**

**A**

**Figure S3. Biological pathways of delirium-associated proteins secondary to inflammation.** IPA systems biology analyses of relevant pathways secondary to inflammation show enrichment of (**A**) the vascularization pathway resulting from the 115 delirium-associated proteins dysregulated preoperatively, and (**B**) the angiogenesis pathway resulting from the 85 delirium-associated proteins dysregulated at postoperative day 2. Blue arrows highlight proteins within the 12 selected biomarker candidates; red arrows highlight the biomarker candidates that compose the final multivariable predictive model. Level of significance: p < 10^−5^.

**A**

**B**

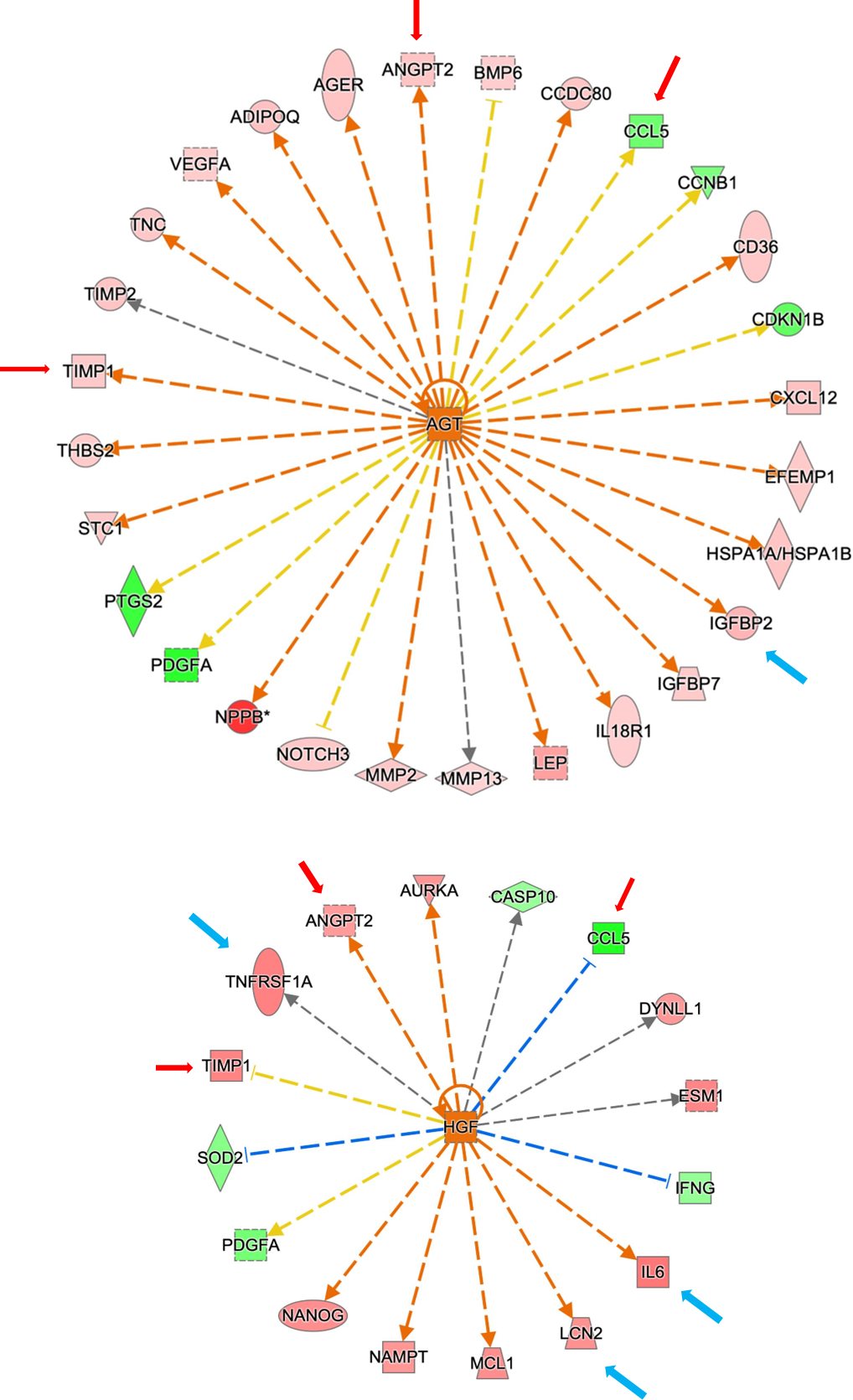

**Figure S4. Upstream regulators of delirium-associated proteins.** IPA systems biology analyses of upstream regulators of functional pathways resulting from (**A**) the 115 delirium-associated proteins dysregulated at preoperative stage, and (**B**) the 85 delirium-associated proteins dysregulated at postoperative day 2. Blue arrows highlight proteins within the 12 selected biomarker candidates; red arrows highlight the biomarker candidates that compose the final multivariable predictive model. Level of significance: p < 10^−5^.

**Table S1.** Overall study population characteristics compared to the propofol subcohort.

| **Characteristics** | **Overall**  **(n = 115)** | | **Propofol subcohort**  **(n = 57)** | ***p*-value (X^2^)** |
| --- | --- | --- | --- | --- |
| **Demographics** | | | |  |
| Male – n (%) | 97 (84.3) | 47 (82.45) | | 0.75 |
| Age – median (IQR), yr | 69 (63-76) | 71 (65-77) | | 0.22 |
| Race – n (%) |  | | |  |
| White | 106 (92.2) | | 53 (92.98) | 0.85 |
| African American | 5 (4.3) | | 2 (3.51) |  |
| Asian | 1 (0.9) | | 1 (1.75) |  |
| Multi-Racial | 1 (0.9) | | 1 (1.75) |  |
| Other | 1 (0.9) | | 0 (.) |  |
| Ethnicity, Hispanic / Latino – n (%) | 3 (2.6) | | 2 (3.51) | 0.74 |
| Body mass index – median (IQR) | 29.6 (26.6-32.7) | | 29.48 (26.48-33.09) | 0.91 |
| Baseline MoCA – median (IQR) | 23 (21-26) | | 24 (21-26) | 0.97 |
| **Comorbidities – n (%)** | | | |  |
| Diabetes | 49 (43.0) | | 21 (36.84) | 0.44 |
| Tobacco use |  | | | ***0.002** |
| Current smoker | 12 (10.4) | | 4 (7.01) |  |
| Former smoker | 60 (52.2) | | 34 (59.64) |  |
| Never smoker | 43 (37.4) | | 16 (28.07) |  |
| Dyslipidemia | 97 (84.3) | | 50 (87.71) | 0.54 |
| Dialysis | 1 (0.9) | | 0 (.) | 0.32 |
| Hypertension | 100 (87.0) | | 49 (85.96) | 0.86 |
| Chronic lung disease | 14 (12.2) | | 0 (.) | 0.60 |
| Prior heart failure | 13 (11.3) | | 6 (10.52) | 0.87 |
| Cerebrovascular disease | 19 (16.5) | | 11 (19.29) | 0.66 |
| Prior MI | 40 (34.8) | | 19 (33.33) | 0.85 |
| **Surgery Type – n (%)** | | | |  |
| CABG only | 71 (61.7) | | 38 (66.66) |  |
| CABG + Valve | 33 (28.7) | | 18 (31.57) |  |
| CABG + Other | 8 (7.0) | | 1 (1.75) | 0.74 |
| CABG + Valve + Other | 2 (1.7) | | 0 (.) |  |
| Valve | 1 (0.9) | | 0 (.) |  |
| **Postoperative delirium (%)** | **21 (18.3)** | | **12 (21.1)** | **0.67** |

*IQR – Inter quartile range, MoCA - Montreal Cognitive Assessment, MI- Myocardial Infarction, CABG - Coronary artery bypass graft.*

**Table S2.** Differential levels of the SOMAscan-screened proteins according to surgery effect.

| **SomaID** | **TargetFullName** | **Target** | **UniProt** | **EntrezGeneID** | **EntrezGeneSymbol** | **tFC** | **BH-p-val** |
| --- | --- | --- | --- | --- | --- | --- | --- |
| SL021043 | Growth/differentiation factor 11/8 | GDF-11/8 | O95390 O14793 | NaN | GDF11 MSTN | -1.419967 | 0 |
| SL019979 | Histone H2A type 3 | H2A3 | Q7L7L0 | NaN | HIST3H2A | 2.035869 | 0 |
| SL018548 | Alpha-1-antichymotrypsin complex | alpha-1-antichymotrypsin complex | P01011 | NaN | SERPINA3 | 1.799866 | 0 |
| SL018509 | R-spondin-3 | RSPO3 | Q9BXY4 | 84870 | RSPO3 | -1.515569 | 0 |
| SL017189 | Kininogen-1 | Kininogen, HMW | P01042 | 3827 | KNG1 | -1.448538 | 0 |
| SL016148 | gp41 C34 peptide, HIV | C34 gp41 HIV Fragment | Q70626 | NaN | Human-virus | 1.673027 | 0 |
| SL014983 | Histone H2B type 2-E | H2B2E | Q16778 | 8349 | HIST2H2BE | 3.036864 | 0 |
| SL014896 | Ankyrin-2 | ANK2 | Q01484 | 287 | ANK2 | 3.360158 | 0 |
| SL014248 | Semaphorin-6B | SEM6B | Q9H3T3 | 10501 | SEMA6B | 1.674373 | 0 |
| SL014092 | Cell adhesion molecule-related/down-regulated by oncogenes | CDON | Q4KMG0 | 50937 | CDON | -1.493299 | 0 |
| SL014008 | Alpha-(1,3)-fucosyltransferase 5 | FUT5 | Q11128 | 2527 | FUT5 | 1.562049 | 0 |
| SL013165 | Non-histone chromosomal protein HMG-14 | HMGN1 | P05114 | 3150 | HMGN1 | -1.50423 | 0 |
| SL012783 | WAP, kazal, immunoglobulin, kunitz and NTR domain-containing protein 1 | WFKN1 | Q96NZ8 | 117166 | WFIKKN1 | -1.422629 | 0 |
| SL012774 | Cysteine-rich with EGF-like domain protein 1 | CREL1 | Q96HD1 | 78987 | CRELD1 | 1.975763 | 0 |
| SL012707 | Proprotein convertase subtilisin/kexin type 9 | PCSK9 | Q8NBP7 | 255738 | PCSK9 | 1.657145 | 0 |
| SL010830 | Protein 4.1 | 41 | P11171 | 2035 | EPB41 | -1.644309 | 0 |
| SL010612 | Dickkopf-related protein 4 | Dkk-4 | Q9UBT3 | 27121 | DKK4 | -1.488827 | 0 |
| SL010328 | Mediator of RNA polymerase II transcription subunit 1 | MED-1 | Q15648 | 5469 | MED1 | -3.072783 | 0 |
| SL010288 | Carbonic anhydrase 6 | Carbonic anhydrase 6 | P23280 | 765 | CA6 | -1.813901 | 0 |
| SL009324 | Follistatin-related protein 3 | FSTL3 | O95633 | 10272 | FSTL3 | 1.520681 | 0 |
| SL009045 | Ectonucleotide pyrophosphatase/phosphodiesterase family member 7 | ENPP7 | Q6UWV6 | 339221 | ENPP7 | -1.782537 | 0 |
| SL008936 | Neural cell adhesion molecule L1-like protein | CHL1 | O00533 | 10752 | CHL1 | -1.468194 | 0 |
| SL008931 | CD177 antigen | CD177 | Q8N6Q3 | 57126 | CD177 | 1.738966 | 0 |
| SL008835 | Asialoglycoprotein receptor 1 | ASGR1 | P07306 | 432 | ASGR1 | 2.730012 | 0 |
| SL008822 | Adhesion G protein-coupled receptor E2 | EMR2 | Q9UHX3 | 30817 | ADGRE2 | -1.437787 | 0 |
| SL008611 | N-acylethanolamine-hydrolyzing acid amidase | ASAHL | Q02083 | 27163 | NAAA | -1.967386 | 0 |
| SL008381 | Cathepsin F | CATF | Q9UBX1 | 8722 | CTSF | -2.010421 | 0 |
| SL008380 | Cathepsin Z | CATZ | Q9UBR2 | 1522 | CTSZ | 1.600572 | 0 |
| SL008372 | Leucine-rich repeat transmembrane protein FLRT3 | FLRT3 | Q9NZU0 | 23767 | FLRT3 | 1.499738 | 0 |
| SL008193 | Nidogen-2 | NID2 | Q14112 | 22795 | NID2 | 1.519864 | 0 |
| SL008178 | Dermatopontin | DERM | Q07507 | 1805 | DPT | -1.530833 | 0 |
| SL008158 | Histone H3.1 | H31 | P68431 | 8350 | HIST1H3A | 1.766529 | 0 |
| SL007237 | Dual specificity mitogen-activated protein kinase kinase 4 | MP2K4 | P45985 | 6416 | MAP2K4 | -2.07002 | 0 |
| SL007207 | Thrombospondin-4 | TSP4 | P35443 | 7060 | THBS4 | -1.552548 | 0 |
| SL007206 | Thrombospondin-2 | TSP2 | P35442 | 7058 | THBS2 | 1.819658 | 0 |
| SL007173 | Granulins | GRN | P28799 | 2896 | GRN | 1.400472 | 0 |
| SL007153 | Lactoperoxidase | PERL | P22079 | 4025 | LPO | 1.703989 | 0 |
| SL007056 | Bone morphogenetic protein 10 | BMP10 | O95393 | 27302 | BMP10 | 1.68322 | 0 |
| SL007049 | Cystatin-F | CYTF | O76096 | 8530 | CST7 | 1.705942 | 0 |
| SL006924 | Agouti-related protein | ART | O00253 | 181 | AGRP | 1.667223 | 0 |
| SL006910 | Cathepsin L2 | Cathepsin V | O60911 | 1515 | CTSV | -2.056401 | 0 |
| SL006777 | Fetuin-B | FETUB | Q9UGM5 | 26998 | FETUB | -1.92966 | 0 |
| SL006694 | Beta-Ala-His dipeptidase | CNDP1 | Q96KN2 | 84735 | CNDP1 | -1.571536 | 0 |
| SL006550 | Extracellular matrix protein 1 | ECM1 | Q16610 | 1893 | ECM1 | -1.476729 | 0 |
| SL006512 | Hepatocyte growth factor activator | HGFA | Q04756 | 3083 | HGFAC | -1.520598 | 0 |
| SL006448 | Histidine-rich glycoprotein | HRG | P04196 | 3273 | HRG | -1.727647 | 0 |
| SL006406 | Plexin-C1 | PLXC1 | O60486 | 10154 | PLXNC1 | -1.439443 | 0 |
| SL005789 | Stanniocalcin-1 | Stanniocalcin-1 | P52823 | 6781 | STC1 | 1.693666 | 0 |
| SL005699 | Apolipoprotein L1 | Apo L1 | O14791 | 8542 | APOL1 | -1.49548 | 0 |
| SL005220 | Sonic hedgehog protein | Sonic Hedgehog | Q15465 | 6469 | SHH | -1.563579 | 0 |
| SL005213 | Tumor necrosis factor receptor superfamily member 19L | RELT | Q969Z4 | 84957 | RELT | 1.452355 | 0 |
| SL005199 | MHC class I polypeptide-related sequence A | MICA | Q29983 | 4276 | MICA | 1.423458 | 0 |
| SL005195 | Lymphocyte activation gene 3 protein | LAG-3 | P18627 | 3902 | LAG3 | -1.636341 | 0 |
| SL005188 | Interleukin-5 receptor subunit alpha | IL-5 Ra | Q01344 | 3568 | IL5RA | 2.129642 | 0 |
| SL005184 | Interleukin-23 | IL-23 | P29460, Q9NPF7 | 3593 51561 | IL12B IL23A | -1.434375 | 0 |
| SL005174 | Interleukin-17 receptor B | IL-17B R | Q9NRM6 | 55540 | IL17RB | -1.458197 | 0 |
| SL005171 | Insulin-like growth factor-binding protein 4 | IGFBP-4 | P22692 | 3487 | IGFBP4 | 1.431616 | 0 |
| SL005156 | Neuroblastoma suppressor of tumorigenicity 1 | DAN | P41271 | 4681 | NBL1 | 1.502203 | 0 |
| SL005153 | Ciliary neurotrophic factor receptor subunit alpha | CNTFR alpha | P26992 | 1271 | CNTFR | -1.449541 | 0 |
| SL004876 | Kallistatin | Kallistatin | P29622 | 5267 | SERPINA4 | -1.750621 | 0 |
| SL004867 | Carbonic anhydrase 3 | Carbonic anhydrase III | P07451 | 761 | CA3 | 2.002668 | 0 |
| SL004866 | Carbonic anhydrase 1 | Carbonic anhydrase I | P00915 | 759 | CA1 | -2.000777 | 0 |
| SL004783 | Protein S100-A12 | S100A12 | P80511 | 6283 | S100A12 | 1.874156 | 0 |
| SL004742 | Afamin | Afamin | P43652 | 173 | AFM | -1.454748 | 0 |
| SL004718 | Importin subunit alpha-1 | Karyopherin-a2 | P52292 | 3838 | KPNA2 | 1.62533 | 0 |
| SL004650 | Secreted frizzled-related protein 3 | sFRP-3 | Q92765 | 2487 | FRZB | -1.603896 | 0 |
| SL004594 | Troponin I, fast skeletal muscle | Troponin I, skeletal, fast twitch | P48788 | 7136 | TNNI2 | 6.836947 | 0 |
| SL004579 | Macrophage mannose receptor 1 | Macrophage mannose receptor | P22897 | 4360 | MRC1 | 1.513855 | 0 |
| SL004536 | Hepcidin | LEAP-1 | P81172 | 57817 | HAMP | 3.891643 | 0 |
| SL004489 | Toll-like receptor 4 | TLR4 | O00206 | 7099 | TLR4 | 1.656259 | 0 |
| SL004489 | Toll-like receptor 4 | TLR4 | O00206 | 7099 | TLR4 | 1.656259 | 0 |
| SL004469 | Amyloid beta A4 protein | amyloid precursor protein | P05067 | 351 | APP | -1.591899 | 0 |
| SL004467 | Protein Z-dependent protease inhibitor | protein Z inhibitor | Q9UK55 | 51156 | SERPINA10 | 1.782611 | 0 |
| SL004466 | Heparin cofactor 2 | Heparin cofactor II | P05546 | 3053 | SERPIND1 | -1.65882 | 0 |
| SL004400 | Coagulation factor IXab | Coagulation Factor IXab | P00740 | 2158 | F9 | 1.427223 | 0 |
| SL004367 | Dickkopf-related protein 1 | DKK1 | O94907 | 22943 | DKK1 | -1.486654 | 0 |
| SL004357 | Growth/differentiation factor 8 | Myostatin | O14793 | 2660 | MSTN | -1.599033 | 0 |
| SL004260 | Resistin | resistin | Q9HD89 | 56729 | RETN | 1.667353 | 0 |
| SL004183 | Cadherin-3 | P-Cadherin | P22223 | 1001 | CDH3 | -2.152574 | 0 |
| SL004154 | Neural cell adhesion molecule L1 | NCAM-L1 | P32004 | 3897 | L1CAM | -1.528782 | 0 |
| SL004152 | Interleukin-18 receptor 1 | IL-18 Ra | Q13478 | 8809 | IL18R1 | 1.450982 | 0 |
| SL004146 | Interleukin-1 receptor-like 1 | IL-1 R4 | Q01638 | 9173 | IL1RL1 | 6.93232 | 0 |
| SL004134 | CD27 antigen | CD27 | P26842 | 939 | CD27 | -1.569329 | 0 |
| SL004016 | C-X-C motif chemokine 16 | CXCL16, soluble | Q9H2A7 | 58191 | CXCL16 | 1.57103 | 0 |
| SL004008 | Myeloblastin | Proteinase-3 | P24158 | 5657 | PRTN3 | 2.654078 | 0 |
| SL003869 | Growth/differentiation factor 15 | MIC-1 | Q99988 | 9518 | GDF15 | 1.747731 | 0 |
| SL003770 | Secreted frizzled-related protein 1 | SARP-2 | Q8N474 | 6422 | SFRP1 | -1.773037 | 0 |
| SL003764 | Neural cell adhesion molecule 1, 120 kDa isoform | NCAM-120 | P13591 | 4684 | NCAM1 | -1.447584 | 0 |
| SL003722 | RAC-beta serine/threonine-protein kinase | PKB beta | P31751 | 208 | AKT2 | -2.447023 | 0 |
| SL003680 | Advanced glycosylation end product-specific receptor, soluble | sRAGE | Q15109 | 177 | AGER | -1.567755 | 0 |
| SL003340 | Chitinase-3-like protein 1 | YKL-40 | P36222 | 1116 | CHI3L1 | 5.943921 | 0 |
| SL003329 | C-C motif chemokine 14 | HCC-1 | Q16627 | 6358 | CCL14 | 1.629337 | 0 |
| SL003326 | C-X-C motif chemokine 11 | I-TAC | O14625 | 6373 | CXCL11 | -1.903122 | 0 |
| SL003309 | Lipopolysaccharide-binding protein | LBP | P18428 | 3929 | LBP | 1.541969 | 0 |
| SL003303 | C-C motif chemokine 28 | CCL28 | Q9NRJ3 | 56477 | CCL28 | -1.593283 | 0 |
| SL003302 | C-C motif chemokine 23 | MPIF-1 | P55773 | 6368 | CCL23 | 3.425021 | 0 |
| SL003301 | Ck-beta-8-1 | Ck-b-8-1 | P55773 | 6368 | CCL23 | 3.366869 | 0 |
| SL003300 | C-C motif chemokine 16 | HCC-4 | O15467 | 6360 | CCL16 | -1.660056 | 0 |
| SL003201 | Vascular endothelial growth factor receptor 2 | VEGF sR2 | P35968 | 3791 | KDR | -1.449614 | 0 |
| SL003189 | C-C motif chemokine 19 | MIP-3b | Q99731 | 6363 | CCL19 | 1.542045 | 0 |
| SL003188 | C-X-C motif chemokine 9 | MIG | Q07325 | 4283 | CXCL9 | -1.49343 | 0 |
| SL003172 | C-X-C motif chemokine 6 | GCP-2 | P80162 | 6372 | CXCL6 | -1.579405 | 0 |
| SL003167 | C-X-C motif chemokine 13 | BLC | O43927 | 10563 | CXCL13 | 1.476656 | 0 |
| SL002785 | N-terminal pro-BNP | N-terminal pro-BNP | P16860 | 4879 | NPPB | 3.474763 | 0 |
| SL002762 | Chromogranin-A | CgA | P10645 | 1113 | CHGA | 1.567788 | 0 |
| SL002755 | Pappalysin-1 | PAPP-A | Q13219 | 5069 | PAPPA | 2.087058 | 0 |
| SL002705 | Thrombospondin-1 | Thrombospondin-1 | P07996 | 7057 | THBS1 | -1.952699 | 0 |
| SL002704 | Pleiotrophin | PTN | P21246 | 5764 | PTN | -1.526857 | 0 |
| SL002688 | Osteopontin | Osteopontin | P10451 | 6696 | SPP1 | 2.162912 | 0 |
| SL002684 | Macrophage colony-stimulating factor 1 | CSF-1 | P09603 | 1435 | CSF1 | 1.509591 | 0 |
| SL002662 | Coagulation Factor XI | Coagulation Factor XI | P03951 | 2160 | F11 | -1.460452 | 0 |
| SL002621 | Midkine | Midkine | P21741 | 4192 | MDK | -2.069289 | 0 |
| SL002528 | Phospholipase A2, membrane associated | NPS-PLA2 | P14555 | 5320 | PLA2G2A | 9.368349 | 0 |
| SL002508 | Interleukin-18-binding protein | IL-18 BPa | O95998 | 10068 | IL18BP | 1.808122 | 0 |
| SL002505 | Atrial natriuretic factor | ANP | P01160 | 4878 | NPPA | 1.773959 | 0 |
| SL002093 | Histone H2A.z | Histone H2A.z | P0C0S5 | 3015 | H2AFZ | 2.976031 | 0 |
| SL001996 | Angiopoietin-2 | Angiopoietin-2 | O15123 | 285 | ANGPT2 | 2.312921 | 0 |
| SL001995 | Angiopoietin-1 | Angiopoietin-1 | Q15389 | 284 | ANGPT1 | -1.496837 | 0 |
| SL001992 | Tumor necrosis factor receptor superfamily member 1A | TNF sR-I | P19438 | 7132 | TNFRSF1A | 1.756859 | 0 |
| SL001990 | Interleukin-1 receptor antagonist protein | IL-1Ra | P18510 | 3557 | IL1RN | 1.822241 | 0 |
| SL001947 | Melanoma-derived growth regulatory protein | MIA | Q16674 | 8190 | MIA | -1.430124 | 0 |
| SL001888 | Antileukoproteinase | SLPI | P03973 | 6590 | SLPI | 1.545633 | 0 |
| SL001800 | Tumor necrosis factor receptor superfamily member 1B | TNF sR-II | P20333 | 7133 | TNFRSF1B | 1.617016 | 0 |
| SL001774 | Fatty acid-binding protein, heart | FABP | P05413 | 2170 | FABP3 | 1.495252 | 0 |
| SL001761 | Troponin I, cardiac muscle | Troponin I | P19429 | 7137 | TNNI3 | 6.37992 | 0 |
| SL001729 | Granulocyte colony-stimulating factor | G-CSF | P09919 | 1440 | CSF3 | 1.680855 | 0 |
| SL000836 | Hemoglobin | Hemoglobin | P69905, P68871 | 3039 3043 | HBA1 HBB | -2.296679 | 0 |
| SL000640 | Nidogen-1 | Nidogen | P14543 | 4811 | NID1 | 1.666783 | 0 |
| SL000598 | Thrombopoietin | Tpo | P40225 | 7066 | THPO | 2.204794 | 0 |
| SL000591 | Metalloproteinase inhibitor 1 | TIMP-1 | P01033 | 7076 | TIMP1 | 2.150487 | 0 |
| SL000572 | Serum amyloid A-1 protein | SAA | P0DJI8 | 6288 | SAA1 | 47.943744 | 0 |
| SL000566 | Retinol-binding protein 4 | RBP | P02753 | 5950 | RBP4 | -1.446875 | 0 |
| SL000563 | C-C motif chemokine 5 | RANTES | P13501 | 6352 | CCL5 | -2.374728 | 0 |
| SL000550 | Plasma serine protease inhibitor | PCI | P05154 | 5104 | SERPINA5 | -3.843791 | 0 |
| SL000545 | Plasma kallikrein | Prekallikrein | P03952 | 3818 | KLKB1 | -1.632923 | 0 |
| SL000541 | Plasminogen | Plasminogen | P00747 | 5340 | PLG | -1.791432 | 0 |
| SL000537 | Platelet-derived growth factor subunit B | PDGF-BB | P01127 | 5155 | PDGFB | -2.317799 | 0 |
| SL000535 | Platelet-derived growth factor subunit A | PDGF-AA | P04085 | 5154 | PDGFA | -1.834954 | 0 |
| SL000532 | SPARC | ON | P09486 | 6678 | SPARC | -1.929011 | 0 |
| SL000528 | NADPH--cytochrome P450 reductase | NADPH-P450 Oxidoreductase | P16435 | 5447 | POR | -1.652632 | 0 |
| SL000522 | Macrophage metalloelastase | MMP-12 | P39900 | 4321 | MMP12 | -1.439746 | 0 |
| SL000521 | Interstitial collagenase | MMP-1 | P03956 | 4312 | MMP1 | -1.447815 | 0 |
| SL000507 | Lymphotoxin alpha1:beta2 | Lymphotoxin a1/b2 | P01374, Q06643 | 4049 4050 | LTA LTB | -1.633256 | 0 |
| SL000506 | Luteinizing hormone | Luteinizing hormone | P01215, P01229 | 1081 3972 | CGA LHB | -1.651403 | 0 |
| SL000496 | Lactotransferrin | Lactoferrin | P02788 | 4057 | LTF | -1.82915 | 0 |
| SL000478 | Interleukin-2 | IL-2 | P60568 | 3558 | IL2 | 1.417921 | 0 |
| SL000468 | Immunoglobulin M | IgM | P01871 | 3507 3512 50802 3535 | IGHM IGJ IGK@ IGL@ | -1.513826 | 0 |
| SL000451 | Heat shock 70 kDa protein 1A | HSP 70 | P0DMV8 | 3303 | HSPA1A | 2.10375 | 0 |
| SL000428 | Follicle stimulating hormone | FSH | P01215, P01225 | 1081 2488 | CGA FSHB | -1.710933 | 0 |
| SL000420 | Ferritin | Ferritin | P02794 P02792 | 2495 2512 | FTH1 FTL | 1.874723 | 0 |
| SL000408 | Erythropoietin | Epo | P01588 | 2056 | EPO | 4.304497 | 0 |
| SL000406 | Eotaxin | Eotaxin | P51671 | 6356 | CCL11 | -1.762473 | 0 |
| SL000383 | Creatine kinase M-type | CK-MM | P06732 | 1158 | CKM | 3.294865 | 0 |
| SL000382 | Creatine kinase M-type:Creatine kinase B-type heterodimer | CK-MB | P12277 P06732 | 1152 1158 | CKB CKM | 6.126634 | 0 |
| SL000344 | Cathepsin D | Cathepsin D | P07339 | 1509 | CTSD | 1.545278 | 0 |
| SL000338 | Calpastatin | Calpastatin | P20810 | 831 | CAST | -1.652613 | 0 |
| SL000314 | Complement C3b | C3b | P01024 | 718 | C3 | -1.695539 | 0 |
| SL000310 | Complement C1r subcomponent | C1r | P00736 | 715 | C1R | 1.544617 | 0 |
| SL000271 | Angiotensinogen | Angiotensinogen | P01019 | 183 | AGT | 1.537448 | 0 |
| SL000268 | Angiostatin | Angiostatin | P00747 | 5340 | PLG | -1.822922 | 0 |
| SL000252 | Alpha-2-macroglobulin | a2-Macroglobulin | P01023 | 2 | A2M | -1.405235 | 0 |
| SL000249 | Alpha-1-antitrypsin | a1-Antitrypsin | P01009 | 5265 | SERPINA1 | 1.604008 | 0 |
| SL000164 | Myoglobin | Myoglobin | P02144 | 4151 | MB | 2.068413 | 0 |
| SL000089 | Transforming growth factor beta-3 | TGF-b3 | P10600 | 7043 | TGFB3 | 1.663424 | 0 |
| SL000087 | Interleukin-6 | IL-6 | P05231 | 3569 | IL6 | 5.868963 | 0 |
| SL000070 | Glypican-3 | Glypican 3 | P51654 | 2719 | GPC3 | -1.575042 | 0 |
| SL000064 | Kallikrein-7 | Kallikrein 7 | P49862 | 5650 | KLK7 | -1.785847 | 0 |
| SL000062 | Prostate-specific antigen | PSA | P07288 | 354 | KLK3 | 1.486764 | 0 |
| SL000052 | Troponin T, cardiac muscle | Troponin T | P45379 | 7139 | TNNT2 | 6.963777 | 0 |
| SL000051 | C-reactive protein | CRP | P02741 | 1401 | CRP | 2.632733 | 0 |
| SL000021 | Insulin | Insulin | P01308 | 3630 | INS | 1.456021 | 0 |
| SL000019 | Apolipoprotein A-I | Apo A-I | P01857 | 335 | APOA1 | -1.620674 | 0 |
| SL013488 | C-type lectin domain family 1 member B | CLC1B | Q9P126 | 51266 | CLEC1B | -1.597125 | 0.000001 |
| SL010388 | Trypsin-2 | Trypsin 2 | P07478 | 5645 | PRSS2 | 1.501958 | 0.000001 |
| SL006523 | Lactadherin | MFGM | Q08431 | 4240 | MFGE8 | 1.939035 | 0.000001 |
| SL003191 | Neutrophil-activating peptide 2 | NAP-2 | P02775 | 5473 | PPBP | -1.80071 | 0.000001 |
| SL003187 | C-C motif chemokine 22 | MDC | O00626 | 6367 | CCL22 | -1.422126 | 0.000001 |
| SL001691 | Fibroblast growth factor 7 | FGF7 | P21781 | 2252 | FGF7 | -1.501007 | 0.000001 |
| SL000603 | Trypsin-1 | Trypsin | P07477 | 5644 | PRSS1 | -1.728136 | 0.000001 |
| SL000565 | Renin | Renin | P00797 | 5972 | REN | 1.531811 | 0.000001 |
| SL005115 | Spondin-1 | Spondin-1 | Q9HCB6 | 10418 | SPON1 | -1.421943 | 0.000002 |
| SL005102 | Sex hormone-binding globulin | SHBG | P04278 | 6462 | SHBG | -1.422654 | 0.000003 |
| SL010391 | WAP, Kazal, immunoglobulin, Kunitz and NTR domain-containing protein 2 | WFKN2 | Q8TEU8 | 124857 | WFIKKN2 | 1.413565 | 0.000004 |
| SL004708 | Connective tissue-activating peptide III | CTAP-III | P02775 | 5473 | PPBP | -1.816836 | 0.000004 |
| SL003993 | Bone morphogenetic protein 6 | BMP-6 | P22004 | 654 | BMP6 | -1.404326 | 0.000004 |
| SL001998 | Tissue factor pathway inhibitor | TFPI | P10646 | 7035 | TFPI | -1.505465 | 0.000006 |
| SL004296 | Adenylate kinase isoenzyme 1 | Myokinase, human | P00568 | 203 | AK1 | -1.671402 | 0.000007 |
| SL007651 | Fibroblast growth factor 23 | FGF23 | Q9GZV9 | 8074 | FGF23 | 1.550432 | 0.000009 |
| SL008466 | 3-hydroxyanthranilate 3,4-dioxygenase | 3HAO | P46952 | 23498 | HAAO | 1.41024 | 0.000011 |
| SL000526 | Neutrophil collagenase | MMP-8 | P22894 | 4317 | MMP8 | 1.673444 | 0.000011 |
| SL007024 | Gremlin-1 | GREM1 | O60565 | 26585 | GREM1 | 1.481451 | 0.000012 |
| SL000546 | Prolactin | PRL | P01236 | 5617 | PRL | 1.626853 | 0.000012 |
| SL003332 | Matrix metalloproteinase-17 | MMP-17 | Q9ULZ9 | 4326 | MMP17 | 1.649023 | 0.000014 |
| SL006830 | Complement factor H-related protein 5 | complement factor H-related 5 | Q9BXR6 | 81494 | CFHR5 | 1.414197 | 0.00002 |
| SL006992 | Matrilin-3 | MATN3 | O15232 | 4148 | MATN3 | 1.459276 | 0.000026 |
| SL004142 | Ephrin-B3 | Ephrin-B3 | Q15768 | 1949 | EFNB3 | 1.482625 | 0.000027 |
| SL005694 | Peroxiredoxin-6 | Peroxiredoxin-6 | P30041 | 9588 | PRDX6 | -1.569743 | 0.000037 |
| SL005629 | N-acetyl-D-glucosamine kinase | NAGK | Q9UJ70 | 55577 | NAGK | -1.548361 | 0.000085 |
| SL004157 | Tumor necrosis factor receptor superfamily member 10B | TRAIL R2 | O14763 | 8795 | TNFRSF10B | 1.497203 | 0.000097 |
| SL000466 | Insulin-like growth factor-binding protein 2 | IGFBP-2 | P18065 | 3485 | IGFBP2 | 1.458578 | 0.000121 |
| SL000589 | Thyroid Stimulating Hormone | TSH | P01215 P01222 | 1081 12372 | CGA TSHB | 1.577651 | 0.000124 |
| SL000426 | Fibronectin | Fibronectin | P02751 | 2335 | FN1 | -1.520872 | 0.000128 |
| SL016557 | 3-hydroxy-3-methylglutaryl-coenzyme A reductase | HMGR | P04035 | 3156 | HMGCR | 1.410911 | 0.000152 |
| SL017614 | RAC-alpha/beta/gamma serine/threonine-protein kinase | PKB a/b/g | P31749 P31751 Q9Y243 | 207 208 10000 | AKT1 AKT2 AKT3 | -1.571498 | 0.0002 |
| SL007229 | Signal transducer and activator of transcription 6 | STAT6 | P42226 | 6778 | STAT6 | -1.439924 | 0.000253 |
| SL000462 | Insulin-like growth factor-binding protein 1 | IGFBP-1 | P08833 | 3484 | IGFBP1 | 1.705807 | 0.000269 |
| SL017289 | PolyUbiquitin K48-linked | PolyUbiquitin K48 | P0CG47 | 7314 | UBB | -1.406714 | 0.000339 |
| SL005358 | Phosphatidylethanolamine-binding protein 1 | prostatic binding protein | P30086 | 5037 | PEBP1 | -1.563466 | 0.000435 |
| SL005215 | Myeloid cell surface antigen CD33 | Siglec-3 | P20138 | 945 | CD33 | -1.424511 | 0.000436 |
| SL004349 | Interferon lambda-2 | IFN-lambda 2 | Q8IZJ0 | 282616 | IFNL2 | 1.454259 | 0.000521 |
| SL004724 | Histone acetyltransferase KAT6A | MOZ | Q92794 | 7994 | KAT6A | 1.429254 | 0.000548 |
| SL004258 | Adiponectin | Adiponectin | Q15848 | 9370 | ADIPOQ | -1.426814 | 0.000663 |
| SL004914 | Inorganic pyrophosphatase | PPase | Q15181 | 5464 | PPA1 | -1.478972 | 0.000677 |
| SL000401 | Neutrophil elastase | Elastase | P08246 | 1991 | ELANE | 1.445816 | 0.000757 |
| SL005846 | Moesin | Moesin | P26038 | 4478 | MSN | -1.514323 | 0.000765 |
| SL007806 | Interleukin-22 receptor subunit alpha-1 | IL22RA1 | Q8N6P7 | 58985 | IL22RA1 | 1.447708 | 0.000769 |
| SL009628 | Inhibitor of growth protein 1 | ING1 | Q9UK53 | 3621 | ING1 | -1.503011 | 0.000903 |
| SL000584 | Transforming growth factor beta-1 | TGF-b1 | P01137 | 7040 | TGFB1 | 1.428287 | 0.000924 |
| SL000320 | C5a anaphylatoxin | C5a | P01031 | 727 | C5 | 1.495413 | 0.001099 |
| SL003196 | C-C motif chemokine 17 | TARC | Q92583 | 6361 | CCL17 | -1.52678 | 0.001201 |
| SL008143 | Ubiquitin-conjugating enzyme E2 N | UBE2N | P61088 | 7334 | UBE2N | -1.516802 | 0.001501 |
| SL008039 | Alcohol dehydrogenase [NADP(+)] | AK1A1 | P14550 | 10327 | AKR1A1 | -1.419339 | 0.00175 |
| SL012561 | Regenerating islet-derived protein 4 | REG4 | Q9BYZ8 | 83998 | REG4 | -1.482732 | 0.003249 |
| SL004757 | Vacuolar protein sorting-associated protein VTA1 homolog | DRG-1 | Q9NP79 | 51534 | VTA1 | -1.508956 | 0.006823 |
| SL018900 | Formimidoyltransferase-cyclodeaminase | FTCD | O95954 | 10841 | FTCD | -2.353564 | 0.007073 |
| SL005034 | GTP-binding nuclear protein Ran | RAN | P62826 | 5901 | RAN | -1.445606 | 0.010052 |
| SL005687 | Eukaryotic translation initiation factor 5A-1 | eIF-5A-1 | P63241 | 1984 | EIF5A | -1.461013 | 0.012451 |
| SL007640 | C-type lectin domain family 7 member A | CLC7A | Q9BXN2 | 64581 | CLEC7A | 1.405634 | 0.013566 |
| SL006705 | Prefoldin subunit 5 | PFD5 | Q99471 | 5204 | PFDN5 | 1.430642 | 0.014924 |
| SL015046 | Amphoterin-induced protein 2 | AMGO2 | Q86SJ2 | 347902 | AMIGO2 | -1.497656 | 0.017344 |
| SL010500 | Tyrosine-protein kinase Lyn, isoform B | LYNB | P07948 | 4067 | LYN | -1.400726 | 0.022029 |
| SL008759 | Platelet glycoprotein VI | GPVI | Q9HCN6 | 51206 | GP6 | -1.437571 | 0.022343 |
| SL004660 | Bone sialoprotein 2 | BSP | P21815 | 3381 | IBSP | 1.515668 | 0.024157 |
| SL000312 | Complement C3 | C3 | P01024 | 718 | C3 | -1.480631 | 0.026581 |
| SL003179 | Integrin alpha-I: beta-1 complex | Integrin a1b1 | P56199, P05556 | 3672 3688 | ITGA1 ITGB1 | 1.406263 | 0.034624 |
| SL005256 | Histone H1.2 | Histone H1.2 | P16403 | 3006 | HIST1H1C | 1.660463 | 0.040518 |
| SL003970 | Parathyroid hormone | PTH | P01270 | 5741 | PTH | 1.505947 | 0.064327 |
| SL000695 | Neutrophil gelatinase-associated lipocalin | Lipocalin 2 | P80188 | 3934 | LCN2 | 1.222895 | 0.041578 |
| SL016969 | Cysteine and glycine-rich protein 3 | CSRP3 | P50461 | 8048 | CSRP3 | 1.320712 | 0 |
| SL016555 | Dual 3',5'-cyclic-AMP and -GMP phosphodiesterase 11A | PDE11 | Q9HCR9 | 50940 | PDE11A | 1.335867 | 0 |
| SL014292 | Sialic acid-binding Ig-like lectin 14 | SIG14 | Q08ET2 | 100049587 | SIGLEC14 | 1.386988 | 0 |
| SL014288 | Cell surface glycoprotein CD200 receptor 1 | MO2R1 | Q8TD46 | 131450 | CD200R1 | -1.209145 | 0 |
| SL014130 | Carbohydrate sulfotransferase 6 | CHST6 | Q9GZX3 | 4166 | CHST6 | 1.314379 | 0 |
| SL014111 | Kin of IRRE-like protein 3 | KIRR3 | Q8IZU9 | 84623 | KIRREL3 | -1.174947 | 0 |
| SL014069 | Proprotein convertase subtilisin/kexin type 7 | PCSK7 | Q16549 | 9159 | PCSK7 | 1.343567 | 0 |
| SL014028 | Ectonucleoside triphosphate diphosphohydrolase 5 | ENTP5 | O75356 | 957 | ENTPD5 | 1.269702 | 0 |
| SL012881 | Glypican-6 | GPC6 | Q9Y625 | 10082 | GPC6 | -1.331221 | 0 |
| SL012469 | Glypican-5 | GPC5 | P78333 | 2262 | GPC5 | -1.352811 | 0 |
| SL010617 | Tryptase beta-2 | TPSB2 | P20231 | 64499 | TPSB2 | -1.381584 | 0 |
| SL010457 | Delta-like protein 4 | DLL4 | Q9NR61 | 54567 | DLL4 | 1.286405 | 0 |
| SL010455 | Contactin-5 | Contactin-5 | O94779 | 53942 | CNTN5 | -1.122076 | 0 |
| SL010454 | Contactin-4 | Contactin-4 | Q8IWV2 | 152330 | CNTN4 | -1.390821 | 0 |
| SL010375 | Neutral ceramidase | ASAH2 | Q9NR71 | 56624 | ASAH2 | -1.341604 | 0 |
| SL010368 | Alpha-L-iduronidase | IDUA | P35475 | 3425 | IDUA | -1.381542 | 0 |
| SL010348 | Fibronectin Fragment 4 | FN1.4 | P02751 | 2335 | FN1 | -1.219892 | 0 |
| SL009951 | Protein Wnt-7a | WNT7A | O00755 | 7476 | WNT7A | -1.122191 | 0 |
| SL009349 | Follistatin-related protein 1 | FSTL1 | Q12841 | 11167 | FSTL1 | 1.314594 | 0 |
| SL009213 | Lysosomal protective protein | Cathepsin A | P10619 | 5476 | CTSA | -1.377018 | 0 |
| SL009089 | Brevican core protein | PGCB | Q96GW7 | 63827 | BCAN | -1.290067 | 0 |
| SL008416 | C-type mannose receptor 2 | MRC2 | Q9UBG0 | 9902 | MRC2 | -1.367446 | 0 |
| SL008023 | Hyaluronan and proteoglycan link protein 1 | HPLN1 | P10915 | 1404 | HAPLN1 | -1.185806 | 0 |
| SL007673 | Netrin-4 | NET4 | Q9HB63 | 59277 | NTN4 | 1.381606 | 0 |
| SL007547 | Hepatitis A virus cellular receptor 2 | TIMD3 | Q8TDQ0 | 84868 | HAVCR2 | 1.29968 | 0 |
| SL007274 | Ephrin-B2 | EFNB2 | P52799 | 1948 | EFNB2 | 1.347401 | 0 |
| SL007217 | Interferon gamma receptor 2 | INGR2 | P38484 | 3460 | IFNGR2 | 1.174872 | 0 |
| SL007100 | Leukotriene A-4 hydrolase | LKHA4 | P09960 | 4048 | LTA4H | 1.311123 | 0 |
| SL006610 | A disintegrin and metalloproteinase with thrombospondin motifs 13 | ATS13 | Q76LX8 | 11093 | ADAMTS13 | -1.380507 | 0 |
| SL006542 | Ficolin-2 | FCN2 | Q15485 | 2220 | FCN2 | 1.298982 | 0 |
| SL006528 | Prolyl endopeptidase FAP | SEPR | Q12884 | 2191 | FAP | -1.391095 | 0 |
| SL006460 | Platelet glycoprotein Ib alpha chain | GP1BA | P07359 | 2811 | GP1BA | -1.223573 | 0 |
| SL006091 | Cytoplasmic protein NCK1 | NCK1 | P16333 | 4690 | NCK1 | -1.221861 | 0 |
| SL005392 | Arylsulfatase A | Arylsulfatase A | P15289 | 197 | ARSA | 1.2465 | 0 |
| SL005352 | Fatty acid-binding protein, epidermal | FABPE | Q01469 | 2171 | FABP5 | 1.231811 | 0 |
| SL005236 | Protein NOV homolog | NovH | P48745 | 4856 | NOV | -1.156006 | 0 |
| SL005205 | Natural cytotoxicity triggering receptor 3 | NKp30 | O14931 | 259197 | NCR3 | -1.13641 | 0 |
| SL005169 | Intercellular adhesion molecule 5 | sICAM-5 | Q9UMF0 | 7087 | ICAM5 | -1.392613 | 0 |
| SL005168 | Growth hormone receptor | Growth hormone receptor | P10912 | 2690 | GHR | -1.288109 | 0 |
| SL005155 | Teratocarcinoma-derived growth factor 1 | Cripto | P13385 | 6997 | TDGF1 | 1.324404 | 0 |
| SL004860 | Serine protease HTRA2, mitochondrial | HTRA2 | O43464 | 27429 | HTRA2 | 1.27073 | 0 |
| SL004855 | Contactin-1 | contactin-1 | Q12860 | 1272 | CNTN1 | -1.3931 | 0 |
| SL004782 | Tumor necrosis factor-inducible gene 6 protein | TSG-6 | P98066 | 7130 | TNFAIP6 | 1.367457 | 0 |
| SL004645 | Kunitz-type protease inhibitor 1 | HAI-1 | O43278 | 6692 | SPINT1 | -1.360485 | 0 |
| SL004644 | Ectodysplasin-A, secreted form | EDA | Q92838 | 1896 | EDA | -1.283341 | 0 |
| SL004642 | Disintegrin and metalloproteinase domain-containing protein 9 | ADAM 9 | Q13443 | 8754 | ADAM9 | 1.295216 | 0 |
| SL004365 | Tumor necrosis factor ligand superfamily member 12 | TWEAK | O43508 | 8742 | TNFSF12 | -1.319671 | 0 |
| SL004350 | Interleukin-17B | IL-17B | Q9UHF5 | 27190 | IL17B | -1.388578 | 0 |
| SL004298 | Granzyme A | granzyme A | P12544 | 3001 | GZMA | -1.287098 | 0 |
| SL004297 | Neuregulin-1 | NEUREGULIN-1 | Q02297 | 3084 | NRG1 | -1.26899 | 0 |
| SL004160 | BDNF/NT-3 growth factors receptor | TrkB | Q16620 | 4915 | NTRK2 | -1.346728 | 0 |
| SL004147 | Interleukin-10 receptor subunit beta | IL-10 Rb | Q08334 | 3588 | IL10RB | 1.277439 | 0 |
| SL004118 | Tartrate-resistant acid phosphatase type 5 | TrATPase | P13686 | 54 | ACP5 | -1.360867 | 0 |
| SL004010 | Mast/stem cell growth factor receptor Kit | SCF sR | P10721 | 3815 | KIT | -1.302787 | 0 |
| SL003739 | Tumor necrosis factor receptor superfamily member 6B | DcR3 | O95407 | 8771 | TNFRSF6B | 1.258192 | 0 |
| SL003192 | Properdin | Properdin | P27918 | 5199 | CFP | -1.13606 | 0 |
| SL003043 | Metalloproteinase inhibitor 3 | TIMP-3 | P35625 | 7078 | TIMP3 | -1.392319 | 0 |
| SL002782 | Adrenomedullin | Adrenomedullin | P35318 | 133 | ADM | -1.2362 | 0 |
| SL002561 | Parathyroid hormone-related protein | PTHrP | P12272 | 5744 | PTHLH | -1.335703 | 0 |
| SL002506 | Urokinase plasminogen activator surface receptor | suPAR | Q03405 | 5329 | PLAUR | 1.259573 | 0 |
| SL001897 | Kunitz-type protease inhibitor 2 | SPINT2 | O43291 | 10653 | SPINT2 | -1.319611 | 0 |
| SL001896 | Clusterin | Clusterin | P10909 | 1191 | CLU | -1.362123 | 0 |
| SL001815 | Superoxide dismutase [Mn], mitochondrial | Mn SOD | P04179 | 6648 | SOD2 | -1.394907 | 0 |
| SL001791 | Osteocalcin | Osteocalcin | P02818 | 632 | BGLAP | -1.149044 | 0 |
| SL001766 | Human Chorionic Gonadotropin | HCG | P01215,P01233 | 1081 1082 | CGA CGB | -1.363632 | 0 |
| SL001728 | Platelet-derived growth factor receptor alpha | PDGFRA | P16234 | 5156 | PDGFRA | 1.305066 | 0 |
| SL001716 | Interleukin-12 | IL-12 | P29459, P29460 | 3592 3593 | IL12A IL12B | -1.125498 | 0 |
| SL000668 | Platelet glycoprotein 4 | CD36 ANTIGEN | P16671 | 948 | CD36 | -1.274012 | 0 |
| SL000601 | Serotransferrin | Transferrin | P02787 | 7018 | TF | -1.13512 | 0 |
| SL000558 | Prothrombin | Prothrombin | P00734 | 2147 | F2 | -1.333014 | 0 |
| SL000515 | C-C motif chemokine 8 | MCP-2 | P80075 | 6355 | CCL8 | -1.144955 | 0 |
| SL000510 | Lysozyme C | Lysozyme | P61626 | 4069 | LYZ | -1.197662 | 0 |
| SL000508 | Lymphotoxin alpha2:beta1 | Lymphotoxin a2/b1 | P01374, Q06643 | 4049 4050 | LTA LTB | -1.21548 | 0 |
| SL000467 | Immunoglobulin G | IgG | P01857 | 3500 3501 3502 3503 50802 3535 | IGHG1 IGHG2 IGHG3 IGHG4 IGK@ IGL@ | -1.253164 | 0 |
| SL000424 | Fibrinogen | Fibrinogen | P02671 P02675 P02679 | 2243 2244 2266 | FGA FGB FGG | 1.084591 | 0 |
| SL000415 | Complement factor H | Factor H | P08603 | 3075 | CFH | -1.304553 | 0 |
| SL000396 | Cytochrome c | Cytochrome c | P99999 | 54205 | CYCS | -1.26126 | 0 |
| SL000357 | Coagulation factor IX | Coagulation Factor IX | P00740 | 2158 | F9 | 1.388675 | 0 |
| SL000316 | Complement C4 | C4 | P0C0L4, P0C0L5 | 720 721 | C4A C4B | -1.131105 | 0 |
| SL000272 | Antithrombin-III | Antithrombin III | P01008 | 462 | SERPINC1 | -1.217775 | 0 |
| SL000251 | Alpha-2-HS-glycoprotein | a2-HS-Glycoprotein | P02765 | 197 | AHSG | -1.389505 | 0 |
| SL000248 | Alpha-1-antichymotrypsin | a1-Antichymotrypsin | P01011 | 12 | SERPINA3 | 1.322255 | 0 |
| SL000084 | Epidermal growth factor | EGF | P01133 | 1950 | EGF | -1.365616 | 0 |
| SL018625 | Toll-like receptor 4:Lymphocyte antigen 96 complex | TLR4:MD-2 complex | O00206 Q9Y6Y9 | 7099 23643 | TLR4 LY96 | -1.204117 | 0.000001 |
| SL009400 | Chordin-like protein 1 | CRDL1 | Q9BU40 | 91851 | CHRDL1 | -1.328908 | 0.000001 |
| SL007680 | Roundabout homolog 2 | ROBO2 | Q9HCK4 | 6092 | ROBO2 | -1.347325 | 0.000001 |
| SL007429 | Transmembrane glycoprotein NMB | GPNMB | Q14956 | 10457 | GPNMB | -1.329842 | 0.000001 |
| SL004646 | Layilin | Layilin | Q6UX15 | 143903 | LAYN | 1.299911 | 0.000001 |
| SL004492 | Toll-like receptor 2 | TLR2 | O60603 | 7097 | TLR2 | 1.190462 | 0.000001 |
| SL004355 | C-C motif chemokine 3-like 1 | LD78-beta | P16619 | 414062 | CCL3L1 | 1.168452 | 0.000001 |
| SL004353 | Interleukin-17D | IL-17D | Q8TAD2 | 53342 | IL17D | 1.149192 | 0.000001 |
| SL003974 | Activated Protein C | Activated Protein C | P04070 | 5624 | PROC | 1.272345 | 0.000001 |
| SL003182 | Integrin alpha-V: beta-5 complex | Integrin aVb5 | P06756, P18084 | 3685 3693 | ITGAV ITGB5 | -1.289718 | 0.000001 |
| SL002519 | Receptor tyrosine-protein kinase erbB-3 | ERBB3 | P21860 | 2065 | ERBB3 | -1.306042 | 0.000001 |
| SL000358 | Coagulation factor VII | Coagulation Factor VII | P08709 | 2155 | F7 | -1.267267 | 0.000001 |
| SL000306 | Brain natriuretic peptide 32 | BNP-32 | P16860 | 4879 | NPPB | 1.318498 | 0.000001 |
| SL000048 | Vitamin K-dependent protein C | Protein C | P04070 | 5624 | PROC | -1.342938 | 0.000001 |
| SL011071 | Platelet-derived growth factor C | PDGF-CC | Q9NRA1 | 56034 | PDGFC | -1.182688 | 0.000002 |
| SL010460 | Low affinity immunoglobulin gamma Fc region receptor II-b | FCG2B | P31994 | 2213 | FCGR2B | -1.290184 | 0.000002 |
| SL007385 | Interleukin-24 | IL24 | Q13007 | 11009 | IL24 | 1.329722 | 0.000002 |
| SL005488 | SPARC-like protein 1 | SPARCL1 | Q14515 | 8404 | SPARCL1 | 1.346567 | 0.000002 |
| SL005185 | Interleukin-23 receptor | IL-23 R | Q5VWK5 | 149233 | IL23R | -1.305674 | 0.000002 |
| SL004759 | Eukaryotic translation initiation factor 5 | eIF-5 | P55010 | 1983 | EIF5 | -1.335029 | 0.000002 |
| SL004131 | T-lymphocyte activation antigen CD86 | B7-2 | P42081 | 942 | CD86 | -1.234658 | 0.000002 |
| SL003198 | Tenascin | Tenascin | P24821 | 3371 | TNC | 1.302339 | 0.000002 |
| SL000339 | Carbonic anhydrase 2 | carbonic anhydrase II | P00918 | 760 | CA2 | -1.1553 | 0.000002 |
| SL008728 | Neurexin-3-beta | NRX3B | Q9HDB5 | 9369 | NRXN3 | -1.282756 | 0.000003 |
| SL004155 | Platelet-derived growth factor receptor beta | PDGF Rb | P09619 | 5159 | PDGFRB | -1.283636 | 0.000003 |
| SL002602 | Trefoil factor 2 | Trefoil factor 2 | Q03403 | 7032 | TFF2 | 1.327584 | 0.000003 |
| SL000570 | Secretin | Secretin | P09683 | 6343 | SCT | -1.093051 | 0.000003 |
| SL010613 | Interleukin-17 receptor D | IL-17 RD | Q8NFM7 | 54756 | IL17RD | -1.147774 | 0.000004 |
| SL008709 | Desmocollin-3 | DSC3 | Q14574 | 1825 | DSC3 | -1.127951 | 0.000004 |
| SL007756 | Growth/differentiation factor 2 | GDF2 | Q9UK05 | 2658 | GDF2 | -1.244823 | 0.000004 |
| SL004845 | Ephrin type-B receptor 4 | EphB4 | P54760 | 2050 | EPHB4 | 1.132414 | 0.000004 |
| SL004747 | Apolipoprotein M | ApoM | O95445 | 55937 | APOM | -1.389308 | 0.000004 |
| SL004352 | Interleukin-17F | IL-17F | Q96PD4 | 112744 | IL17F | -1.234925 | 0.000004 |
| SL004327 | Tumor necrosis factor ligand superfamily member 13B | BAFF | Q9Y275 | 10673 | TNFSF13B | 1.30203 | 0.000004 |
| SL003755 | Induced myeloid leukemia cell differentiation protein Mcl-1 | Mcl-1 | Q07820 | 4170 | MCL1 | 1.382697 | 0.000004 |
| SL002644 | Epidermal growth factor receptor | ERBB1 | P00533 | 1956 | EGFR | -1.26953 | 0.000004 |
| SL000325 | Complement component C9 | C9 | P02748 | 735 | C9 | 1.19492 | 0.000004 |
| SL000299 | Fibroblast growth factor 1 | b-ECGF | P05230 | 2246 | FGF1 | -1.156242 | 0.000004 |
| SL012148 | Peroxisomal targeting signal 1 receptor | PEX5 | P50542 | 5830 | PEX5 | -1.132395 | 0.000005 |
| SL006892 | Tyrosine-protein kinase ABL1 | ABL1 | P00519 | 25 | ABL1 | 1.273108 | 0.000005 |
| SL004865 | Cadherin-6 | Cadherin-6 | P55285 | 1004 | CDH6 | -1.333963 | 0.000005 |
| SL004326 | Tumor necrosis factor ligand superfamily member 18 | TNFSF18 | Q9UNG2 | 8995 | TNFSF18 | 1.149398 | 0.000005 |
| SL000523 | Collagenase 3 | MMP-13 | P45452 | 4322 | MMP13 | 1.2245 | 0.000005 |
| SL004336 | Fibroblast growth factor 18 | FGF-18 | O76093 | 8817 | FGF18 | -1.191159 | 0.000006 |
| SL003177 | Intercellular adhesion molecule 2 | sICAM-2 | P13598 | 3384 | ICAM2 | -1.318682 | 0.000006 |
| SL000022 | D-dimer | D-dimer | P02671 P02675 P02679 | 2243 2244 2266 | FGA FGB FGG | 1.318946 | 0.000006 |
| SL014113 | Neuroligin-4, X-linked | NLGNX | Q8N0W4 | 57502 | NLGN4X | 1.30617 | 0.000009 |
| SL010464 | Leucine-rich repeats and immunoglobulin-like domains protein 3 | LRIG3 | Q6UXM1 | 121227 | LRIG3 | -1.27964 | 0.000009 |
| SL000622 | Coagulation Factor V | Coagulation Factor V | P12259 | 2153 | F5 | -1.146906 | 0.000009 |
| SL006911 | Serine/threonine-protein kinase Chk1 | CHK1 | O14757 | 1111 | CHEK1 | 1.183551 | 0.00001 |
| SL005228 | NKG2D ligand 2 | ULBP-2 | Q9BZM5 | 80328 | ULBP2 | -1.096405 | 0.00001 |
| SL005196 | Limbic system-associated membrane protein | LSAMP | Q13449 | 4045 | LSAMP | -1.339001 | 0.00001 |
| SL000139 | Epiregulin | EPI | O14944 | 2069 | EREG | -1.147598 | 0.00001 |
| SL012395 | Netrin-1 | NET1 | O95631 | 9423 | NTN1 | -1.369543 | 0.000011 |
| SL008609 | Low affinity immunoglobulin gamma Fc region receptor III-B | FCG3B | O75015 | 2215 | FCGR3B | -1.395208 | 0.000011 |
| SL003687 | Nucleoside diphosphate kinase A | Nucleoside diphosphate kinase A | P15531 | 4830 | NME1 | -1.323717 | 0.000012 |
| SL000377 | Creatine kinase B-type | CK-BB | P12277 | 1152 | CKB | 1.317452 | 0.000012 |
| SL017610 | Gro-beta/gamma | Gro-b/g | P19876 P19875 | NaN | CXCL3 CXCL2 | -1.392179 | 0.000014 |
| SL005084 | Periostin | Periostin | Q15063 | 10631 | POSTN | -1.384487 | 0.000014 |
| SL003918 | Kallikrein-13 | kallikrein 13 | Q9UKR3 | 26085 | KLK13 | -1.120245 | 0.000014 |
| SL001943 | Interleukin-6 receptor subunit alpha | IL-6 sRa | P08887 | 3570 | IL6R | -1.374235 | 0.000015 |
| SL000024 | Tissue Factor | TF | P13726 | 2152 | F3 | 1.243412 | 0.000016 |
| SL000442 | Somatotropin | HGH | P01241 | 2688 | GH1 | -1.120399 | 0.000017 |
| SL004060 | Endothelin-converting enzyme 1 | Endothelin-converting enzyme 1 | P42892 | 1889 | ECE1 | -1.358892 | 0.000018 |
| SL008639 | Iduronate 2-sulfatase | IDS | P22304 | 3423 | IDS | 1.375278 | 0.000019 |
| SL003717 | Caspase-10 | Caspase-10 | Q92851 | 843 | CASP10 | 1.111844 | 0.000019 |
| SL002654 | Ephrin type-A receptor 2 | Epithelial cell kinase | P29317 | 1969 | EPHA2 | 1.262624 | 0.000019 |
| SL010451 | Cryptic protein | CFC1 | P0CG37 | 55997 | CFC1 | -1.213999 | 0.00002 |
| SL000309 | Complement C1q subcomponent | C1q | P02745 P02746 P02747 | 712 713 714 | C1QA C1QB C1QC | -1.398072 | 0.000021 |
| SL000250 | Alpha-2-antiplasmin | a2-Antiplasmin | P08697 | 5345 | SERPINF2 | 1.342969 | 0.000021 |
| SL005764 | Scavenger receptor cysteine-rich type 1 protein M130 | sCD163 | Q86VB7 | 9332 | CD163 | -1.331165 | 0.000022 |
| SL011769 | Acidic leucine-rich nuclear phosphoprotein 32 family member B | AN32B | Q92688 | 10541 | ANP32B | 1.3041 | 0.000023 |
| SL011508 | Pituitary adenylate cyclase-activating polypeptide 27 | PACAP-27 | P18509 | 116 | ADCYAP1 | -1.251601 | 0.000023 |
| SL008909 | Legumain | LGMN | Q99538 | 5641 | LGMN | 1.281868 | 0.000024 |
| SL003322 | Vascular endothelial growth factor receptor 3 | VEGF sR3 | P35916 | 2324 | FLT4 | -1.347382 | 0.000024 |
| SL006970 | Delta-like protein 1 | DLL1 | O00548 | 28514 | DLL1 | 1.233749 | 0.000031 |
| SL004683 | Noggin | Noggin | Q13253 | 9241 | NOG | -1.250149 | 0.000031 |
| SL000049 | Vitamin K-dependent protein S | Protein S | P07225 | 5627 | PROS1 | -1.350039 | 0.000031 |
| SL001720 | Vascular cell adhesion protein 1 | VCAM-1 | P19320 | 7412 | VCAM1 | 1.327764 | 0.000032 |
| SL005200 | MHC class I polypeptide-related sequence B | MICB | Q29980 | 4277 | MICB | 1.234766 | 0.000033 |
| SL005059 | Transforming growth factor beta receptor type 3 | TGF-b R III | Q03167 | 7049 | TGFBR3 | 1.285209 | 0.000036 |
| SL000458 | Interferon gamma receptor 1 | IFN-g R1 | P15260 | 3459 | IFNGR1 | 1.318345 | 0.000039 |
| SL003323 | C-C motif chemokine 18 | PARC | P55774 | 6362 | CCL18 | 1.393292 | 0.000041 |
| SL004149 | Interleukin-13 receptor subunit alpha-1 | IL-13 Ra1 | P78552 | 3597 | IL13RA1 | 1.399577 | 0.000047 |
| SL004125 | Insulin receptor | IR | P06213 | 3643 | INSR | -1.322339 | 0.000048 |
| SL005183 | Interleukin-22 receptor subunit alpha-2 | IL-22BP | Q969J5 | 116379 | IL22RA2 | -1.341768 | 0.000051 |
| SL008967 | Tyrosine-protein phosphatase non-receptor type substrate 1 | SHPS1 | P78324 | 140885 | SIRPA | 1.307144 | 0.000054 |
| SL008099 | Macrophage-capping protein | CAPG | P40121 | 822 | CAPG | 1.32894 | 0.000056 |
| SL010530 | OCIA domain-containing protein 1 | OCAD1 | Q9NX40 | 54940 | OCIAD1 | 1.156827 | 0.000057 |
| SL003733 | Diablo homolog, mitochondrial | SMAC | Q9NR28 | 56616 | DIABLO | -1.106957 | 0.000057 |
| SL007306 | Protein FAM3B | FAM3B | P58499 | 54097 | FAM3B | -1.316446 | 0.000058 |
| SL002081 | Cadherin-5 | Cadherin-5 | P33151 | 1003 | CDH5 | -1.317547 | 0.00006 |
| SL002522 | Retinoblastoma-associated protein | Rb | P06400 | 5925 | RB1 | -1.167253 | 0.000065 |
| SL010379 | Semaphorin-3A | Semaphorin 3A | Q14563 | 10371 | SEMA3A | -1.128659 | 0.000071 |
| SL010372 | Enteropeptidase | Enterokinase | P98073 | 5651 | TMPRSS15 | 1.314673 | 0.000071 |
| SL003728 | Baculoviral IAP repeat-containing protein 3 | cIAP-2 | Q13489 | 330 | BIRC3 | -1.135678 | 0.000071 |
| SL001890 | Tumor-associated calcium signal transducer 2 | GA733-1 protein | P09758 | 4070 | TACSTD2 | -1.17202 | 0.000076 |
| SL005210 | Neuronal cell adhesion molecule | Nr-CAM | Q92823 | 4897 | NRCAM | -1.385301 | 0.000078 |
| SL004475 | Interferon alpha/beta receptor 1 | IFN-a/b R1 | P17181 | 3454 | IFNAR1 | 1.272579 | 0.000078 |
| SL007151 | Glycerol-3-phosphate dehydrogenase [NAD(+)], cytoplasmic | GPDA | P21695 | 2819 | GPD1 | 1.329759 | 0.000082 |
| SL000280 | Aspartate aminotransferase, cytoplasmic | GOT1 | P17174 | 2805 | GOT1 | 1.318554 | 0.000083 |
| SL000318 | Complement C4b | C4b | P0C0L4 P0C0L5 | 720 721 | C4A C4B | 1.378028 | 0.000084 |
| SL000461 | Immunoglobulin E | IgE | P01854 | 3497 50802 3535 | IGHE IGK@ IGL@ | 1.367656 | 0.000087 |
| SL000125 | Interleukin-1 alpha | IL-1a | P01583 | 3552 | IL1A | -1.208278 | 0.000089 |
| SL007531 | BMP-binding endothelial regulator protein | BMPER | Q8N8U9 | 168667 | BMPER | 1.327999 | 0.000094 |
| SL018947 | Ubiquitin carboxyl-terminal hydrolase 25 | UBP25 | Q9UHP3 | 29761 | USP25 | -1.257661 | 0.000095 |
| SL001726 | Granulocyte-macrophage colony-stimulating factor | GM-CSF | P04141 | 1437 | CSF2 | 1.354497 | 0.000095 |
| SL004081 | Decorin | Bone proteoglycan II | P07585 | 1634 | DCN | -1.06487 | 0.000096 |
| SL000678 | Granulysin | Granulysin | P22749 | 10578 | GNLY | -1.281987 | 0.000097 |
| SL006114 | Tyrosine-protein kinase transmembrane receptor ROR1 | ROR1 | Q01973 | 4919 | ROR1 | -1.318613 | 0.000103 |
| SL000002 | Vascular endothelial growth factor A | VEGF | P15692 | 7422 | VEGFA | 1.294386 | 0.000103 |
| SL005703 | Neurogenic locus notch homolog protein 1 | Notch 1 | P46531 | 4851 | NOTCH1 | -1.314178 | 0.000107 |
| SL009054 | Neurexin-1-beta | NRX1B | P58400 | 9378 | NRXN1 | -1.215165 | 0.000113 |
| SL007774 | Protein jagged-2 | JAG2 | Q9Y219 | 3714 | JAG2 | -1.107856 | 0.000115 |
| SL010371 | Ectonucleoside triphosphate diphosphohydrolase 1 | CD39 | P49961 | 953 | ENTPD1 | 1.185976 | 0.000119 |
| SL004698 | Protein E7_HPV18 | HPV E7 Type18 | P06788 | 1489089 | Human-virus | 1.226318 | 0.000121 |
| SL011809 | dCTP pyrophosphatase 1 | XTP3A | Q9H773 | 79077 | DCTPP1 | 1.284256 | 0.000136 |
| SL007328 | Protein jagged-1 | JAG1 | P78504 | 182 | JAG1 | -1.238314 | 0.000136 |
| SL004180 | Tumor necrosis factor receptor superfamily member 8 | CD30 | P28908 | 943 | TNFRSF8 | 1.2027 | 0.000144 |
| SL005159 | Erythropoietin receptor | EPO-R | P19235 | 2057 | EPOR | -1.182783 | 0.000147 |
| SL004858 | GDNF family receptor alpha-1 | GFRa-1 | P56159 | 2674 | GFRA1 | -1.169048 | 0.000147 |
| SL003193 | C-C motif chemokine 21 | 6Ckine | O00585 | 6366 | CCL21 | -1.057467 | 0.000149 |
| SL004333 | Fibroblast growth factor 10 | FGF-10 | O15520 | 2255 | FGF10 | -1.190164 | 0.000157 |
| SL005158 | C-type lectin domain family 4 member M | DC-SIGNR | Q9H2X3 | 10332 | CLEC4M | -1.166536 | 0.000163 |
| SL004712 | Stromal cell-derived factor 1 | SDF-1 | P48061 | 6387 | CXCL12 | 1.390208 | 0.000164 |
| SL003690 | Tumor necrosis factor receptor superfamily member 11A | RANK | Q9Y6Q6 | 8792 | TNFRSF11A | 1.103197 | 0.000188 |
| SL005165 | Galectin-4 | Galectin-4 | P56470 | 3960 | LGALS4 | -1.182283 | 0.000192 |
| SL007828 | Neuregulin-4 | NRG4 | Q8WWG1 | 145957 | NRG4 | -1.181145 | 0.000202 |
| SL003304 | Insulin-like growth factor 1 receptor | IGF-I sR | P08069 | 3480 | IGF1R | -1.352528 | 0.000208 |
| SL006108 | CD5 antigen-like | CD5L | O43866 | 922 | CD5L | -1.36324 | 0.000219 |
| SL006029 | Chitotriosidase-1 | Chitotriosidase-1 | Q13231 | 1118 | CHIT1 | -1.291226 | 0.000219 |
| SL011068 | Interleukin-17 receptor C | IL-17 RC | Q8NAC3 | 84818 | IL17RC | 1.216898 | 0.000224 |
| SL004342 | Fibroblast growth factor 8 isoform B | FGF-8B | P55075 | 2253 | FGF8 | 1.397844 | 0.000226 |
| SL005209 | Neurogenic locus notch homolog protein 3 | Notch-3 | Q9UM47 | 4854 | NOTCH3 | -1.25879 | 0.000228 |
| SL004668 | Apolipoprotein E (isoform E3) | Apo E3 | P02649 | 348 | APOE | 0.919713 | 0.000231 |
| SL010462 | Ficolin-1 | FCN1 | O00602 | 2219 | FCN1 | 1.331598 | 0.000238 |
| SL005224 | Thrombopoietin Receptor | Thrombopoietin Receptor | P40238 | 4352 | MPL | -1.196934 | 0.000239 |
| SL004139 | Ephrin-A3 | Ephrin-A3 | P52797 | 1944 | EFNA3 | -1.20124 | 0.000239 |
| SL004067 | Group 10 secretory phospholipase A2 | GX | O15496 | 8399 | PLA2G10 | 1.246637 | 0.00025 |
| SL011770 | Leucine carboxyl methyltransferase 1 | LCMT1 | Q9UIC8 | 51451 | LCMT1 | 1.206732 | 0.000251 |
| SL004354 | Interleukin-19 | IL-19 | Q9UHD0 | 29949 | IL19 | -1.291389 | 0.000259 |
| SL002565 | Myc proto-oncogene protein | c-Myc | P01106 | 4609 | MYC | -1.153148 | 0.000259 |
| SL014091 | VPS10 domain-containing receptor SorCS2 | SORC2 | Q96PQ0 | 57537 | SORCS2 | -1.14907 | 0.000261 |
| SL018891 | Down syndrome cell adhesion molecule | DSCAM | O60469 | 1826 | DSCAM | -1.13974 | 0.000269 |
| SL014093 | Ectonucleoside triphosphate diphosphohydrolase 3 | ENTP3 | O75355 | 956 | ENTPD3 | 1.308911 | 0.000269 |
| SL004143 | GDNF family receptor alpha-2 | GFRa-2 | O00451 | 2675 | GFRA2 | 1.176059 | 0.000269 |
| SL015728 | Glucokinase regulatory protein | GCKR | Q14397 | 2646 | GCKR | 1.185567 | 0.000272 |
| SL004984 | 14-3-3 protein epsilon | 14-3-3E | P62258 | 7531 | YWHAE | 1.290065 | 0.000272 |
| SL004669 | Apolipoprotein E (isoform E4) | Apo E4 | P02649 | 348 | APOE | 0.681281 | 0.000272 |
| SL012108 | 1-phosphatidylinositol 4,5-bisphosphate phosphodiesterase gamma-1 | PLCG1 | P19174 | 5335 | PLCG1 | -1.186597 | 0.000279 |
| SL001753 | Sialoadhesin | Sialoadhesin | Q9BZZ2 | 6614 | SIGLEC1 | -1.210611 | 0.000287 |
| SL010495 | Cyclin-dependent kinase 2:Cyclin-A2 complex | CDK2/cyclin A | P24941 P20248 | 1017 890 | CDK2 CCNA2 | 1.22029 | 0.000323 |
| SL004814 | Coactosin-like protein | Coactosin-like protein | Q14019 | 23406 | COTL1 | 1.178974 | 0.00033 |
| SL007560 | Stabilin-2 | STAB2 | Q8WWQ8 | 55576 | STAB2 | 1.209823 | 0.000333 |
| SL007179 | Ephrin type-B receptor 2 | EPHB2 | P29323 | 2048 | EPHB2 | -1.301508 | 0.000343 |
| SL003849 | Fibroblast growth factor 9 | FGF9 | P31371 | 2254 | FGF9 | -1.121505 | 0.000345 |
| SL004458 | Elafin | Elafin | P19957 | 5266 | PI3 | 1.095341 | 0.000358 |
| SL000145 | Interleukin-1 receptor type 2 | IL-1 sRII | P27930 | 7850 | IL1R2 | 1.340332 | 0.000358 |
| SL003166 | CD166 antigen | ALCAM | Q13740 | 214 | ALCAM | -1.265515 | 0.000361 |
| SL003803 | Receptor tyrosine-protein kinase erbB-4 | ERBB4 | Q15303 | 2066 | ERBB4 | -1.135013 | 0.000362 |
| SL005258 | Serine/threonine-protein kinase PLK1 | PLK-1 | P53350 | 5347 | PLK1 | 1.329738 | 0.00037 |
| SL005357 | Lithostathine-1-alpha | PSP | P05451 | 5967 | REG1A | 1.347889 | 0.000372 |
| SL009216 | Aromatic-L-amino-acid decarboxylase | dopa decarboxylase | P20711 | 1644 | DDC | -1.2659 | 0.000376 |
| SL002524 | T-cell surface glycoprotein CD4 | sCD4 | P01730 | 920 | CD4 | 1.33827 | 0.00038 |
| SL005166 | Galectin-7 | Galectin-7 | P47929 | 3963 | LGALS7 | -1.182354 | 0.000386 |
| SL000131 | Proliferating cell nuclear antigen | PCNA | P12004 | 5111 | PCNA | 1.111204 | 0.000388 |
| SL000524 | Stromelysin-1 | MMP-3 | P08254 | 4314 | MMP3 | 1.065878 | 0.000443 |
| SL006230 | Lumican | Lumican | P51884 | 4060 | LUM | -1.379225 | 0.000459 |
| SL000519 | C-C motif chemokine 3 | MIP-1a | P10147 | 6348 | CCL3 | 1.295883 | 0.000466 |
| SL018587 | Immunoglobulin superfamily containing leucine-rich repeat protein 2 | ISLR2 | Q6UXK2 | 57611 | ISLR2 | -1.314038 | 0.000468 |
| SL000509 | Tumor necrosis factor receptor superfamily member 3 | Lymphotoxin b R | P36941 | 4055 | LTBR | 1.294916 | 0.000485 |
| SL004868 | Carbonic anhydrase 7 | Carbonic anhydrase VII | P43166 | 766 | CA7 | 1.206624 | 0.000499 |
| SL004811 | Transgelin-2 | Transgelin-2 | P37802 | 8407 | TAGLN2 | -1.310759 | 0.000502 |
| SL004716 | Mitogen-activated protein kinase 9 | JNK2 | P45984 | 5601 | MAPK9 | 1.172109 | 0.000535 |
| SL004120 | Discoidin domain-containing receptor 2 | Discoidin domain receptor 2 | Q16832 | 4921 | DDR2 | 1.289393 | 0.000548 |
| SL005261 | Tyrosine-protein phosphatase non-receptor type 2 | TCPTP | P17706 | 5771 | PTPN2 | 1.263738 | 0.00055 |
| SL005221 | Scavenger receptor class F member 1 | SREC-I | Q14162 | 8578 | SCARF1 | -1.112662 | 0.00056 |
| SL000645 | Stromelysin-2 | MMP-10 | P09238 | 4319 | MMP10 | 1.266203 | 0.00056 |
| SL004516 | Mannose-binding protein C | MBL | P11226 | 4153 | MBL2 | 1.30714 | 0.000582 |
| SL005572 | Gelsolin | Gelsolin | P06396 | 2934 | GSN | -1.306563 | 0.000616 |
| SL004635 | Tumor necrosis factor ligand superfamily member 8 | CD30 Ligand | P32971 | 944 | TNFSF8 | -1.142978 | 0.000659 |
| SL000597 | Lymphotoxin-alpha | TNF-b | P01374 | 4049 | LTA | 1.270688 | 0.000674 |
| SL004625 | A disintegrin and metalloproteinase with thrombospondin motifs 4 | ADAMTS-4 | O75173 | 9507 | ADAMTS4 | 1.201366 | 0.000684 |
| SL004248 | Glial cell line-derived neurotrophic factor | GDNF | P39905 | 2668 | GDNF | 1.289684 | 0.000697 |
| SL001999 | E3 ubiquitin-protein ligase Mdm2 | MDM2 | Q00987 | 4193 | MDM2 | 1.279956 | 0.000697 |
| SL011532 | Persulfide dioxygenase ETHE1, mitochondrial | ETHE1 | O95571 | 23474 | ETHE1 | -1.26587 | 0.00071 |
| SL000670 | Glutathione S-transferase A3 | GSTA3 | Q16772 | 2940 | GSTA3 | -1.192763 | 0.000723 |
| SL000347 | Corticosteroid-binding globulin | CBG | P08185 | 866 | SERPINA6 | -1.10669 | 0.00073 |
| SL010461 | High affinity immunoglobulin gamma Fc receptor I | FCGR1 | P12314 | 2209 | FCGR1A | 1.378896 | 0.000747 |
| SL004697 | Protein E7_HPV16 | HPV E7 Type 16 | P03129 | 1489079 | Human-virus | 1.269706 | 0.000758 |
| SL000540 | Plasmin | Plasmin | P00747 | 5340 | PLG | 1.310232 | 0.000775 |
| SL005208 | Reticulon-4 receptor | Nogo Receptor | Q9BZR6 | 65078 | RTN4R | -1.262415 | 0.000794 |
| SL004339 | Fibroblast growth factor 5 | FGF-5 | P12034 | 2250 | FGF5 | 1.388598 | 0.000794 |
| SL000384 | Cytotoxic T-lymphocyte protein 4 | CTLA-4 | P16410 | 1493 | CTLA4 | -1.284267 | 0.000807 |
| SL004804 | Cell adhesion molecule 3 | Nectin-like protein 1 | Q8N126 | 57863 | CADM3 | 1.199155 | 0.000814 |
| SL000345 | Cathepsin G | Cathepsin G | P08311 | 1511 | CTSG | 1.29217 | 0.00085 |
| SL000587 | Thyroglobulin | Thyroglobulin | P01266 | 7038 | TG | 1.184518 | 0.00089 |
| SL004066 | Group IIE secretory phospholipase A2 | GIIE | Q9NZK7 | 30814 | PLA2G2E | 1.180251 | 0.000895 |
| SL003994 | Bone morphogenetic protein 1 | BMP-1 | P13497 | 649 | BMP1 | -1.370839 | 0.000904 |
| SL007310 | Ras-related C3 botulinum toxin substrate 3 | RAC3 | P60763 | 5881 | RAC3 | 1.228494 | 0.000913 |
| SL000590 | Thyroxine-binding globulin | Thyroxine-Binding Globulin | P05543 | 6906 | SERPINA7 | -1.261138 | 0.000913 |
| SL006805 | Serine/threonine-protein kinase MRCK beta | MRCKB | Q9Y5S2 | 9578 | CDC42BPB | 1.269802 | 0.000934 |
| SL005181 | Interleukin-20 receptor subunit alpha | IL-20 Ra | Q9UHF4 | 53832 | IL20RA | -1.124393 | 0.000972 |
| SL010250 | Stress-induced-phosphoprotein 1 | Stress-induced-phosphoprotein 1 | P31948 | 10963 | STIP1 | -1.286517 | 0.000974 |
| SL000088 | Transforming growth factor beta-2 | TGF-b2 | P61812 | 7042 | TGFB2 | 1.275354 | 0.001031 |
| SL000277 | Apolipoprotein E (isoform E2) | Apo E2 | P02649 | 348 | APOE | 1.140887 | 0.001049 |
| SL010465 | Matrilin-2 | MATN2 | O00339 | 4147 | MATN2 | 1.276141 | 0.00107 |
| SL004919 | Peroxiredoxin-1 | Peroxiredoxin-1 | Q06830 | 5052 | PRDX1 | -1.305958 | 0.001084 |
| SL004337 | Fibroblast growth factor 19 | FGF-19 | O95750 | 9965 | FGF19 | 1.274997 | 0.001096 |
| SL004144 | GDNF family receptor alpha-3 | GFRa-3 | O60609 | 2676 | GFRA3 | 1.286452 | 0.001107 |
| SL006132 | Lamin-B1 | Lamin-B1 | P20700 | 4001 | LMNB1 | 1.228757 | 0.001182 |
| SL001721 | Platelet endothelial cell adhesion molecule | PECAM-1 | P16284 | 5175 | PECAM1 | 1.372496 | 0.001207 |
| SL008421 | A disintegrin and metalloproteinase with thrombospondin motifs 1 | ATS1 | Q9UHI8 | 9510 | ADAMTS1 | 1.281436 | 0.001257 |
| SL004063 | Fibroblast growth factor receptor 3 | FGFR-3 | P22607 | 2261 | FGFR3 | 1.240952 | 0.001257 |
| SL002539 | Tumor necrosis factor receptor superfamily member 11B | OPG | O00300 | 4982 | TNFRSF11B | 1.247235 | 0.001319 |
| SL000017 | von Willebrand factor | vWF | P04275 | 7450 | VWF | 0.822058 | 0.001322 |
| SL003308 | Interleukin-4 receptor subunit alpha | IL-4 sR | P24394 | 3566 | IL4R | -1.177384 | 0.001362 |
| SL000283 | Beta-2-microglobulin | b2-Microglobulin | P61769 | 567 | B2M | 1.260524 | 0.001428 |
| SL008773 | CD109 antigen | CD109 | Q6YHK3 | 135228 | CD109 | -1.316993 | 0.001439 |
| SL018971 | E3 ubiquitin-protein ligase ZNRF3 | ZNRF3 | Q9ULT6 | 84133 | ZNRF3 | -1.137932 | 0.001469 |
| SL008402 | Ly6/PLAUR domain-containing protein 3 | LYPD3 | O95274 | 27076 | LYPD3 | -1.242301 | 0.001469 |
| SL004329 | Growth/differentiation factor 5 | BMP-14 | P43026 | 8200 | GDF5 | -1.210455 | 0.001503 |
| SL000615 | Vasoactive Intestinal Peptide | Vasoactive Intestinal Peptide | P01282 | 7432 | VIP | -1.344235 | 0.001516 |
| SL009341 | Basigin | BASI | P35613 | 682 | BSG | 1.213948 | 0.001521 |
| SL013754 | Ras GTPase-activating protein 1 | RASA1 | P20936 | 5921 | RASA1 | -1.277993 | 0.001535 |
| SL003800 | Kallikrein-4 | Kallikrein 4 | Q9Y5K2 | 9622 | KLK4 | 1.25956 | 0.001688 |
| SL003169 | C-X-C motif chemokine 5 | ENA-78 | P42830 | 6374 | CXCL5 | -1.249801 | 0.001688 |
| SL007471 | Collectin-12 | COLEC12 | Q5KU26 | 81035 | COLEC12 | -1.271642 | 0.001709 |
| SL010449 | Carbonic anhydrase-related protein 10 | Carbonic Anhydrase X | Q9NS85 | 56934 | CA10 | -1.129273 | 0.001743 |
| SL003744 | Galectin-3 | Galectin-3 | P17931 | 3958 | LGALS3 | -1.237046 | 0.001743 |
| SL003200 | Angiopoietin-1 receptor, soluble | sTie-2 | Q02763 | 7010 | TEK | -1.249599 | 0.001743 |
| SL000613 | Urokinase-type plasminogen activator | uPA | P00749 | 5328 | PLAU | -1.278746 | 0.001767 |
| SL012248 | Focal adhesion kinase 1 | FAK1 | Q05397 | 5747 | PTK2 | -1.24482 | 0.001814 |
| SL003672 | Extracellular superoxide dismutase [Cu-Zn] | SOD3 | P08294 | 6649 | SOD3 | -1.31456 | 0.001936 |
| SL014289 | Killer cell immunoglobulin-like receptor 3DS1 | KI3S1 | Q14943 | 3813 | KIR3DS1 | -1.090573 | 0.001961 |
| SL006913 | Tyrosine-protein kinase Fyn | FYN | P06241 | 2534 | FYN | -1.390246 | 0.002031 |
| SL000525 | Matrilysin | MMP-7 | P09237 | 4316 | MMP7 | -1.280624 | 0.002089 |
| SL000039 | Interleukin-8 | IL-8 | P10145 | 3576 | CXCL8 | 1.23382 | 0.002089 |
| SL005167 | Galectin-8 | Galectin-8 | O00214 | 3964 | LGALS8 | -1.186974 | 0.002158 |
| SL004672 | Tumor necrosis factor receptor superfamily member 17 | BCMA | Q02223 | 608 | TNFRSF17 | -1.297649 | 0.00225 |
| SL006119 | Trefoil factor 3 | TFF3 | Q07654 | 7033 | TFF3 | 1.279347 | 0.002268 |
| SL004862 | Programmed cell death 1 ligand 2 | PD-L2 | Q9BQ51 | 80380 | PDCD1LG2 | -1.147324 | 0.002402 |
| SL011888 | SPARC-related modular calcium-binding protein 1 | SMOC1 | Q9H4F8 | 64093 | SMOC1 | -0.806498 | 0.002416 |
| SL000003 | Angiogenin | Angiogenin | P03950 | 283 | ANG | -1.346801 | 0.002448 |
| SL000138 | Heparin-binding EGF-like growth factor | HB-EGF | Q99075 | 1839 | HBEGF | -1.223472 | 0.002537 |
| SL010510 | Serine/threonine-protein kinase PAK 7 | PAK7 | Q9P286 | 57144 | PAK7 | 1.184393 | 0.002553 |
| SL004626 | A disintegrin and metalloproteinase with thrombospondin motifs 5 | ADAMTS-5 | Q9UNA0 | 11096 | ADAMTS5 | -1.128586 | 0.002616 |
| SL000557 | Protein kinase C zeta type | PKC-Z | Q05513 | 5590 | PRKCZ | 0.989222 | 0.002616 |
| SL007620 | Interleukin-12 receptor subunit beta-2 | IL-12 RB2 | Q99665 | 3595 | IL12RB2 | -1.175901 | 0.002626 |
| SL010517 | Serine/threonine-protein kinase 16 | STK16 | O75716 | 8576 | STK16 | 1.205305 | 0.002649 |
| SL003440 | Platelet-activating factor acetylhydrolase | PAFAH | Q13093 | 7941 | PLA2G7 | -1.287277 | 0.002729 |
| SL000633 | Tumor necrosis factor ligand superfamily member 6, soluble form | Fas ligand, soluble | P48023 | 356 | FASLG | -1.128289 | 0.002734 |
| SL005437 | Matrix extracellular phosphoglycoprotein | MEPE | Q9NQ76 | 56955 | MEPE | 1.191563 | 0.002749 |
| SL004137 | Ephrin type-A receptor 1 | EphA1 | P21709 | 2041 | EPHA1 | -1.226761 | 0.002757 |
| SL008331 | Proliferation-associated protein 2G4 | PA2G4 | Q9UQ80 | 5036 | PA2G4 | -1.378222 | 0.002781 |
| SL004346 | Interleukin-20 | IL-20 | Q9NYY1 | 50604 | IL20 | 1.193872 | 0.002801 |
| SL002722 | ADP-ribosyl cyclase/cyclic ADP-ribose hydrolase 1 | CD38 | P28907 | 952 | CD38 | -1.151517 | 0.002883 |
| SL007631 | Sclerostin | SOST | Q9BQB4 | 50964 | SOST | -1.39533 | 0.002891 |
| SL004737 | Tropomyosin alpha-1 chain | Tropomyosin 1 alpha chain | P09493 | 7168 | TPM1 | -1.113585 | 0.002903 |
| SL008190 | Spectrin alpha chain, non-erythrocytic 1 | SPTA2 | Q13813 | 6709 | SPTAN1 | 1.266239 | 0.002919 |
| SL008063 | Low molecular weight phosphotyrosine protein phosphatase | PPAC | P24666 | 52 | ACP1 | -1.15731 | 0.002927 |
| SL004515 | Peptidoglycan recognition protein 1 | PGRP-S | O75594 | 8993 | PGLYRP1 | -1.325579 | 0.002981 |
| SL013872 | Low-density lipoprotein receptor-related protein 1B | LRP1B | Q9NZR2 | 53353 | LRP1B | -1.392721 | 0.003042 |
| SL000276 | Apolipoprotein E | Apo E | P02649 | 348 | APOE | 0.915513 | 0.003043 |
| SL014088 | Fc receptor-like protein 3 | FCRL3 | Q96P31 | 115352 | FCRL3 | 1.141561 | 0.003125 |
| SL006544 | Transforming growth factor-beta-induced protein ig-h3 | BGH3 | Q15582 | 7045 | TGFBI | -1.362541 | 0.003125 |
| SL008837 | CD63 antigen | CD63 | P08962 | 967 | CD63 | -1.258263 | 0.003175 |
| SL007025 | Tyrosine-protein kinase JAK2 | JAK2 | O60674 | 3717 | JAK2 | -1.057055 | 0.00322 |
| SL002086 | Ficolin-3 | Ficolin-3 | O75636 | 8547 | FCN3 | -0.989533 | 0.003256 |
| SL010610 | C-type lectin domain family 4 member K | CLC4K | Q9UJ71 | 50489 | CD207 | 1.171506 | 0.003293 |
| SL004752 | Protein FAM107A | DRR1 | O95990 | 11170 | FAM107A | 1.096789 | 0.003332 |
| SL003220 | C3a anaphylatoxin des Arginine | C3adesArg | P01024 | 718 | C3 | -1.228782 | 0.003441 |
| SL004692 | Cytokine receptor-like factor 1:Cardiotrophin-like cytokine factor 1 Complex | CLF-1/CLC Complex | O75462 Q9UBD9 | 9244 23529 | CRLF1 CLCF1 | -1.244601 | 0.00346 |
| SL006993 | Mitogen-activated protein kinase 13 | MK13 | O15264 | 5603 | MAPK13 | -1.351955 | 0.003478 |
| SL009412 | Dickkopf-related protein 3 | DKK3 | Q9UBP4 | 27122 | DKK3 | 1.347367 | 0.003592 |
| SL004673 | Cathepsin S | Cathepsin S | P25774 | 1520 | CTSS | 1.271326 | 0.003607 |
| SL008810 | Neuronal growth regulator 1 | NEGR1 | Q7Z3B1 | 257194 | NEGR1 | -1.305841 | 0.00364 |
| SL005157 | CD209 antigen | DC-SIGN | Q9NNX6 | 30835 | CD209 | 1.294422 | 0.003735 |
| SL005206 | Natural cytotoxicity triggering receptor 2 | NKp44 | O95944 | 9436 | NCR2 | 1.154407 | 0.003809 |
| SL010619 | Tryptase gamma | TPSG1 | Q9NRR2 | 25823 | TPSG1 | 1.19207 | 0.003854 |
| SL014270 | CMRF35-like molecule 6 | CLM6 | Q08708 | 10871 | CD300C | 1.22541 | 0.003883 |
| SL004639 | NT-3 growth factor receptor | TrkC | Q16288 | 4916 | NTRK3 | -1.247375 | 0.003895 |
| SL006803 | Angiopoietin-related protein 3 | ANGL3 | Q9Y5C1 | 27329 | ANGPTL3 | 1.397168 | 0.003905 |
| SL004415 | Angiotensin-converting enzyme 2 | ACE2 | Q9BYF1 | 59272 | ACE2 | -1.287616 | 0.003905 |
| SL012188 | Epidermal growth factor receptor substrate 15-like 1 | EP15R | Q9UBC2 | 58513 | EPS15L1 | 1.273498 | 0.003988 |
| SL004925 | Anterior gradient protein 2 homolog | AGR2 | O95994 | 10551 | AGR2 | -1.170794 | 0.004047 |
| SL006480 | Trypsin-3 | TRY3 | P35030 | 5646 | PRSS3 | -1.195646 | 0.004087 |
| SL004360 | Neurotrophin-4 | Neurotrophin-5 | P34130 | 4909 | NTF4 | 1.228849 | 0.004087 |
| SL005263 | alpha-2-macroglobulin receptor-associated protein | RAP | P30533 | 4043 | LRPAP1 | -1.155904 | 0.00412 |
| SL001777 | Cystatin-C | Cystatin C | P01034 | 1471 | CST3 | 1.294841 | 0.004132 |
| SL004140 | Ephrin-A4 | Ephrin-A4 | P52798 | 1945 | EFNA4 | 1.264871 | 0.004207 |
| SL005234 | Growth/differentiation factor 9 | GDF-9 | O60383 | 2661 | GDF9 | 0.998155 | 0.004211 |
| SL004820 | Phosphoglycerate mutase 1 | Phosphoglycerate mutase 1 | P18669 | 5223 | PGAM1 | 1.07105 | 0.004259 |
| SL003178 | Intercellular adhesion molecule 3 | sICAM-3 | P32942 | 3385 | ICAM3 | 1.375824 | 0.004259 |
| SL004141 | Ephrin-A5 | Ephrin-A5 | P52803 | 1946 | EFNA5 | 1.225455 | 0.00436 |
| SL004015 | Carboxypeptidase B2 | TAFI | Q96IY4 | 1361 | CPB2 | -1.268772 | 0.004408 |
| SL004850 | Interleukin-17 receptor A | IL-17 sR | Q96F46 | 23765 | IL17RA | -1.194331 | 0.004649 |
| SL003655 | Transketolase | Transketolase | P29401 | 7086 | TKT | -1.260292 | 0.004674 |
| SL003643 | Glutathione S-transferase P | Glutathione S-transferase Pi | P09211 | 2950 | GSTP1 | 1.24153 | 0.004674 |
| SL002823 | L-Selectin | sL-Selectin | P14151 | 6402 | SELL | -1.286474 | 0.004674 |
| SL010489 | Calcium/calmodulin-dependent protein kinase type 1 | CAMK1 | Q14012 | 8536 | CAMK1 | -0.90838 | 0.004733 |
| SL010384 | Testican-1 | Testican-1 | Q08629 | 6695 | SPOCK1 | -1.29134 | 0.004788 |
| SL000655 | Keratin, type I cytoskeletal 18 | Keratin 18 | P05783 | 3875 | KRT18 | -1.320562 | 0.004826 |
| SL002731 | Tumor necrosis factor receptor superfamily member 6 | Fas, soluble | P25445 | 355 | FAS | -1.340844 | 0.004855 |
| SL007729 | Endoplasmic reticulum aminopeptidase 1 | ARTS1 | Q9NZ08 | 51752 | ERAP1 | 1.33598 | 0.004917 |
| SL004343 | Fms-related tyrosine kinase 3 ligand | Flt3 ligand | P49771 | 2323 | FLT3LG | -1.140893 | 0.004936 |
| SL007502 | Carbohydrate sulfotransferase 15 | ST4S6 | Q7LFX5 | 51363 | CHST15 | 1.284119 | 0.005013 |
| SL005233 | Tumor necrosis factor receptor superfamily member 27 | XEDAR | Q9HAV5 | 60401 | EDA2R | 1.231835 | 0.005096 |
| SL005223 | Interleukin-27 receptor subunit alpha | TCCR | Q6UWB1 | 9466 | IL27RA | -1.237631 | 0.005134 |
| SL003171 | Fibroblast growth factor 4 | FGF-4 | P08620 | 2249 | FGF4 | 1.223009 | 0.005257 |
| SL003862 | CD40 ligand | CD40 ligand, soluble | P29965 | 959 | CD40LG | -1.316647 | 0.005312 |
| SL016563 | Mitochondrial glutamate carrier 2 | GHC2 | Q9H1K4 | 83733 | SLC25A18 | 1.175341 | 0.005427 |
| SL001713 | Interleukin-17A | IL-17 | Q16552 | 3605 | IL17A | -1.114911 | 0.005427 |
| SL000539 | Glucose-6-phosphate isomerase | PHI | P06744 | 2821 | GPI | 1.234565 | 0.005427 |
| SL010520 | Aurora kinase B | AURKB | Q96GD4 | 9212 | AURKB | 1.13749 | 0.005477 |
| SL000605 | Ubiquitin+1, truncated mutation for UbB | Ubiquitin+1 | P62979 | 6233 | RPS27A | -1.095904 | 0.005597 |
| SL011535 | RNA-binding protein 39 | RBM39 | Q14498 | 9584 | RBM39 | 0.954392 | 0.005811 |
| SL017106 | Interferon alpha-10 | IFN10 | P01566 | 3446 | IFNA10 | -1.204837 | 0.005827 |
| SL014129 | Heparan-sulfate 6-O-sulfotransferase 1 | H6ST1 | O60243 | 9394 | HS6ST1 | 1.285423 | 0.00596 |
| SL003328 | Complement factor I | Factor I | P05156 | 3426 | CFI | -1.241154 | 0.00596 |
| SL016567 | Mitogen-activated protein kinase kinase kinase 7:TGF-beta-activated kinase 1 and MAP3K7-binding protein 1 fusion | TAK1-TAB1 | O43318 Q15750 | 6885 10454 | MAP3K7 TAB1 | -1.182573 | 0.005978 |
| SL002542 | GTPase KRas | K-ras | P01116 | 3845 | KRAS | -1.243815 | 0.005991 |
| SL011400 | Calcium/calmodulin-dependent 3',5'-cyclic nucleotide phosphodiesterase 1A | PDE1A | P54750 | 5136 | PDE1A | 1.306264 | 0.006076 |
| SL011202 | Alpha-soluble NSF attachment protein | SNAA | P54920 | 8775 | NAPA | -1.024646 | 0.006348 |
| SL004438 | Cystatin-M | Cystatin M | Q15828 | 1474 | CST6 | 1.306591 | 0.006621 |
| SL000493 | L-lactate dehydrogenase B chain | LDH-H 1 | P07195 | 3945 | LDHB | 1.225912 | 0.006703 |
| SL000573 | Serum amyloid P-component | SAP | P02743 | 325 | APCS | -1.294583 | 0.006829 |
| SL004861 | Interleukin-18 receptor accessory protein | IL-18 Rb | O95256 | 8807 | IL18RAP | -1.129671 | 0.006972 |
| SL003648 | Rab GDP dissociation inhibitor beta | Rab GDP dissociation inhibitor beta | P50395 | 2665 | GDI2 | -1.390658 | 0.007037 |
| SL010505 | Megakaryocyte-associated tyrosine-protein kinase | MATK | P42679 | 4145 | MATK | 1.33542 | 0.007102 |
| SL008623 | Contactin-2 | CNTN2 | Q02246 | 6900 | CNTN2 | -1.301762 | 0.007348 |
| SL014208 | Cytotoxic and regulatory T-cell molecule | CRTAM | O95727 | 56253 | CRTAM | -1.187223 | 0.007379 |
| SL017528 | Ribonuclease H1 | RNase H1 | O60930 | 246243 | RNASEH1 | -1.182883 | 0.007382 |
| SL005152 | Retinoic acid receptor responder protein 2 | TIG2 | Q99969 | 5919 | RARRES2 | 1.325974 | 0.007414 |
| SL002517 | Tumor necrosis factor | TNF-a | P01375 | 7124 | TNF | -1.139071 | 0.007501 |
| SL000437 | Haptoglobin | Haptoglobin, Mixed Type | P00738 | 3240 | HP | 1.097853 | 0.007665 |
| SL005235 | Homeobox protein NANOG | NANOG | Q9H9S0 | 79923 | NANOG | 1.218321 | 0.007689 |
| SL003520 | Calreticulin | calreticulin | P27797 | 811 | CALR | 1.176336 | 0.007752 |
| SL002922 | Intercellular adhesion molecule 1 | sICAM-1 | P05362 | 3383 | ICAM1 | 1.201908 | 0.007752 |
| SL005508 | Carbonic anhydrase 9 | Carbonic anhydrase 9 | Q16790 | 768 | CA9 | 1.329514 | 0.007864 |
| SL006921 | [Pyruvate dehydrogenase (acetyl-transferring)] kinase isozyme 1, mitochondrial | PDK1 | Q15118 | 5163 | PDK1 | -1.307632 | 0.008048 |
| SL004064 | Phospholipase A2 | GIB | P04054 | 5319 | PLA2G1B | 1.307304 | 0.008048 |
| SL005189 | Interleukin-7 receptor subunit alpha | IL-7 Ra | P16871 | 3575 | IL7R | 1.28723 | 0.008134 |
| SL007059 | Lymphocyte antigen 86 | LY86 | O95711 | 9450 | LY86 | 1.154023 | 0.008354 |
| SL014148 | Roundabout homolog 3 | ROBO3 | Q96MS0 | 64221 | ROBO3 | 1.292815 | 0.008465 |
| SL000011 | alpha-Fetoprotein | AFP | P02771 | 174 | AFP | -1.091542 | 0.008716 |
| SL015510 | Ubiquitin-like protein ISG15 | UCRP | P05161 | 9636 | ISG15 | -1.397174 | 0.008815 |
| SL006372 | Tyrosine-protein kinase Yes | YES | P07947 | 7525 | YES1 | 1.360987 | 0.008815 |
| SL000053 | Tissue-type plasminogen activator | tPA | P00750 | 5327 | PLAT | -1.112729 | 0.008817 |
| SL002071 | Gastrin-releasing peptide | Gastrin-releasing peptide | P07492 | 2922 | GRP | -1.107364 | 0.008889 |
| SL003951 | Brain-derived neurotrophic factor | BDNF | P23560 | 627 | BDNF | 1.070336 | 0.00905 |
| SL014048 | Collagen alpha-1(XXIII) chain | CONA1 | Q86Y22 | 91522 | COL23A1 | 1.234115 | 0.009077 |
| SL004159 | Tumor necrosis factor receptor superfamily member 10D | TRAIL R4 | Q9UBN6 | 8793 | TNFRSF10D | 1.304321 | 0.009446 |
| SL004580 | Macrophage scavenger receptor types I and II | Macrophage scavenger receptor | P21757 | 4481 | MSR1 | -1.12461 | 0.009655 |
| SL004148 | Interleukin-12 receptor subunit beta-1 | IL-12 Rb1 | P42701 | 3594 | IL12RB1 | 1.28771 | 0.009655 |
| SL003183 | C-X-C motif chemokine 10 | IP-10 | P02778 | 3627 | CXCL10 | -1.39322 | 0.00972 |
| SL004726 | CD97 antigen | CD97 | P48960 | 976 | CD97 | 1.165813 | 0.009849 |
| SL011509 | Peptide YY | PYY | P10082 | 5697 | PYY | -1.251143 | 0.009983 |
| SL004649 | Tumor necrosis factor ligand superfamily member 4 | OX40 Ligand | P23510 | 7292 | TNFSF4 | 1.214509 | 0.010052 |
| SL004643 | Angiopoietin-4 | Angiopoietin-4 | Q9Y264 | 51378 | ANGPT4 | 1.219719 | 0.010241 |
| SL010471 | Testican-2 | Testican-2 | Q92563 | 9806 | SPOCK2 | -1.235233 | 0.010647 |
| SL006527 | EGF-containing fibulin-like extracellular matrix protein 1 | FBLN3 | Q12805 | 2202 | EFEMP1 | -1.208702 | 0.011037 |
| SL006189 | Kinesin-like protein KIF23 | KIF23 | Q02241 | 9493 | KIF23 | -0.850528 | 0.011049 |
| SL000136 | Amphiregulin | AREG | P15514 | 374 | AREG | -0.85041 | 0.011049 |
| SL010458 | Endothelial cell-specific molecule 1 | Endocan | Q9NQ30 | 11082 | ESM1 | -0.619753 | 0.011068 |
| SL003310 | Vascular endothelial growth factor A, isoform 121 | VEGF121 | P15692 | 7422 | VEGFA | -1.251549 | 0.011302 |
| SL007122 | Insulin-degrading enzyme | IDE | P14735 | 3416 | IDE | -0.791806 | 0.011394 |
| SL018256 | Choline/ethanolamine kinase | CHKB | Q9Y259 | 1120 | CHKB | -1.253746 | 0.012135 |
| SL004690 | WNT1-inducible-signaling pathway protein 3 | WISP-3 | O95389 | 8838 | WISP3 | -1.145508 | 0.012135 |
| SL000300 | Beta-endorphin | b-Endorphin | P01189 | 5443 | POMC | 1.155137 | 0.012581 |
| SL017328 | Myosin-binding protein C, slow-type | MYPC1 | Q00872 | 4604 | MYBPC1 | 0.734203 | 0.012652 |
| SL006919 | Ribosomal protein S6 kinase alpha-5 | RSK-like protein kinase | O75582 | 9252 | RPS6KA5 | -1.212273 | 0.012801 |
| SL004482 | Endoglin | Endoglin | P17813 | 2022 | ENG | -1.345731 | 0.013306 |
| SL003060 | Fibroblast growth factor receptor 1 | bFGF-R | P11362 | 2260 | FGFR1 | -1.3485 | 0.013373 |
| SL013988 | Carbohydrate sulfotransferase 2 | CHST2 | Q9Y4C5 | 9435 | CHST2 | 0.778081 | 0.013623 |
| SL020172 | Relaxin receptor 1 | RXFP1 | Q9HBX9 | NaN | RXFP1 | -1.167665 | 0.013836 |
| SL004331 | Ciliary neurotrophic factor | CNTF | P26441 | 1270 | CNTF | -1.09865 | 0.014086 |
| SL004156 | Tumor necrosis factor receptor superfamily member 10A | TRAIL R1 | O00220 | 8797 | TNFRSF10A | 1.26577 | 0.014137 |
| SL000055 | Cadherin-1 | Cadherin E | P12830 | 999 | CDH1 | -1.376261 | 0.014476 |
| SL004351 | Interleukin-25 | IL-17E | Q9H293 | 64806 | IL25 | 1.228743 | 0.01477 |
| SL000560 | P-selectin | P-Selectin | P16109 | 6403 | SELP | -1.204422 | 0.014924 |
| SL010493 | Calcium/calmodulin-dependent protein kinase type II subunit delta | CAMK2D | Q13557 | 817 | CAMK2D | -0.806821 | 0.014933 |
| SL005675 | ATP synthase subunit beta, mitochondrial | ATP synthase beta chain | P06576 | 506 | ATP5B | -0.918187 | 0.01525 |
| SL005187 | Interleukin-3 receptor subunit alpha | IL-3 Ra | P26951 | 3563 | IL3RA | -0.910126 | 0.015544 |
| SL010373 | Immunoglobulin alpha Fc receptor | FCAR | P24071 | 2204 | FCAR | 1.200512 | 0.015684 |
| SL004823 | Peptidyl-prolyl cis-trans isomerase A | Cyclophilin A | P62937 | 5478 | PPIA | -0.823353 | 0.015684 |
| SL004605 | 40S ribosomal protein SA | 40S ribosomal protein SA | P08865 | 3921 | RPSA | 1.127105 | 0.015684 |
| SL011499 | Pancreatic hormone | PH | P01298 | 5539 | PPY | 1.206162 | 0.016078 |
| SL011448 | Tumor necrosis factor receptor superfamily member 4 | TNR4 | P43489 | 7293 | TNFRSF4 | 1.220012 | 0.016078 |
| SL001797 | Kallikrein-6 | Kallikrein 6 | Q92876 | 5653 | KLK6 | 1.17163 | 0.016156 |
| SL004396 | Signal transducer and activator of transcription 1-alpha/beta | STAT1 | P42224 | 6772 | STAT1 | -1.099871 | 0.016198 |
| SL004908 | Tropomyosin beta chain | Tropomyosin 2 | P07951 | 7169 | TPM2 | 1.257403 | 0.016417 |
| SL014268 | OX-2 membrane glycoprotein | OX2G | P41217 | 4345 | CD200 | 1.191158 | 0.016974 |
| SL001796 | Myeloperoxidase | Myeloperoxidase | P05164 | 4353 | MPO | 1.371002 | 0.016974 |
| SL001905 | Mesothelin | Mesothelin | Q13421 | 10232 | MSLN | -1.248297 | 0.017003 |
| SL008486 | Galectin-9 | LEG9 | O00182 | 3965 | LGALS9 | -1.144222 | 0.017924 |
| SL010519 | Tyrosine-protein kinase ZAP-70 | ZAP70 | P43403 | 7535 | ZAP70 | 1.259471 | 0.018275 |
| SL016554 | High affinity cGMP-specific 3',5'-cyclic phosphodiesterase 9A | PDE9A | O76083 | 5152 | PDE9A | 1.143444 | 0.018347 |
| SL000403 | Endostatin | Endostatin | P39060 | 80781 | COL18A1 | -1.272374 | 0.018467 |
| SL003080 | Macrophage migration inhibitory factor | MIF | P14174 | 4282 | MIF | -1.303525 | 0.018501 |
| SL006397 | Neuropilin-1 | NRP1 | O14786 | 8829 | NRP1 | 1.130219 | 0.018783 |
| SL004940 | Tyrosine-protein phosphatase non-receptor type 6 | PTP-1C | P29350 | 5777 | PTPN6 | -1.354642 | 0.018965 |
| SL011708 | cAMP-regulated phosphoprotein 19 | ARP19 | P56211 | 10776 | ARPP19 | -1.343556 | 0.018991 |
| SL000592 | Metalloproteinase inhibitor 2 | TIMP-2 | P16035 | 7077 | TIMP2 | -1.324503 | 0.018991 |
| SL012822 | Brain-specific serine protease 4 | BSSP4 | Q9GZN4 | 64063 | PRSS22 | 1.255896 | 0.019025 |
| SL014071 | Leucine-rich repeat transmembrane protein FLRT1 | FLRT1 | Q9NZU1 | 23769 | FLRT1 | -1.235872 | 0.01909 |
| SL005266 | Alpha-synuclein | a-Synuclein | P37840 | 6622 | SNCA | -1.248745 | 0.019268 |
| SL019096 | E3 SUMO-protein ligase PIAS4 | PIAS4 | Q8N2W9 | 51588 | PIAS4 | 1.387652 | 0.019548 |
| SL008176 | Proteasome activator complex subunit 1 | PSME1 | Q06323 | 5720 | PSME1 | -0.969765 | 0.019548 |
| SL008590 | Olfactomedin-4 | Olfactomedin-4 | Q6UX06 | 10562 | OLFM4 | 1.191015 | 0.019674 |
| SL014108 | Leucine-rich repeat transmembrane neuronal protein 3 | LRRT3 | Q86VH5 | 347731 | LRRTM3 | 1.236649 | 0.019925 |
| SL006918 | Mitogen-activated protein kinase 1 | MK01 | P28482 | 5594 | MAPK1 | -1.273166 | 0.020249 |
| SL007266 | 26S proteasome non-ATPase regulatory subunit 7 | PSD7 | P51665 | 5713 | PSMD7 | 1.150245 | 0.020754 |
| SL002075 | Interferon alpha-2 | IFN-aA | P01563 | 3440 | IFNA2 | -1.31533 | 0.020811 |
| SL004760 | Platelet-activating factor acetylhydrolase IB subunit beta | PAFAH beta subunit | P68402 | 5049 | PAFAH1B2 | -0.939466 | 0.021314 |
| SL000409 | Mitogen-activated protein kinase 3 | ERK-1 | P27361 | 5595 | MAPK3 | -1.153159 | 0.021467 |
| SL007121 | Cathepsin E | CATE | P14091 | 1510 | CTSE | 1.261839 | 0.022112 |
| SL007003 | Desert hedgehog protein N-product | DHH | O43323 | 50846 | DHH | -0.669764 | 0.022496 |
| SL000324 | Complement component C8 | C8 | P07357,P07358,P07360 | 731 732 733 | C8A C8B C8G | 1.190898 | 0.022619 |
| SL016551 | Casein kinase II 2-alpha':2-beta heterotetramer | CK2-A2:B | P19784 P67870 | 1459 1460 | CSNK2A2 CSNK2B | -1.353619 | 0.023192 |
| SL000456 | Complement C3b, inactivated | iC3b | P01024 | 718 | C3 | 0.921863 | 0.023192 |
| SL001802 | Interferon gamma | IFN-g | P01579 | 3458 | IFNG | -1.20225 | 0.02467 |
| SL005685 | Elongation factor 1-beta | EF-1-beta | P24534 | 1933 | EEF1B2 | -0.861758 | 0.025186 |
| SL004306 | SUMO-conjugating enzyme UBC9 | UBC9 | P63279 | 7329 | UBE2I | -0.801087 | 0.02519 |
| SL011630 | Seizure 6-like protein 2 | SE6L2 | Q6UXD5 | 26470 | SEZ6L2 | -1.286629 | 0.026001 |
| SL011709 | Pyridoxal phosphate phosphatase | PLPP | Q96GD0 | 57026 | PDXP | -0.900275 | 0.026411 |
| SL003324 | Coagulation factor Xa | Coagulation Factor Xa | P00742 | 2159 | F10 | -0.974759 | 0.026411 |
| SL003186 | Lymphotactin | Lymphotactin | P47992 | 6375 | XCL1 | 1.288595 | 0.026461 |
| SL007327 | Oxidized low-density lipoprotein receptor 1 | OLR1 | P78380 | 4973 | OLR1 | -0.543742 | 0.026581 |
| SL004899 | Heat shock cognate 71 kDa protein | HSP70 protein 8 | P11142 | 3312 | HSPA8 | 0.72244 | 0.026779 |
| SL000158 | Glutamate carboxypeptidase 2 | PSMA | Q04609 | 2346 | FOLH1 | -1.198895 | 0.02687 |
| SL016928 | SLAM family member 7 | SLAF7 | Q9NQ25 | 57823 | SLAMF7 | -1.28358 | 0.026892 |
| SL004920 | Cofilin-1 | Cofilin-1 | P23528 | 1072 | CFL1 | -1.006114 | 0.027101 |
| SL003307 | Cytokine receptor common subunit gamma | IL-2 sRg | P31785 | 3561 | IL2RG | -0.694143 | 0.02717 |
| SL018887 | Quinone oxidoreductase-like protein 1 | QORL1 | O95825 | 9946 | CRYZL1 | 1.215228 | 0.028768 |
| SL008703 | Carboxypeptidase E | CBPE | P16870 | 1363 | CPE | -1.141132 | 0.030051 |
| SL003863 | Kallikrein-5 | kallikrein 5 | Q9Y337 | 25818 | KLK5 | -1.126254 | 0.03023 |
| SL003197 | C-C motif chemokine 25 | TECK | O15444 | 6370 | CCL25 | -0.576829 | 0.031201 |
| SL010502 | Mitogen-activated protein kinase 8 | MK08 | P45983 | 5599 | MAPK8 | -1.389215 | 0.031312 |
| SL003184 | Leptin receptor | sLeptin R | P48357 | 3953 | LEPR | -0.74497 | 0.031427 |
| SL010496 | Cyclin-dependent kinase 5:Cyclin-dependent kinase 5 activator 1 complex | CDK5/p35 | Q00535 Q15078 | 1020 8851 | CDK5 CDK5R1 | 1.174456 | 0.031906 |
| SL003173 | Growth-regulated alpha protein | Gro-a | P09341 | 2919 | CXCL1 | -1.228528 | 0.032134 |
| SL004837 | Inhibin beta A chain:Inhibin beta B chain heterodimer | Activin AB | P08476 P09529 | 3624 3625 | INHBA INHBB | 1.188413 | 0.033177 |
| SL004648 | Tumor necrosis factor ligand superfamily member 14 | LIGHT | O43557 | 8740 | TNFSF14 | -1.155588 | 0.033177 |
| SL004119 | Epithelial discoidin domain-containing receptor 1 | discoidin domain receptor 1 | Q08345 | 780 | DDR1 | -1.233728 | 0.033458 |
| SL009988 | Disintegrin and metalloproteinase domain-containing protein 12 | ADAM12 | O43184 | 8038 | ADAM12 | 1.288409 | 0.033992 |
| SL002702 | Serine/threonine-protein kinase pim-1 | PIM1 | P11309 | 5292 | PIM1 | 1.163988 | 0.034042 |
| SL004915 | Chloride intracellular channel protein 1 | NCC27 | O00299 | 1192 | CLIC1 | -1.301397 | 0.034442 |
| SL002803 | Ubiquitin carboxyl-terminal hydrolase isozyme L1 | PGP9.5 | P09936 | 7345 | UCHL1 | -1.293993 | 0.034624 |
| SL003349 | Hypoxia-inducible factor 1-alpha | HIF-1a | Q16665 | 3091 | HIF1A | -1.161924 | 0.035389 |
| SL000481 | Interleukin-5 | IL-5 | P05113 | 3567 | IL5 | -1.170707 | 0.035625 |
| SL010499 | Tyrosine-protein kinase HCK | HCK | P08631 | 3055 | HCK | 1.159643 | 0.035856 |
| SL001731 | Gamma-enolase | NSE | P09104 | 2026 | ENO2 | -1.323375 | 0.035904 |
| SL004652 | Wnt inhibitory factor 1 | WIF-1 | Q9Y5W5 | 11197 | WIF1 | -1.243648 | 0.035943 |
| SL005730 | cGMP-dependent 3',5'-cyclic phosphodiesterase | cGMP-stimulated PDE | O00408 | 5138 | PDE2A | -1.232297 | 0.036234 |
| SL000038 | C-C motif chemokine 2 | MCP-1 | P13500 | 6347 | CCL2 | -1.128496 | 0.036503 |
| SL008085 | 3-hydroxyisobutyrate dehydrogenase, mitochondrial | 3HIDH | P31937 | 11112 | HIBADH | -1.12131 | 0.037595 |
| SL000551 | Protein kinase C alpha type | PKC-A | P17252 | 5578 | PRKCA | -1.112496 | 0.038388 |
| SL017128 | Leucine-rich repeat serine/threonine-protein kinase 2 | LRRK2 | Q5S007 | NaN | LRRK2 | -1.117585 | 0.038514 |
| SL010491 | Calcium/calmodulin-dependent protein kinase type II subunit alpha | CAMK2A | Q9UQM7 | 815 | CAMK2A | -0.423153 | 0.040579 |
| SL012740 | A disintegrin and metalloproteinase with thrombospondin motifs 15 | ATS15 | Q8TE58 | 170689 | ADAMTS15 | 1.165469 | 0.040593 |
| SL004097 | Mothers against decapentaplegic homolog 3 | SMAD3 | P84022 | 4088 | SMAD3 | -1.021149 | 0.040628 |
| SL005160 | Endothelial cell-selective adhesion molecule | ESAM | Q96AP7 | 90952 | ESAM | 1.232894 | 0.041073 |
| SL003176 | C-C motif chemokine 1 | I-309 | P22362 | 6346 | CCL1 | 1.189481 | 0.041399 |
| SL007281 | Mitogen-activated protein kinase 12 | MK12 | P53778 | 6300 | MAPK12 | 1.169852 | 0.041951 |
| SL000137 | Betacellulin | BTC | P35070 | 685 | BTC | 1.206741 | 0.04269 |
| SL003703 | Bcl-2-related protein A1 | BFL1 | Q16548 | 597 | BCL2A1 | -1.152971 | 0.042879 |
| SL008588 | SLAM family member 5 | SLAF5 | Q9UIB8 | 8832 | CD84 | 1.130486 | 0.043127 |
| SL000313 | C3a anaphylatoxin | C3a | P01024 | 718 | C3 | -1.003452 | 0.043405 |
| SL008157 | Ubiquitin-conjugating enzyme E2 L3 | UB2L3 | P68036 | 7332 | UBE2L3 | -0.550665 | 0.045174 |
| SL007871 | UMP-CMP kinase | Cytidylate kinase | P30085 | 51727 | CMPK1 | -1.155241 | 0.04567 |
| SL011549 | AT-rich interactive domain-containing protein 3A | ARI3A | Q99856 | 1820 | ARID3A | -0.464304 | 0.046789 |
| SL003334 | Endothelial monocyte-activating polypeptide 2 | EMAP-2 | Q12904 | 9255 | AIMP1 | -1.184197 | 0.047594 |
| SL004334 | Fibroblast growth factor 16 | FGF-16 | O43320 | 8823 | FGF16 | -0.85375 | 0.047721 |
| SL007070 | Low-density lipoprotein receptor | LDLR | P01130 | 3949 | LDLR | -1.12955 | 0.047726 |
| SL003658 | Aflatoxin B1 aldehyde reductase member 2 | Aflatoxin B1 aldehyde reductase | O43488 | 8574 | AKR7A2 | -1.269904 | 0.04884 |
| SL003710 | Caspase-2 | Caspase-2 | P42575 | 835 | CASP2 | -1.144097 | 0.049337 |
| SL005194 | Junctional adhesion molecule C | JAM-C | Q9BX67 | 83700 | JAM3 | -0.593925 | 0.049462 |
| SL001945 | E-selectin | sE-Selectin | P16581 | 6401 | SELE | -1.368509 | 0.050677 |
| SL013989 | R-spondin-2 | RSPO2 | Q6UXX9 | 340419 | RSPO2 | 0.651041 | 0.052397 |
| SL008177 | Complement component 1 Q subcomponent-binding protein, mitochondrial | C1QBP | Q07021 | 708 | C1QBP | 1.123021 | 0.052503 |
| SL007181 | Non-receptor tyrosine-protein kinase TYK2 | TYK2 | P29597 | 7297 | TYK2 | 1.192076 | 0.052503 |
| SL010369 | Carbonic anhydrase 4 | Carbonic Anhydrase IV | P22748 | 762 | CA4 | 1.272274 | 0.05289 |
| SL009210 | Pro-opiomelanocortin | Corticotropin-lipotropin | P01189 | 5443 | POMC | -1.188988 | 0.05289 |
| SL000360 | Coagulation Factor X | Coagulation Factor X | P00742 | 2159 | F10 | -1.316527 | 0.054146 |
| SL014229 | CD83 antigen | CD83 | Q01151 | 9308 | CD83 | -1.242773 | 0.054372 |
| SL011808 | Copine-1 | CPNE1 | Q99829 | 8904 | CPNE1 | -0.922541 | 0.056327 |
| SL000076 | Cyclin-dependent kinase inhibitor 1B | p27Kip1 | P46527 | 1027 | CDKN1B | -0.963318 | 0.057724 |
| SL011768 | NudC domain-containing protein 3 | NUDC3 | Q8IVD9 | 23386 | NUDCD3 | 1.156374 | 0.05824 |
| SL008644 | ADP-ribosyl cyclase/cyclic ADP-ribose hydrolase 2 | BST1 | Q10588 | 683 | BST1 | -1.03548 | 0.058904 |
| SL006917 | Tyrosine-protein kinase Lyn | LYN | P07948 | 4067 | LYN | -1.241765 | 0.058955 |
| SL004812 | Triosephosphate isomerase | Triosephosphate isomerase | P60174 | 7167 | TPI1 | -1.118764 | 0.059133 |
| SL004348 | Interferon lambda-1 | IFN-lambda 1 | Q8IU54 | 282618 | IFNL1 | -0.325832 | 0.060305 |
| SL008008 | Arginase-1 | ARGI1 | P05089 | 383 | ARG1 | -0.611713 | 0.061218 |
| SL005212 | Prolactin receptor | Prolactin Receptor | P16471 | 5618 | PRLR | 1.230497 | 0.06224 |
| SL011772 | Pescadillo homolog | PESC | O00541 | 23481 | PES1 | 0.767868 | 0.062839 |
| SL003170 | C-C motif chemokine 26 | Eotaxin-3 | Q9Y258 | 10344 | CCL26 | 1.223243 | 0.063021 |
| SL000527 | Matrix metalloproteinase-9 | MMP-9 | P14780 | 4318 | MMP9 | -0.831552 | 0.063363 |
| SL001997 | Interleukin-1 receptor type 1 | IL-1 sRI | P14778 | 3554 | IL1R1 | 1.184249 | 0.063543 |
| SL004068 | Granzyme B | Granzyme B | P10144 | 3002 | GZMB | -0.460541 | 0.064666 |
| SL000020 | Apolipoprotein B | Apo B | P04114 | 338 | APOB | 0.404078 | 0.064964 |
| SL000057 | Thymidine kinase, cytosolic | Thymidine kinase | P04183 | 7083 | TK1 | -0.421648 | 0.066196 |
| SL004765 | MAP kinase-activated protein kinase 3 | MAPKAPK3 | Q16644 | 7867 | MAPKAPK3 | -0.745286 | 0.067239 |
| SL000470 | Interleukin-11 | IL-11 | P20809 | 3589 | IL11 | 1.265967 | 0.067287 |
| SL000556 | Protein kinase C gamma type | PKC-G | P05129 | 5582 | PRKCG | 1.157428 | 0.067553 |
| SL003041 | Platelet factor 4 | PF-4 | P02776 | 5196 | PF4 | -1.3764 | 0.067622 |
| SL005725 | GRB2-related adapter protein 2 | GRB2-related adapter protein 2 | O75791 | 9402 | GRAP2 | 1.141135 | 0.06804 |
| SL003104 | C-C motif chemokine 24 | Eotaxin-2 | O00175 | 6369 | CCL24 | 1.133481 | 0.068268 |
| SL004589 | Azurocidin | Azurocidin | P20160 | 566 | AZU1 | -0.350306 | 0.069883 |
| SL003542 | Histone-lysine N-methyltransferase EHMT2 | NG36 | Q96KQ7 | 10919 | EHMT2 | 0.612066 | 0.070444 |
| SL008071 | 14-3-3 protein theta | 14-3-3 protein theta | P27348 | 10971 | YWHAQ | -1.213216 | 0.073512 |
| SL000441 | Hepatocyte growth factor | HGF | P14210 | 3082 | HGF | 0.926213 | 0.073847 |
| SL010349 | Fibronectin Fragment 3 | FN1.3 | P02751 | 2335 | FN1 | -1.330249 | 0.075236 |
| SL006713 | Collectin-11 | Collectin Kidney 1 | Q9BWP8 | 78989 | COLEC11 | -1.306188 | 0.075682 |
| SL016553 | cGMP-inhibited 3',5'-cyclic phosphodiesterase A | PDE3A | Q14432 | 5139 | PDE3A | -1.219444 | 0.076884 |
| SL001717 | Interleukin-10 | IL-10 | P22301 | 3586 | IL10 | 1.146616 | 0.076884 |
| SL004827 | Protein S100-A6 | S100A6 | P06703 | 6277 | S100A6 | -0.523577 | 0.077466 |
| SL000530 | Oncostatin-M | OSM | P13725 | 5008 | OSM | -0.679212 | 0.077516 |
| SL010466 | Membrane frizzled-related protein | MFRP | Q9BY79 | 83552 | MFRP | 1.296547 | 0.078244 |
| SL007195 | CD70 antigen | CD70 | P32970 | 970 | CD70 | -1.10414 | 0.079179 |
| SL006914 | Glucocorticoid receptor | Glucocorticoid receptor | P04150 | 2908 | NR3C1 | -1.329733 | 0.080699 |
| SL008378 | Eukaryotic translation initiation factor 4E-binding protein 2 | 4EBP2 | Q13542 | 1979 | EIF4EBP2 | 1.195116 | 0.083052 |
| SL014308 | Fibroblast growth factor 8 isoform A | FGF-8A | P55075 | 2253 | FGF8 | -0.49952 | 0.084345 |
| SL004939 | Tyrosine-protein phosphatase non-receptor type 1 | PTP-1B | P18031 | 5770 | PTPN1 | -1.182257 | 0.084345 |
| SL000414 | Complement factor B | Factor B | P00751 | 629 | CFB | 1.234296 | 0.084345 |
| SL004875 | Interleukin-1 receptor-like 2 | IL-1Rrp2 | Q9HB29 | 8808 | IL1RL2 | -1.100587 | 0.087658 |
| SL018938 | Small ubiquitin-related modifier 3 | SUMO3 | P55854 | 6613 | SUMO3 | -1.378414 | 0.087904 |
| SL006998 | 3-phosphoinositide-dependent protein kinase 1 | PDPK1 | O15530 | 5170 | PDPK1 | -0.916709 | 0.088573 |
| SL007869 | Peptidyl-prolyl cis-trans isomerase B | PPIB | P23284 | 5479 | PPIB | 1.28528 | 0.089305 |
| SL016550 | Casein kinase II 2-alpha:2-beta heterotetramer | CK2-A1:B | P68400 P67870 | 1457 1460 | CSNK2A1 CSNK2B | -0.640586 | 0.08999 |
| SL010381 | Dickkopf-like protein 1 | Soggy-1 | Q9UK85 | 27120 | DKKL1 | -1.215561 | 0.08999 |
| SL004253 | Estradiol 17-beta-dehydrogenase 1 | 17-beta-HSD 1 | P14061 | 3292 | HSD17B1 | -0.497587 | 0.08999 |
| SL008113 | ATP synthase subunit O, mitochondrial | ATPO | P48047 | 539 | ATP5O | -1.334201 | 0.090897 |
| SL013969 | Kynureninase | KYNU | Q16719 | 8942 | KYNU | -1.214014 | 0.091308 |
| SL011049 | Mannan-binding lectin serine protease 1 | MASP3 | P48740 | 5648 | MASP1 | -1.186493 | 0.091308 |
| SL006522 | Galectin-3-binding protein | LG3BP | Q08380 | 3959 | LGALS3BP | -1.307837 | 0.091308 |
| SL005430 | Secreted and transmembrane protein 1 | SECTM1 | Q8WVN6 | 6398 | SECTM1 | 1.157447 | 0.091308 |
| SL004133 | Bone morphogenetic protein receptor type-2 | BMP RII | Q13873 | 659 | BMPR2 | -1.156868 | 0.091308 |
| SL003872 | Interleukin-6 receptor subunit beta | gp130, soluble | P40189 | 3572 | IL6ST | -1.302705 | 0.091308 |
| SL003848 | Annexin A5 | Annexin V | P08758 | 308 | ANXA5 | -1.282633 | 0.091326 |
| SL005214 | Semaphorin-6A | Semaphorin-6A | Q9H2E6 | 57556 | SEMA6A | 0.692188 | 0.091822 |
| SL011588 | 40S ribosomal protein S3a | RS3A | P61247 | 6189 | RPS3A | -0.798108 | 0.092066 |
| SL004299 | Baculoviral IAP repeat-containing protein 7 Isoform beta | Livin B | Q96CA5 | 79444 | BIRC7 | 1.194919 | 0.092066 |
| SL003990 | Fibroblast growth factor receptor 2 | FGFR-2 | P21802 | 2263 | FGFR2 | -1.11089 | 0.092083 |
| SL007280 | Dipeptidyl peptidase 1 | CATC | P53634 | 1075 | CTSC | 1.133051 | 0.09224 |
| SL004901 | Protein disulfide-isomerase | Protein disulfide-isomerase | P07237 | 5034 | P4HB | 1.172252 | 0.09224 |
| SL004872 | Tumor necrosis factor receptor superfamily member EDAR | EDAR | Q9UNE0 | 10913 | EDAR | -1.267306 | 0.093099 |
| SL004768 | Allograft inflammatory factor 1 | AIF1 | P55008 | 199 | AIF1 | -0.634382 | 0.093119 |
| SL004714 | Leukemia inhibitory factor receptor | LIF sR | P42702 | 3977 | LIFR | -1.314549 | 0.094983 |
| SL004637 | Macrophage-stimulating protein receptor | MSP R | Q04912 | 4486 | MST1R | 1.198256 | 0.095143 |
| SL011628 | Drebrin-like protein | DBNL | Q9UJU6 | 28988 | DBNL | 1.271979 | 0.095378 |
| SL003685 | Nicotinamide phosphoribosyltransferase | PBEF | P43490 | 10135 | NAMPT | 1.247406 | 0.095378 |
| SL000045 | Insulin-like growth factor-binding protein 3 | IGFBP-3 | P17936 | 3486 | IGFBP3 | 0.69611 | 0.095539 |
| SL011533 | Small glutamine-rich tetratricopeptide repeat-containing protein alpha | SGTA | O43765 | 6449 | SGTA | -0.807271 | 0.096544 |
| SL003700 | Bcl2-associated agonist of cell death | BAD | Q92934 | 572 | BAD | -0.709985 | 0.099904 |
| SL003679 | Cation-independent mannose-6-phosphate receptor | IGF-II receptor | P11717 | 3482 | IGF2R | -1.27037 | 0.100018 |
| SL014009 | Hepatoma-derived growth factor-related protein 2 | HDGR2 | Q7Z4V5 | 84717 | HDGFRP2 | 1.279145 | 0.100863 |
| SL010463 | Glypican-2 | GPC2 | Q8N158 | 221914 | GPC2 | 0.631962 | 0.100863 |
| SL008760 | SLIT and NTRK-like protein 1 | SLIK1 | Q96PX8 | 114798 | SLITRK1 | 1.256179 | 0.102358 |
| SL000460 | Immunoglobulin D | IgD | P01880 | 3495 50802 3535 | IGHD IGK@ IGL@ | -1.308742 | 0.10324 |
| SL007033 | Latent-transforming growth factor beta-binding protein 4 | LTBP4 | Q8N2S1 | 8425 | LTBP4 | -0.942858 | 0.103963 |
| SL000133 | C-C motif chemokine 20 | MIP-3a | P78556 | 6364 | CCL20 | -0.655825 | 0.105248 |
| SL009202 | Junctional adhesion molecule-like | JAML1 | Q86YT9 | 120425 | AMICA1 | -1.177612 | 0.105717 |
| SL000449 | DnaJ homolog subfamily B member 1 | HSP 40 | P25685 | 3337 | DNAJB1 | -0.57518 | 0.106835 |
| SL006922 | DNA repair protein RAD51 homolog 1 | RAD51 | Q06609 | 5888 | RAD51 | 1.210232 | 0.111748 |
| SL004477 | Protein S100-A9 | calgranulin B | P06702 | 6280 | S100A9 | 1.021349 | 0.112901 |
| SL011510 | Somatostatin-28 | Somatostatin-28 | P61278 | 6750 | SST | 0.77103 | 0.113539 |
| SL010524 | Serine/threonine-protein kinase WNK3 | WNK3 | Q9BYP7 | 65267 | WNK3 | -0.728188 | 0.115269 |
| SL012754 | Acid sphingomyelinase-like phosphodiesterase 3a | ASM3A | Q92484 | 10924 | SMPDL3A | -0.842466 | 0.115708 |
| SL004750 | ATP-dependent RNA helicase DDX19B | DEAD-box protein 19B | Q9UMR2 | 11269 | DDX19B | -0.568744 | 0.116068 |
| SL011406 | High affinity cAMP-specific 3',5'-cyclic phosphodiesterase 7A | PDE7A | Q13946 | 5150 | PDE7A | 1.189933 | 0.116124 |
| SL000638 | Cadherin-2 | Cadherin-2 | P19022 | 1000 | CDH2 | 1.18541 | 0.116124 |
| SL008414 | Ephrin type-B receptor 6 | EphB6 | O15197 | 2051 | EPHB6 | 1.189866 | 0.11645 |
| SL004871 | Tumor necrosis factor receptor superfamily member 21 | DR6 | O75509 | 27242 | TNFRSF21 | -1.274173 | 0.116907 |
| SL007261 | Methionine aminopeptidase 2 | AMPM2 | P50579 | 10988 | METAP2 | -0.586995 | 0.117162 |
| SL005229 | NKG2D ligand 3 | ULBP-3 | Q9BZM4 | 79465 | ULBP3 | -1.118904 | 0.118055 |
| SL004359 | Neurotrophin-3 | Neurotrophin-3 | P20783 | 4908 | NTF3 | -1.204585 | 0.118055 |
| SL010492 | Calcium/calmodulin-dependent protein kinase type II subunit beta | CAMK2B | Q13554 | 816 | CAMK2B | -0.377498 | 0.119943 |
| SL005493 | Serum paraoxonase/arylesterase 1 | paraoxonase 1 | P27169 | 5444 | PON1 | -0.897161 | 0.120281 |
| SL013928 | Peptidyl-prolyl cis-trans isomerase E | PPIE | Q9UNP9 | 10450 | PPIE | -0.498512 | 0.120572 |
| SL004869 | Carbonic anhydrase 13 | Carbonic anhydrase XIII | Q8N1Q1 | 377677 | CA13 | -0.802434 | 0.121102 |
| SL002695 | Cytosolic non-specific dipeptidase | Glutamate carboxypeptidase | Q96KP4 | 55748 | CNDP2 | 1.163815 | 0.12112 |
| SL004588 | Interleukin-1 Receptor accessory protein | IL-1 R AcP | Q9NPH3 | 3556 | IL1RAP | 0.983732 | 0.121601 |
| SL004305 | DNA topoisomerase 1 | Topoisomerase I | P11387 | 7150 | TOP1 | -0.661586 | 0.122916 |
| SL007356 | Neurogenic locus notch homolog protein 2 | NOTC2 | Q04721 | 4853 | NOTCH2 | -1.217154 | 0.122996 |
| SL000247 | 6-phosphogluconate dehydrogenase, decarboxylating | 6-Phosphogluconate dehydrogenase | P52209 | 5226 | PGD | -0.455444 | 0.124066 |
| SL007804 | Biglycan | BGN | P21810 | 633 | BGN | -0.396477 | 0.125719 |
| SL000433 | Glucagon | Glucagon | P01275 | 2641 | GCG | 0.865491 | 0.1262 |
| SL005177 | Interleukin-36 alpha | IL-1F6 | Q9UHA7 | 27179 | IL36A | -1.08203 | 0.126902 |
| SL003726 | Serine/threonine-protein kinase Chk2 | Chk2 | O96017 | 11200 | CHEK2 | -1.163539 | 0.135622 |
| SL003522 | Endoplasmic reticulum resident protein 29 | ERP29 | P30040 | 10961 | ERP29 | 0.633562 | 0.137518 |
| SL016549 | AMP Kinase (alpha2beta2gamma1) | AMPK a2b2g1 | P54646 O43741 P54619 | 5563 5565 5571 | PRKAA2 PRKAB2 PRKAG1 | -0.767052 | 0.138024 |
| SL000445 | Protein Rev_HV2BE | HIV-2 Rev | P18093 | 1724716 | Human-virus | 1.165142 | 0.138538 |
| SL017612 | Heat shock protein HSP 90-alpha/beta | HSP 90a/b | P07900 P08238 | NaN | HSP90AA1 HSP90AB1 | 0.506001 | 0.139403 |
| SL014488 | Proto-oncogene vav | VAV | P15498 | 7409 | VAV1 | -0.589307 | 0.141715 |
| SL004128 | Activin receptor type-1B | Activin RIB | P36896 | 91 | ACVR1B | -0.626011 | 0.143003 |
| SL004080 | Bone morphogenetic protein receptor type-1A | BMPR1A | P36894 | 657 | BMPR1A | -1.05484 | 0.143162 |
| SL000455 | Transcription factor AP-1 | c-Jun | P05412 | 3725 | JUN | -0.461193 | 0.143776 |
| SL004101 | Mothers against decapentaplegic homolog 2 | SMAD2 | Q15796 | 4087 | SMAD2 | -0.884197 | 0.143836 |
| SL003331 | Matrix metalloproteinase-16 | MMP-16 | P51512 | 4325 | MMP16 | -0.553142 | 0.144723 |
| SL008631 | Desmocollin-2 | DSC2 | Q02487 | 1824 | DSC2 | -1.301237 | 0.145437 |
| SL005179 | Interleukin-36 beta | IL-1F8 | Q9NZH7 | 27177 | IL36B | 1.164753 | 0.145437 |
| SL007752 | Death-associated protein kinase 2 | DAPK2 | Q9UIK4 | 23604 | DAPK2 | 0.12512 | 0.146509 |
| SL002706 | Low affinity immunoglobulin epsilon Fc receptor | CD23 | P06734 | 2208 | FCER2 | -1.287334 | 0.147052 |
| SL010616 | Tyrosine-protein phosphatase non-receptor type 11 | SHP-2 | Q06124 | 5781 | PTPN11 | -1.385919 | 0.149913 |
| SL010516 | Proto-oncogene tyrosine-protein kinase Src | SRCN1 | P12931 | 6714 | SRC | -1.012944 | 0.150872 |
| SL003785 | Glyceraldehyde-3-phosphate dehydrogenase | GAPDH, liver | P04406 | 2597 | GAPDH | -0.575157 | 0.152021 |
| SL012517 | R-spondin-4 | RSPO4 | Q2I0M5 | 343637 | RSPO4 | -1.228922 | 0.152159 |
| SL016566 | Serine/threonine-protein kinase 17B | DRAK2 | O94768 | 9262 | STK17B | -0.421081 | 0.154548 |
| SL007228 | Protein kinase C iota type | KPCI | P41743 | 5584 | PRKCI | -0.923858 | 0.154548 |
| SL003753 | Dynein light chain 1, cytoplasmic | DLC8 | P63167 | 8655 | DYNLL1 | -1.145803 | 0.155142 |
| SL003190 | C-C motif chemokine 15 | MIP-5 | Q16663 | 6359 | CCL15 | 0.49285 | 0.156673 |
| SL000104 | Apoptosis regulator Bcl-2 | Bcl-2 | P10415 | 596 | BCL2 | 0.726173 | 0.157167 |
| SL013490 | Brother of CDO | BOC | Q9BWV1 | 91653 | BOC | 1.213759 | 0.157701 |
| SL007022 | Heterogeneous nuclear ribonucleoprotein Q | HNRPQ | O60506 | 10492 | SYNCRIP | -1.24595 | 0.158387 |
| SL002655 | Connective tissue growth factor | CTGF | P29279 | 1490 | CTGF | 1.094737 | 0.158404 |
| SL004654 | Granzyme H | Granzyme H | P20718 | 2999 | GZMH | 1.083366 | 0.159132 |
| SL004921 | Nucleoside diphosphate kinase B | NDP kinase B | P22392 | 4831 | NME2 | -0.784403 | 0.162079 |
| SL014269 | Killer cell immunoglobulin-like receptor 3DL2 | KI3L2 | P43630 | 3812 | KIR3DL2 | 0.792109 | 0.162752 |
| SL000337 | Calpain I | Calpain I | P07384 P04632 | 823 826 | CAPN1 CAPNS1 | -0.452923 | 0.167197 |
| SL004781 | Tyrosine-protein kinase CSK | CSK | P41240 | 1445 | CSK | -0.947806 | 0.16735 |
| SL005231 | Netrin receptor UNC5D | UNC5H4 | Q6UXZ4 | 137970 | UNC5D | 1.234243 | 0.168326 |
| SL007250 | Phosphatidylinositol 4,5-bisphosphate 3-kinase catalytic subunit gamma isoform | PK3CG | P48736 | 5294 | PIK3CG | 1.144099 | 0.169155 |
| SL013489 | Protein amnionless | AMNLS | Q9BXJ7 | 81693 | AMN | -1.14158 | 0.169339 |
| SL011232 | Hsp90 co-chaperone Cdc37 | CDC37 | Q16543 | 11140 | CDC37 | -0.78906 | 0.170558 |
| SL010513 | cAMP-dependent protein kinase catalytic subunit alpha | PRKACA | P17612 | 5566 | PRKACA | 0.654569 | 0.172137 |
| SL012457 | Neurexophilin-1 | NXPH1 | P58417 | 30010 | NXPH1 | 0.399459 | 0.172746 |
| SL005679 | Translationally-controlled tumor protein | TCTP | P13693 | 7178 | TPT1 | -1.056152 | 0.172746 |
| SL000130 | G2/mitotic-specific cyclin-B1 | Cyclin B1 | P14635 | 891 | CCNB1 | 1.188355 | 0.174412 |
| SL011631 | Nascent polypeptide-associated complex subunit alpha | NACA | Q13765 | 4666 | NACA | -0.869001 | 0.175557 |
| SL006912 | Tyrosine-protein kinase Fgr | FGR | P09769 | 2268 | FGR | -0.194506 | 0.177873 |
| SL011616 | Eukaryotic initiation factor 4A-III | IF4A3 | P38919 | 9775 | EIF4A3 | -0.917111 | 0.179145 |
| SL000342 | Catalase | Catalase | P04040 | 847 | CAT | -1.002038 | 0.180716 |
| SL004151 | Interleukin-15 receptor subunit alpha | IL-15 Ra | Q13261 | 3601 | IL15RA | 1.186654 | 0.180767 |
| SL010467 | Repulsive guidance molecule A | RGMA | Q96B86 | 56963 | RGMA | -1.20417 | 0.182181 |
| SL004938 | Calcium/calmodulin-dependent protein kinase kinase 1 | CaMKK alpha | Q8N5S9 | 84254 | CAMKK1 | -0.514112 | 0.183251 |
| SL004484 | Pulmonary surfactant-associated protein D | SP-D | P35247 | 6441 | SFTPD | -0.550347 | 0.183979 |
| SL006197 | Mitochondrial import inner membrane translocase subunit TIM14 | DnaJ homolog | Q96DA6 | 131118 | DNAJC19 | 1.202821 | 0.184888 |
| SL004457 | Glia-derived nexin | Protease nexin I | P07093 | 5270 | SERPINE2 | 0.363167 | 0.184888 |
| SL004269 | Appetite-regulating hormone | ghrelin | Q9UBU3 | 51738 | GHRL | -1.143007 | 0.184888 |
| SL006675 | Cytoskeleton-associated protein 2 | CKAP2 | Q8WWK9 | 26586 | CKAP2 | 1.147412 | 0.185376 |
| SL008059 | 40S ribosomal protein S3 | RS3 | P23396 | 6188 | RPS3 | -0.644671 | 0.186093 |
| SL008102 | Malate dehydrogenase, cytoplasmic | MDHC | P40925 | 4190 | MDH1 | -0.89624 | 0.186501 |
| SL003280 | High mobility group protein B1 | HMG-1 | P09429 | 3146 | HMGB1 | 0.274658 | 0.187705 |
| SL007284 | Cysteine-rich secretory protein 3 | CRIS3 | P54108 | 10321 | CRISP3 | -0.419162 | 0.188301 |
| SL007108 | Interferon regulatory factor 1 | IRF1 | P10914 | 3659 | IRF1 | -1.046217 | 0.188943 |
| SL000588 | Thyroid peroxidase | TMA | P07202 | 7173 | TPO | 1.147565 | 0.188943 |
| SL000617 | Alanine aminotransferase 1 | ALT | P24298 | 2875 | GPT | 1.179793 | 0.189645 |
| SL007747 | Serine/threonine-protein kinase TBK1 | TBK1 | Q9UHD2 | 29110 | TBK1 | 1.152349 | 0.190171 |
| SL003341 | Fibrinogen gamma chain | Fibrinogen g-chain dimer | P02679 | 2266 | FGG | 0.375637 | 0.190171 |
| SL003650 | Alpha-enolase | Alpha enolase | P06733 | 2023 | ENO1 | -0.633303 | 0.190176 |
| SL005793 | Peptidyl-prolyl cis-trans isomerase F, mitochondrial | Cyclophilin F | P30405 | 10105 | PPIF | -0.550324 | 0.190808 |
| SL008904 | Lymphatic vessel endothelial hyaluronic acid receptor 1 | LYVE1 | Q9Y5Y7 | 10894 | LYVE1 | 1.219359 | 0.191797 |
| SL010456 | Cystatin-SN | CYTN | P01037 | 1469 | CST1 | -1.055178 | 0.192822 |
| SL005193 | Junctional adhesion molecule B | JAM-B | P57087 | 58494 | JAM2 | -1.151159 | 0.192822 |
| SL005308 | Proteasome activator complex subunit 3 | PSME3 | P61289 | 10197 | PSME3 | -0.603609 | 0.194858 |
| SL003916 | Kallikrein-12 | kallikrein 12 | Q9UKR0 | 43849 | KLK12 | 1.221063 | 0.195874 |
| SL003919 | Kallikrein-14 | kallikrein 14 | Q9P0G3 | 43847 | KLK14 | -1.147874 | 0.195975 |
| SL001973 | Mammaglobin-B | Mammaglobin 2 | O75556 | 4246 | SCGB2A1 | -0.393036 | 0.196155 |
| SL000479 | Interleukin-3 | IL-3 | P08700 | 3562 | IL3 | 1.156112 | 0.198364 |
| SL004340 | Fibroblast growth factor 6 | FGF-6 | P10767 | 2251 | FGF6 | -0.515086 | 0.205522 |
| SL011404 | cAMP-specific 3',5'-cyclic phosphodiesterase 4D | PDE4D | Q08499 | 5144 | PDE4D | -0.511307 | 0.205544 |
| SL009948 | Plexin-B2 | PLXB2 | O15031 | 23654 | PLXNB2 | 0.202886 | 0.205812 |
| SL000074 | Cellular tumor antigen p53 | p53 | P04637 | 7157 | TP53 | 1.151425 | 0.209008 |
| SL004591 | Granulocyte colony-stimulating factor receptor | G-CSF-R | Q99062 | 1441 | CSF3R | 1.231917 | 0.209174 |
| SL010378 | Proto-oncogene tyrosine-protein kinase receptor Ret | RET | P07949 | 5979 | RET | 1.297162 | 0.209199 |
| SL010504 | MAP kinase-activated protein kinase 5 | MAPK5 | Q8IW41 | 8550 | MAPKAPK5 | -1.125433 | 0.210679 |
| SL008504 | N-acetylglucosamine-6-sulfatase | GNS | P15586 | 2799 | GNS | 1.260031 | 0.213386 |
| SL004739 | Inter-alpha-trypsin inhibitor heavy chain H4 | ITI heavy chain H4 | Q14624 | 3700 | ITIH4 | 0.686181 | 0.215971 |
| SL009431 | Histidine triad nucleotide-binding protein 1 | HINT1 | P49773 | 3094 | HINT1 | -0.41598 | 0.223533 |
| SL011100 | CD226 antigen | CD226 | Q15762 | 10666 | CD226 | 1.169223 | 0.226619 |
| SL004704 | COMM domain-containing protein 7 | COMMD7 | Q86VX2 | 149951 | COMMD7 | 1.312726 | 0.226619 |
| SL016129 | Protein FAM107B | FAM107B | Q9H098 | 83641 | FAM107B | -1.023947 | 0.22663 |
| SL004805 | Cell adhesion molecule 1 | Nectin-like protein 2 | Q9BY67 | 23705 | CADM1 | -1.1362 | 0.227704 |
| SL004671 | Tumor necrosis factor receptor superfamily member 13C | BAFF Receptor | Q96RJ3 | 115650 | TNFRSF13C | -0.666632 | 0.231857 |
| SL004864 | Cadherin-12 | Cadherin-12 | P55289 | 1010 | CDH12 | 0.263812 | 0.234237 |
| SL004338 | Fibroblast growth factor 20 | FGF-20 | Q9NP95 | 26281 | FGF20 | 1.165454 | 0.234343 |
| SL006916 | Tyrosine-protein kinase Lck | LCK | P06239 | 3932 | LCK | -1.207957 | 0.240908 |
| SL011180 | Hexokinase-1 | HXK1 | P19367 | 3098 | HK1 | 1.048238 | 0.243286 |
| SL008094 | Phosphoglucomutase-1 | PGM1 | P36871 | 5236 | PGM1 | -0.923691 | 0.244473 |
| SL011529 | Ribosome maturation protein SBDS | SBDS | Q9Y3A5 | 51119 | SBDS | -0.52187 | 0.245987 |
| SL004723 | Histone deacetylase 8 | HDAC8 | Q9BY41 | 55869 | HDAC8 | -1.162098 | 0.246508 |
| SL010468 | RGM domain family member B | RGMB | Q6NW40 | 285704 | RGMB | -1.221156 | 0.247113 |
| SL010529 | Ubiquitin-fold modifier-conjugating enzyme 1 | UFC1 | Q9Y3C8 | 51506 | UFC1 | -0.352411 | 0.250225 |
| SL010508 | Serine/threonine-protein kinase PAK 3 | PAK3 | O75914 | 5063 | PAK3 | -0.640189 | 0.25184 |
| SL001795 | Interleukin-1 beta | IL-1b | P01584 | 3553 | IL1B | 1.313644 | 0.25306 |
| SL007953 | Pyridoxal kinase | PDXK | O00764 | 8566 | PDXK | -0.576609 | 0.253979 |
| SL010488 | Abelson tyrosine-protein kinase 2 | ABL2 | P42684 | 27 | ABL2 | 1.20296 | 0.255234 |
| SL004844 | Ephrin type-A receptor 5 | EphA5 | P54756 | 2044 | EPHA5 | -0.656322 | 0.257329 |
| SL000398 | Cytochrome P450 3A4 | Cytochrome P450 3A4 | P08684 | 1576 | CYP3A4 | -1.288698 | 0.257789 |
| SL003327 | Complement factor D | Factor D | P00746 | 1675 | CFD | -1.143786 | 0.257885 |
| SL000498 | Leptin | Leptin | P41159 | 3952 | LEP | -0.575255 | 0.258751 |
| SL008916 | Dipeptidyl peptidase 2 | DPP2 | Q9UHL4 | 29952 | DPP7 | 1.076703 | 0.259891 |
| SL005173 | Interleukin-10 receptor subunit alpha | IL-10 Ra | Q13651 | 3587 | IL10RA | 1.22808 | 0.262338 |
| SL004661 | Aggrecan core protein | Aggrecan | P16112 | 176 | ACAN | 1.107763 | 0.262338 |
| SL004863 | Tumor necrosis factor receptor superfamily member 19 | TAJ | Q9NS68 | 55504 | TNFRSF19 | -1.220466 | 0.263971 |
| SL019100 | E3 ubiquitin-protein ligase CHIP | CHIP | Q9UNE7 | 10273 | STUB1 | -0.307194 | 0.265155 |
| SL000319 | Complement C5 | C5 | P01031 | 727 | C5 | -1.249478 | 0.266687 |
| SL004330 | B-cell receptor CD22 | CD22 | P20273 | 933 | CD22 | 1.14829 | 0.267543 |
| SL005222 | Scavenger receptor class F member 2 | SREC-II | Q96GP6 | 91179 | SCARF2 | -1.166627 | 0.26798 |
| SL014070 | SLIT and NTRK-like protein 5 | SLIK5 | O94991 | 26050 | SLITRK5 | -0.681328 | 0.268238 |
| SL003305 | Interleukin-2 receptor subunit alpha | IL-2 sRa | P01589 | 3559 | IL2RA | 1.137757 | 0.269239 |
| SL000553 | Protein kinase C beta type (splice variant beta-II) | PKC-B-II | P05771 | 5579 | PRKCB | -0.900842 | 0.269681 |
| SL010521 | Tyrosine-protein kinase BTK | BTK | Q06187 | 695 | BTK | -0.675704 | 0.27426 |
| SL010498 | Ephrin type-A receptor 3 | EPHA3 | P29320 | 2042 | EPHA3 | 1.102993 | 0.277999 |
| SL000658 | Growth arrest-specific protein 1 | GAS1 | P54826 | 2619 | GAS1 | 0.57498 | 0.283149 |
| SL005588 | Tyrosine-protein kinase Fer | FER | P16591 | 2241 | FER | -0.683614 | 0.285623 |
| SL005202 | Hepatocyte growth factor-like protein | MSP | P26927 | 4485 | MST1 | -1.275685 | 0.289446 |
| SL000674 | Follistatin | FST | P19883 | 10468 | FST | -1.169998 | 0.294817 |
| SL002650 | Pyruvate kinase PKM | M2-PK | P14618 | 5315 | PKM2 | -0.429632 | 0.300166 |
| SL007136 | CCAAT/enhancer-binding protein beta | CEBPB | P17676 | 1051 | CEBPB | 0.232661 | 0.301266 |
| SL006629 | NAD-dependent protein deacetylase sirtuin-2 | SIRT2 | Q8IXJ6 | 22933 | SIRT2 | -0.29234 | 0.303742 |
| SL010973 | Proteasome subunit alpha type-1 | PSA1 | P25786 | 5682 | PSMA1 | -0.269935 | 0.307397 |
| SL006915 | Interleukin-27 | IL-27 | Q8NEV9 Q14213 | 246778 10148 | IL27 EBI3 | 1.094655 | 0.308614 |
| SL004687 | Thymic stromal lymphopoietin | TSLP | Q969D9 | 85480 | TSLP | -0.318522 | 0.308861 |
| SL004364 | Tumor necrosis factor receptor superfamily member 13B | TACI | O14836 | 23495 | TNFRSF13B | 1.218852 | 0.308861 |
| SL005574 | Aminoacylase-1 | Aminoacylase-1 | Q03154 | 95 | ACY1 | 0.275418 | 0.314681 |
| SL000448 | Heat shock protein beta-1 | HSP 27 | P04792 | 3315 | HSPB1 | -0.444917 | 0.316839 |
| SL004332 | Prokineticin-1 | EG-VEGF | P58294 | 84432 | PROK1 | 0.413059 | 0.317095 |
| SL003792 | Growth factor receptor-bound protein 2 | GRB2 adapter protein | P62993 | 2885 | GRB2 | -0.601334 | 0.322422 |
| SL006131 | Fatty acid-binding protein, liver | FABPL | P07148 | 2168 | FABP1 | -0.660073 | 0.325907 |
| SL003915 | Kallikrein-8 | kallikrein 8 | O60259 | 11202 | KLK8 | 0.831626 | 0.326656 |
| SL006378 | S-formylglutathione hydrolase | Esterase D | P10768 | 2098 | ESD | -1.087166 | 0.328973 |
| SL014470 | Breast cancer anti-estrogen resistance protein 3 | BCAR3 | O75815 | 8412 | BCAR3 | 1.220072 | 0.330003 |
| SL004815 | 14-3-3 protein beta/alpha | 14-3-3 protein beta/alpha | P31946 | 7529 | YWHAB | -0.356512 | 0.330003 |
| SL017611 | 14-3-3 protein family | 14-3-3 | P31946, P62258, P61981, Q04917, P27348, P63104, P31947 | 7529, 7531, 7532, 7533, 10971, 7534, 2810 | YWHAB,YWHAE,YWHAG,YWHAH,YWHAQ,YWHAZ,SFN | 0.51863 | 0.331505 |
| SL010927 | Importin subunit beta-1 | IMB1 | Q14974 | 3837 | KPNB1 | -0.363753 | 0.331505 |
| SL014684 | 2'-5'-oligoadenylate synthase 1 | OAS1 | P00973 | 4938 | OAS1 | -0.441831 | 0.331525 |
| SL003653 | Phosphoglycerate kinase 1 | phosphoglycerate kinase 1 | P00558 | 5230 | PGK1 | 0.412909 | 0.331525 |
| SL003647 | Annexin A6 | annexin VI | P08133 | 309 | ANXA6 | -1.190242 | 0.331525 |
| SL009791 | Heterogeneous nuclear ribonucleoprotein A/B | hnRNP A/B | Q99729 | 3182 | HNRNPAB | -0.577472 | 0.33289 |
| SL002640 | Placenta growth factor | PlGF | P49763 | 5228 | PGF | -1.123723 | 0.33756 |
| SL005170 | Inducible T-cell costimulator | ICOS | Q9Y6W8 | 29851 | ICOS | 0.689175 | 0.338613 |
| SL004182 | Calcium-dependent phospholipase A2 | GV | P39877 | 5322 | PLA2G5 | 1.159506 | 0.339724 |
| SL001880 | Interferon beta | IFN-b | P01574 | 3456 | IFNB1 | -1.159236 | 0.344942 |
| SL014228 | SLAM family member 6 | SLAF6 | Q96DU3 | 114836 | SLAMF6 | 1.189108 | 0.345616 |
| SL004795 | 3-hydroxyacyl-CoA dehydrogenase type-2 | ERAB | Q99714 | 3028 | HSD17B10 | -0.401027 | 0.349964 |
| SL011405 | cGMP-specific 3',5'-cyclic phosphodiesterase | PDE5A | O76074 | 8654 | PDE5A | 0.660317 | 0.352128 |
| SL008360 | Leucine-rich repeat transmembrane protein FLRT2 | FLRT2 | O43155 | 23768 | FLRT2 | -0.845544 | 0.355088 |
| SL004345 | Growth/differentiation factor 11 | GDF-11 | O95390 | 10220 | GDF11 | 1.197265 | 0.356813 |
| SL004557 | CD59 glycoprotein | CD59 | P13987 | 966 | CD59 | 0.759198 | 0.357707 |
| SL004843 | Ephrin-A2 | Ephrin-A2 | O43921 | 1943 | EFNA2 | 1.065283 | 0.358461 |
| SL005204 | NKG2-D type II integral membrane protein | NKG2D | P26718 | 22914 | KLRK1 | 1.153076 | 0.359296 |
| SL010518 | Tyrosine-protein kinase Tec | TEC | P42680 | 7006 | TEC | -0.431241 | 0.361158 |
| SL013548 | Interleukin-34 | IL-34 | Q6ZMJ4 | 146433 | IL34 | 1.115634 | 0.36636 |
| SL008614 | Ephrin-B1 | EFNB1 | P98172 | 1947 | EFNB1 | -1.077716 | 0.37748 |
| SL003646 | Tropomyosin alpha-4 chain | Tropomyosin 4 | P67936 | 7171 | TPM4 | -0.561842 | 0.379596 |
| SL008574 | Osteomodulin | OMD | Q99983 | 4958 | OMD | 0.927587 | 0.38218 |
| SL000459 | Immunoglobulin A | IgA | P01876 P01877 | 3493 3494 | IGHA1 IGHA2 | -1.257448 | 0.386088 |
| SL005361 | Apolipoprotein D | Apo D | P05090 | 347 | APOD | 1.200635 | 0.387266 |
| SL004733 | Low-density lipoprotein receptor-related protein 1, soluble | sLRP1 | Q07954 | 4035 | LRP1 | -1.200965 | 0.387289 |
| SL004362 | Stem Cell Growth Factor-beta | SCGF-beta | Q9Y240 | 6320 | CLEC11A | -0.802414 | 0.387391 |
| SL000006 | Plasminogen activator inhibitor 1 | PAI-1 | P05121 | 5054 | SERPINE1 | -1.175063 | 0.387968 |
| SL001737 | 14-3-3 protein sigma | STRATIFIN | P31947 | 2810 | SFN | 0.790864 | 0.388461 |
| SL005688 | 14-3-3 protein zeta/delta | 14-3-3 protein zeta/delta | P63104 | 7534 | YWHAZ | -0.415625 | 0.388828 |
| SL001902 | Basal Cell Adhesion Molecule | BCAM | P50895 | 4059 | BCAM | -1.21268 | 0.398744 |
| SL010503 | MAP kinase-activated protein kinase 2 | MAPK2 | P49137 | 9261 | MAPKAPK2 | -0.420651 | 0.402403 |
| SL013240 | Adapter molecule crk | CRK | P46108 | 1398 | CRK | 0.395027 | 0.405431 |
| SL003066 | Pigment epithelium-derived factor | PEDF | P36955 | 5176 | SERPINF1 | 0.501283 | 0.405431 |
| SL017290 | PolyUbiquitin K63-linked | PolyUbiquitin K63 | P0CG48 | 7316 | UBC | -0.974276 | 0.40665 |
| SL000474 | Interleukin-16 | IL-16 | Q14005 | 3603 | IL16 | 0.093308 | 0.407038 |
| SL017188 | Glycogen synthase kinase-3 alpha/beta | GSK-3 alpha/beta | P49840 P49841 | 2931 2932 | GSK3A GSK3B | -0.360122 | 0.409037 |
| SL005630 | Proteasome subunit alpha type-6 | PSA6 | P60900 | 5687 | PSMA6 | -0.217884 | 0.409607 |
| SL002036 | Fibroblast growth factor receptor 4 | FGFR4 | P22455 | 2264 | FGFR4 | 1.176127 | 0.410956 |
| SL004685 | Persephin | Persephin | O60542 | 5623 | PSPN | -1.116581 | 0.411661 |
| SL004301 | X-ray repair cross-complementing protein 6 | Ku70 | P12956 | 2547 | XRCC6 | -0.444091 | 0.412197 |
| SL010459 | Low affinity immunoglobulin gamma Fc region receptor II-a | FCG2A | P12318 | 2212 | FCGR2A | -0.899011 | 0.418069 |
| SL000254 | Serum albumin | Albumin | P02768 | 213 | ALB | -1.085475 | 0.419734 |
| SL007696 | Complement component C1q receptor | C1QR1 | Q9NPY3 | 22918 | CD93 | 0.195957 | 0.420098 |
| SL003738 | T-lymphocyte activation antigen CD80 | B7 | P33681 | 941 | CD80 | 1.09203 | 0.420098 |
| SL008591 | Trefoil factor 1 | TFF1 | P04155 | 7031 | TFF1 | 0.511603 | 0.422918 |
| SL005178 | Interleukin-37 | IL-1F7 | Q9NZH6 | 27178 | IL37 | 1.151526 | 0.422963 |
| SL004363 | Stem cell growth factor-alpha | SCGF-alpha | Q9Y240 | 6320 | CLEC11A | -0.629031 | 0.422963 |
| SL002792 | beta-adrenergic receptor kinase 1 | BARK1 | P25098 | 156 | ADRBK1 | -0.302955 | 0.422963 |
| SL004208 | Annexin A1 | annexin I | P04083 | 301 | ANXA1 | 0.377085 | 0.436017 |
| SL000308 | Plasma protease C1 inhibitor | C1-Esterase Inhibitor | P05155 | 710 | SERPING1 | -1.291085 | 0.437304 |
| SL017424 | Platelet receptor Gi24 | GI24 | Q9H7M9 | NaN | C10orf54 | -1.001978 | 0.438104 |
| SL000616 | Vitronectin | Vitronectin | P04004 | 7448 | VTN | -1.098548 | 0.439598 |
| SL011211 | Eukaryotic translation initiation factor 4 gamma 2 | IF4G2 | P78344 | 1982 | EIF4G2 | 0.639493 | 0.440543 |
| SL006088 | Sphingosine kinase 1 | Sphingosine kinase 1 | Q9NYA1 | 8877 | SPHK1 | -0.552957 | 0.440543 |
| SL003199 | Tyrosine-protein kinase receptor Tie-1, soluble | sTie-1 | P35590 | 7075 | TIE1 | -0.48887 | 0.445668 |
| SL000305 | beta-nerve growth factor | b-NGF | P01138 | 4803 | NGF | -1.165724 | 0.44922 |
| SL005250 | Chymase | Chymase | P23946 | 1215 | CMA1 | 0.168615 | 0.452243 |
| SL005218 | Sialic acid-binding Ig-like lectin 7 | Siglec-7 | Q9Y286 | 27036 | SIGLEC7 | -1.183429 | 0.45612 |
| SL005087 | Insulin-like growth factor-binding protein 7 | IGFBP-7 | Q16270 | 3490 | IGFBP7 | -0.517909 | 0.460403 |
| SL004356 | C-C motif chemokine 4-like | LAG-1 | Q8NHW4 | 388372 | CCL4L1 | -1.209485 | 0.460873 |
| SL018946 | Ubiquitin-conjugating enzyme E2 G2 | UB2G2 | P60604 | 7327 | UBE2G2 | 0.830627 | 0.461294 |
| SL007336 | Protein SET | SET | Q01105 | 6418 | SET | -1.193453 | 0.461294 |
| SL005191 | Leukocyte immunoglobulin-like receptor subfamily B member 2 | ILT-4 | Q8N423 | 10288 | LILRB2 | 0.194259 | 0.461294 |
| SL004821 | Protein S100-A4 | S100A4 | P26447 | 6275 | S100A4 | -0.318848 | 0.461294 |
| SL005403 | Vesicular integral-membrane protein VIP36 | Lectin, mannose-binding 2 | Q12907 | 10960 | LMAN2 | -0.655956 | 0.469549 |
| SL004859 | Tumor necrosis factor receptor superfamily member 18 | GITR | Q9Y5U5 | 8784 | TNFRSF18 | 1.041902 | 0.475549 |
| SL001938 | Inhibin beta A chain | Activin A | P08476 | 3624 | INHBA | -1.18649 | 0.479785 |
| SL000343 | Cathepsin B | Cathepsin B | P07858 | 1508 | CTSB | -1.191851 | 0.479785 |
| SL005227 | NKG2D ligand 1 | ULBP-1 | Q9BZM6 | 80329 | ULBP1 | 1.024249 | 0.480549 |
| SL000485 | Interleukin-9 | IL-9 | P15248 | 3578 | IL9 | 1.14022 | 0.480549 |
| SL005230 | Netrin receptor UNC5C | UNC5H3 | O95185 | 8633 | UNC5C | -1.219346 | 0.483612 |
| SL002783 | Cardiotrophin-1 | Cardiotrophin-1 | Q16619 | 1489 | CTF1 | 0.806511 | 0.484965 |
| SL004511 | Bactericidal permeability-increasing protein | BPI | P17213 | 671 | BPI | 0.677954 | 0.503543 |
| SL000497 | Laminin | Laminin | P25391, P07942, P11047 | 284217 3912 3915 | LAMA1 LAMB1 LAMC1 | -0.86204 | 0.503601 |
| SL005201 | Muellerian-inhibiting factor | MIS | P03971 | 268 | AMH | -0.803077 | 0.503921 |
| SL004009 | Ras-related C3 botulinum toxin substrate 1 | RAC1 | P63000 | 5879 | RAC1 | -0.47397 | 0.50748 |
| SL004838 | Leukocyte surface antigen CD47 | CD47 | Q08722 | 961 | CD47 | -0.931551 | 0.507617 |
| SL016548 | AMP Kinase (alpha1beta1gamma1) | AMPK a1b1g1 | Q13131 Q9Y478 P54619 | 5562 5564 5571 | PRKAA1 PRKAB1 PRKAG1 | -1.097131 | 0.508895 |
| SL008933 | Protein deglycase DJ-1 | PARK7 | Q99497 | 11315 | PARK7 | 1.144954 | 0.511444 |
| SL004725 | Histone acetyltransferase type B catalytic subunit | Hat1 | O14929 | 8520 | HAT1 | 0.325119 | 0.516162 |
| SL007453 | Mitogen-activated protein kinase 11 | MK11 | Q15759 | 5600 | MAPK11 | -0.732743 | 0.518105 |
| SL013570 | Dual specificity tyrosine-phosphorylation-regulated kinase 3 | DYRK3 | O43781 | 8444 | DYRK3 | 1.118257 | 0.520704 |
| SL009328 | Chorionic somatomammotropin hormone | CSH | P0DML2 P0DML3 | 1442 1443 | CSH1 CSH2 | 0.280232 | 0.524193 |
| SL004209 | Annexin A2 | annexin II | P07355 | 302 | ANXA2 | -0.588988 | 0.525866 |
| SL004751 | Heme oxygenase 2 | HO-2 | P30519 | 3163 | HMOX2 | 0.595206 | 0.526079 |
| SL016128 | Thioredoxin domain-containing protein 12 | TXD12 | O95881 | 51060 | TXNDC12 | -1.199934 | 0.529414 |
| SL000007 | Estrogen receptor | ER | P03372 | 2099 | ESR1 | -1.158348 | 0.529414 |
| SL007888 | Cystatin-S | Cystatin-S | P01036 | 1472 | CST4 | -1.150599 | 0.534457 |
| SL000450 | 60 kDa heat shock protein, mitochondrial | HSP 60 | P10809 | 3329 | HSPD1 | 0.289691 | 0.535004 |
| SL005197 | Lysosome membrane protein 2 | LIMP II | Q14108 | 950 | SCARB2 | 1.141747 | 0.535639 |
| SL008522 | Semaphorin-5A | SEM5A | Q13591 | 9037 | SEMA5A | -1.257624 | 0.538195 |
| SL005226 | Cytokine receptor-like factor 2 | TSLP R | Q9HC73 | 64109 | CRLF2 | -0.209706 | 0.546147 |
| SL004851 | Serine/threonine-protein kinase receptor R3 | ALK-1 | P37023 | 94 | ACVRL1 | 0.519239 | 0.548698 |
| SL010523 | Homeodomain-interacting protein kinase 3 | HIPK3 | Q9H422 | 10114 | HIPK3 | -1.115072 | 0.549223 |
| SL003362 | Complement C3d fragment | C3d | P01024 | 718 | C3 | 0.14262 | 0.557875 |
| SL004932 | Peroxiredoxin-5, mitochondrial | Peroxiredoxin-5 | P30044 | 25824 | PRDX5 | -1.086451 | 0.558715 |
| SL000027 | Prostaglandin G/H synthase 2 | COX-2 | P35354 | 5743 | PTGS2 | -0.33301 | 0.559418 |
| SL004610 | Low-density lipoprotein receptor-related protein 8 | LRP8 | Q14114 | 7804 | LRP8 | -0.936837 | 0.560354 |
| SL000440 | Hemopexin | Hemopexin | P02790 | 3263 | HPX | 0.441017 | 0.560354 |
| SL007674 | T-lymphocyte surface antigen Ly-9 | LY9 | Q9HBG7 | 4063 | LY9 | -1.248997 | 0.56049 |
| SL000554 | Protein kinase C delta type | PKC-D | Q05655 | 5580 | PRKCD | 0.655383 | 0.56049 |
| SL014096 | Leucine-rich repeat transmembrane neuronal protein 1 | LRRT1 | Q86UE6 | 347730 | LRRTM1 | -1.15305 | 0.560808 |
| SL004924 | Eukaryotic translation initiation factor 4H | eIF-4H | Q15056 | 7458 | EIF4H | 0.622753 | 0.561876 |
| SL004486 | Vascular endothelial growth factor C | VEGF-C | P49767 | 7424 | VEGFC | 0.520008 | 0.566308 |
| SL002763 | Kallikrein-11 | Kallikrein 11 | Q9UBX7 | 11012 | KLK11 | -1.198334 | 0.566308 |
| SL003761 | Phosphatidylinositol 3,4,5-trisphosphate 3-phosphatase and dual-specificity protein phosphatase PTEN | pTEN | P60484 | 5728 | PTEN | -1.06272 | 0.569112 |
| SL014735 | Inosine-5'-monophosphate dehydrogenase 1 | IMDH1 | P20839 | 3614 | IMPDH1 | 0.487413 | 0.571839 |
| SL010469 | Hemojuvelin | RGM-C | Q6ZVN8 | 148738 | HFE2 | 0.28105 | 0.574087 |
| SL000142 | Thymidylate synthase | TS | P04818 | 7298 | TYMS | -1.113533 | 0.583841 |
| SL004556 | Complement decay-accelerating factor | DAF | P08174 | 1604 | CD55 | -0.689756 | 0.584944 |
| SL003774 | Bcl-2-like protein 2 | Apoptosis regulator Bcl-W | Q92843 | 599 | BCL2L2 | 1.130414 | 0.589094 |
| SL012168 | Protein lin-7 homolog B | LIN7B | Q9HAP6 | 64130 | LIN7B | -0.254471 | 0.596146 |
| SL000480 | Interleukin-4 | IL-4 | P05112 | 3565 | IL4 | -1.085932 | 0.598419 |
| SL006920 | Mitogen-activated protein kinase 14 | MAPK14 | Q16539 | 1432 | MAPK14 | -0.296888 | 0.600976 |
| SL008122 | Dual specificity protein phosphatase 3 | DUS3 | P51452 | 1845 | DUSP3 | -0.312191 | 0.610066 |
| SL004829 | Protein S100-A7 | S100A7 | P31151 | 6278 | S100A7 | 1.017272 | 0.610066 |
| SL011629 | AH receptor-interacting protein | AIP | O00170 | 9049 | AIP | -0.264428 | 0.615015 |
| SL016828 | Epidermal growth factor receptor variant III | EGFRvIII | P00533 | NaN | EGFR | -0.56006 | 0.61586 |
| SL012769 | Protein FAM3D | FAM3D | Q96BQ1 | 131177 | FAM3D | 0.792319 | 0.61586 |
| SL000322 | Complement component C6 | C6 | P13671 | 729 | C6 | -1.125516 | 0.61586 |
| SL000582 | Baculoviral IAP repeat-containing protein 5 | Survivin | O15392 | 332 | BIRC5 | 1.098756 | 0.621448 |
| SL001718 | Interleukin-13 | IL-13 | P35225 | 3596 | IL13 | -0.467884 | 0.623811 |
| SL003461 | Corticotropin | ACTH | P01189 | 5443 | POMC | -1.157963 | 0.623962 |
| SL010928 | Inosine-5'-monophosphate dehydrogenase 2 | IMDH2 | P12268 | 3615 | IMPDH2 | -0.231832 | 0.624309 |
| SL000586 | Thrombin | Thrombin | P00734 | 2147 | F2 | 1.034178 | 0.624309 |
| SL012538 | Cerebral dopamine neurotrophic factor | ARMEL | Q49AH0 | 441549 | CDNF | 0.155083 | 0.633144 |
| SL003711 | Caspase-3 | Caspase-3 | P42574 | 836 | CASP3 | -0.473871 | 0.633144 |
| SL005372 | Sorting nexin-4 | Sorting nexin 4 | O95219 | 8723 | SNX4 | -0.289729 | 0.635463 |
| SL008382 | Cystatin-D | CYTD | P28325 | 1473 | CST5 | -0.342219 | 0.63743 |
| SL004153 | Macrophage colony-stimulating factor 1 receptor | M-CSF R | P07333 | 1436 | CSF1R | -1.222394 | 0.640572 |
| SL006476 | Glycylpeptide N-tetradecanoyltransferase 1 | NMT1 | P30419 | 4836 | NMT1 | -0.260563 | 0.641341 |
| SL006374 | Cytoplasmic tyrosine-protein kinase BMX | BMX | P51813 | 660 | BMX | -0.743684 | 0.641341 |
| SL005225 | High affinity nerve growth factor receptor | TrkA | P04629 | 4914 | NTRK1 | 0.750738 | 0.641341 |
| SL005575 | Galactoside 3(4)-L-fucosyltransferase | Fucosyltransferase 3 | P21217 | 2525 | FUT3 | -1.238591 | 0.642887 |
| SL010528 | Ubiquitin-fold modifier 1 | UFM1 | P61960 | 51569 | UFM1 | 0.119742 | 0.648905 |
| SL010522 | Cyclin-dependent kinase 8:Cyclin-C complex | CDK8/cyclin C | P49336 P24863 | 1024 892 | CDK8 CCNC | -1.196519 | 0.648905 |
| SL010490 | Calcium/calmodulin-dependent protein kinase type 1D | CAMK1D | Q8IU85 | 57118 | CAMK1D | 1.136166 | 0.648905 |
| SL008956 | Arylsulfatase B | ARSB | P15848 | 411 | ARSB | -1.10644 | 0.648905 |
| SL007324 | Casein kinase II subunit alpha | CSK21 | P68400 | 1457 | CSNK2A1 | 1.00951 | 0.648905 |
| SL019978 | Interferon alpha-7 | IFNA7 | P01567 | NaN | IFNA7 | -1.096202 | 0.650518 |
| SL005491 | Opioid-binding protein/cell adhesion molecule | OBCAM | Q14982 | 4978 | OPCML | -1.124254 | 0.661534 |
| SL011498 | Pituitary adenylate cyclase-activating polypeptide 38 | PACAP-38 | P18509 | 116 | ADCYAP1 | 1.098073 | 0.662571 |
| SL004335 | Fibroblast growth factor 17 | FGF-17 | O60258 | 8822 | FGF17 | 1.0833 | 0.665043 |
| SL000483 | Interleukin-7 | IL-7 | P13232 | 3574 | IL7 | -0.484408 | 0.665043 |
| SL004910 | Fructose-bisphosphate aldolase A | aldolase A | P04075 | 226 | ALDOA | 0.193285 | 0.66854 |
| SL004853 | ICOS ligand | B7-H2 | O75144 | 23308 | ICOSLG | 0.520942 | 0.670129 |
| SL003735 | Tumor necrosis factor ligand superfamily member 9 | 4-1BB ligand | P41273 | 8744 | TNFSF9 | 1.109337 | 0.670129 |
| SL002541 | Tumor necrosis factor ligand superfamily member 11 | sRANKL | O14788 | 8600 | TNFSF11 | 1.074621 | 0.675162 |
| SL018629 | E3 ubiquitin-protein ligase RNF43 | RNF43 | Q68DV7 | 54894 | RNF43 | 1.129233 | 0.676671 |
| SL004636 | Receptor-type tyrosine-protein kinase FLT3 | Flt-3 | P36888 | 2322 | FLT3 | 0.221495 | 0.6771 |
| SL009790 | Small nuclear ribonucleoprotein F | RUXF | P62306 | 6636 | SNRPF | 1.123175 | 0.684472 |
| SL014188 | Natural killer cell receptor 2B4 | CD244 | Q9BZW8 | 51744 | CD244 | 1.100509 | 0.686044 |
| SL000516 | C-C motif chemokine 7 | MCP-3 | P80098 | 6354 | CCL7 | 0.933244 | 0.692522 |
| SL004852 | Programmed cell death 1 ligand 1 | B7-H1 | Q9NZQ7 | 29126 | CD274 | 0.60193 | 0.697632 |
| SL004126 | Tumor necrosis factor receptor superfamily member 9 | 4-1BB | Q07011 | 3604 | TNFRSF9 | 0.786206 | 0.707141 |
| SL004676 | Insulin-like growth factor-binding protein 5 | IGFBP-5 | P24593 | 3488 | IGFBP5 | 0.512199 | 0.708352 |
| SL011069 | Serine protease 27 | Marapsin | Q9BQR3 | 83886 | PRSS27 | -1.179842 | 0.710509 |
| SL018921 | HemK methyltransferase family member 2 | HEMK2 | Q9Y5N5 | 29104 | N6AMT1 | 0.153317 | 0.714542 |
| SL004366 | Tumor necrosis factor receptor superfamily member 12A | TWEAKR | Q9NP84 | 51330 | TNFRSF12A | -0.989988 | 0.716361 |
| SL009768 | Chromobox protein homolog 5 | CBX5 | P45973 | 23468 | CBX5 | 1.094795 | 0.722596 |
| SL004670 | Artemin | Artemin | Q5T4W7 | 9048 | ARTN | -1.161059 | 0.722596 |
| SL008865 | Proteasome subunit alpha type-2 | PSA2 | P25787 | 5683 | PSMA2 | -0.127839 | 0.727597 |
| SL019019 | PILR alpha-associated neural protein | PIANP | Q8IYJ0 | 196500 | PIANP | -0.414616 | 0.728329 |
| SL008808 | S-phase kinase-associated protein 1 | SKP1 | P63208 | 6500 | SKP1 | -0.740064 | 0.728329 |
| SL003657 | Calcineurin | Calcineurin | Q08209 P63098 | 5530 5534 | PPP3CA PPP3R1 | 0.594166 | 0.733131 |
| SL007358 | Protein kinase C theta type | KPCT | Q04759 | 5588 | PRKCQ | 0.26922 | 0.738671 |
| SL007373 | Peptidyl-prolyl cis-trans isomerase D | PPID | Q08752 | 5481 | PPID | 0.345569 | 0.746073 |
| SL000542 | Integrin alpha-IIb: beta-3 complex | gpIIbIIIa | P08514 P05106 | 3674 3690 | ITGA2B ITGB3 | 1.158329 | 0.749953 |
| SL000346 | Cathepsin H | Cathepsin H | P09668 | 1512 | CTSH | -0.912835 | 0.75175 |
| SL010514 | Protein-tyrosine kinase 6 | PTK6 | Q13882 | 5753 | PTK6 | -1.106072 | 0.759403 |
| SL000323 | Complement component C7 | C7 | P10643 | 730 | C7 | -0.69791 | 0.759403 |
| SL000427 | Fractalkine | Fractalkine/CX3CL-1 | P78423 | 6376 | CX3CL1 | 0.876163 | 0.781183 |
| SL004849 | X-linked interleukin-1 receptor accessory protein-like 2 | IL-1 sR9 | Q9NP60 | 26280 | IL1RAPL2 | 1.096354 | 0.783757 |
| SL014468 | SH2 domain-containing protein 1A | SH21A | O60880 | 4068 | SH2D1A | -0.907415 | 0.788093 |
| SL010501 | Dual specificity mitogen-activated protein kinase kinase 2 | MP2K2 | P36507 | 5605 | MAP2K2 | -1.182472 | 0.788093 |
| SL004145 | Tumor necrosis factor receptor superfamily member 14 | HVEM | Q92956 | 8764 | TNFRSF14 | 1.096545 | 0.790247 |
| SL003930 | 15-hydroxyprostaglandin dehydrogenase [NAD(+)] | HPG- | P15428 | 3248 | HPGD | 1.158744 | 0.793126 |
| SL002525 | Complement C2 | C2 | P06681 | 717 | C2 | -0.819868 | 0.79425 |
| SL007295 | Cadherin-15 | CAD15 | P55291 | 1013 | CDH15 | 0.632692 | 0.795573 |
| SL004230 | Microtubule-associated protein tau | tau | P10636 | 4137 | MAPT | 1.152187 | 0.795891 |
| SL004771 | Aurora kinase A | Aurora kinase A | O14965 | 6790 | AURKA | -1.152567 | 0.796623 |
| SL005797 | Polymeric immunoglobulin receptor | PIGR | P01833 | 5284 | PIGR | -1.119287 | 0.796663 |
| SL004347 | Interleukin-22 | IL-22 | Q9GZX6 | 50616 | IL22 | -1.267046 | 0.797698 |
| SL004070 | Ubiquitin | Ubiquitin | P62979 | 6233 | RPS27A | -0.986505 | 0.801792 |
| SL003320 | Vascular endothelial growth factor D | VEGF-D | O43915 | 2277 | FIGF | 0.571048 | 0.808024 |
| SL007361 | Galectin-10 | LPPL | Q05315 | 1178 | CLC | 1.079489 | 0.813899 |
| SL000454 | Heat shock protein HSP 90-beta | HSP 90b | P08238 | 3326 | HSP90AB1 | 0.375957 | 0.818731 |
| SL017529 | Kelch-like ECH-associated protein 1 | KEAP1 | Q14145 | 9817 | KEAP1 | -0.616297 | 0.820447 |
| SL003704 | BH3-interacting domain death agonist | BID | P55957 | 637 | BID | -0.998645 | 0.8212 |
| SL010393 | Kremen protein 2 | KREM2 | Q8NCW0 | 79412 | KREMEN2 | -0.184229 | 0.825246 |
| SL003524 | Protein disulfide-isomerase A3 | Protein disulfide isomerase A3 | P30101 | 2923 | PDIA3 | 1.239244 | 0.826075 |
| SL011528 | 40S ribosomal protein S7 | RS7 | P62081 | 6201 | RPS7 | 0.44071 | 0.827423 |
| SL006698 | Ligand-dependent nuclear receptor corepressor-like protein | transcription factor MLR1, isoform CRA_b | Q8N3X6 | 254251 | LCORL | 1.103621 | 0.827706 |
| SL010509 | Serine/threonine-protein kinase PAK 6 | PAK6 | Q9NQU5 | 56924 | PAK6 | 0.600404 | 0.830859 |
| SL014209 | Killer cell lectin-like receptor subfamily F member 1 | KLRF1 | Q9NZS2 | 51348 | KLRF1 | -0.973982 | 0.834697 |
| SL000517 | C-C motif chemokine 13 | MCP-4 | Q99616 | 6357 | CCL13 | 0.28463 | 0.834697 |
| SL000581 | Superoxide dismutase [Cu-Zn] | SOD | P00441 | 6647 | SOD1 | -0.222707 | 0.835554 |
| SL000004 | Fibroblast growth factor 2 | bFGF | P09038 | 2247 | FGF2 | 0.5974 | 0.835554 |
| SL005164 | Galectin-2 | Galectin-2 | P05162 | 3957 | LGALS2 | 1.142461 | 0.835836 |
| SL007145 | Nuclear receptor subfamily 1 group D member 1 | NR1D1 | P20393 | 9572 | NR1D1 | 0.930783 | 0.836885 |
| SL010515 | Ribosomal protein S6 kinase alpha-3 | RPS6KA3 | P51812 | 6197 | RPS6KA3 | 0.318352 | 0.849154 |
| SL014294 | Ephrin type-A receptor 10 | EPHAA | Q5JZY3 | 284656 | EPHA10 | -1.138268 | 0.856002 |
| SL005217 | Sialic acid-binding Ig-like lectin 6 | Siglec-6 | O43699 | 946 | SIGLEC6 | 0.751029 | 0.86982 |
| SL016130 | Transcription factor IIIB 90 kDa subunit | BRF-1 | Q92994 | 2972 | BRF1 | -0.433604 | 0.872788 |
| SL008516 | Cystatin-SA | CYTT | P09228 | 1470 | CST2 | 0.270311 | 0.878276 |
| SL004686 | Tumor necrosis factor ligand superfamily member 15 | TNFSF15 | O95150 | 9966 | TNFSF15 | -0.303673 | 0.878276 |
| SL006268 | NSFL1 cofactor p47 | NSF1C | Q9UNZ2 | 55968 | NSFL1C | 0.264532 | 0.878846 |
| SL020171 | HERV-H LTR-associating protein 2 | HHLA2 | Q9UM44 | NaN | HHLA2 | -1.163813 | 0.881436 |
| SL007272 | Hexokinase-2 | HXK2 | P52789 | 3099 | HK2 | 0.846776 | 0.881436 |
| SL011073 | Xaa-Pro aminopeptidase 1 | XPNPEP1 | Q9NQW7 | 7511 | XPNPEP1 | -0.300999 | 0.885091 |
| SL000321 | Complement C5b-C6 complex | C5b, 6 Complex | P01031,P13671 | 727 729 | C5 C6 | -1.271947 | 0.88671 |
| SL006923 | TATA-box-binding protein | TBP | P20226 | 6908 | TBP | 0.256168 | 0.895094 |
| SL002756 | Heterogeneous nuclear ribonucleoprotein K | hnRNP K | P61978 | 3190 | HNRNPK | -1.185998 | 0.898697 |
| SL000047 | Insulin-like growth factor I | IGF-I | P05019 | 3479 | IGF1 | 1.11419 | 0.898697 |
| SL003674 | Bcl-2-like protein 1 | BCL2-like 1 protein | Q07817 | 598 | BCL2L1 | -1.11482 | 0.900156 |
| SL002078 | TGF-beta receptor type-2 | TGF-b R II | P37173 | 7048 | TGFBR2 | 0.926944 | 0.900156 |
| SL004891 | Heterogeneous nuclear ribonucleoproteins A2/B1 | hnRNP A2/B1 | P22626 | 3181 | HNRNPA2B1 | 0.673952 | 0.900685 |
| SL002646 | Matrix metalloproteinase-14 | MMP-14 | P50281 | 4323 | MMP14 | 0.919647 | 0.903763 |
| SL008072 | Collagen alpha-1(VIII) chain | CO8A1 | P27658 | 1295 | COL8A1 | 0.941318 | 0.90406 |
| SL005161 | Fibroblast growth factor 12 | FGF-12 | P61328 | 2257 | FGF12 | 0.55428 | 0.90406 |
| SL004304 | Syntaxin-1A | STX1a | Q16623 | 6804 | STX1A | 0.869212 | 0.90406 |
| SL000001 | Mucin-1 | MUC1 | P15941 | 4582 | MUC1 | 1.119683 | 0.90406 |
| SL008945 | Protein-glutamine gamma-glutamyltransferase E | TGM3 | Q08188 | 7053 | TGM3 | -0.212494 | 0.911278 |
| SL007223 | B-cell lymphoma 6 protein | BCL6 | P41182 | 604 | BCL6 | 0.771299 | 0.911749 |
| SL000134 | Hepatocyte growth factor receptor | Met | P08581 | 4233 | MET | -0.427952 | 0.911749 |
| SL007311 | Synaptosomal-associated protein 25 | SNP25 | P60880 | 6616 | SNAP25 | 0.134963 | 0.912768 |
| SL007242 | Dual specificity mitogen-activated protein kinase kinase 3 | MP2K3 | P46734 | 5606 | MAP2K3 | 0.125703 | 0.912768 |
| SL005190 | Leukocyte immunoglobulin-like receptor subfamily B member 1 | ILT-2 | Q8NHL6 | 10859 | LILRB1 | -0.948077 | 0.913251 |
| SL005207 | Natural cytotoxicity triggering receptor 1 | NKp46 | O76036 | 9437 | NCR1 | 1.10535 | 0.919361 |
| SL005172 | Insulin-like growth factor-binding protein 6 | IGFBP-6 | P24592 | 3489 | IGFBP6 | -1.190208 | 0.925418 |
| SL010450 | CD48 antigen | CD48 | P09326 | 962 | CD48 | -1.141134 | 0.937051 |
| SL004689 | WNT1-inducible-signaling pathway protein 1 | WISP-1 | O95388 | 8840 | WISP1 | 1.049828 | 0.938294 |
| SL014469 | SHC-transforming protein 1 | SHC1 | P29353 | 6464 | SHC1 | 0.148368 | 0.952052 |
| SL010376 | Membrane metallo-endopeptidase-like 1 | MMEL2 | Q495T6 | 79258 | MMEL1 | 1.101638 | 0.95549 |
| SL009792 | Adenylosuccinate lyase | PUR8 | P30566 | 158 | ADSL | 0.306788 | 0.960167 |
| SL010494 | Cyclin-dependent kinase 1:G2/mitotic-specific cyclin-B1 complex | CDK1/cyclin B | P06493 P14635 | 983 891 | CDC2 CCNB1 | 1.176247 | 0.960299 |
| SL009868 | FACT complex subunit SSRP1 | SSRP1 | Q08945 | 6749 | SSRP1 | 0.161151 | 0.961912 |
| SL004136 | Tyrosine-protein kinase receptor TYRO3 | Dtk | Q06418 | 7301 | TYRO3 | 0.919292 | 0.964398 |
| SL014029 | Sphingosine kinase 2 | SPHK2 | Q9NRA0 | 56848 | SPHK2 | 1.089834 | 0.965144 |
| SL014094 | Adhesion G-protein coupled receptor G5 | GP114 | Q8IZF4 | 221188 | ADGRG5 | -0.862403 | 0.965571 |
| SL012698 | Killer cell immunoglobulin-like receptor 2DL4 | KI2L4 | Q99706 | 3805 | KIR2DL4 | -0.405562 | 0.966871 |
| SL004720 | Calcineurin subunit B type 1 | Calcineurin B a | P63098 | 5534 | PPP3R1 | 0.790094 | 0.96738 |
| SL011530 | Dynein light chain roadblock-type 1 | DLRB1 | Q9NP97 | 83658 | DYNLRB1 | 0.390994 | 0.968536 |
| SL000124 | 72 kDa type IV collagenase | MMP-2 | P08253 | 4313 | MMP2 | -0.888977 | 0.969783 |
| SL010512 | PIK3CA/PIK3R1 | PIK3CA/PIK3R1 | P42336 P27986 | 5290 5295 | PIK3CA PIK3R1 | 0.810904 | 0.974269 |
| SL010470 | Semaphorin-3E | Semaphorin 3E | O15041 | 9723 | SEMA3E | 0.1906 | 0.974269 |
| SL010390 | Coiled-coil domain-containing protein 80 | URB | Q76M96 | 151887 | CCDC80 | 0.819964 | 0.974269 |
| SL007642 | Angiopoietin-related protein 4 | ANGL4 | Q9BY76 | 51129 | ANGPTL4 | 0.181465 | 0.974269 |
| SL000009 | Receptor tyrosine-protein kinase erbB-2 | ERBB2 | P04626 | 2064 | ERBB2 | -1.163703 | 0.974269 |
| SL003793 | Dual specificity mitogen-activated protein kinase kinase 1 | MEK1 | Q02750 | 5604 | MAP2K1 | 1.01976 | 0.976136 |
| SL010374 | Methionine aminopeptidase 1 | METAP1 | P53582 | 23173 | METAP1 | -0.222203 | 0.981002 |
| SL009207 | Dynactin subunit 2 | Dynactin subunit 2 | Q13561 | 10540 | DCTN2 | 0.989717 | 0.983965 |
| SL009629 | Methyl-CpG-binding domain protein 4 | MBD4 | O95243 | 8930 | MBD4 | -0.674778 | 0.985115 |
| SL005219 | Sialic acid-binding Ig-like lectin 9 | Siglec-9 | Q9Y336 | 27180 | SIGLEC9 | 0.102857 | 0.988922 |
| SL007221 | Signal transducer and activator of transcription 3 | STAT3 | P40763 | 6774 | STAT3 | 0.256912 | 0.992771 |
| SL003168 | C-C motif chemokine 27 | CTACK | Q9Y4X3 | 10850 | CCL27 | -1.127333 | 0.992771 |
| SL004271 | Glial fibrillary acidic protein | GFAP | P14136 | 2670 | GFAP | 1.112419 | 0.994638 |
| SL004078 | Bone morphogenetic protein 7 | BMP-7 | P18075 | 655 | BMP7 | -0.500965 | 0.994638 |
| SL004856 | Desmoglein-1 | Desmoglein-1 | Q02413 | 1828 | DSG1 | -0.630897 | 0.996236 |

**Table S3a.** Differential levels of the SOMAscan-screened proteins according to delirium effect at PREOP.

| **SomaId** | **TargetFullName** | **Target** | **UniProt** | **EntrezGeneID** | **EntrezGeneSymbol** | **tFC** | **p-val** |
| --- | --- | --- | --- | --- | --- | --- | --- |
| SL000546 | Prolactin | PRL | P01236 | 5617 | PRL | 2.184902 | 0.000005 |
| SL001774 | Fatty acid-binding protein, heart | FABP | P05413 | 2170 | FABP3 | 1.903164 | 0.000261 |
| SL007547 | Hepatitis A virus cellular receptor 2 | TIMD3 | Q8TDQ0 | 84868 | HAVCR2 | 1.343225 | 0.00126 |
| SL002506 | Urokinase plasminogen activator surface receptor | suPAR | Q03405 | 5329 | PLAUR | 1.290655 | 0.001441 |
| SL003167 | C-X-C motif chemokine 13 | BLC | O43927 | 10563 | CXCL13 | 1.371996 | 0.002326 |
| SL004360 | Neurotrophin-4 | Neurotrophin-5 | P34130 | 4909 | NTF4 | 1.365336 | 0.002409 |
| SL000306 | Brain natriuretic peptide 32 | BNP-32 | P16860 | 4879 | NPPB | 2.49412 | 0.003546 |
| SL003320 | Vascular endothelial growth factor D | VEGF-D | O43915 | 2277 | FIGF | 1.325242 | 0.00364 |
| SL005352 | Fatty acid-binding protein, epidermal | FABPE | Q01469 | 2171 | FABP5 | 1.352895 | 0.003694 |
| SL003970 | Parathyroid hormone | PTH | P01270 | 5741 | PTH | 1.786068 | 0.004241 |
| SL006527 | EGF-containing fibulin-like extracellular matrix protein 1 | FBLN3 | Q12805 | 2202 | EFEMP1 | 1.245849 | 0.004716 |
| SL000323 | Complement component C7 | C7 | P10643 | 730 | C7 | 1.316714 | 0.004972 |
| SL002785 | N-terminal pro-BNP | N-terminal pro-BNP | P16860 | 4879 | NPPB | 2.476798 | 0.004981 |
| SL005789 | Stanniocalcin-1 | Stanniocalcin-1 | P52823 | 6781 | STC1 | 1.305675 | 0.005209 |
| SL001992 | Tumor necrosis factor receptor superfamily member 1A | TNF sR-I | P19438 | 7132 | TNFRSF1A | 1.387836 | 0.005604 |
| SL004579 | Macrophage mannose receptor 1 | Macrophage mannose receptor | P22897 | 4360 | MRC1 | 1.297919 | 0.00565 |
| SL004712 | Stromal cell-derived factor 1 | SDF-1 | P48061 | 6387 | CXCL12 | 1.333273 | 0.006253 |
| SL000459 | Immunoglobulin A | IgA | P01876 P01877 | 3493 3494 | IGHA1 IGHA2 | 1.462692 | 0.006518 |
| SL000483 | Interleukin-7 | IL-7 | P13232 | 3574 | IL7 | 2.651973 | 0.007956 |
| SL007206 | Thrombospondin-2 | TSP2 | P35442 | 7058 | THBS2 | 1.969039 | 0.007969 |
| SL009400 | Chordin-like protein 1 | CRDL1 | Q9BU40 | 91851 | CHRDL1 | 1.334294 | 0.008092 |
| SL004837 | Inhibin beta A chain:Inhibin beta B chain heterodimer | Activin AB | P08476 P09529 | 3624 3625 | INHBA INHBB | 1.902621 | 0.008135 |
| SL004723 | Histone deacetylase 8 | HDAC8 | Q9BY41 | 55869 | HDAC8 | -1.332329 | 0.008729 |
| SL000252 | Alpha-2-macroglobulin | a2-Macroglobulin | P01023 | 2 | A2M | 1.3938 | 0.008773 |
| SL000124 | 72 kDa type IV collagenase | MMP-2 | P08253 | 4313 | MMP2 | 1.290785 | 0.009181 |
| SL007828 | Neuregulin-4 | NRG4 | Q8WWG1 | 145957 | NRG4 | 1.887455 | 0.009707 |
| SL004253 | Estradiol 17-beta-dehydrogenase 1 | 17-beta-HSD 1 | P14061 | 3292 | HSD17B1 | -1.904106 | 0.009951 |
| SL000530 | Oncostatin-M | OSM | P13725 | 5008 | OSM | -1.247379 | 0.011326 |
| SL011069 | Serine protease 27 | Marapsin | Q9BQR3 | 83886 | PRSS27 | -1.445366 | 0.011589 |
| SL005263 | alpha-2-macroglobulin receptor-associated protein | RAP | P30533 | 4043 | LRPAP1 | 1.449148 | 0.013032 |
| SL003340 | Chitinase-3-like protein 1 | YKL-40 | P36222 | 1116 | CHI3L1 | 1.952492 | 0.013345 |
| SL003680 | Advanced glycosylation end product-specific receptor, soluble | sRAGE | Q15109 | 177 | AGER | 1.617123 | 0.013371 |
| SL004925 | Anterior gradient protein 2 homolog | AGR2 | O95994 | 10551 | AGR2 | -1.149021 | 0.013784 |
| SL003735 | Tumor necrosis factor ligand superfamily member 9 | 4-1BB ligand | P41273 | 8744 | TNFSF9 | 1.345539 | 0.013814 |
| SL000523 | Collagenase 3 | MMP-13 | P45452 | 4322 | MMP13 | 1.43702 | 0.014483 |
| SL006522 | Galectin-3-binding protein | LG3BP | Q08380 | 3959 | LGALS3BP | 1.418102 | 0.014813 |
| SL008611 | N-acylethanolamine-hydrolyzing acid amidase | ASAHL | Q02083 | 27163 | NAAA | -1.501487 | 0.015068 |
| SL005797 | Polymeric immunoglobulin receptor | PIGR | P01833 | 5284 | PIGR | 1.282915 | 0.015564 |
| SL000133 | C-C motif chemokine 20 | MIP-3a | P78556 | 6364 | CCL20 | -1.896198 | 0.016218 |
| SL010381 | Dickkopf-like protein 1 | Soggy-1 | Q9UK85 | 27120 | DKKL1 | -1.174462 | 0.016333 |
| SL000591 | Metalloproteinase inhibitor 1 | TIMP-1 | P01033 | 7076 | TIMP1 | 1.246977 | 0.016529 |
| SL003331 | Matrix metalloproteinase-16 | MMP-16 | P51512 | 4325 | MMP16 | -1.71623 | 0.016919 |
| SL004843 | Ephrin-A2 | Ephrin-A2 | O43921 | 1943 | EFNA2 | 1.278909 | 0.01701 |
| SL010617 | Tryptase beta-2 | TPSB2 | P20231 | 64499 | TPSB2 | 1.466695 | 0.017459 |
| SL005169 | Intercellular adhesion molecule 5 | sICAM-5 | Q9UMF0 | 7087 | ICAM5 | 1.286923 | 0.017952 |
| SL004016 | C-X-C motif chemokine 16 | CXCL16, soluble | Q9H2A7 | 58191 | CXCL16 | 1.249345 | 0.017962 |
| SL005190 | Leukocyte immunoglobulin-like receptor subfamily B member 1 | ILT-2 | Q8NHL6 | 10859 | LILRB1 | 1.298843 | 0.01804 |
| SL019019 | PILR alpha-associated neural protein | PIANP | Q8IYJ0 | 196500 | PIANP | 1.311674 | 0.018266 |
| SL006830 | Complement factor H-related protein 5 | complement factor H-related 5 | Q9BXR6 | 81494 | CFHR5 | 1.291595 | 0.01831 |
| SL004068 | Granzyme B | Granzyme B | P10144 | 3002 | GZMB | -1.753155 | 0.019473 |
| SL010390 | Coiled-coil domain-containing protein 80 | URB | Q76M96 | 151887 | CCDC80 | 1.250178 | 0.019911 |
| SL008360 | Leucine-rich repeat transmembrane protein FLRT2 | FLRT2 | O43155 | 23768 | FLRT2 | 1.270596 | 0.020948 |
| SL005235 | Homeobox protein NANOG | NANOG | Q9H9S0 | 79923 | NANOG | 1.196426 | 0.022208 |
| SL005250 | Chymase | Chymase | P23946 | 1215 | CMA1 | -2.461898 | 0.022328 |
| SL007680 | Roundabout homolog 2 | ROBO2 | Q9HCK4 | 6092 | ROBO2 | 1.228509 | 0.022669 |
| SL000466 | Insulin-like growth factor-binding protein 2 | IGFBP-2 | P18065 | 3485 | IGFBP2 | 1.445188 | 0.023757 |
| SL000592 | Metalloproteinase inhibitor 2 | TIMP-2 | P16035 | 7077 | TIMP2 | 1.229168 | 0.024348 |
| SL003198 | Tenascin | Tenascin | P24821 | 3371 | TNC | 1.245199 | 0.025458 |
| SL000002 | Vascular endothelial growth factor A | VEGF | P15692 | 7422 | VEGFA | 1.174023 | 0.025512 |
| SL004332 | Prokineticin-1 | EG-VEGF | P58294 | 84432 | PROK1 | -1.591292 | 0.025877 |
| SL004661 | Aggrecan core protein | Aggrecan | P16112 | 176 | ACAN | 1.199711 | 0.025899 |
| SL004338 | Fibroblast growth factor 20 | FGF-20 | Q9NP95 | 26281 | FGF20 | 1.150775 | 0.02619 |
| SL008059 | 40S ribosomal protein S3 | RS3 | P23396 | 6188 | RPS3 | -2.392878 | 0.026614 |
| SL004128 | Activin receptor type-1B | Activin RIB | P36896 | 91 | ACVR1B | -2.375082 | 0.02678 |
| SL009324 | Follistatin-related protein 3 | FSTL3 | O95633 | 10272 | FSTL3 | 1.31347 | 0.026916 |
| SL000498 | Leptin | Leptin | P41159 | 3952 | LEP | 1.764685 | 0.027702 |
| SL008728 | Neurexin-3-beta | NRX3B | Q9HDB5 | 9369 | NRXN3 | 1.26483 | 0.02778 |
| SL000076 | Cyclin-dependent kinase inhibitor 1B | p27Kip1 | P46527 | 1027 | CDKN1B | -1.695379 | 0.028622 |
| SL012754 | Acid sphingomyelinase-like phosphodiesterase 3a | ASM3A | Q92484 | 10924 | SMPDL3A | -1.404357 | 0.028868 |
| SL007025 | Tyrosine-protein kinase JAK2 | JAK2 | O60674 | 3717 | JAK2 | 1.047468 | 0.029348 |
| SL016566 | Serine/threonine-protein kinase 17B | DRAK2 | O94768 | 9262 | STK17B | -2.519446 | 0.030387 |
| SL005087 | Insulin-like growth factor-binding protein 7 | IGFBP-7 | Q16270 | 3490 | IGFBP7 | 1.260923 | 0.031983 |
| SL003169 | C-X-C motif chemokine 5 | ENA-78 | P42830 | 6374 | CXCL5 | -1.213453 | 0.032208 |
| SL000027 | Prostaglandin G/H synthase 2 | COX-2 | P35354 | 5743 | PTGS2 | -1.998221 | 0.032806 |
| SL012774 | Cysteine-rich with EGF-like domain protein 1 | CREL1 | Q96HD1 | 78987 | CRELD1 | 1.195331 | 0.03314 |
| SL005161 | Fibroblast growth factor 12 | FGF-12 | P61328 | 2257 | FGF12 | -1.675407 | 0.033245 |
| SL004557 | CD59 glycoprotein | CD59 | P13987 | 966 | CD59 | 1.209182 | 0.033295 |
| SL003334 | Endothelial monocyte-activating polypeptide 2 | EMAP-2 | Q12904 | 9255 | AIMP1 | -1.195831 | 0.033344 |
| SL008865 | Proteasome subunit alpha type-2 | PSA2 | P25787 | 5683 | PSMA2 | -1.30006 | 0.034935 |
| SL010288 | Carbonic anhydrase 6 | Carbonic anhydrase 6 | P23280 | 765 | CA6 | -1.385691 | 0.035033 |
| SL005206 | Natural cytotoxicity triggering receptor 2 | NKp44 | O95944 | 9436 | NCR2 | 1.14473 | 0.035572 |
| SL000668 | Platelet glycoprotein 4 | CD36 ANTIGEN | P16671 | 948 | CD36 | 1.305517 | 0.035718 |
| SL005210 | Neuronal cell adhesion molecule | Nr-CAM | Q92823 | 4897 | NRCAM | 1.30411 | 0.036035 |
| SL000535 | Platelet-derived growth factor subunit A | PDGF-AA | P04085 | 5154 | PDGFA | -1.367616 | 0.03678 |
| SL004845 | Ephrin type-B receptor 4 | EphB4 | P54760 | 2050 | EPHB4 | -1.117532 | 0.036853 |
| SL005227 | NKG2D ligand 1 | ULBP-1 | Q9BZM6 | 80329 | ULBP1 | -1.17539 | 0.037273 |
| SL017128 | Leucine-rich repeat serine/threonine-protein kinase 2 | LRRK2 | Q5S007 | NaN | LRRK2 | -1.164815 | 0.037385 |
| SL001996 | Angiopoietin-2 | Angiopoietin-2 | O15123 | 285 | ANGPT2 | 1.530404 | 0.037544 |
| SL000517 | C-C motif chemokine 13 | MCP-4 | Q99616 | 6357 | CCL13 | -1.686276 | 0.038109 |
| SL006993 | Mitogen-activated protein kinase 13 | MK13 | O15264 | 5603 | MAPK13 | -2.391163 | 0.038683 |
| SL006119 | Trefoil factor 3 | TFF3 | Q07654 | 7033 | TFF3 | 1.292202 | 0.038684 |
| SL005209 | Neurogenic locus notch homolog protein 3 | Notch-3 | Q9UM47 | 4854 | NOTCH3 | 1.210475 | 0.038882 |
| SL011068 | Interleukin-17 receptor C | IL-17 RC | Q8NAC3 | 84818 | IL17RC | 5.457345 | 0.039659 |
| SL008414 | Ephrin type-B receptor 6 | EphB6 | O15197 | 2051 | EPHB6 | 1.289246 | 0.039909 |
| SL003930 | 15-hydroxyprostaglandin dehydrogenase [NAD(+)] | HPG- | P15428 | 3248 | HPGD | -1.140036 | 0.040158 |
| SL003280 | High mobility group protein B1 | HMG-1 | P09429 | 3146 | HMGB1 | -3.424896 | 0.040819 |
| SL006923 | TATA-box-binding protein | TBP | P20226 | 6908 | TBP | -1.601313 | 0.04135 |
| SL000022 | D-dimer | D-dimer | P02671 P02675 P02679 | 2243 2244 2266 | FGA FGB FGG | 1.181076 | 0.04149 |
| SL014470 | Breast cancer anti-estrogen resistance protein 3 | BCAR3 | O75815 | 8412 | BCAR3 | -1.207613 | 0.042046 |
| SL005764 | Scavenger receptor cysteine-rich type 1 protein M130 | sCD163 | Q86VB7 | 9332 | CD163 | 1.317395 | 0.042076 |
| SL004704 | COMM domain-containing protein 7 | COMMD7 | Q86VX2 | 149951 | COMMD7 | -1.314646 | 0.042887 |
| SL004152 | Interleukin-18 receptor 1 | IL-18 Ra | Q13478 | 8809 | IL18R1 | 1.23618 | 0.043294 |
| SL003774 | Bcl-2-like protein 2 | Apoptosis regulator Bcl-W | Q92843 | 599 | BCL2L2 | 1.637466 | 0.043423 |
| SL004258 | Adiponectin | Adiponectin | Q15848 | 9370 | ADIPOQ | 1.361058 | 0.043807 |
| SL000130 | G2/mitotic-specific cyclin-B1 | Cyclin B1 | P14635 | 891 | CCNB1 | -1.161855 | 0.044249 |
| SL001938 | Inhibin beta A chain | Activin A | P08476 | 3624 | INHBA | 1.288472 | 0.044425 |
| SL003993 | Bone morphogenetic protein 6 | BMP-6 | P22004 | 654 | BMP6 | 1.50216 | 0.044782 |
| SL000451 | Heat shock 70 kDa protein 1A | HSP 70 | P0DMV8 | 3303 | HSPA1A | 1.213136 | 0.046585 |
| SL004690 | WNT1-inducible-signaling pathway protein 3 | WISP-3 | O95389 | 8838 | WISP3 | 1.243729 | 0.046612 |
| SL003329 | C-C motif chemokine 14 | HCC-1 | Q16627 | 6358 | CCL14 | 1.319119 | 0.046624 |
| SL004720 | Calcineurin subunit B type 1 | Calcineurin B a | P63098 | 5534 | PPP3R1 | -1.14034 | 0.047212 |
| SL007696 | Complement component C1q receptor | C1QR1 | Q9NPY3 | 22918 | CD93 | 1.303635 | 0.047785 |
| SL002706 | Low affinity immunoglobulin epsilon Fc receptor | CD23 | P06734 | 2208 | FCER2 | 1.56935 | 0.048299 |
| SL005493 | Serum paraoxonase/arylesterase 1 | paraoxonase 1 | P27169 | 5444 | PON1 | -1.224229 | 0.049485 |
| SL004133 | Bone morphogenetic protein receptor type-2 | BMP RII | Q13873 | 659 | BMPR2 | -1.192095 | 0.049719 |
| SL006916 | Tyrosine-protein kinase Lck | LCK | P06239 | 3932 | LCK | -1.288115 | 0.049775 |
| SL010450 | CD48 antigen | CD48 | P09326 | 962 | CD48 | 1.50582 | 0.050672 |
| SL006924 | Agouti-related protein | ART | O00253 | 181 | AGRP | 1.19291 | 0.050941 |
| SL005156 | Neuroblastoma suppressor of tumorigenicity 1 | DAN | P41271 | 4681 | NBL1 | 1.501698 | 0.052797 |
| SL016567 | Mitogen-activated protein kinase kinase kinase 7:TGF-beta-activated kinase 1 and MAP3K7-binding protein 1 fusion | TAK1-TAB1 | O43318 Q15750 | 6885 10454 | MAP3K7 TAB1 | -1.370991 | 0.053304 |
| SL003848 | Annexin A5 | Annexin V | P08758 | 308 | ANXA5 | -1.479594 | 0.053499 |
| SL008113 | ATP synthase subunit O, mitochondrial | ATPO | P48047 | 539 | ATP5O | 1.575267 | 0.053517 |
| SL005115 | Spondin-1 | Spondin-1 | Q9HCB6 | 10418 | SPON1 | 1.395832 | 0.053893 |
| SL000474 | Interleukin-16 | IL-16 | Q14005 | 3603 | IL16 | -1.809822 | 0.054793 |
| SL004984 | 14-3-3 protein epsilon | 14-3-3E | P62258 | 7531 | YWHAE | 1.163405 | 0.055163 |
| SL000408 | Erythropoietin | Epo | P01588 | 2056 | EPO | 1.705008 | 0.055553 |
| SL000537 | Platelet-derived growth factor subunit B | PDGF-BB | P01127 | 5155 | PDGFB | -1.388428 | 0.056256 |
| SL004457 | Glia-derived nexin | Protease nexin I | P07093 | 5270 | SERPINE2 | -1.507509 | 0.057196 |
| SL008631 | Desmocollin-2 | DSC2 | Q02487 | 1824 | DSC2 | 1.360242 | 0.057471 |
| SL010498 | Ephrin type-A receptor 3 | EPHA3 | P29320 | 2042 | EPHA3 | 1.16854 | 0.057794 |
| SL003302 | C-C motif chemokine 23 | MPIF-1 | P55773 | 6368 | CCL23 | 1.240035 | 0.05784 |
| SL004139 | Ephrin-A3 | Ephrin-A3 | P52797 | 1944 | EFNA3 | 1.846788 | 0.058067 |
| SL004301 | X-ray repair cross-complementing protein 6 | Ku70 | P12956 | 2547 | XRCC6 | -2.300385 | 0.058136 |
| SL000550 | Plasma serine protease inhibitor | PCI | P05154 | 5104 | SERPINA5 | -1.119658 | 0.059095 |
| SL003301 | Ck-beta-8-1 | Ck-b-8-1 | P55773 | 6368 | CCL23 | 1.311778 | 0.059437 |
| SL010328 | Mediator of RNA polymerase II transcription subunit 1 | MED-1 | Q15648 | 5469 | MED1 | -1.124707 | 0.059486 |
| SL014208 | Cytotoxic and regulatory T-cell molecule | CRTAM | O95727 | 56253 | CRTAM | -1.47718 | 0.059807 |
| SL011588 | 40S ribosomal protein S3a | RS3A | P61247 | 6189 | RPS3A | -2.026224 | 0.060204 |
| SL003166 | CD166 antigen | ALCAM | Q13740 | 214 | ALCAM | 1.171909 | 0.060567 |
| SL004626 | A disintegrin and metalloproteinase with thrombospondin motifs 5 | ADAMTS-5 | Q9UNA0 | 11096 | ADAMTS5 | 1.118998 | 0.060837 |
| SL007033 | Latent-transforming growth factor beta-binding protein 4 | LTBP4 | Q8N2S1 | 8425 | LTBP4 | 1.229915 | 0.060891 |
| SL004867 | Carbonic anhydrase 3 | Carbonic anhydrase III | P07451 | 761 | CA3 | 1.351863 | 0.060952 |
| SL004484 | Pulmonary surfactant-associated protein D | SP-D | P35247 | 6441 | SFTPD | -4.809182 | 0.061736 |
| SL003869 | Growth/differentiation factor 15 | MIC-1 | Q99988 | 9518 | GDF15 | 1.251983 | 0.061794 |
| SL005488 | SPARC-like protein 1 | SPARCL1 | Q14515 | 8404 | SPARCL1 | 1.219829 | 0.061856 |
| SL007295 | Cadherin-15 | CAD15 | P55291 | 1013 | CDH15 | -1.224109 | 0.062455 |
| SL007752 | Death-associated protein kinase 2 | DAPK2 | Q9UIK4 | 23604 | DAPK2 | -1.458773 | 0.062517 |
| SL004208 | Annexin A1 | annexin I | P04083 | 301 | ANXA1 | -2.310173 | 0.063224 |
| SL010375 | Neutral ceramidase | ASAH2 | Q9NR71 | 56624 | ASAH2 | -1.168823 | 0.063717 |
| SL000125 | Interleukin-1 alpha | IL-1a | P01583 | 3552 | IL1A | -1.33458 | 0.064444 |
| SL003309 | Lipopolysaccharide-binding protein | LBP | P18428 | 3929 | LBP | 1.078226 | 0.064508 |
| SL001777 | Cystatin-C | Cystatin C | P01034 | 1471 | CST3 | 1.226785 | 0.065111 |
| SL010348 | Fibronectin Fragment 4 | FN1.4 | P02751 | 2335 | FN1 | 1.10032 | 0.065976 |
| SL005217 | Sialic acid-binding Ig-like lectin 6 | Siglec-6 | O43699 | 946 | SIGLEC6 | 1.147236 | 0.066178 |
| SL004671 | Tumor necrosis factor receptor superfamily member 13C | BAFF Receptor | Q96RJ3 | 115650 | TNFRSF13C | -1.350997 | 0.066363 |
| SL004591 | Granulocyte colony-stimulating factor receptor | G-CSF-R | Q99062 | 1441 | CSF3R | -1.245958 | 0.06802 |
| SL001795 | Interleukin-1 beta | IL-1b | P01584 | 3553 | IL1B | -1.358045 | 0.068491 |
| SL007871 | UMP-CMP kinase | Cytidylate kinase | P30085 | 51727 | CMPK1 | -1.092507 | 0.068889 |
| SL004768 | Allograft inflammatory factor 1 | AIF1 | P55008 | 199 | AIF1 | -1.476887 | 0.069204 |
| SL010522 | Cyclin-dependent kinase 8:Cyclin-C complex | CDK8/cyclin C | P49336 P24863 | 1024 892 | CDK8 CCNC | -1.205953 | 0.072011 |
| SL001902 | Basal Cell Adhesion Molecule | BCAM | P50895 | 4059 | BCAM | 1.15371 | 0.072042 |
| SL013488 | C-type lectin domain family 1 member B | CLC1B | Q9P126 | 51266 | CLEC1B | -1.253721 | 0.072694 |
| SL002688 | Osteopontin | Osteopontin | P10451 | 6696 | SPP1 | 1.385845 | 0.07282 |
| SL000277 | Apolipoprotein E (isoform E2) | Apo E2 | P02649 | 348 | APOE | -1.31105 | 0.073017 |
| SL004751 | Heme oxygenase 2 | HO-2 | P30519 | 3163 | HMOX2 | -1.328352 | 0.073394 |
| SL004683 | Noggin | Noggin | Q13253 | 9241 | NOG | 1.320064 | 0.073969 |
| SL004209 | Annexin A2 | annexin II | P07355 | 302 | ANXA2 | -2.748081 | 0.074245 |
| SL004305 | DNA topoisomerase 1 | Topoisomerase I | P11387 | 7150 | TOP1 | -1.604601 | 0.074618 |
| SL004708 | Connective tissue-activating peptide III | CTAP-III | P02775 | 5473 | PPBP | -1.346308 | 0.075668 |
| SL004396 | Signal transducer and activator of transcription 1-alpha/beta | STAT1 | P42224 | 6772 | STAT1 | -1.815988 | 0.075833 |
| SL012108 | 1-phosphatidylinositol 4,5-bisphosphate phosphodiesterase gamma-1 | PLCG1 | P19174 | 5335 | PLCG1 | -1.137069 | 0.075847 |
| SL011616 | Eukaryotic initiation factor 4A-III | IF4A3 | P38919 | 9775 | EIF4A3 | -1.258019 | 0.07595 |
| SL000613 | Urokinase-type plasminogen activator | uPA | P00749 | 5328 | PLAU | 1.170264 | 0.076679 |
| SL000516 | C-C motif chemokine 7 | MCP-3 | P80098 | 6354 | CCL7 | 1.175078 | 0.077281 |
| SL006374 | Cytoplasmic tyrosine-protein kinase BMX | BMX | P51813 | 660 | BMX | -1.135184 | 0.077412 |
| SL004820 | Phosphoglycerate mutase 1 | Phosphoglycerate mutase 1 | P18669 | 5223 | PGAM1 | -1.681394 | 0.077586 |
| SL006397 | Neuropilin-1 | NRP1 | O14786 | 8829 | NRP1 | 1.211579 | 0.078108 |
| SL008504 | N-acetylglucosamine-6-sulfatase | GNS | P15586 | 2799 | GNS | -1.262938 | 0.079475 |
| SL001990 | Interleukin-1 receptor antagonist protein | IL-1Ra | P18510 | 3557 | IL1RN | 1.147346 | 0.079841 |
| SL003041 | Platelet factor 4 | PF-4 | P02776 | 5196 | PF4 | -1.591916 | 0.080034 |
| SL005224 | Thrombopoietin Receptor | Thrombopoietin Receptor | P40238 | 4352 | MPL | -1.209567 | 0.080641 |
| SL004891 | Heterogeneous nuclear ribonucleoproteins A2/B1 | hnRNP A2/B1 | P22626 | 3181 | HNRNPA2B1 | -2.648764 | 0.080991 |
| SL004939 | Tyrosine-protein phosphatase non-receptor type 1 | PTP-1B | P18031 | 5770 | PTPN1 | -1.276547 | 0.08116 |
| SL005170 | Inducible T-cell costimulator | ICOS | Q9Y6W8 | 29851 | ICOS | -1.33428 | 0.081897 |
| SL002654 | Ephrin type-A receptor 2 | Epithelial cell kinase | P29317 | 1969 | EPHA2 | 1.209078 | 0.082993 |
| SL000496 | Lactotransferrin | Lactoferrin | P02788 | 4057 | LTF | -1.530483 | 0.084378 |
| SL007651 | Fibroblast growth factor 23 | FGF23 | Q9GZV9 | 8074 | FGF23 | 1.856063 | 0.085584 |
| SL004143 | GDNF family receptor alpha-2 | GFRa-2 | O00451 | 2675 | GFRA2 | 1.148244 | 0.087835 |
| SL003172 | C-X-C motif chemokine 6 | GCP-2 | P80162 | 6372 | CXCL6 | -1.287178 | 0.090274 |
| SL013165 | Non-histone chromosomal protein HMG-14 | HMGN1 | P05114 | 3150 | HMGN1 | -1.219652 | 0.090778 |
| SL000139 | Epiregulin | EPI | O14944 | 2069 | EREG | -1.460007 | 0.090792 |
| SL002783 | Cardiotrophin-1 | Cardiotrophin-1 | Q16619 | 1489 | CTF1 | -1.158748 | 0.092124 |
| SL004872 | Tumor necrosis factor receptor superfamily member EDAR | EDAR | Q9UNE0 | 10913 | EDAR | -1.279427 | 0.092278 |
| SL000283 | Beta-2-microglobulin | b2-Microglobulin | P61769 | 567 | B2M | 1.172171 | 0.093364 |
| SL007272 | Hexokinase-2 | HXK2 | P52789 | 3099 | HK2 | -3.420292 | 0.093476 |
| SL004698 | Protein E7_HPV18 | HPV E7 Type18 | P06788 | 1489089 | Human-virus | -1.089213 | 0.093488 |
| SL011404 | cAMP-specific 3',5'-cyclic phosphodiesterase 4D | PDE4D | Q08499 | 5144 | PDE4D | -1.127295 | 0.094479 |
| SL004726 | CD97 antigen | CD97 | P48960 | 976 | CD97 | -1.091683 | 0.09464 |
| SL002519 | Receptor tyrosine-protein kinase erbB-3 | ERBB3 | P21860 | 2065 | ERBB3 | 1.187701 | 0.094702 |
| SL004668 | Apolipoprotein E (isoform E3) | Apo E3 | P02649 | 348 | APOE | -1.436478 | 0.096135 |
| SL000598 | Thrombopoietin | Tpo | P40225 | 7066 | THPO | 1.314398 | 0.096452 |
| SL004511 | Bactericidal permeability-increasing protein | BPI | P17213 | 671 | BPI | -2.104828 | 0.097113 |
| SL005160 | Endothelial cell-selective adhesion molecule | ESAM | Q96AP7 | 90952 | ESAM | 1.154251 | 0.098759 |
| SL008967 | Tyrosine-protein phosphatase non-receptor type substrate 1 | SHPS1 | P78324 | 140885 | SIRPA | 1.268186 | 0.098919 |
| SL011528 | 40S ribosomal protein S7 | RS7 | P62081 | 6201 | RPS7 | -3.243558 | 0.098947 |
| SL010519 | Tyrosine-protein kinase ZAP-70 | ZAP70 | P43403 | 7535 | ZAP70 | -1.134594 | 0.100452 |
| SL003199 | Tyrosine-protein kinase receptor Tie-1, soluble | sTie-1 | P35590 | 7075 | TIE1 | 1.184025 | 0.10051 |
| SL005730 | cGMP-dependent 3',5'-cyclic phosphodiesterase | cGMP-stimulated PDE | O00408 | 5138 | PDE2A | -1.130468 | 0.100978 |
| SL003761 | Phosphatidylinositol 3,4,5-trisphosphate 3-phosphatase and dual-specificity protein phosphatase PTEN | pTEN | P60484 | 5728 | PTEN | 1.588013 | 0.101941 |
| SL000658 | Growth arrest-specific protein 1 | GAS1 | P54826 | 2619 | GAS1 | 1.158458 | 0.10254 |
| SL005102 | Sex hormone-binding globulin | SHBG | P04278 | 6462 | SHBG | 1.238938 | 0.103287 |
| SL003191 | Neutrophil-activating peptide 2 | NAP-2 | P02775 | 5473 | PPBP | -1.2608 | 0.104601 |
| SL009791 | Heterogeneous nuclear ribonucleoprotein A/B | hnRNP A/B | Q99729 | 3182 | HNRNPAB | -1.753965 | 0.104876 |
| SL004669 | Apolipoprotein E (isoform E4) | Apo E4 | P02649 | 348 | APOE | -1.341617 | 0.107829 |
| SL007280 | Dipeptidyl peptidase 1 | CATC | P53634 | 1075 | CTSC | 4.181129 | 0.108386 |
| SL007250 | Phosphatidylinositol 4,5-bisphosphate 3-kinase catalytic subunit gamma isoform | PK3CG | P48736 | 5294 | PIK3CG | -1.151591 | 0.109948 |
| SL006912 | Tyrosine-protein kinase Fgr | FGR | P09769 | 2268 | FGR | -2.323609 | 0.11006 |
| SL000338 | Calpastatin | Calpastatin | P20810 | 831 | CAST | 1.428816 | 0.111589 |
| SL017106 | Interferon alpha-10 | IFN10 | P01566 | 3446 | IFNA10 | 1.668953 | 0.111808 |
| SL004363 | Stem cell growth factor-alpha | SCGF-alpha | Q9Y240 | 6320 | CLEC11A | 1.310949 | 0.112059 |
| SL003341 | Fibrinogen gamma chain | Fibrinogen g-chain dimer | P02679 | 2266 | FGG | 1.109299 | 0.112092 |
| SL003918 | Kallikrein-13 | kallikrein 13 | Q9UKR3 | 26085 | KLK13 | -1.405889 | 0.113056 |
| SL004610 | Low-density lipoprotein receptor-related protein 8 | LRP8 | Q14114 | 7804 | LRP8 | 1.156606 | 0.113345 |
| SL000481 | Interleukin-5 | IL-5 | P05113 | 3567 | IL5 | -1.310798 | 0.113423 |
| SL016554 | High affinity cGMP-specific 3',5'-cyclic phosphodiesterase 9A | PDE9A | O76083 | 5152 | PDE9A | 1.108538 | 0.113938 |
| SL006230 | Lumican | Lumican | P51884 | 4060 | LUM | 1.21507 | 0.114014 |
| SL018587 | Immunoglobulin superfamily containing leucine-rich repeat protein 2 | ISLR2 | Q6UXK2 | 57611 | ISLR2 | 1.162303 | 0.114256 |
| SL004362 | Stem Cell Growth Factor-beta | SCGF-beta | Q9Y240 | 6320 | CLEC11A | 1.278652 | 0.114771 |
| SL009628 | Inhibitor of growth protein 1 | ING1 | Q9UK53 | 3621 | ING1 | -1.287028 | 0.115138 |
| SL016828 | Epidermal growth factor receptor variant III | EGFRvIII | P00533 | NaN | EGFR | -1.293396 | 0.116554 |
| SL007361 | Galectin-10 | LPPL | Q05315 | 1178 | CLC | -1.385528 | 0.116609 |
| SL007869 | Peptidyl-prolyl cis-trans isomerase B | PPIB | P23284 | 5479 | PPIB | -1.244096 | 0.118276 |
| SL011535 | RNA-binding protein 39 | RBM39 | Q14498 | 9584 | RBM39 | -1.369949 | 0.119095 |
| SL010928 | Inosine-5'-monophosphate dehydrogenase 2 | IMDH2 | P12268 | 3615 | IMPDH2 | -1.347752 | 0.119447 |
| SL000138 | Heparin-binding EGF-like growth factor | HB-EGF | Q99075 | 1839 | HBEGF | -1.144244 | 0.119489 |
| SL004304 | Syntaxin-1A | STX1a | Q16623 | 6804 | STX1A | -1.311956 | 0.119813 |
| SL000249 | Alpha-1-antitrypsin | a1-Antitrypsin | P01009 | 5265 | SERPINA1 | 1.165978 | 0.119935 |
| SL003349 | Hypoxia-inducible factor 1-alpha | HIF-1a | Q16665 | 3091 | HIF1A | 1.226632 | 0.120131 |
| SL005191 | Leukocyte immunoglobulin-like receptor subfamily B member 2 | ILT-4 | Q8N423 | 10288 | LILRB2 | 1.236145 | 0.122667 |
| SL000062 | Prostate-specific antigen | PSA | P07288 | 354 | KLK3 | -1.4552 | 0.123058 |
| SL006114 | Tyrosine-protein kinase transmembrane receptor ROR1 | ROR1 | Q01973 | 4919 | ROR1 | 1.189592 | 0.123325 |
| SL017610 | Gro-beta/gamma | Gro-b/g | P19876 P19875 | NaN | CXCL3 CXCL2 | -1.082392 | 0.123587 |
| SL014289 | Killer cell immunoglobulin-like receptor 3DS1 | KI3S1 | Q14943 | 3813 | KIR3DS1 | -1.19672 | 0.12457 |
| SL014270 | CMRF35-like molecule 6 | CLM6 | Q08708 | 10871 | CD300C | 1.162648 | 0.124644 |
| SL008402 | Ly6/PLAUR domain-containing protein 3 | LYPD3 | O95274 | 27076 | LYPD3 | 1.18336 | 0.125545 |
| SL004739 | Inter-alpha-trypsin inhibitor heavy chain H4 | ITI heavy chain H4 | Q14624 | 3700 | ITIH4 | 1.118418 | 0.125642 |
| SL000325 | Complement component C9 | C9 | P02748 | 735 | C9 | 1.102813 | 0.125783 |
| SL012698 | Killer cell immunoglobulin-like receptor 2DL4 | KI2L4 | Q99706 | 3805 | KIR2DL4 | -1.361751 | 0.126633 |
| SL004297 | Neuregulin-1 | NEUREGULIN-1 | Q02297 | 3084 | NRG1 | 5.346371 | 0.128379 |
| SL004863 | Tumor necrosis factor receptor superfamily member 19 | TAJ | Q9NS68 | 55504 | TNFRSF19 | 1.284335 | 0.128551 |
| SL008591 | Trefoil factor 1 | TFF1 | P04155 | 7031 | TFF1 | 1.299537 | 0.13058 |
| SL000038 | C-C motif chemokine 2 | MCP-1 | P13500 | 6347 | CCL2 | -2.685862 | 0.132017 |
| SL004838 | Leukocyte surface antigen CD47 | CD47 | Q08722 | 961 | CD47 | -1.169962 | 0.132737 |
| SL002036 | Fibroblast growth factor receptor 4 | FGFR4 | P22455 | 2264 | FGFR4 | -1.3172 | 0.133738 |
| SL008759 | Platelet glycoprotein VI | GPVI | Q9HCN6 | 51206 | GP6 | -1.319701 | 0.134003 |
| SL001998 | Tissue factor pathway inhibitor | TFPI | P10646 | 7035 | TFPI | 1.199732 | 0.134077 |
| SL004637 | Macrophage-stimulating protein receptor | MSP R | Q04912 | 4486 | MST1R | -1.112311 | 0.135503 |
| SL004685 | Persephin | Persephin | O60542 | 5623 | PSPN | -1.141013 | 0.135861 |
| SL018887 | Quinone oxidoreductase-like protein 1 | QORL1 | O95825 | 9946 | CRYZL1 | -1.29578 | 0.136763 |
| SL007774 | Protein jagged-2 | JAG2 | Q9Y219 | 3714 | JAG2 | -1.630937 | 0.137242 |
| SL010471 | Testican-2 | Testican-2 | Q92563 | 9806 | SPOCK2 | 1.229912 | 0.13806 |
| SL000563 | C-C motif chemokine 5 | RANTES | P13501 | 6352 | CCL5 | -1.272489 | 0.138195 |
| SL005218 | Sialic acid-binding Ig-like lectin 7 | Siglec-7 | Q9Y286 | 27036 | SIGLEC7 | 1.157363 | 0.138249 |
| SL006922 | DNA repair protein RAD51 homolog 1 | RAD51 | Q06609 | 5888 | RAD51 | 1.349339 | 0.13865 |
| SL000051 | C-reactive protein | CRP | P02741 | 1401 | CRP | 1.307463 | 0.138805 |
| SL014070 | SLIT and NTRK-like protein 5 | SLIK5 | O94991 | 26050 | SLITRK5 | 1.197605 | 0.139098 |
| SL007145 | Nuclear receptor subfamily 1 group D member 1 | NR1D1 | P20393 | 9572 | NR1D1 | -1.091967 | 0.140321 |
| SL011772 | Pescadillo homolog | PESC | O00541 | 23481 | PES1 | -1.152366 | 0.140411 |
| SL004335 | Fibroblast growth factor 17 | FGF-17 | O60258 | 8822 | FGF17 | 1.133541 | 0.14075 |
| SL014088 | Fc receptor-like protein 3 | FCRL3 | Q96P31 | 115352 | FCRL3 | 1.098256 | 0.141227 |
| SL005675 | ATP synthase subunit beta, mitochondrial | ATP synthase beta chain | P06576 | 506 | ATP5B | -1.726258 | 0.141541 |
| SL003726 | Serine/threonine-protein kinase Chk2 | Chk2 | O96017 | 11200 | CHEK2 | -1.893209 | 0.141732 |
| SL003177 | Intercellular adhesion molecule 2 | sICAM-2 | P13598 | 3384 | ICAM2 | -1.156755 | 0.141921 |
| SL004814 | Coactosin-like protein | Coactosin-like protein | Q14019 | 23406 | COTL1 | -1.086876 | 0.142692 |
| SL014735 | Inosine-5'-monophosphate dehydrogenase 1 | IMDH1 | P20839 | 3614 | IMPDH1 | -2.020899 | 0.143049 |
| SL004345 | Growth/differentiation factor 11 | GDF-11 | O95390 | 10220 | GDF11 | 1.825525 | 0.144058 |
| SL007195 | CD70 antigen | CD70 | P32970 | 970 | CD70 | 1.098065 | 0.144308 |
| SL010619 | Tryptase gamma | TPSG1 | Q9NRR2 | 25823 | TPSG1 | 1.110847 | 0.144669 |
| SL010509 | Serine/threonine-protein kinase PAK 6 | PAK6 | Q9NQU5 | 56924 | PAK6 | -1.756353 | 0.145175 |
| SL014094 | Adhesion G-protein coupled receptor G5 | GP114 | Q8IZF4 | 221188 | ADGRG5 | 1.206554 | 0.145182 |
| SL005196 | Limbic system-associated membrane protein | LSAMP | Q13449 | 4045 | LSAMP | 1.143181 | 0.145279 |
| SL000515 | C-C motif chemokine 8 | MCP-2 | P80075 | 6355 | CCL8 | 1.260854 | 0.145673 |
| SL000525 | Matrilysin | MMP-7 | P09237 | 4316 | MMP7 | 1.272728 | 0.147381 |
| SL008904 | Lymphatic vessel endothelial hyaluronic acid receptor 1 | LYVE1 | Q9Y5Y7 | 10894 | LYVE1 | 1.27972 | 0.147431 |
| SL008916 | Dipeptidyl peptidase 2 | DPP2 | Q9UHL4 | 29952 | DPP7 | -1.114197 | 0.147718 |
| SL003650 | Alpha-enolase | Alpha enolase | P06733 | 2023 | ENO1 | -1.424077 | 0.147891 |
| SL018256 | Choline/ethanolamine kinase | CHKB | Q9Y259 | 1120 | CHKB | -1.111636 | 0.147894 |
| SL006805 | Serine/threonine-protein kinase MRCK beta | MRCKB | Q9Y5S2 | 9578 | CDC42BPB | -1.097917 | 0.148031 |
| SL003193 | C-C motif chemokine 21 | 6Ckine | O00585 | 6366 | CCL21 | 1.344657 | 0.148617 |
| SL007729 | Endoplasmic reticulum aminopeptidase 1 | ARTS1 | Q9NZ08 | 51752 | ERAP1 | 1.156347 | 0.148839 |
| SL005225 | High affinity nerve growth factor receptor | TrkA | P04629 | 4914 | NTRK1 | -1.396427 | 0.148842 |
| SL005152 | Retinoic acid receptor responder protein 2 | TIG2 | Q99969 | 5919 | RARRES2 | 1.137081 | 0.148976 |
| SL007022 | Heterogeneous nuclear ribonucleoprotein Q | HNRPQ | O60506 | 10492 | SYNCRIP | -1.151813 | 0.149085 |
| SL006131 | Fatty acid-binding protein, liver | FABPL | P07148 | 2168 | FABP1 | 1.48894 | 0.149087 |
| SL012457 | Neurexophilin-1 | NXPH1 | P58417 | 30010 | NXPH1 | -1.224468 | 0.149588 |
| SL007281 | Mitogen-activated protein kinase 12 | MK12 | P53778 | 6300 | MAPK12 | -1.138464 | 0.149859 |
| SL001995 | Angiopoietin-1 | Angiopoietin-1 | Q15389 | 284 | ANGPT1 | -1.266038 | 0.149976 |
| SL001815 | Superoxide dismutase [Mn], mitochondrial | Mn SOD | P04179 | 6648 | SOD2 | -1.134312 | 0.151608 |
| SL004648 | Tumor necrosis factor ligand superfamily member 14 | LIGHT | O43557 | 8740 | TNFSF14 | -1.140068 | 0.152324 |
| SL010465 | Matrilin-2 | MATN2 | O00339 | 4147 | MATN2 | 1.17702 | 0.153397 |
| SL004155 | Platelet-derived growth factor receptor beta | PDGF Rb | P09619 | 5159 | PDGFRB | 1.222333 | 0.154372 |
| SL003197 | C-C motif chemokine 25 | TECK | O15444 | 6370 | CCL25 | 1.204496 | 0.155991 |
| SL004141 | Ephrin-A5 | Ephrin-A5 | P52803 | 1946 | EFNA5 | 1.140735 | 0.156732 |
| SL000633 | Tumor necrosis factor ligand superfamily member 6, soluble form | Fas ligand, soluble | P48023 | 356 | FASLG | 1.157323 | 0.157506 |
| SL000164 | Myoglobin | Myoglobin | P02144 | 4151 | MB | 1.153999 | 0.157813 |
| SL014292 | Sialic acid-binding Ig-like lectin 14 | SIG14 | Q08ET2 | 100049587 | SIGLEC14 | 1.211033 | 0.160375 |
| SL008609 | Low affinity immunoglobulin gamma Fc region receptor III-B | FCG3B | O75015 | 2215 | FCGR3B | 1.156403 | 0.161323 |
| SL004660 | Bone sialoprotein 2 | BSP | P21815 | 3381 | IBSP | 1.324314 | 0.162108 |
| SL010502 | Mitogen-activated protein kinase 8 | MK08 | P45983 | 5599 | MAPK8 | -1.551184 | 0.162554 |
| SL000247 | 6-phosphogluconate dehydrogenase, decarboxylating | 6-Phosphogluconate dehydrogenase | P52209 | 5226 | PGD | -1.669255 | 0.163771 |
| SL000007 | Estrogen receptor | ER | P03372 | 2099 | ESR1 | -1.105024 | 0.164549 |
| SL009868 | FACT complex subunit SSRP1 | SSRP1 | Q08945 | 6749 | SSRP1 | -1.268887 | 0.164766 |
| SL000017 | von Willebrand factor | vWF | P04275 | 7450 | VWF | 1.265811 | 0.165105 |
| SL004306 | SUMO-conjugating enzyme UBC9 | UBC9 | P63279 | 7329 | UBE2I | -1.489545 | 0.166076 |
| SL002646 | Matrix metalloproteinase-14 | MMP-14 | P50281 | 4323 | MMP14 | -1.427018 | 0.167401 |
| SL003653 | Phosphoglycerate kinase 1 | phosphoglycerate kinase 1 | P00558 | 5230 | PGK1 | -1.410649 | 0.168322 |
| SL010515 | Ribosomal protein S6 kinase alpha-3 | RPS6KA3 | P51812 | 6197 | RPS6KA3 | -1.473961 | 0.168325 |
| SL001797 | Kallikrein-6 | Kallikrein 6 | Q92876 | 5653 | KLK6 | -1.090913 | 0.16864 |
| SL004356 | C-C motif chemokine 4-like | LAG-1 | Q8NHW4 | 388372 | CCL4L1 | -1.249598 | 0.169517 |
| SL000403 | Endostatin | Endostatin | P39060 | 80781 | COL18A1 | 1.168057 | 0.170294 |
| SL003703 | Bcl-2-related protein A1 | BFL1 | Q16548 | 597 | BCL2A1 | -1.065545 | 0.170609 |
| SL003872 | Interleukin-6 receptor subunit beta | gp130, soluble | P40189 | 3572 | IL6ST | 1.148497 | 0.170842 |
| SL005157 | CD209 antigen | DC-SIGN | Q9NNX6 | 30835 | CD209 | 1.167073 | 0.170904 |
| SL005233 | Tumor necrosis factor receptor superfamily member 27 | XEDAR | Q9HAV5 | 60401 | EDA2R | 1.108682 | 0.172285 |
| SL004078 | Bone morphogenetic protein 7 | BMP-7 | P18075 | 655 | BMP7 | 1.144692 | 0.172451 |
| SL014096 | Leucine-rich repeat transmembrane neuronal protein 1 | LRRT1 | Q86UE6 | 347730 | LRRTM1 | 1.217407 | 0.172457 |
| SL003060 | Fibroblast growth factor receptor 1 | bFGF-R | P11362 | 2260 | FGFR1 | 1.144322 | 0.172898 |
| SL003362 | Complement C3d fragment | C3d | P01024 | 718 | C3 | 1.167955 | 0.173785 |
| SL007207 | Thrombospondin-4 | TSP4 | P35443 | 7060 | THBS4 | 1.205793 | 0.175317 |
| SL003327 | Complement factor D | Factor D | P00746 | 1675 | CFD | 1.12436 | 0.17568 |
| SL004868 | Carbonic anhydrase 7 | Carbonic anhydrase VII | P43166 | 766 | CA7 | 1.140333 | 0.176947 |
| SL003182 | Integrin alpha-V: beta-5 complex | Integrin aVb5 | P06756, P18084 | 3685 3693 | ITGAV ITGB5 | 1.240688 | 0.177364 |
| SL002702 | Serine/threonine-protein kinase pim-1 | PIM1 | P11309 | 5292 | PIM1 | -1.89601 | 0.177761 |
| SL002528 | Phospholipase A2, membrane associated | NPS-PLA2 | P14555 | 5320 | PLA2G2A | 1.229773 | 0.17872 |
| SL007274 | Ephrin-B2 | EFNB2 | P52799 | 1948 | EFNB2 | 1.197112 | 0.178904 |
| SL004871 | Tumor necrosis factor receptor superfamily member 21 | DR6 | O75509 | 27242 | TNFRSF21 | 1.151033 | 0.179105 |
| SL003187 | C-C motif chemokine 22 | MDC | O00626 | 6367 | CCL22 | 1.1566 | 0.179167 |
| SL004636 | Receptor-type tyrosine-protein kinase FLT3 | Flt-3 | P36888 | 2322 | FLT3 | -1.329642 | 0.179259 |
| SL007284 | Cysteine-rich secretory protein 3 | CRIS3 | P54108 | 10321 | CRISP3 | -2.114774 | 0.179743 |
| SL002792 | beta-adrenergic receptor kinase 1 | BARK1 | P25098 | 156 | ADRBK1 | -1.913865 | 0.181248 |
| SL014308 | Fibroblast growth factor 8 isoform A | FGF-8A | P55075 | 2253 | FGF8 | -1.417643 | 0.181344 |
| SL002505 | Atrial natriuretic factor | ANP | P01160 | 4878 | NPPA | 1.194145 | 0.181468 |
| SL001720 | Vascular cell adhesion protein 1 | VCAM-1 | P19320 | 7412 | VCAM1 | 1.193457 | 0.181767 |
| SL006915 | Interleukin-27 | IL-27 | Q8NEV9 Q14213 | 246778 10148 | IL27 EBI3 | 1.164015 | 0.182077 |
| SL008094 | Phosphoglucomutase-1 | PGM1 | P36871 | 5236 | PGM1 | 1.292485 | 0.182508 |
| SL010460 | Low affinity immunoglobulin gamma Fc region receptor II-b | FCG2B | P31994 | 2213 | FCGR2B | 1.267693 | 0.182562 |
| SL007324 | Casein kinase II subunit alpha | CSK21 | P68400 | 1457 | CSNK2A1 | -1.219078 | 0.182616 |
| SL003915 | Kallikrein-8 | kallikrein 8 | O60259 | 11202 | KLK8 | 1.168642 | 0.182815 |
| SL014091 | VPS10 domain-containing receptor SorCS2 | SORC2 | Q96PQ0 | 57537 | SORCS2 | -6.620584 | 0.183216 |
| SL000319 | Complement C5 | C5 | P01031 | 727 | C5 | 1.104049 | 0.183299 |
| SL007429 | Transmembrane glycoprotein NMB | GPNMB | Q14956 | 10457 | GPNMB | 1.153874 | 0.183411 |
| SL011530 | Dynein light chain roadblock-type 1 | DLRB1 | Q9NP97 | 83658 | DYNLRB1 | -1.322914 | 0.184014 |
| SL009341 | Basigin | BASI | P35613 | 682 | BSG | -1.082555 | 0.185082 |
| SL010454 | Contactin-4 | Contactin-4 | Q8IWV2 | 152330 | CNTN4 | 1.189621 | 0.185693 |
| SL000558 | Prothrombin | Prothrombin | P00734 | 2147 | F2 | -1.061027 | 0.187363 |
| SL004153 | Macrophage colony-stimulating factor 1 receptor | M-CSF R | P07333 | 1436 | CSF1R | 1.13236 | 0.188378 |
| SL014111 | Kin of IRRE-like protein 3 | KIRR3 | Q8IZU9 | 84623 | KIRREL3 | -1.116839 | 0.189235 |
| SL003080 | Macrophage migration inhibitory factor | MIF | P14174 | 4282 | MIF | -1.21561 | 0.189641 |
| SL004482 | Endoglin | Endoglin | P17813 | 2022 | ENG | 1.185121 | 0.189833 |
| SL006694 | Beta-Ala-His dipeptidase | CNDP1 | Q96KN2 | 84735 | CNDP1 | -1.169181 | 0.190618 |
| SL014069 | Proprotein convertase subtilisin/kexin type 7 | PCSK7 | Q16549 | 9159 | PCSK7 | -1.11091 | 0.192326 |
| SL000581 | Superoxide dismutase [Cu-Zn] | SOD | P00441 | 6647 | SOD1 | -1.422894 | 0.192491 |
| SL004008 | Myeloblastin | Proteinase-3 | P24158 | 5657 | PRTN3 | -1.484051 | 0.193618 |
| SL000522 | Macrophage metalloelastase | MMP-12 | P39900 | 4321 | MMP12 | 1.166407 | 0.193745 |
| SL020172 | Relaxin receptor 1 | RXFP1 | Q9HBX9 | NaN | RXFP1 | -1.257802 | 0.195256 |
| SL010391 | WAP, Kazal, immunoglobulin, Kunitz and NTR domain-containing protein 2 | WFKN2 | Q8TEU8 | 124857 | WFIKKN2 | 1.176127 | 0.195265 |
| SL004862 | Programmed cell death 1 ligand 2 | PD-L2 | Q9BQ51 | 80380 | PDCD1LG2 | -1.127551 | 0.195332 |
| SL002705 | Thrombospondin-1 | Thrombospondin-1 | P07996 | 7057 | THBS1 | -1.236417 | 0.195375 |
| SL005256 | Histone H1.2 | Histone H1.2 | P16403 | 3006 | HIST1H1C | -1.567794 | 0.195732 |
| SL004458 | Elafin | Elafin | P19957 | 5266 | PI3 | 1.275093 | 0.196765 |
| SL004137 | Ephrin type-A receptor 1 | EphA1 | P21709 | 2041 | EPHA1 | 1.20813 | 0.197937 |
| SL005178 | Interleukin-37 | IL-1F7 | Q9NZH6 | 27178 | IL37 | -1.07814 | 0.199109 |
| SL014684 | 2'-5'-oligoadenylate synthase 1 | OAS1 | P00973 | 4938 | OAS1 | -1.23177 | 0.199172 |
| SL000458 | Interferon gamma receptor 1 | IFN-g R1 | P15260 | 3459 | IFNGR1 | 1.311206 | 0.200084 |
| SL004136 | Tyrosine-protein kinase receptor TYRO3 | Dtk | Q06418 | 7301 | TYRO3 | 1.290105 | 0.201733 |
| SL009349 | Follistatin-related protein 1 | FSTL1 | Q12841 | 11167 | FSTL1 | 1.105088 | 0.201803 |
| SL004359 | Neurotrophin-3 | Neurotrophin-3 | P20783 | 4908 | NTF3 | 1.174601 | 0.202165 |
| SL005231 | Netrin receptor UNC5D | UNC5H4 | Q6UXZ4 | 137970 | UNC5D | 1.155867 | 0.202381 |
| SL011888 | SPARC-related modular calcium-binding protein 1 | SMOC1 | Q9H4F8 | 64093 | SMOC1 | 1.224139 | 0.20335 |
| SL014071 | Leucine-rich repeat transmembrane protein FLRT1 | FLRT1 | Q9NZU1 | 23769 | FLRT1 | -1.105722 | 0.203838 |
| SL000582 | Baculoviral IAP repeat-containing protein 5 | Survivin | O15392 | 332 | BIRC5 | -1.081516 | 0.203974 |
| SL005629 | N-acetyl-D-glucosamine kinase | NAGK | Q9UJ70 | 55577 | NAGK | -1.280633 | 0.204753 |
| SL007173 | Granulins | GRN | P28799 | 2896 | GRN | 1.102069 | 0.205678 |
| SL010493 | Calcium/calmodulin-dependent protein kinase type II subunit delta | CAMK2D | Q13557 | 817 | CAMK2D | -1.649457 | 0.205781 |
| SL001880 | Interferon beta | IFN-b | P01574 | 3456 | IFNB1 | 1.447144 | 0.20668 |
| SL003179 | Integrin alpha-I: beta-1 complex | Integrin a1b1 | P56199, P05556 | 3672 3688 | ITGA1 ITGB1 | 1.084382 | 0.207072 |
| SL000572 | Serum amyloid A-1 protein | SAA | P0DJI8 | 6288 | SAA1 | 2.603783 | 0.207381 |
| SL016553 | cGMP-inhibited 3',5'-cyclic phosphodiesterase A | PDE3A | Q14432 | 5139 | PDE3A | -1.134238 | 0.208717 |
| SL005188 | Interleukin-5 receptor subunit alpha | IL-5 Ra | Q01344 | 3568 | IL5RA | 1.122259 | 0.208818 |
| SL004750 | ATP-dependent RNA helicase DDX19B | DEAD-box protein 19B | Q9UMR2 | 11269 | DDX19B | -1.242184 | 0.209422 |
| SL004853 | ICOS ligand | B7-H2 | O75144 | 23308 | ICOSLG | -2.904334 | 0.209466 |
| SL010520 | Aurora kinase B | AURKB | Q96GD4 | 9212 | AURKB | -1.050078 | 0.210886 |
| SL005230 | Netrin receptor UNC5C | UNC5H3 | O95185 | 8633 | UNC5C | 1.143726 | 0.211765 |
| SL004646 | Layilin | Layilin | Q6UX15 | 143903 | LAYN | 1.094714 | 0.211996 |
| SL000601 | Serotransferrin | Transferrin | P02787 | 7018 | TF | -1.049532 | 0.212636 |
| SL003764 | Neural cell adhesion molecule 1, 120 kDa isoform | NCAM-120 | P13591 | 4684 | NCAM1 | 1.142132 | 0.212916 |
| SL019978 | Interferon alpha-7 | IFNA7 | P01567 | NaN | IFNA7 | 17.104677 | 0.21309 |
| SL008837 | CD63 antigen | CD63 | P08962 | 967 | CD63 | -1.113676 | 0.213196 |
| SL000276 | Apolipoprotein E | Apo E | P02649 | 348 | APOE | -1.47409 | 0.21361 |
| SL013548 | Interleukin-34 | IL-34 | Q6ZMJ4 | 146433 | IL34 | 1.126691 | 0.21562 |
| SL011708 | cAMP-regulated phosphoprotein 19 | ARP19 | P56211 | 10776 | ARPP19 | -1.280391 | 0.215983 |
| SL004672 | Tumor necrosis factor receptor superfamily member 17 | BCMA | Q02223 | 608 | TNFRSF17 | 1.118678 | 0.216659 |
| SL008936 | Neural cell adhesion molecule L1-like protein | CHL1 | O00533 | 10752 | CHL1 | 1.137446 | 0.217913 |
| SL005575 | Galactoside 3(4)-L-fucosyltransferase | Fucosyltransferase 3 | P21217 | 2525 | FUT3 | 1.093512 | 0.218072 |
| SL004357 | Growth/differentiation factor 8 | Myostatin | O14793 | 2660 | MSTN | -1.124922 | 0.218437 |
| SL010523 | Homeodomain-interacting protein kinase 3 | HIPK3 | Q9H422 | 10114 | HIPK3 | -1.221793 | 0.218771 |
| SL000428 | Follicle stimulating hormone | FSH | P01215, P01225 | 1081 2488 | CGA FSHB | 1.386173 | 0.218925 |
| SL014488 | Proto-oncogene vav | VAV | P15498 | 7409 | VAV1 | -1.768109 | 0.219484 |
| SL016550 | Casein kinase II 2-alpha:2-beta heterotetramer | CK2-A1:B | P68400 P67870 | 1457 1460 | CSNK2A1 CSNK2B | -1.201977 | 0.220001 |
| SL004725 | Histone acetyltransferase type B catalytic subunit | Hat1 | O14929 | 8520 | HAT1 | -1.170648 | 0.220357 |
| SL005084 | Periostin | Periostin | Q15063 | 10631 | POSTN | 1.132308 | 0.220501 |
| SL000615 | Vasoactive Intestinal Peptide | Vasoactive Intestinal Peptide | P01282 | 7432 | VIP | -1.171342 | 0.22173 |
| SL010349 | Fibronectin Fragment 3 | FN1.3 | P02751 | 2335 | FN1 | 1.154781 | 0.2218 |
| SL018921 | HemK methyltransferase family member 2 | HEMK2 | Q9Y5N5 | 29104 | N6AMT1 | -1.262233 | 0.223038 |
| SL000597 | Lymphotoxin-alpha | TNF-b | P01374 | 4049 | LTA | -1.127362 | 0.223453 |
| SL002695 | Cytosolic non-specific dipeptidase | Glutamate carboxypeptidase | Q96KP4 | 55748 | CNDP2 | -1.067087 | 0.223779 |
| SL005685 | Elongation factor 1-beta | EF-1-beta | P24534 | 1933 | EEF1B2 | -1.229314 | 0.224086 |
| SL012707 | Proprotein convertase subtilisin/kexin type 9 | PCSK9 | Q8NBP7 | 255738 | PCSK9 | 1.146289 | 0.22433 |
| SL003690 | Tumor necrosis factor receptor superfamily member 11A | RANK | Q9Y6Q6 | 8792 | TNFRSF11A | -1.251052 | 0.224339 |
| SL000424 | Fibrinogen | Fibrinogen | P02671 P02675 P02679 | 2243 2244 2266 | FGA FGB FGG | 1.029746 | 0.225259 |
| SL010492 | Calcium/calmodulin-dependent protein kinase type II subunit beta | CAMK2B | Q13554 | 816 | CAMK2B | -1.383221 | 0.226004 |
| SL000308 | Plasma protease C1 inhibitor | C1-Esterase Inhibitor | P05155 | 710 | SERPING1 | 1.147232 | 0.226567 |
| SL011629 | AH receptor-interacting protein | AIP | O00170 | 9049 | AIP | -1.320505 | 0.227735 |
| SL003687 | Nucleoside diphosphate kinase A | Nucleoside diphosphate kinase A | P15531 | 4830 | NME1 | -1.124575 | 0.227873 |
| SL007358 | Protein kinase C theta type | KPCT | Q04759 | 5588 | PRKCQ | -1.376255 | 0.228305 |
| SL007328 | Protein jagged-1 | JAG1 | P78504 | 182 | JAG1 | 1.108115 | 0.230068 |
| SL011100 | CD226 antigen | CD226 | Q15762 | 10666 | CD226 | -1.103302 | 0.230477 |
| SL015046 | Amphoterin-induced protein 2 | AMGO2 | Q86SJ2 | 347902 | AMIGO2 | 1.261579 | 0.230492 |
| SL001888 | Antileukoproteinase | SLPI | P03973 | 6590 | SLPI | 1.086254 | 0.231548 |
| SL003300 | C-C motif chemokine 16 | HCC-4 | O15467 | 6360 | CCL16 | 1.121547 | 0.231781 |
| SL001800 | Tumor necrosis factor receptor superfamily member 1B | TNF sR-II | P20333 | 7133 | TNFRSF1B | 1.092512 | 0.232344 |
| SL003717 | Caspase-10 | Caspase-10 | Q92851 | 843 | CASP10 | -1.057047 | 0.232742 |
| SL009629 | Methyl-CpG-binding domain protein 4 | MBD4 | O95243 | 8930 | MBD4 | -1.222999 | 0.233842 |
| SL007179 | Ephrin type-B receptor 2 | EPHB2 | P29323 | 2048 | EPHB2 | 1.129862 | 0.23563 |
| SL004366 | Tumor necrosis factor receptor superfamily member 12A | TWEAKR | Q9NP84 | 51330 | TNFRSF12A | -2.270884 | 0.237179 |
| SL003700 | Bcl2-associated agonist of cell death | BAD | Q92934 | 572 | BAD | -1.226613 | 0.237202 |
| SL014108 | Leucine-rich repeat transmembrane neuronal protein 3 | LRRT3 | Q86VH5 | 347731 | LRRTM3 | 1.146934 | 0.237252 |
| SL012881 | Glypican-6 | GPC6 | Q9Y625 | 10082 | GPC6 | 1.079248 | 0.237709 |
| SL004140 | Ephrin-A4 | Ephrin-A4 | P52798 | 1945 | EFNA4 | 1.099809 | 0.23801 |
| SL002803 | Ubiquitin carboxyl-terminal hydrolase isozyme L1 | PGP9.5 | P09936 | 7345 | UCHL1 | -1.135431 | 0.238549 |
| SL004260 | Resistin | resistin | Q9HD89 | 56729 | RETN | 1.106857 | 0.239194 |
| SL004804 | Cell adhesion molecule 3 | Nectin-like protein 1 | Q8N126 | 57863 | CADM3 | 1.217628 | 0.240657 |
| SL010458 | Endothelial cell-specific molecule 1 | Endocan | Q9NQ30 | 11082 | ESM1 | 1.29693 | 0.240745 |
| SL000655 | Keratin, type I cytoskeletal 18 | Keratin 18 | P05783 | 3875 | KRT18 | -1.120131 | 0.241393 |
| SL010530 | OCIA domain-containing protein 1 | OCAD1 | Q9NX40 | 54940 | OCIAD1 | -1.066655 | 0.241437 |
| SL004477 | Protein S100-A9 | calgranulin B | P06702 | 6280 | S100A9 | -1.801416 | 0.243447 |
| SL007306 | Protein FAM3B | FAM3B | P58499 | 54097 | FAM3B | 1.177476 | 0.244838 |
| SL011630 | Seizure 6-like protein 2 | SE6L2 | Q6UXD5 | 26470 | SEZ6L2 | -1.195358 | 0.245544 |
| SL004588 | Interleukin-1 Receptor accessory protein | IL-1 R AcP | Q9NPH3 | 3556 | IL1RAP | -1.169316 | 0.246331 |
| SL000557 | Protein kinase C zeta type | PKC-Z | Q05513 | 5590 | PRKCZ | -1.175094 | 0.247808 |
| SL003171 | Fibroblast growth factor 4 | FGF-4 | P08620 | 2249 | FGF4 | 1.13527 | 0.247963 |
| SL005171 | Insulin-like growth factor-binding protein 4 | IGFBP-4 | P22692 | 3487 | IGFBP4 | 1.123527 | 0.247993 |
| SL004861 | Interleukin-18 receptor accessory protein | IL-18 Rb | O95256 | 8807 | IL18RAP | -1.122717 | 0.248526 |
| SL008158 | Histone H3.1 | H31 | P68431 | 8350 | HIST1H3A | -1.134148 | 0.24876 |
| SL010508 | Serine/threonine-protein kinase PAK 3 | PAK3 | O75914 | 5063 | PAK3 | -1.803493 | 0.248996 |
| SL017188 | Glycogen synthase kinase-3 alpha/beta | GSK-3 alpha/beta | P49840 P49841 | 2931 2932 | GSK3A GSK3B | -1.503114 | 0.249141 |
| SL014092 | Cell adhesion molecule-related/down-regulated by oncogenes | CDON | Q4KMG0 | 50937 | CDON | -1.110579 | 0.250438 |
| SL009210 | Pro-opiomelanocortin | Corticotropin-lipotropin | P01189 | 5443 | POMC | 1.275049 | 0.252712 |
| SL002756 | Heterogeneous nuclear ribonucleoprotein K | hnRNP K | P61978 | 3190 | HNRNPK | 1.08181 | 0.252805 |
| SL018938 | Small ubiquitin-related modifier 3 | SUMO3 | P55854 | 6613 | SUMO3 | -1.255843 | 0.252995 |
| SL004724 | Histone acetyltransferase KAT6A | MOZ | Q92794 | 7994 | KAT6A | -1.11202 | 0.253026 |
| SL000064 | Kallikrein-7 | Kallikrein 7 | P49862 | 5650 | KLK7 | -1.253016 | 0.253029 |
| SL005166 | Galectin-7 | Galectin-7 | P47929 | 3963 | LGALS7 | -1.096236 | 0.25303 |
| SL004151 | Interleukin-15 receptor subunit alpha | IL-15 Ra | Q13261 | 3601 | IL15RA | -1.227666 | 0.253462 |
| SL005308 | Proteasome activator complex subunit 3 | PSME3 | P61289 | 10197 | PSME3 | 1.314442 | 0.253819 |
| SL004815 | 14-3-3 protein beta/alpha | 14-3-3 protein beta/alpha | P31946 | 7529 | YWHAB | -1.525642 | 0.253915 |
| SL000409 | Mitogen-activated protein kinase 3 | ERK-1 | P27361 | 5595 | MAPK3 | -1.50881 | 0.254612 |
| SL012769 | Protein FAM3D | FAM3D | Q96BQ1 | 131177 | FAM3D | 1.162206 | 0.25481 |
| SL005226 | Cytokine receptor-like factor 2 | TSLP R | Q9HC73 | 64109 | CRLF2 | -1.251447 | 0.255008 |
| SL005179 | Interleukin-36 beta | IL-1F8 | Q9NZH7 | 27177 | IL36B | 1.187139 | 0.255227 |
| SL003178 | Intercellular adhesion molecule 3 | sICAM-3 | P32942 | 3385 | ICAM3 | -1.127548 | 0.255392 |
| SL004331 | Ciliary neurotrophic factor | CNTF | P26441 | 1270 | CNTF | 1.232825 | 0.257388 |
| SL000449 | DnaJ homolog subfamily B member 1 | HSP 40 | P25685 | 3337 | DNAJB1 | -1.139351 | 0.257435 |
| SL008071 | 14-3-3 protein theta | 14-3-3 protein theta | P27348 | 10971 | YWHAQ | -1.084525 | 0.257487 |
| SL004689 | WNT1-inducible-signaling pathway protein 1 | WISP-1 | O95388 | 8840 | WISP1 | -1.113375 | 0.257515 |
| SL013570 | Dual specificity tyrosine-phosphorylation-regulated kinase 3 | DYRK3 | O43781 | 8444 | DYRK3 | -1.092317 | 0.257818 |
| SL013928 | Peptidyl-prolyl cis-trans isomerase E | PPIE | Q9UNP9 | 10450 | PPIE | -1.290311 | 0.258756 |
| SL004852 | Programmed cell death 1 ligand 1 | B7-H1 | Q9NZQ7 | 29126 | CD274 | 2.38166 | 0.259022 |
| SL000462 | Insulin-like growth factor-binding protein 1 | IGFBP-1 | P08833 | 3484 | IGFBP1 | 1.147082 | 0.259389 |
| SL010491 | Calcium/calmodulin-dependent protein kinase type II subunit alpha | CAMK2A | Q9UQM7 | 815 | CAMK2A | -1.322374 | 0.260419 |
| SL004765 | MAP kinase-activated protein kinase 3 | MAPKAPK3 | Q16644 | 7867 | MAPKAPK3 | -1.264968 | 0.261407 |
| SL000052 | Troponin T, cardiac muscle | Troponin T | P45379 | 7139 | TNNT2 | 1.095376 | 0.261884 |
| SL004327 | Tumor necrosis factor ligand superfamily member 13B | BAFF | Q9Y275 | 10673 | TNFSF13B | 1.104823 | 0.262164 |
| SL005187 | Interleukin-3 receptor subunit alpha | IL-3 Ra | P26951 | 3563 | IL3RA | -1.200096 | 0.263671 |
| SL010468 | RGM domain family member B | RGMB | Q6NW40 | 285704 | RGMB | 1.110368 | 0.263726 |
| SL017614 | RAC-alpha/beta/gamma serine/threonine-protein kinase | PKB a/b/g | P31749 P31751 Q9Y243 | 207 208 10000 | AKT1 AKT2 AKT3 | -1.306985 | 0.264082 |
| SL000134 | Hepatocyte growth factor receptor | Met | P08581 | 4233 | MET | 1.11895 | 0.264208 |
| SL006629 | NAD-dependent protein deacetylase sirtuin-2 | SIRT2 | Q8IXJ6 | 22933 | SIRT2 | -1.562608 | 0.265487 |
| SL011549 | AT-rich interactive domain-containing protein 3A | ARI3A | Q99856 | 1820 | ARID3A | -1.818994 | 0.266027 |
| SL011448 | Tumor necrosis factor receptor superfamily member 4 | TNR4 | P43489 | 7293 | TNFRSF4 | 1.070943 | 0.268046 |
| SL003657 | Calcineurin | Calcineurin | Q08209 P63098 | 5530 5534 | PPP3CA PPP3R1 | -1.323078 | 0.268246 |
| SL009412 | Dickkopf-related protein 3 | DKK3 | Q9UBP4 | 27122 | DKK3 | 1.131704 | 0.268657 |
| SL003307 | Cytokine receptor common subunit gamma | IL-2 sRg | P31785 | 3561 | IL2RG | -1.197027 | 0.268735 |
| SL000084 | Epidermal growth factor | EGF | P01133 | 1950 | EGF | -1.122782 | 0.271049 |
| SL011628 | Drebrin-like protein | DBNL | Q9UJU6 | 28988 | DBNL | -1.085499 | 0.271931 |
| SL004759 | Eukaryotic translation initiation factor 5 | eIF-5 | P55010 | 1983 | EIF5 | 1.275997 | 0.272096 |
| SL011533 | Small glutamine-rich tetratricopeptide repeat-containing protein alpha | SGTA | O43765 | 6449 | SGTA | -1.169714 | 0.272356 |
| SL004351 | Interleukin-25 | IL-17E | Q9H293 | 64806 | IL25 | 1.104667 | 0.272483 |
| SL004060 | Endothelin-converting enzyme 1 | Endothelin-converting enzyme 1 | P42892 | 1889 | ECE1 | 1.096511 | 0.273016 |
| SL019979 | Histone H2A type 3 | H2A3 | Q7L7L0 | NaN | HIST3H2A | -1.145767 | 0.273274 |
| SL000009 | Receptor tyrosine-protein kinase erbB-2 | ERBB2 | P04626 | 2064 | ERBB2 | -1.242802 | 0.274851 |
| SL006892 | Tyrosine-protein kinase ABL1 | ABL1 | P00519 | 25 | ABL1 | 1.067427 | 0.275943 |
| SL008372 | Leucine-rich repeat transmembrane protein FLRT3 | FLRT3 | Q9NZU0 | 23767 | FLRT3 | 1.135733 | 0.276744 |
| SL007502 | Carbohydrate sulfotransferase 15 | ST4S6 | Q7LFX5 | 51363 | CHST15 | 1.106086 | 0.276799 |
| SL004920 | Cofilin-1 | Cofilin-1 | P23528 | 1072 | CFL1 | -1.262993 | 0.276854 |
| SL004149 | Interleukin-13 receptor subunit alpha-1 | IL-13 Ra1 | P78552 | 3597 | IL13RA1 | 1.140735 | 0.277758 |
| SL007471 | Collectin-12 | COLEC12 | Q5KU26 | 81035 | COLEC12 | 1.104144 | 0.278107 |
| SL010393 | Kremen protein 2 | KREM2 | Q8NCW0 | 79412 | KREMEN2 | -1.073163 | 0.278569 |
| SL004348 | Interferon lambda-1 | IFN-lambda 1 | Q8IU54 | 282618 | IFNL1 | -1.179097 | 0.278927 |
| SL007674 | T-lymphocyte surface antigen Ly-9 | LY9 | Q9HBG7 | 4063 | LY9 | 1.137564 | 0.279439 |
| SL004347 | Interleukin-22 | IL-22 | Q9GZX6 | 50616 | IL22 | -1.180034 | 0.27998 |
| SL000053 | Tissue-type plasminogen activator | tPA | P00750 | 5327 | PLAT | 1.155512 | 0.279991 |
| SL003728 | Baculoviral IAP repeat-containing protein 3 | cIAP-2 | Q13489 | 330 | BIRC3 | -1.798629 | 0.280145 |
| SL003200 | Angiopoietin-1 receptor, soluble | sTie-2 | Q02763 | 7010 | TEK | 1.090039 | 0.280741 |
| SL003647 | Annexin A6 | annexin VI | P08133 | 309 | ANXA6 | -1.346898 | 0.281909 |
| SL000272 | Antithrombin-III | Antithrombin III | P01008 | 462 | SERPINC1 | -1.034382 | 0.283056 |
| SL010466 | Membrane frizzled-related protein | MFRP | Q9BY79 | 83552 | MFRP | -1.249997 | 0.283521 |
| SL008382 | Cystatin-D | CYTD | P28325 | 1473 | CST5 | 1.201268 | 0.283548 |
| SL008644 | ADP-ribosyl cyclase/cyclic ADP-ribose hydrolase 2 | BST1 | Q10588 | 683 | BST1 | 1.210716 | 0.283945 |
| SL000324 | Complement component C8 | C8 | P07357,P07358,P07360 | 731 732 733 | C8A C8B C8G | 1.073152 | 0.284034 |
| SL010512 | PIK3CA/PIK3R1 | PIK3CA/PIK3R1 | P42336 P27986 | 5290 5295 | PIK3CA PIK3R1 | -1.263707 | 0.284136 |
| SL006918 | Mitogen-activated protein kinase 1 | MK01 | P28482 | 5594 | MAPK1 | -1.303929 | 0.284223 |
| SL003192 | Properdin | Properdin | P27918 | 5199 | CFP | -1.088 | 0.287257 |
| SL017424 | Platelet receptor Gi24 | GI24 | Q9H7M9 | NaN | C10orf54 | -1.119359 | 0.287659 |
| SL000145 | Interleukin-1 receptor type 2 | IL-1 sRII | P27930 | 7850 | IL1R2 | 1.14731 | 0.288474 |
| SL006777 | Fetuin-B | FETUB | Q9UGM5 | 26998 | FETUB | 1.129084 | 0.28865 |
| SL005208 | Reticulon-4 receptor | Nogo Receptor | Q9BZR6 | 65078 | RTN4R | 1.111923 | 0.290298 |
| SL008157 | Ubiquitin-conjugating enzyme E2 L3 | UB2L3 | P68036 | 7332 | UBE2L3 | -1.151439 | 0.290426 |
| SL003744 | Galectin-3 | Galectin-3 | P17931 | 3958 | LGALS3 | 1.062214 | 0.29056 |
| SL000020 | Apolipoprotein B | Apo B | P04114 | 338 | APOB | 1.216453 | 0.290875 |
| SL008574 | Osteomodulin | OMD | Q99983 | 4958 | OMD | 1.17511 | 0.291152 |
| SL002565 | Myc proto-oncogene protein | c-Myc | P01106 | 4609 | MYC | 1.933116 | 0.29147 |
| SL005229 | NKG2D ligand 3 | ULBP-3 | Q9BZM4 | 79465 | ULBP3 | -1.084362 | 0.292397 |
| SL016549 | AMP Kinase (alpha2beta2gamma1) | AMPK a2b2g1 | P54646 O43741 P54619 | 5563 5565 5571 | PRKAA2 PRKAB2 PRKAG1 | -1.39331 | 0.293033 |
| SL004850 | Interleukin-17 receptor A | IL-17 sR | Q96F46 | 23765 | IL17RA | 1.143906 | 0.293595 |
| SL016128 | Thioredoxin domain-containing protein 12 | TXD12 | O95881 | 51060 | TXNDC12 | 1.109211 | 0.293769 |
| SL001761 | Troponin I, cardiac muscle | Troponin I | P19429 | 7137 | TNNI3 | -1.068222 | 0.293879 |
| SL003183 | C-X-C motif chemokine 10 | IP-10 | P02778 | 3627 | CXCL10 | 1.074108 | 0.294102 |
| SL004081 | Decorin | Bone proteoglycan II | P07585 | 1634 | DCN | 1.253768 | 0.295119 |
| SL000554 | Protein kinase C delta type | PKC-D | Q05655 | 5580 | PRKCD | -1.119248 | 0.297044 |
| SL005184 | Interleukin-23 | IL-23 | P29460, Q9NPF7 | 3593 51561 | IL12B IL23A | 1.108162 | 0.297438 |
| SL008931 | CD177 antigen | CD177 | Q8N6Q3 | 57126 | CD177 | 1.402666 | 0.297973 |
| SL002561 | Parathyroid hormone-related protein | PTHrP | P12272 | 5744 | PTHLH | 1.314219 | 0.298608 |
| SL014209 | Killer cell lectin-like receptor subfamily F member 1 | KLRF1 | Q9NZS2 | 51348 | KLRF1 | -1.177693 | 0.298747 |
| SL003326 | C-X-C motif chemokine 11 | I-TAC | O14625 | 6373 | CXCL11 | -1.362222 | 0.298946 |
| SL010451 | Cryptic protein | CFC1 | P0CG37 | 55997 | CFC1 | -1.102777 | 0.299622 |
| SL006698 | Ligand-dependent nuclear receptor corepressor-like protein | transcription factor MLR1, isoform CRA_b | Q8N3X6 | 254251 | LCORL | -1.132905 | 0.299822 |
| SL006919 | Ribosomal protein S6 kinase alpha-5 | RSK-like protein kinase | O75582 | 9252 | RPS6KA5 | -1.191538 | 0.300735 |
| SL004697 | Protein E7_HPV16 | HPV E7 Type 16 | P03129 | 1489079 | Human-virus | 1.063469 | 0.301841 |
| SL000347 | Corticosteroid-binding globulin | CBG | P08185 | 866 | SERPINA6 | -1.221342 | 0.303619 |
| SL000136 | Amphiregulin | AREG | P15514 | 374 | AREG | -1.120927 | 0.304495 |
| SL003672 | Extracellular superoxide dismutase [Cu-Zn] | SOD3 | P08294 | 6649 | SOD3 | -1.011911 | 0.304969 |
| SL006091 | Cytoplasmic protein NCK1 | NCK1 | P16333 | 4690 | NCK1 | -1.074915 | 0.305829 |
| SL005725 | GRB2-related adapter protein 2 | GRB2-related adapter protein 2 | O75791 | 9402 | GRAP2 | -1.295958 | 0.306951 |
| SL003524 | Protein disulfide-isomerase A3 | Protein disulfide isomerase A3 | P30101 | 2923 | PDIA3 | -1.117679 | 0.307579 |
| SL005172 | Insulin-like growth factor-binding protein 6 | IGFBP-6 | P24592 | 3489 | IGFBP6 | 1.124875 | 0.309239 |
| SL000539 | Glucose-6-phosphate isomerase | PHI | P06744 | 2821 | GPI | -1.481261 | 0.309745 |
| SL003176 | C-C motif chemokine 1 | I-309 | P22362 | 6346 | CCL1 | -1.044561 | 0.310496 |
| SL009054 | Neurexin-1-beta | NRX1B | P58400 | 9378 | NRXN1 | 1.137977 | 0.312318 |
| SL008773 | CD109 antigen | CD109 | Q6YHK3 | 135228 | CD109 | 1.124721 | 0.31276 |
| SL007373 | Peptidyl-prolyl cis-trans isomerase D | PPID | Q08752 | 5481 | PPID | -1.484568 | 0.312798 |
| SL007223 | B-cell lymphoma 6 protein | BCL6 | P41182 | 604 | BCL6 | 1.924367 | 0.313179 |
| SL000322 | Complement component C6 | C6 | P13671 | 729 | C6 | -1.058469 | 0.314364 |
| SL003186 | Lymphotactin | Lymphotactin | P47992 | 6375 | XCL1 | -3.387432 | 0.314778 |
| SL004400 | Coagulation factor IXab | Coagulation Factor IXab | P00740 | 2158 | F9 | 1.094303 | 0.315303 |
| SL009951 | Protein Wnt-7a | WNT7A | O00755 | 7476 | WNT7A | 1.1906 | 0.316412 |
| SL000089 | Transforming growth factor beta-3 | TGF-b3 | P10600 | 7043 | TGFB3 | 1.067861 | 0.316644 |
| SL004771 | Aurora kinase A | Aurora kinase A | O14965 | 6790 | AURKA | 1.244651 | 0.317714 |
| SL000248 | Alpha-1-antichymotrypsin | a1-Antichymotrypsin | P01011 | 12 | SERPINA3 | 1.040824 | 0.318452 |
| SL014129 | Heparan-sulfate 6-O-sulfotransferase 1 | H6ST1 | O60243 | 9394 | HS6ST1 | 1.120992 | 0.318576 |
| SL010494 | Cyclin-dependent kinase 1:G2/mitotic-specific cyclin-B1 complex | CDK1/cyclin B | P06493 P14635 | 983 891 | CDC2 CCNB1 | -1.06621 | 0.318609 |
| SL003674 | Bcl-2-like protein 1 | BCL2-like 1 protein | Q07817 | 598 | BCL2L1 | -1.081938 | 0.319249 |
| SL000448 | Heat shock protein beta-1 | HSP 27 | P04792 | 3315 | HSPB1 | -1.395083 | 0.319273 |
| SL011709 | Pyridoxal phosphate phosphatase | PLPP | Q96GD0 | 57026 | PDXP | -1.165597 | 0.319476 |
| SL003753 | Dynein light chain 1, cytoplasmic | DLC8 | P63167 | 8655 | DYNLL1 | 1.032149 | 0.31972 |
| SL007336 | Protein SET | SET | Q01105 | 6418 | SET | 1.085369 | 0.320974 |
| SL012740 | A disintegrin and metalloproteinase with thrombospondin motifs 15 | ATS15 | Q8TE58 | 170689 | ADAMTS15 | 1.047361 | 0.322414 |
| SL000271 | Angiotensinogen | Angiotensinogen | P01019 | 183 | AGT | 1.099825 | 0.322475 |
| SL007153 | Lactoperoxidase | PERL | P22079 | 4025 | LPO | -1.184221 | 0.322971 |
| SL004855 | Contactin-1 | contactin-1 | Q12860 | 1272 | CNTN1 | 1.084069 | 0.32411 |
| SL003173 | Growth-regulated alpha protein | Gro-a | P09341 | 2919 | CXCL1 | 1.033005 | 0.324951 |
| SL008063 | Low molecular weight phosphotyrosine protein phosphatase | PPAC | P24666 | 52 | ACP1 | 1.079087 | 0.325165 |
| SL005167 | Galectin-8 | Galectin-8 | O00214 | 3964 | LGALS8 | -2.755974 | 0.325427 |
| SL000450 | 60 kDa heat shock protein, mitochondrial | HSP 60 | P10809 | 3329 | HSPD1 | 1.1342 | 0.325554 |
| SL008378 | Eukaryotic translation initiation factor 4E-binding protein 2 | 4EBP2 | Q13542 | 1979 | EIF4EBP2 | -1.122471 | 0.3269 |
| SL005194 | Junctional adhesion molecule C | JAM-C | Q9BX67 | 83700 | JAM3 | -1.132632 | 0.327998 |
| SL004340 | Fibroblast growth factor 6 | FGF-6 | P10767 | 2251 | FGF6 | -1.102764 | 0.328701 |
| SL004812 | Triosephosphate isomerase | Triosephosphate isomerase | P60174 | 7167 | TPI1 | -1.233168 | 0.328901 |
| SL004742 | Afamin | Afamin | P43652 | 173 | AFM | -1.067075 | 0.329221 |
| SL008085 | 3-hydroxyisobutyrate dehydrogenase, mitochondrial | 3HIDH | P31937 | 11112 | HIBADH | -2.690931 | 0.332395 |
| SL011808 | Copine-1 | CPNE1 | Q99829 | 8904 | CPNE1 | -1.444641 | 0.332854 |
| SL004146 | Interleukin-1 receptor-like 1 | IL-1 R4 | Q01638 | 9173 | IL1RL1 | 1.222613 | 0.332889 |
| SL004864 | Cadherin-12 | Cadherin-12 | P55289 | 1010 | CDH12 | -1.089561 | 0.333175 |
| SL011770 | Leucine carboxyl methyltransferase 1 | LCMT1 | Q9UIC8 | 51451 | LCMT1 | -1.046474 | 0.333726 |
| SL000104 | Apoptosis regulator Bcl-2 | Bcl-2 | P10415 | 596 | BCL2 | -1.07107 | 0.334183 |
| SL000142 | Thymidylate synthase | TS | P04818 | 7298 | TYMS | -1.185575 | 0.336418 |
| SL004940 | Tyrosine-protein phosphatase non-receptor type 6 | PTP-1C | P29350 | 5777 | PTPN6 | -1.338218 | 0.336567 |
| SL005213 | Tumor necrosis factor receptor superfamily member 19L | RELT | Q969Z4 | 84957 | RELT | 1.082261 | 0.336794 |
| SL000087 | Interleukin-6 | IL-6 | P05231 | 3569 | IL6 | 1.465451 | 0.337042 |
| SL002541 | Tumor necrosis factor ligand superfamily member 11 | sRANKL | O14788 | 8600 | TNFSF11 | -1.136384 | 0.337714 |
| SL010529 | Ubiquitin-fold modifier-conjugating enzyme 1 | UFC1 | Q9Y3C8 | 51506 | UFC1 | -1.225394 | 0.339457 |
| SL008380 | Cathepsin Z | CATZ | Q9UBR2 | 1522 | CTSZ | 1.099053 | 0.340008 |
| SL007122 | Insulin-degrading enzyme | IDE | P14735 | 3416 | IDE | 1.199917 | 0.340066 |
| SL011406 | High affinity cAMP-specific 3',5'-cyclic phosphodiesterase 7A | PDE7A | Q13946 | 5150 | PDE7A | -1.073698 | 0.340725 |
| SL000445 | Protein Rev_HV2BE | HIV-2 Rev | P18093 | 1724716 | Human-virus | 1.037791 | 0.34129 |
| SL004676 | Insulin-like growth factor-binding protein 5 | IGFBP-5 | P24593 | 3488 | IGFBP5 | 1.104614 | 0.343194 |
| SL008416 | C-type mannose receptor 2 | MRC2 | Q9UBG0 | 9902 | MRC2 | 1.133093 | 0.345398 |
| SL000441 | Hepatocyte growth factor | HGF | P14210 | 3082 | HGF | 1.442545 | 0.346026 |
| SL011232 | Hsp90 co-chaperone Cdc37 | CDC37 | Q16543 | 11140 | CDC37 | -1.396479 | 0.346126 |
| SL010374 | Methionine aminopeptidase 1 | METAP1 | P53582 | 23173 | METAP1 | -1.312384 | 0.346671 |
| SL007217 | Interferon gamma receptor 2 | INGR2 | P38484 | 3460 | IFNGR2 | -1.097549 | 0.347408 |
| SL004131 | T-lymphocyte activation antigen CD86 | B7-2 | P42081 | 942 | CD86 | -1.119179 | 0.347681 |
| SL000519 | C-C motif chemokine 3 | MIP-1a | P10147 | 6348 | CCL3 | 1.13815 | 0.349287 |
| SL003919 | Kallikrein-14 | kallikrein 14 | Q9P0G3 | 43847 | KLK14 | -1.047708 | 0.349531 |
| SL006029 | Chitotriosidase-1 | Chitotriosidase-1 | Q13231 | 1118 | CHIT1 | 1.188969 | 0.349585 |
| SL003803 | Receptor tyrosine-protein kinase erbB-4 | ERBB4 | Q15303 | 2066 | ERBB4 | 1.05176 | 0.349641 |
| SL002508 | Interleukin-18-binding protein | IL-18 BPa | O95998 | 10068 | IL18BP | 1.101174 | 0.349662 |
| SL012148 | Peroxisomal targeting signal 1 receptor | PEX5 | P50542 | 5830 | PEX5 | -1.157999 | 0.349961 |
| SL006911 | Serine/threonine-protein kinase Chk1 | CHK1 | O14757 | 1111 | CHEK1 | -1.035656 | 0.350125 |
| SL004899 | Heat shock cognate 71 kDa protein | HSP70 protein 8 | P11142 | 3312 | HSPA8 | -1.112681 | 0.351197 |
| SL007953 | Pyridoxal kinase | PDXK | O00764 | 8566 | PDXK | -1.242165 | 0.352038 |
| SL009216 | Aromatic-L-amino-acid decarboxylase | dopa decarboxylase | P20711 | 1644 | DDC | -1.090587 | 0.352052 |
| SL000532 | SPARC | ON | P09486 | 6678 | SPARC | -1.129003 | 0.352752 |
| SL003520 | Calreticulin | calreticulin | P27797 | 811 | CALR | 1.442383 | 0.352908 |
| SL010501 | Dual specificity mitogen-activated protein kinase kinase 2 | MP2K2 | P36507 | 5605 | MAP2K2 | 1.085369 | 0.353115 |
| SL018509 | R-spondin-3 | RSPO3 | Q9BXY4 | 84870 | RSPO3 | 1.284896 | 0.353599 |
| SL011073 | Xaa-Pro aminopeptidase 1 | XPNPEP1 | Q9NQW7 | 7511 | XPNPEP1 | -1.244876 | 0.35582 |
| SL008178 | Dermatopontin | DERM | Q07507 | 1805 | DPT | 1.093643 | 0.356002 |
| SL003043 | Metalloproteinase inhibitor 3 | TIMP-3 | P35625 | 7078 | TIMP3 | -1.131784 | 0.357301 |
| SL002602 | Trefoil factor 2 | Trefoil factor 2 | Q03403 | 7032 | TFF2 | 1.182226 | 0.35789 |
| HCE004359 | HCE004359 | HCE004359 | HCE004359 | HCE004359 | HCE004359 | 1.067264 | 0.358744 |
| SL005358 | Phosphatidylethanolamine-binding protein 1 | prostatic binding protein | P30086 | 5037 | PEBP1 | -1.103514 | 0.358776 |
| SL006406 | Plexin-C1 | PLXC1 | O60486 | 10154 | PLXNC1 | 1.099412 | 0.359085 |
| SL007003 | Desert hedgehog protein N-product | DHH | O43323 | 50846 | DHH | -1.13178 | 0.360472 |
| SL014248 | Semaphorin-6B | SEM6B | Q9H3T3 | 10501 | SEMA6B | 1.10133 | 0.361721 |
| SL002722 | ADP-ribosyl cyclase/cyclic ADP-ribose hydrolase 1 | CD38 | P28907 | 952 | CD38 | 2.526524 | 0.361916 |
| SL011510 | Somatostatin-28 | Somatostatin-28 | P61278 | 6750 | SST | 1.05297 | 0.362929 |
| SL010463 | Glypican-2 | GPC2 | Q8N158 | 221914 | GPC2 | -1.103052 | 0.363519 |
| SL003304 | Insulin-like growth factor 1 receptor | IGF-I sR | P08069 | 3480 | IGF1R | 1.095574 | 0.363693 |
| SL009328 | Chorionic somatomammotropin hormone | CSH | P0DML2 P0DML3 | 1442 1443 | CSH1 CSH2 | -1.206123 | 0.363954 |
| SL004334 | Fibroblast growth factor 16 | FGF-16 | O43320 | 8823 | FGF16 | -1.093153 | 0.365664 |
| SL010518 | Tyrosine-protein kinase Tec | TEC | P42680 | 7006 | TEC | -1.251841 | 0.365714 |
| SL004938 | Calcium/calmodulin-dependent protein kinase kinase 1 | CaMKK alpha | Q8N5S9 | 84254 | CAMKK1 | -1.074347 | 0.367696 |
| SL007560 | Stabilin-2 | STAB2 | Q8WWQ8 | 55576 | STAB2 | -1.041499 | 0.369336 |
| SL007453 | Mitogen-activated protein kinase 11 | MK11 | Q15759 | 5600 | MAPK11 | 1.075874 | 0.370096 |
| SL007261 | Methionine aminopeptidase 2 | AMPM2 | P50579 | 10988 | METAP2 | -1.283939 | 0.370766 |
| SL002075 | Interferon alpha-2 | IFN-aA | P01563 | 3440 | IFNA2 | -1.119919 | 0.372475 |
| SL003722 | RAC-beta serine/threonine-protein kinase | PKB beta | P31751 | 208 | AKT2 | -1.217253 | 0.372989 |
| SL004645 | Kunitz-type protease inhibitor 1 | HAI-1 | O43278 | 6692 | SPINT1 | -1.073383 | 0.375032 |
| SL004101 | Mothers against decapentaplegic homolog 2 | SMAD2 | Q15796 | 4087 | SMAD2 | -2.022022 | 0.375441 |
| SL004757 | Vacuolar protein sorting-associated protein VTA1 homolog | DRG-1 | Q9NP79 | 51534 | VTA1 | -1.293659 | 0.375553 |
| SL000048 | Vitamin K-dependent protein C | Protein C | P04070 | 5624 | PROC | -1.105392 | 0.377353 |
| SL006913 | Tyrosine-protein kinase Fyn | FYN | P06241 | 2534 | FYN | -1.288825 | 0.378268 |
| SL008143 | Ubiquitin-conjugating enzyme E2 N | UBE2N | P61088 | 7334 | UBE2N | -1.115472 | 0.378717 |
| SL011400 | Calcium/calmodulin-dependent 3',5'-cyclic nucleotide phosphodiesterase 1A | PDE1A | P54750 | 5136 | PDE1A | -1.158417 | 0.378724 |
| SL008122 | Dual specificity protein phosphatase 3 | DUS3 | P51452 | 1845 | DUSP3 | -1.19168 | 0.380157 |
| SL004355 | C-C motif chemokine 3-like 1 | LD78-beta | P16619 | 414062 | CCL3L1 | -1.048931 | 0.380254 |
| SL004142 | Ephrin-B3 | Ephrin-B3 | Q15768 | 1949 | EFNB3 | 1.094565 | 0.380339 |
| SL006998 | 3-phosphoinositide-dependent protein kinase 1 | PDPK1 | O15530 | 5170 | PDPK1 | -1.333468 | 0.3807 |
| SL006705 | Prefoldin subunit 5 | PFD5 | Q99471 | 5204 | PFDN5 | -1.154824 | 0.381427 |
| SL014148 | Roundabout homolog 3 | ROBO3 | Q96MS0 | 64221 | ROBO3 | -1.14329 | 0.381847 |
| SL010490 | Calcium/calmodulin-dependent protein kinase type 1D | CAMK1D | Q8IU85 | 57118 | CAMK1D | -1.0473 | 0.382813 |
| SL002081 | Cadherin-5 | Cadherin-5 | P33151 | 1003 | CDH5 | 1.137072 | 0.383181 |
| SL000398 | Cytochrome P450 3A4 | Cytochrome P450 3A4 | P08684 | 1576 | CYP3A4 | -1.12646 | 0.38381 |
| SL004080 | Bone morphogenetic protein receptor type-1A | BMPR1A | P36894 | 657 | BMPR1A | 1.095188 | 0.383875 |
| SL004915 | Chloride intracellular channel protein 1 | NCC27 | O00299 | 1192 | CLIC1 | -1.238224 | 0.384648 |
| SL000343 | Cathepsin B | Cathepsin B | P07858 | 1508 | CTSB | 1.062994 | 0.385173 |
| SL011529 | Ribosome maturation protein SBDS | SBDS | Q9Y3A5 | 51119 | SBDS | -1.232745 | 0.385936 |
| SL006550 | Extracellular matrix protein 1 | ECM1 | Q16610 | 1893 | ECM1 | 1.114712 | 0.387042 |
| SL008102 | Malate dehydrogenase, cytoplasmic | MDHC | P40925 | 4190 | MDH1 | -1.044539 | 0.387165 |
| SL004119 | Epithelial discoidin domain-containing receptor 1 | discoidin domain receptor 1 | Q08345 | 780 | DDR1 | -1.106205 | 0.388145 |
| SL000455 | Transcription factor AP-1 | c-Jun | P05412 | 3725 | JUN | -1.043229 | 0.388523 |
| SL014048 | Collagen alpha-1(XXIII) chain | CONA1 | Q86Y22 | 91522 | COL23A1 | -1.114349 | 0.388523 |
| SL002650 | Pyruvate kinase PKM | M2-PK | P14618 | 5315 | PKM2 | -1.451381 | 0.39058 |
| SL008808 | S-phase kinase-associated protein 1 | SKP1 | P63208 | 6500 | SKP1 | -1.043357 | 0.39103 |
| SL000556 | Protein kinase C gamma type | PKC-G | P05129 | 5582 | PRKCG | 1.321241 | 0.391247 |
| SL004747 | Apolipoprotein M | ApoM | O95445 | 55937 | APOM | 1.133024 | 0.391909 |
| SL003440 | Platelet-activating factor acetylhydrolase | PAFAH | Q13093 | 7941 | PLA2G7 | 1.062093 | 0.392165 |
| SL001943 | Interleukin-6 receptor subunit alpha | IL-6 sRa | P08887 | 3570 | IL6R | 1.076483 | 0.39268 |
| SL012188 | Epidermal growth factor receptor substrate 15-like 1 | EP15R | Q9UBC2 | 58513 | EPS15L1 | -1.063514 | 0.393061 |
| SL006460 | Platelet glycoprotein Ib alpha chain | GP1BA | P07359 | 2811 | GP1BA | 1.044368 | 0.394921 |
| SL013989 | R-spondin-2 | RSPO2 | Q6UXX9 | 340419 | RSPO2 | -1.091891 | 0.395101 |
| SL008466 | 3-hydroxyanthranilate 3,4-dioxygenase | 3HAO | P46952 | 23498 | HAAO | 1.074834 | 0.395642 |
| SL017290 | PolyUbiquitin K63-linked | PolyUbiquitin K63 | P0CG48 | 7316 | UBC | 1.056176 | 0.39594 |
| SL004330 | B-cell receptor CD22 | CD22 | P20273 | 933 | CD22 | -1.036373 | 0.396459 |
| SL004589 | Azurocidin | Azurocidin | P20160 | 566 | AZU1 | -1.083044 | 0.398252 |
| SL012517 | R-spondin-4 | RSPO4 | Q2I0M5 | 343637 | RSPO4 | 1.073515 | 0.398468 |
| SL003655 | Transketolase | Transketolase | P29401 | 7086 | TKT | -1.21912 | 0.398739 |
| SL000427 | Fractalkine | Fractalkine/CX3CL-1 | P78423 | 6376 | CX3CL1 | 1.061537 | 0.398769 |
| SL001791 | Osteocalcin | Osteocalcin | P02818 | 632 | BGLAP | -1.088794 | 0.399296 |
| SL007056 | Bone morphogenetic protein 10 | BMP10 | O95393 | 27302 | BMP10 | 1.101387 | 0.400009 |
| SL007310 | Ras-related C3 botulinum toxin substrate 3 | RAC3 | P60763 | 5881 | RAC3 | -1.098165 | 0.400345 |
| SL016148 | gp41 C34 peptide, HIV | C34 gp41 HIV Fragment | Q70626 | NaN | Human-virus | 1.199107 | 0.400765 |
| SL004580 | Macrophage scavenger receptor types I and II | Macrophage scavenger receptor | P21757 | 4481 | MSR1 | -1.09332 | 0.401391 |
| SL005793 | Peptidyl-prolyl cis-trans isomerase F, mitochondrial | Cyclophilin F | P30405 | 10105 | PPIF | -1.358885 | 0.401638 |
| SL004924 | Eukaryotic translation initiation factor 4H | eIF-4H | Q15056 | 7458 | EIF4H | -1.356931 | 0.404083 |
| SL000454 | Heat shock protein HSP 90-beta | HSP 90b | P08238 | 3326 | HSP90AB1 | -1.209153 | 0.405174 |
| SL000506 | Luteinizing hormone | Luteinizing hormone | P01215, P01229 | 1081 3972 | CGA LHB | 1.090712 | 0.405313 |
| SL000357 | Coagulation factor IX | Coagulation Factor IX | P00740 | 2158 | F9 | 1.080956 | 0.405703 |
| SL010528 | Ubiquitin-fold modifier 1 | UFM1 | P61960 | 51569 | UFM1 | -1.198577 | 0.405907 |
| SL014469 | SHC-transforming protein 1 | SHC1 | P29353 | 6464 | SHC1 | -1.173098 | 0.405907 |
| SL018548 | Alpha-1-antichymotrypsin complex | alpha-1-antichymotrypsin complex | P01011 | NaN | SERPINA3 | 1.090576 | 0.406555 |
| SL004821 | Protein S100-A4 | S100A4 | P26447 | 6275 | S100A4 | 1.05487 | 0.407656 |
| SL004063 | Fibroblast growth factor receptor 3 | FGFR-3 | P22607 | 2261 | FGFR3 | -1.033544 | 0.407927 |
| SL006920 | Mitogen-activated protein kinase 14 | MAPK14 | Q16539 | 1432 | MAPK14 | -1.179192 | 0.409423 |
| SL000497 | Laminin | Laminin | P25391, P07942, P11047 | 284217 3912 3915 | LAMA1 LAMB1 LAMC1 | -1.080192 | 0.41059 |
| SL005185 | Interleukin-23 receptor | IL-23 R | Q5VWK5 | 149233 | IL23R | -1.075666 | 0.410681 |
| SL000674 | Follistatin | FST | P19883 | 10468 | FST | -1.084613 | 0.412723 |
| SL004733 | Low-density lipoprotein receptor-related protein 1, soluble | sLRP1 | Q07954 | 4035 | LRP1 | 1.087892 | 0.412891 |
| SL000524 | Stromelysin-1 | MMP-3 | P08254 | 4314 | MMP3 | 1.354063 | 0.413032 |
| SL000384 | Cytotoxic T-lymphocyte protein 4 | CTLA-4 | P16410 | 1493 | CTLA4 | -3.309048 | 0.41346 |
| SL006448 | Histidine-rich glycoprotein | HRG | P04196 | 3273 | HRG | 1.114316 | 0.413674 |
| SL000313 | C3a anaphylatoxin | C3a | P01024 | 718 | C3 | 1.112321 | 0.414113 |
| SL005357 | Lithostathine-1-alpha | PSP | P05451 | 5967 | REG1A | 1.08342 | 0.4155 |
| SL008023 | Hyaluronan and proteoglycan link protein 1 | HPLN1 | P10915 | 1404 | HAPLN1 | -1.080281 | 0.416945 |
| SL003323 | C-C motif chemokine 18 | PARC | P55774 | 6362 | CCL18 | 1.114342 | 0.418532 |
| SL005202 | Hepatocyte growth factor-like protein | MSP | P26927 | 4485 | MST1 | 1.031871 | 0.419109 |
| SL005372 | Sorting nexin-4 | Sorting nexin 4 | O95219 | 8723 | SNX4 | -1.269982 | 0.419676 |
| SL010517 | Serine/threonine-protein kinase 16 | STK16 | O75716 | 8576 | STK16 | -1.05856 | 0.420212 |
| SL000638 | Cadherin-2 | Cadherin-2 | P19022 | 1000 | CDH2 | 1.70568 | 0.422086 |
| SL005034 | GTP-binding nuclear protein Ran | RAN | P62826 | 5901 | RAN | 1.006189 | 0.422087 |
| SL004230 | Microtubule-associated protein tau | tau | P10636 | 4137 | MAPT | 1.404848 | 0.423197 |
| HCE004333 | HCE004333 | HCE004333 | HCE004333 | HCE004333 | HCE004333 | 1.060451 | 0.423373 |
| SL007531 | BMP-binding endothelial regulator protein | BMPER | Q8N8U9 | 168667 | BMPER | 1.057748 | 0.423519 |
| SL000316 | Complement C4 | C4 | P0C0L4, P0C0L5 | 720 721 | C4A C4B | -1.039785 | 0.424231 |
| SL001997 | Interleukin-1 receptor type 1 | IL-1 sRI | P14778 | 3554 | IL1R1 | 1.110748 | 0.424335 |
| SL010459 | Low affinity immunoglobulin gamma Fc region receptor II-a | FCG2A | P12318 | 2212 | FCGR2A | 1.407018 | 0.42521 |
| SL010503 | MAP kinase-activated protein kinase 2 | MAPK2 | P49137 | 9261 | MAPKAPK2 | -1.317071 | 0.42574 |
| SL004643 | Angiopoietin-4 | Angiopoietin-4 | Q9Y264 | 51378 | ANGPT4 | -1.117056 | 0.427588 |
| SL000507 | Lymphotoxin alpha1:beta2 | Lymphotoxin a1/b2 | P01374, Q06643 | 4049 4050 | LTA LTB | 1.088688 | 0.428477 |
| SL000383 | Creatine kinase M-type | CK-MM | P06732 | 1158 | CKM | -1.173507 | 0.428937 |
| SL000305 | beta-nerve growth factor | b-NGF | P01138 | 4803 | NGF | 1.363678 | 0.429556 |
| HCE000483 | HCE000483 | HCE000483 | HCE000483 | HCE000483 | HCE000483 | 1.06749 | 0.430166 |
| HCE003300 | HCE003300 | HCE003300 | HCE003300 | HCE003300 | HCE003300 | 1.050384 | 0.430518 |
| SL010371 | Ectonucleoside triphosphate diphosphohydrolase 1 | CD39 | P49961 | 953 | ENTPD1 | -1.044159 | 0.431461 |
| SL006917 | Tyrosine-protein kinase Lyn | LYN | P07948 | 4067 | LYN | -1.335003 | 0.431632 |
| SL004858 | GDNF family receptor alpha-1 | GFRa-1 | P56159 | 2674 | GFRA1 | 1.215296 | 0.431742 |
| SL008177 | Complement component 1 Q subcomponent-binding protein, mitochondrial | C1QBP | Q07021 | 708 | C1QBP | 1.043014 | 0.431865 |
| SL007620 | Interleukin-12 receptor subunit beta-2 | IL-12 RB2 | Q99665 | 3595 | IL12RB2 | 1.04181 | 0.432193 |
| SL003104 | C-C motif chemokine 24 | Eotaxin-2 | O00175 | 6369 | CCL24 | 1.042256 | 0.433118 |
| SL003785 | Glyceraldehyde-3-phosphate dehydrogenase | GAPDH, liver | P04406 | 2597 | GAPDH | -1.199651 | 0.43376 |
| SL004851 | Serine/threonine-protein kinase receptor R3 | ALK-1 | P37023 | 94 | ACVRL1 | -1.439165 | 0.434932 |
| SL005679 | Translationally-controlled tumor protein | TCTP | P13693 | 7178 | TPT1 | -1.19091 | 0.435305 |
| SL006542 | Ficolin-2 | FCN2 | Q15485 | 2220 | FCN2 | -1.02627 | 0.436566 |
| SL005572 | Gelsolin | Gelsolin | P06396 | 2934 | GSN | 1.07129 | 0.436888 |
| SL002731 | Tumor necrosis factor receptor superfamily member 6 | Fas, soluble | P25445 | 355 | FAS | 1.073671 | 0.436893 |
| SL000587 | Thyroglobulin | Thyroglobulin | P01266 | 7038 | TG | 1.034492 | 0.436951 |
| SL002525 | Complement C2 | C2 | P06681 | 717 | C2 | 1.058881 | 0.438229 |
| SL004687 | Thymic stromal lymphopoietin | TSLP | Q969D9 | 85480 | TSLP | -1.04301 | 0.438365 |
| SL005059 | Transforming growth factor beta receptor type 3 | TGF-b R III | Q03167 | 7049 | TGFBR3 | 1.108168 | 0.439676 |
| SL004486 | Vascular endothelial growth factor C | VEGF-C | P49767 | 7424 | VEGFC | -1.053903 | 0.440385 |
| SL006970 | Delta-like protein 1 | DLL1 | O00548 | 28514 | DLL1 | 1.071412 | 0.441398 |
| SL010388 | Trypsin-2 | Trypsin 2 | P07478 | 5645 | PRSS2 | -1.007056 | 0.441732 |
| HCE003167 | HCE003167 | HCE003167 | HCE003167 | HCE003167 | HCE003167 | 1.06057 | 0.441861 |
| SL003792 | Growth factor receptor-bound protein 2 | GRB2 adapter protein | P62993 | 2885 | GRB2 | -1.342886 | 0.44323 |
| SL006378 | S-formylglutathione hydrolase | Esterase D | P10768 | 2098 | ESD | 1.099805 | 0.44495 |
| SL000526 | Neutrophil collagenase | MMP-8 | P22894 | 4317 | MMP8 | -1.345956 | 0.446347 |
| SL004070 | Ubiquitin | Ubiquitin | P62979 | 6233 | RPS27A | -1.130224 | 0.448112 |
| SL016551 | Casein kinase II 2-alpha':2-beta heterotetramer | CK2-A2:B | P19784 P67870 | 1459 1460 | CSNK2A2 CSNK2B | -1.140613 | 0.448135 |
| SL007327 | Oxidized low-density lipoprotein receptor 1 | OLR1 | P78380 | 4973 | OLR1 | -1.339827 | 0.448423 |
| SL011508 | Pituitary adenylate cyclase-activating polypeptide 27 | PACAP-27 | P18509 | 116 | ADCYAP1 | 2.197198 | 0.449125 |
| SL002086 | Ficolin-3 | Ficolin-3 | O75636 | 8547 | FCN3 | -1.415156 | 0.449756 |
| SL005181 | Interleukin-20 receptor subunit alpha | IL-20 Ra | Q9UHF4 | 53832 | IL20RA | 1.958384 | 0.450121 |
| SL006108 | CD5 antigen-like | CD5L | O43866 | 922 | CD5L | 1.043723 | 0.452069 |
| SL007806 | Interleukin-22 receptor subunit alpha-1 | IL22RA1 | Q8N6P7 | 58985 | IL22RA1 | -1.103173 | 0.452345 |
| SL007673 | Netrin-4 | NET4 | Q9HB63 | 59277 | NTN4 | 1.057646 | 0.453155 |
| SL007242 | Dual specificity mitogen-activated protein kinase kinase 3 | MP2K3 | P46734 | 5606 | MAP2K3 | -1.118145 | 0.453164 |
| SL004783 | Protein S100-A12 | S100A12 | P80511 | 6283 | S100A12 | -1.407355 | 0.453572 |
| SL004365 | Tumor necrosis factor ligand superfamily member 12 | TWEAK | O43508 | 8742 | TNFSF12 | 1.203658 | 0.453928 |
| SL004869 | Carbonic anhydrase 13 | Carbonic anhydrase XIII | Q8N1Q1 | 377677 | CA13 | -1.299611 | 0.455341 |
| SL011631 | Nascent polypeptide-associated complex subunit alpha | NACA | Q13765 | 4666 | NACA | -1.020275 | 0.456934 |
| SL005197 | Lysosome membrane protein 2 | LIMP II | Q14108 | 950 | SCARB2 | -1.1613 | 0.457094 |
| SL007804 | Biglycan | BGN | P21810 | 633 | BGN | 1.300157 | 0.4579 |
| SL010250 | Stress-induced-phosphoprotein 1 | Stress-induced-phosphoprotein 1 | P31948 | 10963 | STIP1 | -1.05536 | 0.459146 |
| SL006675 | Cytoskeleton-associated protein 2 | CKAP2 | Q8WWK9 | 26586 | CKAP2 | 1.033034 | 0.460467 |
| SL007228 | Protein kinase C iota type | KPCI | P41743 | 5584 | PRKCI | -1.079408 | 0.460606 |
| SL014008 | Alpha-(1,3)-fucosyltransferase 5 | FUT5 | Q11128 | 2527 | FUT5 | 1.0888 | 0.461915 |
| SL014028 | Ectonucleoside triphosphate diphosphohydrolase 5 | ENTP5 | O75356 | 957 | ENTPD5 | -1.060466 | 0.463546 |
| SL004718 | Importin subunit alpha-1 | Karyopherin-a2 | P52292 | 3838 | KPNA2 | 1.061245 | 0.464868 |
| SL000337 | Calpain I | Calpain I | P07384 P04632 | 823 826 | CAPN1 CAPNS1 | -1.176219 | 0.465227 |
| SL000678 | Granulysin | Granulysin | P22749 | 10578 | GNLY | -1.064262 | 0.467092 |
| SL000382 | Creatine kinase M-type:Creatine kinase B-type heterodimer | CK-MB | P12277 P06732 | 1152 1158 | CKB CKM | -1.255138 | 0.467412 |
| SL010456 | Cystatin-SN | CYTN | P01037 | 1469 | CST1 | 1.097265 | 0.467818 |
| SL004849 | X-linked interleukin-1 receptor accessory protein-like 2 | IL-1 sR9 | Q9NP60 | 26280 | IL1RAPL2 | -1.112526 | 0.468235 |
| SL016928 | SLAM family member 7 | SLAF7 | Q9NQ25 | 57823 | SLAMF7 | 1.074426 | 0.468398 |
| SL018629 | E3 ubiquitin-protein ligase RNF43 | RNF43 | Q68DV7 | 54894 | RNF43 | 1.039371 | 0.468834 |
| SL004299 | Baculoviral IAP repeat-containing protein 7 Isoform beta | Livin B | Q96CA5 | 79444 | BIRC7 | -1.142584 | 0.470471 |
| SL004639 | NT-3 growth factor receptor | TrkC | Q16288 | 4916 | NTRK3 | 1.078151 | 0.470543 |
| SL007059 | Lymphocyte antigen 86 | LY86 | O95711 | 9450 | LY86 | -1.034823 | 0.472339 |
| SL006372 | Tyrosine-protein kinase Yes | YES | P07947 | 7525 | YES1 | -1.165 | 0.473967 |
| SL008945 | Protein-glutamine gamma-glutamyltransferase E | TGM3 | Q08188 | 7053 | TGM3 | -1.803086 | 0.474782 |
| SL005212 | Prolactin receptor | Prolactin Receptor | P16471 | 5618 | PRLR | -1.022415 | 0.474833 |
| SL010376 | Membrane metallo-endopeptidase-like 1 | MMEL2 | Q495T6 | 79258 | MMEL1 | 1.558766 | 0.474947 |
| SL003739 | Tumor necrosis factor receptor superfamily member 6B | DcR3 | O95407 | 8771 | TNFRSF6B | 1.357204 | 0.475252 |
| HCE003183 | HCE003183 | HCE003183 | HCE003183 | HCE003183 | HCE003183 | 1.050015 | 0.475644 |
| SL008703 | Carboxypeptidase E | CBPE | P16870 | 1363 | CPE | -1.169343 | 0.476384 |
| SL004686 | Tumor necrosis factor ligand superfamily member 15 | TNFSF15 | O95150 | 9966 | TNFSF15 | -1.003812 | 0.476609 |
| SL008623 | Contactin-2 | CNTN2 | Q02246 | 6900 | CNTN2 | 1.046996 | 0.477773 |
| SL001737 | 14-3-3 protein sigma | STRATIFIN | P31947 | 2810 | SFN | 1.04262 | 0.478316 |
| SL011498 | Pituitary adenylate cyclase-activating polypeptide 38 | PACAP-38 | P18509 | 116 | ADCYAP1 | -1.114969 | 0.47837 |
| SL005688 | 14-3-3 protein zeta/delta | 14-3-3 protein zeta/delta | P63104 | 7534 | YWHAZ | -1.266318 | 0.479073 |
| SL017612 | Heat shock protein HSP 90-alpha/beta | HSP 90a/b | P07900 P08238 | NaN | HSP90AA1 HSP90AB1 | -1.159465 | 0.481378 |
| SL003793 | Dual specificity mitogen-activated protein kinase kinase 1 | MEK1 | Q02750 | 5604 | MAP2K1 | -1.09252 | 0.481425 |
| SL004860 | Serine protease HTRA2, mitochondrial | HTRA2 | O43464 | 27429 | HTRA2 | 1.028317 | 0.481593 |
| SL000001 | Mucin-1 | MUC1 | P15941 | 4582 | MUC1 | 1.065175 | 0.481724 |
| SL008331 | Proliferation-associated protein 2G4 | PA2G4 | Q9UQ80 | 5036 | PA2G4 | -1.118247 | 0.482365 |
| SL004823 | Peptidyl-prolyl cis-trans isomerase A | Cyclophilin A | P62937 | 5478 | PPIA | -1.159321 | 0.482459 |
| SL005177 | Interleukin-36 alpha | IL-1F6 | Q9UHA7 | 27179 | IL36A | -1.065897 | 0.482917 |
| SL007631 | Sclerostin | SOST | Q9BQB4 | 50964 | SOST | 1.10911 | 0.484553 |
| SL002755 | Pappalysin-1 | PAPP-A | Q13219 | 5069 | PAPPA | 1.073376 | 0.485426 |
| SL005228 | NKG2D ligand 2 | ULBP-2 | Q9BZM5 | 80328 | ULBP2 | -1.093358 | 0.486122 |
| SL005214 | Semaphorin-6A | Semaphorin-6A | Q9H2E6 | 57556 | SEMA6A | 1.091478 | 0.486293 |
| SL010616 | Tyrosine-protein phosphatase non-receptor type 11 | SHP-2 | Q06124 | 5781 | PTPN11 | -1.103896 | 0.48911 |
| SL008486 | Galectin-9 | LEG9 | O00182 | 3965 | LGALS9 | 1.042834 | 0.489867 |
| SL000670 | Glutathione S-transferase A3 | GSTA3 | Q16772 | 2940 | GSTA3 | -1.133638 | 0.490006 |
| SL000045 | Insulin-like growth factor-binding protein 3 | IGFBP-3 | P17936 | 3486 | IGFBP3 | -1.138238 | 0.49007 |
| SL012561 | Regenerating islet-derived protein 4 | REG4 | Q9BYZ8 | 83998 | REG4 | 1.068192 | 0.490286 |
| SL008381 | Cathepsin F | CATF | Q9UBX1 | 8722 | CTSF | 1.064309 | 0.491149 |
| SL000299 | Fibroblast growth factor 1 | b-ECGF | P05230 | 2246 | FGF1 | 1.048669 | 0.493009 |
| SL008835 | Asialoglycoprotein receptor 1 | ASGR1 | P07306 | 432 | ASGR1 | 1.08179 | 0.494333 |
| SL014229 | CD83 antigen | CD83 | Q01151 | 9308 | CD83 | 1.063903 | 0.495467 |
| SL004650 | Secreted frizzled-related protein 3 | sFRP-3 | Q92765 | 2487 | FRZB | 1.221556 | 0.495611 |
| SL000426 | Fibronectin | Fibronectin | P02751 | 2335 | FN1 | 1.071631 | 0.496051 |
| SL009792 | Adenylosuccinate lyase | PUR8 | P30566 | 158 | ADSL | 1.119935 | 0.497234 |
| SL004333 | Fibroblast growth factor 10 | FGF-10 | O15520 | 2255 | FGF10 | -1.121228 | 0.498952 |
| SL003522 | Endoplasmic reticulum resident protein 29 | ERP29 | P30040 | 10961 | ERP29 | 1.049472 | 0.498998 |
| SL000415 | Complement factor H | Factor H | P08603 | 3075 | CFH | 1.024873 | 0.499512 |
| HCE000104 | HCE000104 | HCE000104 | HCE000104 | HCE000104 | HCE000104 | 1.048887 | 0.500333 |
| SL006189 | Kinesin-like protein KIF23 | KIF23 | Q02241 | 9493 | KIF23 | -1.03138 | 0.500716 |
| SL005159 | Erythropoietin receptor | EPO-R | P19235 | 2057 | EPOR | -1.064186 | 0.503447 |
| SL014294 | Ephrin type-A receptor 10 | EPHAA | Q5JZY3 | 284656 | EPHA10 | 1.098176 | 0.503925 |
| SL000480 | Interleukin-4 | IL-4 | P05112 | 3565 | IL4 | -1.04387 | 0.503938 |
| SL010927 | Importin subunit beta-1 | IMB1 | Q14974 | 3837 | KPNB1 | 1.129901 | 0.50529 |
| SL004605 | 40S ribosomal protein SA | 40S ribosomal protein SA | P08865 | 3921 | RPSA | -1.113592 | 0.5059 |
| SL000396 | Cytochrome c | Cytochrome c | P99999 | 54205 | CYCS | -1.065112 | 0.506497 |
| SL004673 | Cathepsin S | Cathepsin S | P25774 | 1520 | CTSS | 1.035208 | 0.507595 |
| SL004805 | Cell adhesion molecule 1 | Nectin-like protein 2 | Q9BY67 | 23705 | CADM1 | 1.050804 | 0.508857 |
| SL001726 | Granulocyte-macrophage colony-stimulating factor | GM-CSF | P04141 | 1437 | CSF2 | -1.645729 | 0.508895 |
| SL000320 | C5a anaphylatoxin | C5a | P01031 | 727 | C5 | 1.072542 | 0.510022 |
| SL004876 | Kallistatin | Kallistatin | P29622 | 5267 | SERPINA4 | -1.057762 | 0.511415 |
| SL011532 | Persulfide dioxygenase ETHE1, mitochondrial | ETHE1 | O95571 | 23474 | ETHE1 | 1.044747 | 0.511801 |
| SL004438 | Cystatin-M | Cystatin M | Q15828 | 1474 | CST6 | -1.091799 | 0.512069 |
| SL005703 | Neurogenic locus notch homolog protein 1 | Notch 1 | P46531 | 4851 | NOTCH1 | 1.053657 | 0.512386 |
| SL001753 | Sialoadhesin | Sialoadhesin | Q9BZZ2 | 6614 | SIGLEC1 | 1.837181 | 0.513442 |
| SL007229 | Signal transducer and activator of transcription 6 | STAT6 | P42226 | 6778 | STAT6 | -1.155352 | 0.513612 |
| SL009948 | Plexin-B2 | PLXB2 | O15031 | 23654 | PLXNB2 | 1.050336 | 0.51462 |
| SL015728 | Glucokinase regulatory protein | GCKR | Q14397 | 2646 | GCKR | 1.080088 | 0.514847 |
| SL005220 | Sonic hedgehog protein | Sonic Hedgehog | Q15465 | 6469 | SHH | 1.0793 | 0.516551 |
| SL014983 | Histone H2B type 2-E | H2B2E | Q16778 | 8349 | HIST2H2BE | 1.118889 | 0.517713 |
| SL004154 | Neural cell adhesion molecule L1 | NCAM-L1 | P32004 | 3897 | L1CAM | 1.085439 | 0.51831 |
| SL004875 | Interleukin-1 receptor-like 2 | IL-1Rrp2 | Q9HB29 | 8808 | IL1RL2 | 1.024792 | 0.51911 |
| SL004134 | CD27 antigen | CD27 | P26842 | 939 | CD27 | 1.042797 | 0.519223 |
| SL002823 | L-Selectin | sL-Selectin | P14151 | 6402 | SELL | 1.060832 | 0.520737 |
| SL007100 | Leukotriene A-4 hydrolase | LKHA4 | P09960 | 4048 | LTA4H | -1.053877 | 0.521329 |
| SL000280 | Aspartate aminotransferase, cytoplasmic | GOT1 | P17174 | 2805 | GOT1 | 1.034236 | 0.521551 |
| SL009207 | Dynactin subunit 2 | Dynactin subunit 2 | Q13561 | 10540 | DCTN2 | 1.049258 | 0.521721 |
| SL010516 | Proto-oncogene tyrosine-protein kinase Src | SRCN1 | P12931 | 6714 | SRC | -1.360243 | 0.521814 |
| SL017611 | 14-3-3 protein family | 14-3-3 | P31946, P62258, P61981, Q04917, P27348, P63104, P31947 | 7529, 7531, 7532, 7533, 10971, 7534, 2810 | YWHAB,YWHAE,YWHAG,YWHAH,YWHAQ,YWHAZ,SFN | -1.2227 | 0.52194 |
| SL004009 | Ras-related C3 botulinum toxin substrate 1 | RAC1 | P63000 | 5879 | RAC1 | -1.165471 | 0.522768 |
| SL010524 | Serine/threonine-protein kinase WNK3 | WNK3 | Q9BYP7 | 65267 | WNK3 | -1.089094 | 0.522845 |
| SL012783 | WAP, kazal, immunoglobulin, kunitz and NTR domain-containing protein 1 | WFKN1 | Q96NZ8 | 117166 | WFIKKN1 | -1.092733 | 0.524367 |
| SL005491 | Opioid-binding protein/cell adhesion molecule | OBCAM | Q14982 | 4978 | OPCML | -1.027313 | 0.525364 |
| SL005508 | Carbonic anhydrase 9 | Carbonic anhydrase 9 | Q16790 | 768 | CA9 | -1.121702 | 0.526648 |
| SL004337 | Fibroblast growth factor 19 | FGF-19 | O95750 | 9965 | FGF19 | 1.010304 | 0.526671 |
| SL004782 | Tumor necrosis factor-inducible gene 6 protein | TSG-6 | P98066 | 7130 | TNFAIP6 | 1.031785 | 0.526892 |
| SL007642 | Angiopoietin-related protein 4 | ANGL4 | Q9BY76 | 51129 | ANGPTL4 | -1.019102 | 0.527733 |
| SL006476 | Glycylpeptide N-tetradecanoyltransferase 1 | NMT1 | P30419 | 4836 | NMT1 | -1.264355 | 0.528212 |
| SL000485 | Interleukin-9 | IL-9 | P15248 | 3578 | IL9 | 1.047816 | 0.529516 |
| SL012822 | Brain-specific serine protease 4 | BSSP4 | Q9GZN4 | 64063 | PRSS22 | 1.043755 | 0.533047 |
| SL001945 | E-selectin | sE-Selectin | P16581 | 6401 | SELE | 1.180292 | 0.533069 |
| SL001766 | Human Chorionic Gonadotropin | HCG | P01215,P01233 | 1081 1082 | CGA CGB | 1.687159 | 0.534161 |
| SL001731 | Gamma-enolase | NSE | P09104 | 2026 | ENO2 | -1.112406 | 0.534178 |
| SL011202 | Alpha-soluble NSF attachment protein | SNAA | P54920 | 8775 | NAPA | -1.038008 | 0.535628 |
| SL007756 | Growth/differentiation factor 2 | GDF2 | Q9UK05 | 2658 | GDF2 | -1.06393 | 0.5359 |
| SL010467 | Repulsive guidance molecule A | RGMA | Q96B86 | 56963 | RGMA | 1.053912 | 0.536082 |
| SL007356 | Neurogenic locus notch homolog protein 2 | NOTC2 | Q04721 | 4853 | NOTCH2 | -1.724092 | 0.536327 |
| SL000570 | Secretin | Secretin | P09683 | 6343 | SCT | -1.060922 | 0.538315 |
| SL002522 | Retinoblastoma-associated protein | Rb | P06400 | 5925 | RB1 | -1.061912 | 0.541474 |
| SL005173 | Interleukin-10 receptor subunit alpha | IL-10 Ra | Q13651 | 3587 | IL10RA | 1.049593 | 0.542595 |
| SL004642 | Disintegrin and metalloproteinase domain-containing protein 9 | ADAM 9 | Q13443 | 8754 | ADAM9 | -1.02125 | 0.542919 |
| SL008709 | Desmocollin-3 | DSC3 | Q14574 | 1825 | DSC3 | -1.070432 | 0.544452 |
| SL002078 | TGF-beta receptor type-2 | TGF-b R II | P37173 | 7048 | TGFBR2 | 1.255178 | 0.546823 |
| SL004556 | Complement decay-accelerating factor | DAF | P08174 | 1604 | CD55 | 1.070698 | 0.547507 |
| SL000586 | Thrombin | Thrombin | P00734 | 2147 | F2 | 1.095479 | 0.548044 |
| SL014228 | SLAM family member 6 | SLAF6 | Q96DU3 | 114836 | SLAMF6 | -1.005226 | 0.548114 |
| SL016130 | Transcription factor IIIB 90 kDa subunit | BRF-1 | Q92994 | 2972 | BRF1 | 1.107685 | 0.548267 |
| SL014269 | Killer cell immunoglobulin-like receptor 3DL2 | KI3L2 | P43630 | 3812 | KIR3DL2 | -1.51505 | 0.548363 |
| SL010384 | Testican-1 | Testican-1 | Q08629 | 6695 | SPOCK1 | -1.058773 | 0.548694 |
| SL004859 | Tumor necrosis factor receptor superfamily member 18 | GITR | Q9Y5U5 | 8784 | TNFRSF18 | -1.294056 | 0.548834 |
| SL018625 | Toll-like receptor 4:Lymphocyte antigen 96 complex | TLR4:MD-2 complex | O00206 Q9Y6Y9 | 7099 23643 | TLR4 LY96 | -1.04822 | 0.551913 |
| SL017528 | Ribonuclease H1 | RNase H1 | O60930 | 246243 | RNASEH1 | -2.01682 | 0.554413 |
| SL000640 | Nidogen-1 | Nidogen | P14543 | 4811 | NID1 | 1.043693 | 0.555307 |
| SL004326 | Tumor necrosis factor ligand superfamily member 18 | TNFSF18 | Q9UNG2 | 8995 | TNFSF18 | -1.017126 | 0.556617 |
| SL017189 | Kininogen-1 | Kininogen, HMW | P01042 | 3827 | KNG1 | 1.048894 | 0.556803 |
| SL004182 | Calcium-dependent phospholipase A2 | GV | P39877 | 5322 | PLA2G5 | -1.051824 | 0.559557 |
| SL010500 | Tyrosine-protein kinase Lyn, isoform B | LYNB | P07948 | 4067 | LYN | -1.275715 | 0.559885 |
| SL000527 | Matrix metalloproteinase-9 | MMP-9 | P14780 | 4318 | MMP9 | 1.097908 | 0.559931 |
| SL010372 | Enteropeptidase | Enterokinase | P98073 | 5651 | TMPRSS15 | 1.026005 | 0.561105 |
| SL003220 | C3a anaphylatoxin des Arginine | C3adesArg | P01024 | 718 | C3 | -1.062346 | 0.564108 |
| SL004919 | Peroxiredoxin-1 | Peroxiredoxin-1 | Q06830 | 5052 | PRDX1 | -1.002014 | 0.564385 |
| SL004910 | Fructose-bisphosphate aldolase A | aldolase A | P04075 | 226 | ALDOA | -1.118855 | 0.564655 |
| SL000309 | Complement C1q subcomponent | C1q | P02745 P02746 P02747 | 712 713 714 | C1QA C1QB C1QC | 1.014073 | 0.566835 |
| SL009045 | Ectonucleotide pyrophosphatase/phosphodiesterase family member 7 | ENPP7 | Q6UWV6 | 339221 | ENPP7 | 1.239456 | 0.566858 |
| SL000300 | Beta-endorphin | b-Endorphin | P01189 | 5443 | POMC | -1.088486 | 0.568174 |
| SL003168 | C-C motif chemokine 27 | CTACK | Q9Y4X3 | 10850 | CCL27 | -1.104206 | 0.569018 |
| SL005261 | Tyrosine-protein phosphatase non-receptor type 2 | TCPTP | P17706 | 5771 | PTPN2 | -1.115969 | 0.569324 |
| SL004467 | Protein Z-dependent protease inhibitor | protein Z inhibitor | Q9UK55 | 51156 | SERPINA10 | 1.065063 | 0.569344 |
| SL011405 | cGMP-specific 3',5'-cyclic phosphodiesterase | PDE5A | O76074 | 8654 | PDE5A | -1.215901 | 0.569696 |
| SL011180 | Hexokinase-1 | HXK1 | P19367 | 3098 | HK1 | -1.033985 | 0.570144 |
| SL000551 | Protein kinase C alpha type | PKC-A | P17252 | 5578 | PRKCA | -1.375767 | 0.571066 |
| SL009202 | Junctional adhesion molecule-like | JAML1 | Q86YT9 | 120425 | AMICA1 | -2.800435 | 0.571292 |
| SL007151 | Glycerol-3-phosphate dehydrogenase [NAD(+)], cytoplasmic | GPDA | P21695 | 2819 | GPD1 | -1.054423 | 0.571483 |
| SL016555 | Dual 3',5'-cyclic-AMP and -GMP phosphodiesterase 11A | PDE11 | Q9HCR9 | 50940 | PDE11A | -1.077721 | 0.573343 |
| SL000509 | Tumor necrosis factor receptor superfamily member 3 | Lymphotoxin b R | P36941 | 4055 | LTBR | 1.049839 | 0.573968 |
| SL001905 | Mesothelin | Mesothelin | Q13421 | 10232 | MSLN | -1.10868 | 0.575185 |
| SL001721 | Platelet endothelial cell adhesion molecule | PECAM-1 | P16284 | 5175 | PECAM1 | -1.058855 | 0.57564 |
| SL000057 | Thymidine kinase, cytosolic | Thymidine kinase | P04183 | 7083 | TK1 | -1.104553 | 0.575726 |
| SL001999 | E3 ubiquitin-protein ligase Mdm2 | MDM2 | Q00987 | 4193 | MDM2 | -1.555437 | 0.576395 |
| HCE001796 | HCE001796 | HCE001796 | HCE001796 | HCE001796 | HCE001796 | -1.108006 | 0.576762 |
| SL014188 | Natural killer cell receptor 2B4 | CD244 | Q9BZW8 | 51744 | CD244 | 1.212776 | 0.577045 |
| SL005687 | Eukaryotic translation initiation factor 5A-1 | eIF-5A-1 | P63241 | 1984 | EIF5A | -1.084477 | 0.577586 |
| SL008810 | Neuronal growth regulator 1 | NEGR1 | Q7Z3B1 | 257194 | NEGR1 | 1.052744 | 0.580352 |
| SL010462 | Ficolin-1 | FCN1 | O00602 | 2219 | FCN1 | -1.053845 | 0.580364 |
| SL004827 | Protein S100-A6 | S100A6 | P06703 | 6277 | S100A6 | 1.146457 | 0.581395 |
| SL006523 | Lactadherin | MFGM | Q08431 | 4240 | MFGE8 | 1.018991 | 0.582442 |
| SL002093 | Histone H2A.z | Histone H2A.z | P0C0S5 | 3015 | H2AFZ | -1.091659 | 0.583434 |
| SL004148 | Interleukin-12 receptor subunit beta-1 | IL-12 Rb1 | P42701 | 3594 | IL12RB1 | 1.039421 | 0.58464 |
| SL002922 | Intercellular adhesion molecule 1 | sICAM-1 | P05362 | 3383 | ICAM1 | 1.06012 | 0.585256 |
| SL004781 | Tyrosine-protein kinase CSK | CSK | P41240 | 1445 | CSK | -1.315253 | 0.586832 |
| SL010489 | Calcium/calmodulin-dependent protein kinase type 1 | CAMK1 | Q14012 | 8536 | CAMK1 | -1.066669 | 0.588388 |
| SL004469 | Amyloid beta A4 protein | amyloid precursor protein | P05067 | 351 | APP | -1.068346 | 0.590068 |
| SL017529 | Kelch-like ECH-associated protein 1 | KEAP1 | Q14145 | 9817 | KEAP1 | -3.302763 | 0.591929 |
| SL014113 | Neuroligin-4, X-linked | NLGNX | Q8N0W4 | 57502 | NLGN4X | 1.092531 | 0.592254 |
| SL004644 | Ectodysplasin-A, secreted form | EDA | Q92838 | 1896 | EDA | 1.041884 | 0.592328 |
| SL020171 | HERV-H LTR-associating protein 2 | HHLA2 | Q9UM44 | NaN | HHLA2 | -1.577044 | 0.592612 |
| SL000589 | Thyroid Stimulating Hormone | TSH | P01215 P01222 | 1081 12372 | CGA TSHB | 1.144085 | 0.593238 |
| SL004144 | GDNF family receptor alpha-3 | GFRa-3 | O60609 | 2676 | GFRA3 | -1.071388 | 0.597092 |
| SL001897 | Kunitz-type protease inhibitor 2 | SPINT2 | O43291 | 10653 | SPINT2 | 1.024188 | 0.598374 |
| SL002684 | Macrophage colony-stimulating factor 1 | CSF-1 | P09603 | 1435 | CSF1 | 1.052473 | 0.60067 |
| SL005164 | Galectin-2 | Galectin-2 | P05162 | 3957 | LGALS2 | 1.085726 | 0.602042 |
| SL004714 | Leukemia inhibitory factor receptor | LIF sR | P42702 | 3977 | LIFR | 1.071107 | 0.603055 |
| SL002762 | Chromogranin-A | CgA | P10645 | 1113 | CHGA | -1.364433 | 0.603889 |
| SL007640 | C-type lectin domain family 7 member A | CLC7A | Q9BXN2 | 64581 | CLEC7A | -1.066631 | 0.606644 |
| SL010973 | Proteasome subunit alpha type-1 | PSA1 | P25786 | 5682 | PSMA1 | -1.990679 | 0.607294 |
| SL007888 | Cystatin-S | Cystatin-S | P01036 | 1472 | CST4 | -1.343768 | 0.609098 |
| SL000468 | Immunoglobulin M | IgM | P01871 | 3507 3512 50802 3535 | IGHM IGJ IGK@ IGL@ | 1.025378 | 0.610191 |
| SL004635 | Tumor necrosis factor ligand superfamily member 8 | CD30 Ligand | P32971 | 944 | TNFSF8 | -1.09603 | 0.611231 |
| SL010610 | C-type lectin domain family 4 member K | CLC4K | Q9UJ71 | 50489 | CD207 | 1.004799 | 0.611701 |
| SL004180 | Tumor necrosis factor receptor superfamily member 8 | CD30 | P28908 | 943 | TNFRSF8 | -1.056778 | 0.612038 |
| SL000510 | Lysozyme C | Lysozyme | P61626 | 4069 | LYZ | 1.019413 | 0.612278 |
| SL005223 | Interleukin-27 receptor subunit alpha | TCCR | Q6UWB1 | 9466 | IL27RA | 1.036049 | 0.612543 |
| SL006992 | Matrilin-3 | MATN3 | O15232 | 4148 | MATN3 | -1.034849 | 0.613759 |
| SL001973 | Mammaglobin-B | Mammaglobin 2 | O75556 | 4246 | SCGB2A1 | -1.023065 | 0.614744 |
| SL001947 | Melanoma-derived growth regulatory protein | MIA | Q16674 | 8190 | MIA | 1.045721 | 0.615075 |
| SL000645 | Stromelysin-2 | MMP-10 | P09238 | 4319 | MMP10 | 1.071111 | 0.61636 |
| SL003170 | C-C motif chemokine 26 | Eotaxin-3 | Q9Y258 | 10344 | CCL26 | -1.112906 | 0.616986 |
| SL013988 | Carbohydrate sulfotransferase 2 | CHST2 | Q9Y4C5 | 9435 | CHST2 | -1.038152 | 0.618395 |
| SL011211 | Eukaryotic translation initiation factor 4 gamma 2 | IF4G2 | P78344 | 1982 | EIF4G2 | -1.275608 | 0.618968 |
| SL004649 | Tumor necrosis factor ligand superfamily member 4 | OX40 Ligand | P23510 | 7292 | TNFSF4 | 1.100057 | 0.62025 |
| SL010470 | Semaphorin-3E | Semaphorin 3E | O15041 | 9723 | SEMA3E | 1.061193 | 0.62272 |
| SL004921 | Nucleoside diphosphate kinase B | NDP kinase B | P22392 | 4831 | NME2 | -1.120263 | 0.623689 |
| SL004120 | Discoidin domain-containing receptor 2 | Discoidin domain receptor 2 | Q16832 | 4921 | DDR2 | -1.045332 | 0.625415 |
| SL012538 | Cerebral dopamine neurotrophic factor | ARMEL | Q49AH0 | 441549 | CDNF | 1.024866 | 0.626957 |
| SL003461 | Corticotropin | ACTH | P01189 | 5443 | POMC | -1.093603 | 0.627315 |
| SL005193 | Junctional adhesion molecule B | JAM-B | P57087 | 58494 | JAM2 | 1.027968 | 0.630587 |
| SL003305 | Interleukin-2 receptor subunit alpha | IL-2 sRa | P01589 | 3559 | IL2RA | -1.139679 | 0.630628 |
| SL003770 | Secreted frizzled-related protein 1 | SARP-2 | Q8N474 | 6422 | SFRP1 | 1.233436 | 0.630942 |
| SL005846 | Moesin | Moesin | P26038 | 4478 | MSN | -1.057004 | 0.632789 |
| SL003188 | C-X-C motif chemokine 9 | MIG | Q07325 | 4283 | CXCL9 | 1.078027 | 0.633621 |
| SL003994 | Bone morphogenetic protein 1 | BMP-1 | P13497 | 649 | BMP1 | -1.068703 | 0.634178 |
| SL007121 | Cathepsin E | CATE | P14091 | 1510 | CTSE | -1.28611 | 0.634185 |
| SL002662 | Coagulation Factor XI | Coagulation Factor XI | P03951 | 2160 | F11 | 1.051434 | 0.63471 |
| SL000433 | Glucagon | Glucagon | P01275 | 2641 | GCG | 1.006779 | 0.635007 |
| SL014029 | Sphingosine kinase 2 | SPHK2 | Q9NRA0 | 56848 | SPHK2 | -1.02811 | 0.636323 |
| SL006544 | Transforming growth factor-beta-induced protein ig-h3 | BGH3 | Q15582 | 7045 | TGFBI | 1.073754 | 0.63653 |
| SL001691 | Fibroblast growth factor 7 | FGF7 | P21781 | 2252 | FGF7 | -1.037763 | 0.636999 |
| SL000493 | L-lactate dehydrogenase B chain | LDH-H 1 | P07195 | 3945 | LDHB | 1.026628 | 0.637948 |
| SL004147 | Interleukin-10 receptor subunit beta | IL-10 Rb | Q08334 | 3588 | IL10RB | 1.02383 | 0.638844 |
| SL003303 | C-C motif chemokine 28 | CCL28 | Q9NRJ3 | 56477 | CCL28 | 1.230326 | 0.641529 |
| SL003643 | Glutathione S-transferase P | Glutathione S-transferase Pi | P09211 | 2950 | GSTP1 | -1.030599 | 0.642348 |
| SL003648 | Rab GDP dissociation inhibitor beta | Rab GDP dissociation inhibitor beta | P50395 | 2665 | GDI2 | 1.023367 | 0.642881 |
| SL010373 | Immunoglobulin alpha Fc receptor | FCAR | P24071 | 2204 | FCAR | -1.036469 | 0.643341 |
| SL000470 | Interleukin-11 | IL-11 | P20809 | 3589 | IL11 | -1.16235 | 0.644373 |
| SL004716 | Mitogen-activated protein kinase 9 | JNK2 | P45984 | 5601 | MAPK9 | 1.015332 | 0.644855 |
| SL000401 | Neutrophil elastase | Elastase | P08246 | 1991 | ELANE | -1.115028 | 0.645094 |
| SL004125 | Insulin receptor | IR | P06213 | 3643 | INSR | 1.063606 | 0.645855 |
| SL010455 | Contactin-5 | Contactin-5 | O94779 | 53942 | CNTN5 | 1.01109 | 0.64702 |
| SL008193 | Nidogen-2 | NID2 | Q14112 | 22795 | NID2 | 1.019726 | 0.647448 |
| SL004349 | Interferon lambda-2 | IFN-lambda 2 | Q8IZJ0 | 282616 | IFNL2 | 1.008144 | 0.647802 |
| SL008008 | Arginase-1 | ARGI1 | P05089 | 383 | ARG1 | 1.258449 | 0.649638 |
| SL005201 | Muellerian-inhibiting factor | MIS | P03971 | 268 | AMH | 1.045033 | 0.649703 |
| SL004346 | Interleukin-20 | IL-20 | Q9NYY1 | 50604 | IL20 | 1.026048 | 0.649836 |
| SL000318 | Complement C4b | C4b | P0C0L4 P0C0L5 | 720 721 | C4A C4B | -1.133637 | 0.650121 |
| SL000588 | Thyroid peroxidase | TMA | P07202 | 7173 | TPO | -1.032032 | 0.650568 |
| SL010612 | Dickkopf-related protein 4 | Dkk-4 | Q9UBT3 | 27121 | DKK4 | 1.051875 | 0.651249 |
| SL019096 | E3 SUMO-protein ligase PIAS4 | PIAS4 | Q8N2W9 | 51588 | PIAS4 | -1.023784 | 0.652396 |
| SL004760 | Platelet-activating factor acetylhydrolase IB subunit beta | PAFAH beta subunit | P68402 | 5049 | PAFAH1B2 | -1.029534 | 0.654227 |
| SL004160 | BDNF/NT-3 growth factors receptor | TrkB | Q16620 | 4915 | NTRK2 | 1.040917 | 0.654938 |
| SL008176 | Proteasome activator complex subunit 1 | PSME1 | Q06323 | 5720 | PSME1 | -1.09122 | 0.65598 |
| SL008039 | Alcohol dehydrogenase [NADP(+)] | AK1A1 | P14550 | 10327 | AKR1A1 | -1.065569 | 0.661032 |
| SL016548 | AMP Kinase (alpha1beta1gamma1) | AMPK a1b1g1 | Q13131 Q9Y478 P54619 | 5562 5564 5571 | PRKAA1 PRKAB1 PRKAG1 | -1.007241 | 0.661143 |
| SL005588 | Tyrosine-protein kinase Fer | FER | P16591 | 2241 | FER | -1.209894 | 0.661894 |
| SL000055 | Cadherin-1 | Cadherin E | P12830 | 999 | CDH1 | 1.026493 | 0.662196 |
| SL001718 | Interleukin-13 | IL-13 | P35225 | 3596 | IL13 | -1.063353 | 0.662399 |
| SL005174 | Interleukin-17 receptor B | IL-17B R | Q9NRM6 | 55540 | IL17RB | 1.181943 | 0.662872 |
| SL008590 | Olfactomedin-4 | Olfactomedin-4 | Q6UX06 | 10562 | OLFM4 | -1.01951 | 0.66302 |
| HCE000414 | HCE000414 | HCE000414 | HCE000414 | HCE000414 | HCE000414 | 1.022449 | 0.663444 |
| SL004865 | Cadherin-6 | Cadherin-6 | P55285 | 1004 | CDH6 | -1.022615 | 0.66367 |
| SL007221 | Signal transducer and activator of transcription 3 | STAT3 | P40763 | 6774 | STAT3 | -1.168548 | 0.66447 |
| SL000346 | Cathepsin H | Cathepsin H | P09668 | 1512 | CTSH | 1.015426 | 0.669009 |
| SL004466 | Heparin cofactor 2 | Heparin cofactor II | P05546 | 3053 | SERPIND1 | 1.051888 | 0.669726 |
| SL005204 | NKG2-D type II integral membrane protein | NKG2D | P26718 | 22914 | KLRK1 | -1.054852 | 0.67079 |
| SL003990 | Fibroblast growth factor receptor 2 | FGFR-2 | P21802 | 2263 | FGFR2 | -2.499395 | 0.672157 |
| SL000377 | Creatine kinase B-type | CK-BB | P12277 | 1152 | CKB | 1.08693 | 0.674692 |
| SL004097 | Mothers against decapentaplegic homolog 3 | SMAD3 | P84022 | 4088 | SMAD3 | -1.241718 | 0.674765 |
| SL000553 | Protein kinase C beta type (splice variant beta-II) | PKC-B-II | P05771 | 5579 | PRKCB | -1.31146 | 0.674943 |
| SL011809 | dCTP pyrophosphatase 1 | XTP3A | Q9H773 | 79077 | DCTPP1 | 1.04631 | 0.676204 |
| SL016557 | 3-hydroxy-3-methylglutaryl-coenzyme A reductase | HMGR | P04035 | 3156 | HMGCR | 1.094883 | 0.676234 |
| SL004064 | Phospholipase A2 | GIB | P04054 | 5319 | PLA2G1B | -1.061211 | 0.676514 |
| SL004183 | Cadherin-3 | P-Cadherin | P22223 | 1001 | CDH3 | -1.027454 | 0.677098 |
| SL000456 | Complement C3b, inactivated | iC3b | P01024 | 718 | C3 | 1.044465 | 0.677256 |
| SL010379 | Semaphorin-3A | Semaphorin 3A | Q14563 | 10371 | SEMA3A | 1.033676 | 0.6787 |
| SL003190 | C-C motif chemokine 15 | MIP-5 | Q16663 | 6359 | CCL15 | 1.005523 | 0.679301 |
| SL002782 | Adrenomedullin | Adrenomedullin | P35318 | 133 | ADM | -1.018855 | 0.680085 |
| SL008614 | Ephrin-B1 | EFNB1 | P98172 | 1947 | EFNB1 | -1.024139 | 0.680491 |
| SL016563 | Mitochondrial glutamate carrier 2 | GHC2 | Q9H1K4 | 83733 | SLC25A18 | -1.021493 | 0.680587 |
| SL005222 | Scavenger receptor class F member 2 | SREC-II | Q96GP6 | 91179 | SCARF2 | -1.052859 | 0.680609 |
| SL000695 | Neutrophil gelatinase-associated lipocalin | Lipocalin 2 | P80188 | 3934 | LCN2 | 1.037396 | 0.681015 |
| SL005430 | Secreted and transmembrane protein 1 | SECTM1 | Q8WVN6 | 6398 | SECTM1 | -1.03417 | 0.681476 |
| SL004415 | Angiotensin-converting enzyme 2 | ACE2 | Q9BYF1 | 59272 | ACE2 | -1.022052 | 0.682805 |
| SL000358 | Coagulation factor VII | Coagulation Factor VII | P08709 | 2155 | F7 | 1.034793 | 0.686327 |
| SL003201 | Vascular endothelial growth factor receptor 2 | VEGF sR2 | P35968 | 3791 | KDR | 1.043515 | 0.688103 |
| SL000345 | Cathepsin G | Cathepsin G | P08311 | 1511 | CTSG | -1.089697 | 0.688552 |
| SL004652 | Wnt inhibitory factor 1 | WIF-1 | Q9Y5W5 | 11197 | WIF1 | 1.016546 | 0.688753 |
| SL002704 | Pleiotrophin | PTN | P21246 | 5764 | PTN | 1.027274 | 0.689716 |
| SL004489 | Toll-like receptor 4 | TLR4 | O00206 | 7099 | TLR4 | -1.195711 | 0.690446 |
| SL003679 | Cation-independent mannose-6-phosphate receptor | IGF-II receptor | P11717 | 3482 | IGF2R | 1.021664 | 0.691157 |
| SL013969 | Kynureninase | KYNU | Q16719 | 8942 | KYNU | 1.055273 | 0.691398 |
| SL000339 | Carbonic anhydrase 2 | carbonic anhydrase II | P00918 | 760 | CA2 | -1.141448 | 0.693321 |
| SL005699 | Apolipoprotein L1 | Apo L1 | O14791 | 8542 | APOL1 | 1.047744 | 0.695603 |
| SL005165 | Galectin-4 | Galectin-4 | P56470 | 3960 | LGALS4 | -1.079338 | 0.69724 |
| SL000251 | Alpha-2-HS-glycoprotein | a2-HS-Glycoprotein | P02765 | 197 | AHSG | 1.04576 | 0.698895 |
| SL010378 | Proto-oncogene tyrosine-protein kinase receptor Ret | RET | P07949 | 5979 | RET | 1.027137 | 0.698976 |
| SL004336 | Fibroblast growth factor 18 | FGF-18 | O76093 | 8817 | FGF18 | 1.015624 | 0.699275 |
| SL007311 | Synaptosomal-associated protein 25 | SNP25 | P60880 | 6616 | SNAP25 | -1.014822 | 0.700313 |
| SL008072 | Collagen alpha-1(VIII) chain | CO8A1 | P27658 | 1295 | COL8A1 | -1.671095 | 0.701014 |
| SL009089 | Brevican core protein | PGCB | Q96GW7 | 63827 | BCAN | -1.072003 | 0.701751 |
| SL000460 | Immunoglobulin D | IgD | P01880 | 3495 50802 3535 | IGHD IGK@ IGL@ | -1.453016 | 0.703098 |
| SL003862 | CD40 ligand | CD40 ligand, soluble | P29965 | 959 | CD40LG | -1.014184 | 0.703341 |
| SL010830 | Protein 4.1 | 41 | P11171 | 2035 | EPB41 | 1.408789 | 0.70452 |
| SL000605 | Ubiquitin+1, truncated mutation for UbB | Ubiquitin+1 | P62979 | 6233 | RPS27A | 1.052233 | 0.70614 |
| SL010496 | Cyclin-dependent kinase 5:Cyclin-dependent kinase 5 activator 1 complex | CDK5/p35 | Q00535 Q15078 | 1020 8851 | CDK5 CDK5R1 | 1.094125 | 0.706293 |
| SL010457 | Delta-like protein 4 | DLL4 | Q9NR61 | 54567 | DLL4 | 1.020002 | 0.706823 |
| SL000360 | Coagulation Factor X | Coagulation Factor X | P00742 | 2159 | F10 | -1.043946 | 0.707734 |
| SL004159 | Tumor necrosis factor receptor superfamily member 10D | TRAIL R4 | Q9UBN6 | 8793 | TNFRSF10D | -1.007349 | 0.707856 |
| SL003066 | Pigment epithelium-derived factor | PEDF | P36955 | 5176 | SERPINF1 | 1.038881 | 0.708852 |
| SL008516 | Cystatin-SA | CYTT | P09228 | 1470 | CST2 | 1.004131 | 0.709062 |
| SL001890 | Tumor-associated calcium signal transducer 2 | GA733-1 protein | P09758 | 4070 | TACSTD2 | -1.113378 | 0.709567 |
| SL003658 | Aflatoxin B1 aldehyde reductase member 2 | Aflatoxin B1 aldehyde reductase | O43488 | 8574 | AKR7A2 | -1.133251 | 0.710305 |
| SL005200 | MHC class I polypeptide-related sequence B | MICB | Q29980 | 4277 | MICB | -1.284071 | 0.711578 |
| SL008956 | Arylsulfatase B | ARSB | P15848 | 411 | ARSB | 1.052045 | 0.712936 |
| SL004515 | Peptidoglycan recognition protein 1 | PGRP-S | O75594 | 8993 | PGLYRP1 | -2.690988 | 0.714931 |
| SL009988 | Disintegrin and metalloproteinase domain-containing protein 12 | ADAM12 | O43184 | 8038 | ADAM12 | -1.771829 | 0.716038 |
| SL001717 | Interleukin-10 | IL-10 | P22301 | 3586 | IL10 | -1.158558 | 0.717539 |
| SL003196 | C-C motif chemokine 17 | TARC | Q92583 | 6361 | CCL17 | 1.010728 | 0.71798 |
| SL013490 | Brother of CDO | BOC | Q9BWV1 | 91653 | BOC | 1.043523 | 0.718121 |
| SL000565 | Renin | Renin | P00797 | 5972 | REN | -1.256498 | 0.718268 |
| SL003328 | Complement factor I | Factor I | P05156 | 3426 | CFI | -1.028781 | 0.719841 |
| SL008588 | SLAM family member 5 | SLAF5 | Q9UIB8 | 8832 | CD84 | 1.025972 | 0.721743 |
| SL021043 | Growth/differentiation factor 11/8 | GDF-11/8 | O95390 O14793 | NaN | GDF11 MSTN | -1.053591 | 0.722333 |
| SL006713 | Collectin-11 | Collectin Kidney 1 | Q9BWP8 | 78989 | COLEC11 | 1.069049 | 0.723323 |
| SL000528 | NADPH--cytochrome P450 reductase | NADPH-P450 Oxidoreductase | P16435 | 5447 | POR | -1.005779 | 0.723427 |
| SL011768 | NudC domain-containing protein 3 | NUDC3 | Q8IVD9 | 23386 | NUDCD3 | 1.391637 | 0.723454 |
| SL008822 | Adhesion G protein-coupled receptor E2 | EMR2 | Q9UHX3 | 30817 | ADGRE2 | -1.025055 | 0.724762 |
| SL010495 | Cyclin-dependent kinase 2:Cyclin-A2 complex | CDK2/cyclin A | P24941 P20248 | 1017 890 | CDK2 CCNA2 | 1.01899 | 0.72504 |
| SL004367 | Dickkopf-related protein 1 | DKK1 | O94907 | 22943 | DKK1 | 1.01303 | 0.725078 |
| SL004594 | Troponin I, fast skeletal muscle | Troponin I, skeletal, fast twitch | P48788 | 7136 | TNNI2 | -1.128306 | 0.726034 |
| SL018946 | Ubiquitin-conjugating enzyme E2 G2 | UB2G2 | P60604 | 7327 | UBE2G2 | -1.017318 | 0.72768 |
| SL004354 | Interleukin-19 | IL-19 | Q9UHD0 | 29949 | IL19 | 1.095222 | 0.727903 |
| SL005183 | Interleukin-22 receptor subunit alpha-2 | IL-22BP | Q969J5 | 116379 | IL22RA2 | -1.062469 | 0.728796 |
| SL006480 | Trypsin-3 | TRY3 | P35030 | 5646 | PRSS3 | -1.001576 | 0.728922 |
| SL000541 | Plasminogen | Plasminogen | P00747 | 5340 | PLG | 1.018481 | 0.729876 |
| SL003733 | Diablo homolog, mitochondrial | SMAC | Q9NR28 | 56616 | DIABLO | 1.011429 | 0.730276 |
| SL019100 | E3 ubiquitin-protein ligase CHIP | CHIP | Q9UNE7 | 10273 | STUB1 | -1.107048 | 0.731538 |
| SL005403 | Vesicular integral-membrane protein VIP36 | Lectin, mannose-binding 2 | Q12907 | 10960 | LMAN2 | -1.010309 | 0.73243 |
| SL010521 | Tyrosine-protein kinase BTK | BTK | Q06187 | 695 | BTK | -1.227063 | 0.734285 |
| SL000590 | Thyroxine-binding globulin | Thyroxine-Binding Globulin | P05543 | 6906 | SERPINA7 | -1.034261 | 0.736811 |
| SL006088 | Sphingosine kinase 1 | Sphingosine kinase 1 | Q9NYA1 | 8877 | SPHK1 | -1.128547 | 0.737965 |
| SL000006 | Plasminogen activator inhibitor 1 | PAI-1 | P05121 | 5054 | SERPINE1 | -1.099131 | 0.739813 |
| SL000540 | Plasmin | Plasmin | P00747 | 5340 | PLG | -1.249737 | 0.741312 |
| SL018947 | Ubiquitin carboxyl-terminal hydrolase 25 | UBP25 | Q9UHP3 | 29761 | USP25 | -1.059745 | 0.742176 |
| HCE004331 | HCE004331 | HCE004331 | HCE004331 | HCE004331 | HCE004331 | 1.025127 | 0.742219 |
| SL000617 | Alanine aminotransferase 1 | ALT | P24298 | 2875 | GPT | 1.010006 | 0.745123 |
| SL010514 | Protein-tyrosine kinase 6 | PTK6 | Q13882 | 5753 | PTK6 | -1.226907 | 0.745551 |
| SL000521 | Interstitial collagenase | MMP-1 | P03956 | 4312 | MMP1 | 1.380812 | 0.747411 |
| SL008760 | SLIT and NTRK-like protein 1 | SLIK1 | Q96PX8 | 114798 | SLITRK1 | 1.159844 | 0.750264 |
| SL000560 | P-selectin | P-Selectin | P16109 | 6403 | SELP | -1.065615 | 0.7505 |
| SL000478 | Interleukin-2 | IL-2 | P60568 | 3558 | IL2 | -1.057451 | 0.750823 |
| SL000047 | Insulin-like growth factor I | IGF-I | P05019 | 3479 | IGF1 | 1.018795 | 0.751185 |
| SL008522 | Semaphorin-5A | SEM5A | Q13591 | 9037 | SEMA5A | -1.143174 | 0.752698 |
| SL004156 | Tumor necrosis factor receptor superfamily member 10A | TRAIL R1 | O00220 | 8797 | TNFRSF10A | -1.040838 | 0.754038 |
| SL005155 | Teratocarcinoma-derived growth factor 1 | Cripto | P13385 | 6997 | TDGF1 | -1.264387 | 0.754561 |
| SL006268 | NSFL1 cofactor p47 | NSF1C | Q9UNZ2 | 55968 | NSFL1C | 1.097699 | 0.757958 |
| SL000011 | alpha-Fetoprotein | AFP | P02771 | 174 | AFP | -1.028786 | 0.760647 |
| SL004901 | Protein disulfide-isomerase | Protein disulfide-isomerase | P07237 | 5034 | P4HB | -1.039433 | 0.760933 |
| SL000024 | Tissue Factor | TF | P13726 | 2152 | F3 | -1.033003 | 0.76107 |
| SL000836 | Hemoglobin | Hemoglobin | P69905, P68871 | 3039 3043 | HBA1 HBB | -2.161941 | 0.761591 |
| SL005266 | Alpha-synuclein | a-Synuclein | P37840 | 6622 | SNCA | -1.01475 | 0.761673 |
| SL005153 | Ciliary neurotrophic factor receptor subunit alpha | CNTFR alpha | P26992 | 1271 | CNTFR | 1.051108 | 0.762403 |
| SL000442 | Somatotropin | HGH | P01241 | 2688 | GH1 | -1.040526 | 0.762837 |
| SL000003 | Angiogenin | Angiogenin | P03950 | 283 | ANG | 1.026013 | 0.763202 |
| SL014093 | Ectonucleoside triphosphate diphosphohydrolase 3 | ENTP3 | O75355 | 956 | ENTPD3 | -1.18626 | 0.766163 |
| SL010368 | Alpha-L-iduronidase | IDUA | P35475 | 3425 | IDUA | 1.042272 | 0.767041 |
| SL007181 | Non-receptor tyrosine-protein kinase TYK2 | TYK2 | P29597 | 7297 | TYK2 | -1.024834 | 0.767218 |
| SL000004 | Fibroblast growth factor 2 | bFGF | P09038 | 2247 | FGF2 | 1.148184 | 0.76891 |
| SL005236 | Protein NOV homolog | NovH | P48745 | 4856 | NOV | 1.02243 | 0.769118 |
| SL004350 | Interleukin-17B | IL-17B | Q9UHF5 | 27190 | IL17B | 1.107008 | 0.769344 |
| SL004692 | Cytokine receptor-like factor 1:Cardiotrophin-like cytokine factor 1 Complex | CLF-1/CLC Complex | O75462 Q9UBD9 | 9244 23529 | CRLF1 CLCF1 | 1.071015 | 0.770223 |
| SL000314 | Complement C3b | C3b | P01024 | 718 | C3 | 1.096457 | 0.772922 |
| SL004492 | Toll-like receptor 2 | TLR2 | O60603 | 7097 | TLR2 | 1.021517 | 0.773098 |
| SL016129 | Protein FAM107B | FAM107B | Q9H098 | 83641 | FAM107B | 1.021814 | 0.77415 |
| SL006132 | Lamin-B1 | Lamin-B1 | P20700 | 4001 | LMNB1 | 1.035347 | 0.774745 |
| SL000414 | Complement factor B | Factor B | P00751 | 629 | CFB | 1.011554 | 0.775163 |
| SL013872 | Low-density lipoprotein receptor-related protein 1B | LRP1B | Q9NZR2 | 53353 | LRP1B | 1.041429 | 0.775503 |
| SL004516 | Mannose-binding protein C | MBL | P11226 | 4153 | MBL2 | -1.037634 | 0.775586 |
| SL018900 | Formimidoyltransferase-cyclodeaminase | FTCD | O95954 | 10841 | FTCD | -1.644117 | 0.775831 |
| SL004010 | Mast/stem cell growth factor receptor Kit | SCF sR | P10721 | 3815 | KIT | -1.00391 | 0.777927 |
| SL003542 | Histone-lysine N-methyltransferase EHMT2 | NG36 | Q96KQ7 | 10919 | EHMT2 | -1.00523 | 0.78377 |
| SL000440 | Hemopexin | Hemopexin | P02790 | 3263 | HPX | -1.001 | 0.783808 |
| SL004866 | Carbonic anhydrase 1 | Carbonic anhydrase I | P00915 | 759 | CA1 | 1.3442 | 0.784023 |
| SL004067 | Group 10 secretory phospholipase A2 | GX | O15496 | 8399 | PLA2G10 | -1.01154 | 0.786116 |
| SL002763 | Kallikrein-11 | Kallikrein 11 | Q9UBX7 | 11012 | KLK11 | 1.013112 | 0.786141 |
| SL014468 | SH2 domain-containing protein 1A | SH21A | O60880 | 4068 | SH2D1A | 1.132459 | 0.786481 |
| SL003951 | Brain-derived neurotrophic factor | BDNF | P23560 | 627 | BDNF | -1.079694 | 0.786579 |
| SL008909 | Legumain | LGMN | Q99538 | 5641 | LGMN | 1.016847 | 0.789305 |
| SL008639 | Iduronate 2-sulfatase | IDS | P22304 | 3423 | IDS | -1.039011 | 0.789815 |
| SL001796 | Myeloperoxidase | Myeloperoxidase | P05164 | 4353 | MPO | 1.018252 | 0.789906 |
| SL003755 | Induced myeloid leukemia cell differentiation protein Mcl-1 | Mcl-1 | Q07820 | 4170 | MCL1 | -1.023567 | 0.790754 |
| SL000131 | Proliferating cell nuclear antigen | PCNA | P12004 | 5111 | PCNA | -1.009559 | 0.79135 |
| SL005574 | Aminoacylase-1 | Aminoacylase-1 | Q03154 | 95 | ACY1 | -1.051977 | 0.792109 |
| SL006910 | Cathepsin L2 | Cathepsin V | O60911 | 1515 | CTSV | 1.039081 | 0.793154 |
| SL008421 | A disintegrin and metalloproteinase with thrombospondin motifs 1 | ATS1 | Q9UHI8 | 9510 | ADAMTS1 | -1.008368 | 0.793903 |
| SL015510 | Ubiquitin-like protein ISG15 | UCRP | P05161 | 9636 | ISG15 | 1.119535 | 0.795293 |
| SL014288 | Cell surface glycoprotein CD200 receptor 1 | MO2R1 | Q8TD46 | 131450 | CD200R1 | -1.115539 | 0.795512 |
| SL006197 | Mitochondrial import inner membrane translocase subunit TIM14 | DnaJ homolog | Q96DA6 | 131118 | DNAJC19 | -1.03122 | 0.799562 |
| SL002071 | Gastrin-releasing peptide | Gastrin-releasing peptide | P07492 | 2922 | GRP | 1.035571 | 0.799739 |
| SL005158 | C-type lectin domain family 4 member M | DC-SIGNR | Q9H2X3 | 10332 | CLEC4M | 1.009996 | 0.799989 |
| SL005630 | Proteasome subunit alpha type-6 | PSA6 | P60900 | 5687 | PSMA6 | -1.013524 | 0.801786 |
| SL004752 | Protein FAM107A | DRR1 | O95990 | 11170 | FAM107A | -1.012218 | 0.803794 |
| SL006512 | Hepatocyte growth factor activator | HGFA | Q04756 | 3083 | HGFAC | 1.012764 | 0.808683 |
| SL001896 | Clusterin | Clusterin | P10909 | 1191 | CLU | 1.021874 | 0.808915 |
| SL004271 | Glial fibrillary acidic protein | GFAP | P14136 | 2670 | GFAP | -1.107093 | 0.811081 |
| SL010464 | Leucine-rich repeats and immunoglobulin-like domains protein 3 | LRIG3 | Q6UXM1 | 121227 | LRIG3 | -1.017703 | 0.812264 |
| SL009431 | Histidine triad nucleotide-binding protein 1 | HINT1 | P49773 | 3094 | HINT1 | 1.048189 | 0.812959 |
| SL004908 | Tropomyosin beta chain | Tropomyosin 2 | P07951 | 7169 | TPM2 | -1.011152 | 0.813435 |
| SL010505 | Megakaryocyte-associated tyrosine-protein kinase | MATK | P42679 | 4145 | MATK | -1.024172 | 0.815143 |
| SL003738 | T-lymphocyte activation antigen CD80 | B7 | P33681 | 941 | CD80 | 1.005243 | 0.815406 |
| SL010488 | Abelson tyrosine-protein kinase 2 | ABL2 | P42684 | 27 | ABL2 | 1.351722 | 0.817721 |
| SL006528 | Prolyl endopeptidase FAP | SEPR | Q12884 | 2191 | FAP | 1.042651 | 0.818999 |
| SL013240 | Adapter molecule crk | CRK | P46108 | 1398 | CRK | -1.009441 | 0.819956 |
| SL004066 | Group IIE secretory phospholipase A2 | GIIE | Q9NZK7 | 30814 | PLA2G2E | -1.002321 | 0.820398 |
| SL001729 | Granulocyte colony-stimulating factor | G-CSF | P09919 | 1440 | CSF3 | -1.037662 | 0.823243 |
| SL003322 | Vascular endothelial growth factor receptor 3 | VEGF sR3 | P35916 | 2324 | FLT4 | 1.030654 | 0.823824 |
| SL011071 | Platelet-derived growth factor C | PDGF-CC | Q9NRA1 | 56034 | PDGFC | 1.14768 | 0.825165 |
| SL008933 | Protein deglycase DJ-1 | PARK7 | Q99497 | 11315 | PARK7 | -1.013543 | 0.826189 |
| SL004118 | Tartrate-resistant acid phosphatase type 5 | TrATPase | P13686 | 54 | ACP5 | 1.001258 | 0.826721 |
| SL007747 | Serine/threonine-protein kinase TBK1 | TBK1 | Q9UHD2 | 29110 | TBK1 | -1.039992 | 0.826974 |
| SL003324 | Coagulation factor Xa | Coagulation Factor Xa | P00742 | 2159 | F10 | -1.034535 | 0.829062 |
| SL014130 | Carbohydrate sulfotransferase 6 | CHST6 | Q9GZX3 | 4166 | CHST6 | -1.009327 | 0.829523 |
| SL004339 | Fibroblast growth factor 5 | FGF-5 | P12034 | 2250 | FGF5 | -1.063197 | 0.829852 |
| SL012248 | Focal adhesion kinase 1 | FAK1 | Q05397 | 5747 | PTK2 | 1.01216 | 0.830542 |
| SL004829 | Protein S100-A7 | S100A7 | P31151 | 6278 | S100A7 | 1.559122 | 0.831299 |
| SL000312 | Complement C3 | C3 | P01024 | 718 | C3 | -1.034917 | 0.831578 |
| SL007385 | Interleukin-24 | IL24 | Q13007 | 11009 | IL24 | 1.005763 | 0.832719 |
| SL012469 | Glypican-5 | GPC5 | P78333 | 2262 | GPC5 | -1.072353 | 0.832931 |
| SL005205 | Natural cytotoxicity triggering receptor 3 | NKp30 | O14931 | 259197 | NCR3 | -1.031603 | 0.833126 |
| SL005189 | Interleukin-7 receptor subunit alpha | IL-7 Ra | P16871 | 3575 | IL7R | 1.046895 | 0.833332 |
| SL004329 | Growth/differentiation factor 5 | BMP-14 | P43026 | 8200 | GDF5 | -1.041556 | 0.833898 |
| SL010369 | Carbonic anhydrase 4 | Carbonic Anhydrase IV | P22748 | 762 | CA4 | 1.004859 | 0.834653 |
| SL009213 | Lysosomal protective protein | Cathepsin A | P10619 | 5476 | CTSA | 1.03791 | 0.835806 |
| SL003711 | Caspase-3 | Caspase-3 | P42574 | 836 | CASP3 | -1.065189 | 0.836039 |
| SL014896 | Ankyrin-2 | ANK2 | Q01484 | 287 | ANK2 | -1.213134 | 0.836266 |
| SL001728 | Platelet-derived growth factor receptor alpha | PDGFRA | P16234 | 5156 | PDGFRA | -2.196119 | 0.837197 |
| SL012168 | Protein lin-7 homolog B | LIN7B | Q9HAP6 | 64130 | LIN7B | 1.051197 | 0.838023 |
| SL000137 | Betacellulin | BTC | P35070 | 685 | BTC | 1.015505 | 0.838511 |
| SL007070 | Low-density lipoprotein receptor | LDLR | P01130 | 3949 | LDLR | -1.034428 | 0.839355 |
| SL010469 | Hemojuvelin | RGM-C | Q6ZVN8 | 148738 | HFE2 | -1.018382 | 0.841056 |
| SL000070 | Glypican-3 | Glypican 3 | P51654 | 2719 | GPC3 | -1.04021 | 0.841084 |
| SL005234 | Growth/differentiation factor 9 | GDF-9 | O60383 | 2661 | GDF9 | -1.014576 | 0.841971 |
| SL003646 | Tropomyosin alpha-4 chain | Tropomyosin 4 | P67936 | 7171 | TPM4 | -1.121233 | 0.842707 |
| SL005219 | Sialic acid-binding Ig-like lectin 9 | Siglec-9 | Q9Y336 | 27180 | SIGLEC9 | 1.02397 | 0.844331 |
| SL000310 | Complement C1r subcomponent | C1r | P00736 | 715 | C1R | 1.03374 | 0.84808 |
| SL003184 | Leptin receptor | sLeptin R | P48357 | 3953 | LEPR | 1.033271 | 0.848712 |
| SL013489 | Protein amnionless | AMNLS | Q9BXJ7 | 81693 | AMN | -1.022131 | 0.851822 |
| SL005195 | Lymphocyte activation gene 3 protein | LAG-3 | P18627 | 3902 | LAG3 | -1.060607 | 0.851827 |
| SL003332 | Matrix metalloproteinase-17 | MMP-17 | Q9ULZ9 | 4326 | MMP17 | 1.02103 | 0.852938 |
| SL004536 | Hepcidin | LEAP-1 | P81172 | 57817 | HAMP | 1.011634 | 0.854642 |
| SL002542 | GTPase KRas | K-ras | P01116 | 3845 | KRAS | -1.003343 | 0.856125 |
| SL000461 | Immunoglobulin E | IgE | P01854 | 3497 50802 3535 | IGHE IGK@ IGL@ | 3.047612 | 0.856373 |
| SL010499 | Tyrosine-protein kinase HCK | HCK | P08631 | 3055 | HCK | -1.046814 | 0.856645 |
| SL009768 | Chromobox protein homolog 5 | CBX5 | P45973 | 23468 | CBX5 | 1.041821 | 0.856914 |
| SL010510 | Serine/threonine-protein kinase PAK 7 | PAK7 | Q9P286 | 57144 | PAK7 | 1.016294 | 0.857201 |
| SL004811 | Transgelin-2 | Transgelin-2 | P37802 | 8407 | TAGLN2 | -1.022978 | 0.857252 |
| SL004145 | Tumor necrosis factor receptor superfamily member 14 | HVEM | Q92956 | 8764 | TNFRSF14 | -2.190476 | 0.857345 |
| SL005199 | MHC class I polypeptide-related sequence A | MICA | Q29983 | 4276 | MICA | 1.120955 | 0.859028 |
| SL017289 | PolyUbiquitin K48-linked | PolyUbiquitin K48 | P0CG47 | 7314 | UBB | 1.19555 | 0.859562 |
| SL004352 | Interleukin-17F | IL-17F | Q96PD4 | 112744 | IL17F | -1.03933 | 0.860881 |
| SL014268 | OX-2 membrane glycoprotein | OX2G | P41217 | 4345 | CD200 | 1.013532 | 0.861153 |
| SL004475 | Interferon alpha/beta receptor 1 | IFN-a/b R1 | P17181 | 3454 | IFNAR1 | 1.032971 | 0.862394 |
| SL005168 | Growth hormone receptor | Growth hormone receptor | P10912 | 2690 | GHR | 1.028007 | 0.863164 |
| SL000344 | Cathepsin D | Cathepsin D | P07339 | 1509 | CTSD | -1.144353 | 0.86578 |
| SL000088 | Transforming growth factor beta-2 | TGF-b2 | P61812 | 7042 | TGFB2 | -1.02396 | 0.866925 |
| SL000437 | Haptoglobin | Haptoglobin, Mixed Type | P00738 | 3240 | HP | -1.029013 | 0.868762 |
| SL004343 | Fms-related tyrosine kinase 3 ligand | Flt3 ligand | P49771 | 2323 | FLT3LG | -1.042476 | 0.868814 |
| SL014009 | Hepatoma-derived growth factor-related protein 2 | HDGR2 | Q7Z4V5 | 84717 | HDGFRP2 | 1.124329 | 0.869877 |
| SL002539 | Tumor necrosis factor receptor superfamily member 11B | OPG | O00300 | 4982 | TNFRSF11B | 1.014261 | 0.871028 |
| SL004296 | Adenylate kinase isoenzyme 1 | Myokinase, human | P00568 | 203 | AK1 | 1.207582 | 0.872424 |
| SL010513 | cAMP-dependent protein kinase catalytic subunit alpha | PRKACA | P17612 | 5566 | PRKACA | -1.044566 | 0.872683 |
| SL011049 | Mannan-binding lectin serine protease 1 | MASP3 | P48740 | 5648 | MASP1 | 1.015185 | 0.873861 |
| SL003800 | Kallikrein-4 | Kallikrein 4 | Q9Y5K2 | 9622 | KLK4 | -1.001661 | 0.874086 |
| SL017328 | Myosin-binding protein C, slow-type | MYPC1 | Q00872 | 4604 | MYBPC1 | 1.090675 | 0.877002 |
| SL010461 | High affinity immunoglobulin gamma Fc receptor I | FCGR1 | P12314 | 2209 | FCGR1A | 1.032227 | 0.877326 |
| SL003308 | Interleukin-4 receptor subunit alpha | IL-4 sR | P24394 | 3566 | IL4R | -1.064934 | 0.879127 |
| SL011769 | Acidic leucine-rich nuclear phosphoprotein 32 family member B | AN32B | Q92688 | 10541 | ANP32B | -1.007991 | 0.879785 |
| SL000321 | Complement C5b-C6 complex | C5b, 6 Complex | P01031,P13671 | 727 729 | C5 C6 | -1.040509 | 0.879858 |
| SL003189 | C-C motif chemokine 19 | MIP-3b | Q99731 | 6363 | CCL19 | -1.063698 | 0.880676 |
| SL006803 | Angiopoietin-related protein 3 | ANGL3 | Q9Y5C1 | 27329 | ANGPTL3 | -1.022709 | 0.881198 |
| SL003974 | Activated Protein C | Activated Protein C | P04070 | 5624 | PROC | -1.512861 | 0.883196 |
| SL003685 | Nicotinamide phosphoribosyltransferase | PBEF | P43490 | 10135 | NAMPT | -1.097244 | 0.884281 |
| SL000508 | Lymphotoxin alpha2:beta1 | Lymphotoxin a2/b1 | P01374, Q06643 | 4049 4050 | LTA LTB | 1.008511 | 0.885421 |
| SL002621 | Midkine | Midkine | P21741 | 4192 | MDK | 1.038726 | 0.886302 |
| SL000584 | Transforming growth factor beta-1 | TGF-b1 | P01137 | 7040 | TGFB1 | -1.016672 | 0.889089 |
| SL000479 | Interleukin-3 | IL-3 | P08700 | 3562 | IL3 | 1.058407 | 0.889693 |
| SL000342 | Catalase | Catalase | P04040 | 847 | CAT | 1.040978 | 0.891789 |
| SL000049 | Vitamin K-dependent protein S | Protein S | P07225 | 5627 | PROS1 | 1.008579 | 0.89336 |
| SL000250 | Alpha-2-antiplasmin | a2-Antiplasmin | P08697 | 5345 | SERPINF2 | 1.001866 | 0.893395 |
| SL016969 | Cysteine and glycine-rich protein 3 | CSRP3 | P50461 | 8048 | CSRP3 | -1.007887 | 0.893455 |
| SL004015 | Carboxypeptidase B2 | TAFI | Q96IY4 | 1361 | CPB2 | 1.021072 | 0.89469 |
| SL002517 | Tumor necrosis factor | TNF-a | P01375 | 7124 | TNF | 1.017262 | 0.895481 |
| SL000019 | Apolipoprotein A-I | Apo A-I | P02647 | 335 | APOA1 | 1.011415 | 0.89662 |
| SL006914 | Glucocorticoid receptor | Glucocorticoid receptor | P04150 | 2908 | NR3C1 | -1.178408 | 0.900251 |
| SL004856 | Desmoglein-1 | Desmoglein-1 | Q02413 | 1828 | DSG1 | 1.002074 | 0.903873 |
| SL006921 | [Pyruvate dehydrogenase (acetyl-transferring)] kinase isozyme 1, mitochondrial | PDK1 | Q15118 | 5163 | PDK1 | 1.153837 | 0.905558 |
| SL005437 | Matrix extracellular phosphoglycoprotein | MEPE | Q9NQ76 | 56955 | MEPE | -1.056035 | 0.906186 |
| SL001716 | Interleukin-12 | IL-12 | P29459, P29460 | 3592 3593 | IL12A IL12B | -1.00551 | 0.906803 |
| SL005258 | Serine/threonine-protein kinase PLK1 | PLK-1 | P53350 | 5347 | PLK1 | -1.076717 | 0.906883 |
| SL004914 | Inorganic pyrophosphatase | PPase | Q15181 | 5464 | PPA1 | 1.151681 | 0.907718 |
| SL004654 | Granzyme H | Granzyme H | P20718 | 2999 | GZMH | -1.01791 | 0.911031 |
| SL000158 | Glutamate carboxypeptidase 2 | PSMA | Q04609 | 2346 | FOLH1 | -1.018441 | 0.911199 |
| SL007266 | 26S proteasome non-ATPase regulatory subunit 7 | PSD7 | P51665 | 5713 | PSMD7 | -1.008429 | 0.911213 |
| SL004353 | Interleukin-17D | IL-17D | Q8TAD2 | 53342 | IL17D | 1.085041 | 0.91275 |
| SL000616 | Vitronectin | Vitronectin | P04004 | 7448 | VTN | -1.010269 | 0.913706 |
| SL000545 | Plasma kallikrein | Prekallikrein | P03952 | 3818 | KLKB1 | -1.005082 | 0.914505 |
| SL000603 | Trypsin-1 | Trypsin | P07477 | 5644 | PRSS1 | -1.079688 | 0.916815 |
| SL012395 | Netrin-1 | NET1 | O95631 | 9423 | NTN1 | -1.083876 | 0.917867 |
| SL000254 | Serum albumin | Albumin | P02768 | 213 | ALB | 1.038095 | 0.918443 |
| SL011499 | Pancreatic hormone | PH | P01298 | 5539 | PPY | -1.131118 | 0.92009 |
| SL002640 | Placenta growth factor | PlGF | P49763 | 5228 | PGF | -1.004651 | 0.923302 |
| SL006610 | A disintegrin and metalloproteinase with thrombospondin motifs 13 | ATS13 | Q76LX8 | 11093 | ADAMTS13 | 1.00098 | 0.924018 |
| SL013754 | Ras GTPase-activating protein 1 | RASA1 | P20936 | 5921 | RASA1 | 1.000678 | 0.924086 |
| SL004248 | Glial cell line-derived neurotrophic factor | GDNF | P39905 | 2668 | GDNF | 1.010285 | 0.925362 |
| SL000268 | Angiostatin | Angiostatin | P00747 | 5340 | PLG | -1.005758 | 0.926264 |
| SL010449 | Carbonic anhydrase-related protein 10 | Carbonic Anhydrase X | Q9NS85 | 56934 | CA10 | 1.008073 | 0.927678 |
| SL005392 | Arylsulfatase A | Arylsulfatase A | P15289 | 410 | ARSA | 1.006492 | 0.927735 |
| SL000573 | Serum amyloid P-component | SAP | P02743 | 325 | APCS | 1.003217 | 0.929823 |
| SL005361 | Apolipoprotein D | Apo D | P05090 | 347 | APOD | 1.007395 | 0.930696 |
| SL000021 | Insulin | Insulin | P01308 | 3630 | INS | -1.027836 | 0.930965 |
| SL004795 | 3-hydroxyacyl-CoA dehydrogenase type-2 | ERAB | Q99714 | 3028 | HSD17B10 | -1.063905 | 0.931333 |
| SL000074 | Cellular tumor antigen p53 | p53 | P04637 | 7157 | TP53 | 1.003902 | 0.931383 |
| SL000406 | Eotaxin | Eotaxin | P51671 | 6356 | CCL11 | -1.012888 | 0.931808 |
| SL005207 | Natural cytotoxicity triggering receptor 1 | NKp46 | O76036 | 9437 | NCR1 | -1.103754 | 0.93744 |
| SL000467 | Immunoglobulin G | IgG | P01857 | 3500 3501 3502 3503 50802 3535 | IGHG1 IGHG2 IGHG3 IGHG4 IGK@ IGL@ | -1.001141 | 0.938649 |
| SL003849 | Fibroblast growth factor 9 | FGF9 | P31371 | 2254 | FGF9 | -1.020391 | 0.939678 |
| SL001713 | Interleukin-17A | IL-17 | Q16552 | 3605 | IL17A | -1.000066 | 0.941031 |
| SL000039 | Interleukin-8 | IL-8 | P10145 | 3576 | CXCL8 | -1.112449 | 0.941603 |
| HCE000342 | HCE000342 | HCE000342 | HCE000342 | HCE000342 | HCE000342 | -1.005026 | 0.941773 |
| SL004298 | Granzyme A | granzyme A | P12544 | 3001 | GZMA | -1.029385 | 0.941986 |
| SL008190 | Spectrin alpha chain, non-erythrocytic 1 | SPTA2 | Q13813 | 6709 | SPTAN1 | 1.02787 | 0.945946 |
| SL005694 | Peroxiredoxin-6 | Peroxiredoxin-6 | P30041 | 9588 | PRDX6 | 1.129571 | 0.947279 |
| SL007108 | Interferon regulatory factor 1 | IRF1 | P10914 | 3659 | IRF1 | -1.167609 | 0.950034 |
| SL007136 | CCAAT/enhancer-binding protein beta | CEBPB | P17676 | 1051 | CEBPB | -1.162081 | 0.950347 |
| SL007049 | Cystatin-F | CYTF | O76096 | 8530 | CST7 | -1.345449 | 0.950764 |
| SL004932 | Peroxiredoxin-5, mitochondrial | Peroxiredoxin-5 | P30044 | 25824 | PRDX5 | 1.028891 | 0.95117 |
| SL009790 | Small nuclear ribonucleoprotein F | RUXF | P62306 | 6636 | SNRPF | 1.004275 | 0.951384 |
| SL004625 | A disintegrin and metalloproteinase with thrombospondin motifs 4 | ADAMTS-4 | O75173 | 9507 | ADAMTS4 | -1.010545 | 0.952607 |
| SL001802 | Interferon gamma | IFN-g | P01579 | 3458 | IFNG | -1.015273 | 0.952701 |
| SL000622 | Coagulation Factor V | Coagulation Factor V | P12259 | 2153 | F5 | -1.013564 | 0.953039 |
| SL018971 | E3 ubiquitin-protein ligase ZNRF3 | ZNRF3 | Q9ULT6 | 84133 | ZNRF3 | -1.007726 | 0.953334 |
| SL008099 | Macrophage-capping protein | CAPG | P40121 | 822 | CAPG | -1.066622 | 0.954022 |
| SL003710 | Caspase-2 | Caspase-2 | P42575 | 835 | CASP2 | 1.027553 | 0.954144 |
| HCE004152 | HCE004152 | HCE004152 | HCE004152 | HCE004152 | HCE004152 | 1.047981 | 0.955094 |
| SL003916 | Kallikrein-12 | kallikrein 12 | Q9UKR0 | 43849 | KLK12 | 1.01699 | 0.955675 |
| SL002655 | Connective tissue growth factor | CTGF | P29279 | 1490 | CTGF | -1.007573 | 0.956312 |
| SL002524 | T-cell surface glycoprotein CD4 | sCD4 | P01730 | 920 | CD4 | -1.042254 | 0.957397 |
| SL004269 | Appetite-regulating hormone | ghrelin | Q9UBU3 | 51738 | GHRL | 1.094061 | 0.957484 |
| SL005215 | Myeloid cell surface antigen CD33 | Siglec-3 | P20138 | 945 | CD33 | 1.055951 | 0.958329 |
| SL004737 | Tropomyosin alpha-1 chain | Tropomyosin 1 alpha chain | P09493 | 7168 | TPM1 | -1.004695 | 0.958927 |
| SL004844 | Ephrin type-A receptor 5 | EphA5 | P54756 | 2044 | EPHA5 | -1.002456 | 0.962898 |
| SL003310 | Vascular endothelial growth factor A, isoform 121 | VEGF121 | P15692 | 7422 | VEGFA | -1.062757 | 0.963332 |
| SL003863 | Kallikrein-5 | kallikrein 5 | Q9Y337 | 25818 | KLK5 | -1.225183 | 0.964696 |
| SL004364 | Tumor necrosis factor receptor superfamily member 13B | TACI | O14836 | 23495 | TNFRSF13B | 1.004871 | 0.975404 |
| SL010504 | MAP kinase-activated protein kinase 5 | MAPK5 | Q8IW41 | 8550 | MAPKAPK5 | 1.223629 | 0.978442 |
| SL004670 | Artemin | Artemin | Q5T4W7 | 9048 | ARTN | 1.030888 | 0.980581 |
| SL011509 | Peptide YY | PYY | P10082 | 5697 | PYY | 1.003715 | 0.982398 |
| SL005221 | Scavenger receptor class F member 1 | SREC-I | Q14162 | 8578 | SCARF1 | -1.008818 | 0.984352 |
| SL010613 | Interleukin-17 receptor D | IL-17 RD | Q8NFM7 | 54756 | IL17RD | 1.001241 | 0.985349 |
| SL004157 | Tumor necrosis factor receptor superfamily member 10B | TRAIL R2 | O14763 | 8795 | TNFRSF10B | 1.007081 | 0.985976 |
| SL004126 | Tumor necrosis factor receptor superfamily member 9 | 4-1BB | Q07011 | 3604 | TNFRSF9 | -1.618109 | 0.986119 |
| SL007237 | Dual specificity mitogen-activated protein kinase kinase 4 | MP2K4 | P45985 | 6416 | MAP2K4 | -1.002234 | 0.987656 |
| SL000420 | Ferritin | Ferritin | P02794 P02792 | 2495 2512 | FTH1 FTL | -1.112924 | 0.988666 |
| SL000542 | Integrin alpha-IIb: beta-3 complex | gpIIbIIIa | P08514 P05106 | 3674 3690 | ITGA2B ITGB3 | 1.00399 | 0.98895 |
| SL000566 | Retinol-binding protein 4 | RBP | P02753 | 5950 | RBP4 | 1.021913 | 0.98978 |
| SL004342 | Fibroblast growth factor 8 isoform B | FGF-8B | P55075 | 2253 | FGF8 | 1.014985 | 0.990181 |
| SL002644 | Epidermal growth factor receptor | ERBB1 | P00533 | 1956 | EGFR | 1.004822 | 0.99209 |
| SL018891 | Down syndrome cell adhesion molecule | DSCAM | O60469 | 1826 | DSCAM | -1.025748 | 0.993714 |
| SL007024 | Gremlin-1 | GREM1 | O60565 | 26585 | GREM1 | 1.011927 | 0.994808 |
| SL003704 | BH3-interacting domain death agonist | BID | P55957 | 637 | BID | -2.013764 | 0.998466 |

**Table S3b.** Differential levels of the SOMAscan-screened proteins according to delirium effect at POD2.

| **SomaId** | **TargetFullName** | **Target** | **UniProt** | **EntrezGeneID** | **EntrezGeneSymbol** | **tFC** | **p-val** |
| --- | --- | --- | --- | --- | --- | --- | --- |
| SL001774 | Fatty acid-binding protein, heart | FABP | P05413 | 2170 | FABP3 | 1.932971 | 0.000248 |
| SL000084 | Epidermal growth factor | EGF | P01133 | 1950 | EGF | -1.286166 | 0.000281 |
| SL005233 | Tumor necrosis factor receptor superfamily member 27 | XEDAR | Q9HAV5 | 60401 | EDA2R | 1.25966 | 0.000942 |
| SL021043 | Growth/differentiation factor 11/8 | GDF-11/8 | O95390 O14793 | NaN | GDF11 MSTN | -1.322293 | 0.00114 |
| SL002093 | Histone H2A.z | Histone H2A.z | P0C0S5 | 3015 | H2AFZ | 1.611476 | 0.002358 |
| SL000322 | Complement component C6 | C6 | P13671 | 729 | C6 | -1.231442 | 0.002585 |
| SL014092 | Cell adhesion molecule-related/down-regulated by oncogenes | CDON | Q4KMG0 | 50937 | CDON | -1.301259 | 0.003227 |
| SL005797 | Polymeric immunoglobulin receptor | PIGR | P01833 | 5284 | PIGR | 1.704651 | 0.003321 |
| SL018946 | Ubiquitin-conjugating enzyme E2 G2 | UB2G2 | P60604 | 7327 | UBE2G2 | -1.298644 | 0.004497 |
| SL000591 | Metalloproteinase inhibitor 1 | TIMP-1 | P01033 | 7076 | TIMP1 | 1.324939 | 0.005171 |
| SL000563 | C-C motif chemokine 5 | RANTES | P13501 | 6352 | CCL5 | -1.731573 | 0.005527 |
| SL005403 | Vesicular integral-membrane protein VIP36 | Lectin, mannose-binding 2 | Q12907 | 10960 | LMAN2 | -1.250802 | 0.006754 |
| SL003755 | Induced myeloid leukemia cell differentiation protein Mcl-1 | Mcl-1 | Q07820 | 4170 | MCL1 | 1.397277 | 0.007155 |
| SL001995 | Angiopoietin-1 | Angiopoietin-1 | Q15389 | 284 | ANGPT1 | -1.277224 | 0.007291 |
| SL004360 | Neurotrophin-4 | Neurotrophin-5 | P34130 | 4909 | NTF4 | 1.260345 | 0.007603 |
| SL001797 | Kallikrein-6 | Kallikrein 6 | Q92876 | 5653 | KLK6 | -1.149392 | 0.008381 |
| SL003172 | C-X-C motif chemokine 6 | GCP-2 | P80162 | 6372 | CXCL6 | -1.376466 | 0.008393 |
| SL014009 | Hepatoma-derived growth factor-related protein 2 | HDGR2 | Q7Z4V5 | 84717 | HDGFRP2 | 1.274322 | 0.008943 |
| SL007828 | Neuregulin-4 | NRG4 | Q8WWG1 | 145957 | NRG4 | 1.724248 | 0.009622 |
| SL007547 | Hepatitis A virus cellular receptor 2 | TIMD3 | Q8TDQ0 | 84868 | HAVCR2 | 1.255009 | 0.009782 |
| SL005263 | alpha-2-macroglobulin receptor-associated protein | RAP | P30533 | 4043 | LRPAP1 | 1.292035 | 0.011491 |
| SL007181 | Non-receptor tyrosine-protein kinase TYK2 | TYK2 | P29597 | 7297 | TYK2 | -1.299183 | 0.011763 |
| SL000483 | Interleukin-7 | IL-7 | P13232 | 3574 | IL7 | 2.072644 | 0.011857 |
| SL004131 | T-lymphocyte activation antigen CD86 | B7-2 | P42081 | 942 | CD86 | -1.276762 | 0.01407 |
| SL010375 | Neutral ceramidase | ASAH2 | Q9NR71 | 56624 | ASAH2 | -1.195455 | 0.01438 |
| SL004899 | Heat shock cognate 71 kDa protein | HSP70 protein 8 | P11142 | 3312 | HSPA8 | 1.217293 | 0.014754 |
| SL004868 | Carbonic anhydrase 7 | Carbonic anhydrase VII | P43166 | 766 | CA7 | 1.165345 | 0.014791 |
| SL004645 | Kunitz-type protease inhibitor 1 | HAI-1 | O43278 | 6692 | SPINT1 | -1.3392 | 0.016703 |
| SL004357 | Growth/differentiation factor 8 | Myostatin | O14793 | 2660 | MSTN | -1.338952 | 0.017042 |
| SL017128 | Leucine-rich repeat serine/threonine-protein kinase 2 | LRRK2 | Q5S007 | NaN | LRRK2 | -1.16278 | 0.017115 |
| SL000306 | Brain natriuretic peptide 32 | BNP-32 | P16860 | 4879 | NPPB | 2.299345 | 0.017327 |
| SL004016 | C-X-C motif chemokine 16 | CXCL16, soluble | Q9H2A7 | 58191 | CXCL16 | 1.248126 | 0.018201 |
| SL013488 | C-type lectin domain family 1 member B | CLC1B | Q9P126 | 51266 | CLEC1B | -1.35881 | 0.018274 |
| SL014069 | Proprotein convertase subtilisin/kexin type 7 | PCSK7 | Q16549 | 9159 | PCSK7 | -1.198256 | 0.019875 |
| SL010469 | Hemojuvelin | RGM-C | Q6ZVN8 | 148738 | HFE2 | -1.170389 | 0.020596 |
| SL000320 | C5a anaphylatoxin | C5a | P01031 | 727 | C5 | -1.274766 | 0.020728 |
| SL000537 | Platelet-derived growth factor subunit B | PDGF-BB | P01127 | 5155 | PDGFB | -1.746779 | 0.021844 |
| SL004690 | WNT1-inducible-signaling pathway protein 3 | WISP-3 | O95389 | 8838 | WISP3 | 1.319664 | 0.021874 |
| SL001802 | Interferon gamma | IFN-g | P01579 | 3458 | IFNG | -1.167412 | 0.022743 |
| SL001761 | Troponin I, cardiac muscle | Troponin I | P19429 | 7137 | TNNI3 | 1.810825 | 0.022794 |
| SL005491 | Opioid-binding protein/cell adhesion molecule | OBCAM | Q14982 | 4978 | OPCML | -1.138131 | 0.023192 |
| SL013988 | Carbohydrate sulfotransferase 2 | CHST2 | Q9Y4C5 | 9435 | CHST2 | 1.189242 | 0.023213 |
| SL001992 | Tumor necrosis factor receptor superfamily member 1A | TNF sR-I | P19438 | 7132 | TNFRSF1A | 1.307527 | 0.024583 |
| SL003685 | Nicotinamide phosphoribosyltransferase | PBEF | P43490 | 10135 | NAMPT | 1.321171 | 0.026353 |
| SL001815 | Superoxide dismutase [Mn], mitochondrial | Mn SOD | P04179 | 6648 | SOD2 | -1.201564 | 0.026421 |
| SL000064 | Kallikrein-7 | Kallikrein 7 | P49862 | 5650 | KLK7 | -1.218786 | 0.027286 |
| SL016129 | Protein FAM107B | FAM107B | Q9H098 | 83641 | FAM107B | -1.167311 | 0.027386 |
| SL003753 | Dynein light chain 1, cytoplasmic | DLC8 | P63167 | 8655 | DYNLL1 | 1.406757 | 0.028353 |
| SL005224 | Thrombopoietin Receptor | Thrombopoietin Receptor | P40238 | 4352 | MPL | -1.205509 | 0.02855 |
| SL014028 | Ectonucleoside triphosphate diphosphohydrolase 5 | ENTP5 | O75356 | 957 | ENTPD5 | -1.200772 | 0.02968 |
| SL004159 | Tumor necrosis factor receptor superfamily member 10D | TRAIL R4 | Q9UBN6 | 8793 | TNFRSF10D | 1.283235 | 0.029859 |
| SL005173 | Interleukin-10 receptor subunit alpha | IL-10 Ra | Q13651 | 3587 | IL10RA | -1.166234 | 0.030264 |
| SL012457 | Neurexophilin-1 | NXPH1 | P58417 | 30010 | NXPH1 | -1.217451 | 0.030622 |
| SL009341 | Basigin | BASI | P35613 | 682 | BSG | 1.104215 | 0.030786 |
| SL014111 | Kin of IRRE-like protein 3 | KIRR3 | Q8IZU9 | 84623 | KIRREL3 | -1.143901 | 0.031601 |
| SL000603 | Trypsin-1 | Trypsin | P07477 | 5644 | PRSS1 | -1.512756 | 0.031946 |
| SL004939 | Tyrosine-protein phosphatase non-receptor type 1 | PTP-1B | P18031 | 5770 | PTPN1 | -1.225808 | 0.032358 |
| SL003179 | Integrin alpha-I: beta-1 complex | Integrin a1b1 | P56199, P05556 | 3672 3688 | ITGA1 ITGB1 | 1.612867 | 0.032933 |
| SL005087 | Insulin-like growth factor-binding protein 7 | IGFBP-7 | Q16270 | 3490 | IGFBP7 | 1.180024 | 0.033342 |
| SL004865 | Cadherin-6 | Cadherin-6 | P55285 | 1004 | CDH6 | -1.16682 | 0.033592 |
| SL004771 | Aurora kinase A | Aurora kinase A | O14965 | 6790 | AURKA | 1.350579 | 0.033772 |
| SL010288 | Carbonic anhydrase 6 | Carbonic anhydrase 6 | P23280 | 765 | CA6 | -1.541549 | 0.033828 |
| SL000087 | Interleukin-6 | IL-6 | P05231 | 3569 | IL6 | 1.342734 | 0.035631 |
| SL004837 | Inhibin beta A chain:Inhibin beta B chain heterodimer | Activin AB | P08476 P09529 | 3624 3625 | INHBA INHBB | 1.709308 | 0.035771 |
| SL000268 | Angiostatin | Angiostatin | P00747 | 5340 | PLG | -1.198572 | 0.035924 |
| SL000668 | Platelet glycoprotein 4 | CD36 ANTIGEN | P16671 | 948 | CD36 | 1.23342 | 0.035976 |
| SL008639 | Iduronate 2-sulfatase | IDS | P22304 | 3423 | IDS | -1.274182 | 0.039042 |
| SL006372 | Tyrosine-protein kinase Yes | YES | P07947 | 7525 | YES1 | 1.226503 | 0.039633 |
| SL000535 | Platelet-derived growth factor subunit A | PDGF-AA | P04085 | 5154 | PDGFA | -1.443885 | 0.040179 |
| SL019019 | PILR alpha-associated neural protein | PIANP | Q8IYJ0 | 196500 | PIANP | 1.324582 | 0.040594 |
| SL002756 | Heterogeneous nuclear ribonucleoprotein K | hnRNP K | P61978 | 3190 | HNRNPK | 1.695806 | 0.04148 |
| SL004860 | Serine protease HTRA2, mitochondrial | HTRA2 | O43464 | 27429 | HTRA2 | 1.282187 | 0.041508 |
| SL005235 | Homeobox protein NANOG | NANOG | Q9H9S0 | 79923 | NANOG | 1.12655 | 0.042632 |
| SL001996 | Angiopoietin-2 | Angiopoietin-2 | O15123 | 285 | ANGPT2 | 1.335408 | 0.043441 |
| SL010498 | Ephrin type-A receptor 3 | EPHA3 | P29320 | 2042 | EPHA3 | 1.087012 | 0.046108 |
| SL006528 | Prolyl endopeptidase FAP | SEPR | Q12884 | 2191 | FAP | -1.164875 | 0.046206 |
| SL000498 | Leptin | Leptin | P41159 | 3952 | LEP | 1.543157 | 0.04657 |
| SL003717 | Caspase-10 | Caspase-10 | Q92851 | 843 | CASP10 | -1.100382 | 0.046619 |
| SL014983 | Histone H2B type 2-E | H2B2E | Q16778 | 8349 | HIST2H2BE | 1.390071 | 0.047177 |
| SL014113 | Neuroligin-4, X-linked | NLGNX | Q8N0W4 | 57502 | NLGN4X | 1.301284 | 0.047279 |
| SL000459 | Immunoglobulin A | IgA | P01876 P01877 | 3493 3494 | IGHA1 IGHA2 | 1.232909 | 0.047431 |
| SL010458 | Endothelial cell-specific molecule 1 | Endocan | Q9NQ30 | 11082 | ESM1 | 1.25191 | 0.047447 |
| SL000546 | Prolactin | PRL | P01236 | 5617 | PRL | 1.340062 | 0.047912 |
| SL004363 | Stem cell growth factor-alpha | SCGF-alpha | Q9Y240 | 6320 | CLEC11A | 1.407138 | 0.047957 |
| SL014094 | Adhesion G-protein coupled receptor G5 | GP114 | Q8IZF4 | 221188 | ADGRG5 | 1.181261 | 0.049166 |
| SL004625 | A disintegrin and metalloproteinase with thrombospondin motifs 4 | ADAMTS-4 | O75173 | 9507 | ADAMTS4 | 1.116222 | 0.051288 |
| SL004588 | Interleukin-1 Receptor accessory protein | IL-1 R AcP | Q9NPH3 | 3556 | IL1RAP | -1.26244 | 0.05179 |
| SL003043 | Metalloproteinase inhibitor 3 | TIMP-3 | P35625 | 7078 | TIMP3 | -1.31196 | 0.052222 |
| SL000309 | Complement C1q subcomponent | C1q | P02745 P02746 P02747 | 712 713 714 | C1QA C1QB C1QC | 1.175275 | 0.052421 |
| SL004160 | BDNF/NT-3 growth factors receptor | TrkB | Q16620 | 4915 | NTRK2 | -1.244329 | 0.052661 |
| SL010378 | Proto-oncogene tyrosine-protein kinase receptor Ret | RET | P07949 | 5979 | RET | -1.20048 | 0.052672 |
| SL003647 | Annexin A6 | annexin VI | P08133 | 309 | ANXA6 | -1.497966 | 0.053078 |
| SL000451 | Heat shock 70 kDa protein 1A | HSP 70 | P0DMV8 | 3303 | HSPA1A | 1.33049 | 0.053444 |
| SL008760 | SLIT and NTRK-like protein 1 | SLIK1 | Q96PX8 | 114798 | SLITRK1 | 1.221413 | 0.054107 |
| SL006913 | Tyrosine-protein kinase Fyn | FYN | P06241 | 2534 | FYN | -1.763179 | 0.055102 |
| SL002655 | Connective tissue growth factor | CTGF | P29279 | 1490 | CTGF | -1.112539 | 0.055137 |
| SL000493 | L-lactate dehydrogenase B chain | LDH-H 1 | P07195 | 3945 | LDHB | 1.152334 | 0.055906 |
| SL013969 | Kynureninase | KYNU | Q16719 | 8942 | KYNU | 1.305762 | 0.055964 |
| SL007195 | CD70 antigen | CD70 | P32970 | 970 | CD70 | 1.095079 | 0.056118 |
| SL008094 | Phosphoglucomutase-1 | PGM1 | P36871 | 5236 | PGM1 | 1.321072 | 0.05846 |
| SL005789 | Stanniocalcin-1 | Stanniocalcin-1 | P52823 | 6781 | STC1 | 1.233638 | 0.060238 |
| SL003994 | Bone morphogenetic protein 1 | BMP-1 | P13497 | 649 | BMP1 | -1.186488 | 0.061172 |
| SL008382 | Cystatin-D | CYTD | P28325 | 1473 | CST5 | 1.586015 | 0.061773 |
| SL000125 | Interleukin-1 alpha | IL-1a | P01583 | 3552 | IL1A | -1.200938 | 0.062741 |
| SL001777 | Cystatin-C | Cystatin C | P01034 | 1471 | CST3 | 1.241986 | 0.062864 |
| SL000516 | C-C motif chemokine 7 | MCP-3 | P80098 | 6354 | CCL7 | 1.164756 | 0.063379 |
| SL000617 | Alanine aminotransferase 1 | ALT | P24298 | 2875 | GPT | 1.688531 | 0.063959 |
| SL010456 | Cystatin-SN | CYTN | P01037 | 1469 | CST1 | 2.225795 | 0.06433 |
| SL007206 | Thrombospondin-2 | TSP2 | P35442 | 7058 | THBS2 | 1.656098 | 0.064509 |
| SL003542 | Histone-lysine N-methyltransferase EHMT2 | NG36 | Q96KQ7 | 10919 | EHMT2 | -1.165834 | 0.064923 |
| SL000051 | C-reactive protein | CRP | P02741 | 1401 | CRP | -1.095873 | 0.067128 |
| SL002654 | Ephrin type-A receptor 2 | Epithelial cell kinase | P29317 | 1969 | EPHA2 | 1.226232 | 0.067623 |
| SL004362 | Stem Cell Growth Factor-beta | SCGF-beta | Q9Y240 | 6320 | CLEC11A | 1.359999 | 0.067766 |
| SL006131 | Fatty acid-binding protein, liver | FABPL | P07148 | 2168 | FABP1 | 1.800992 | 0.06941 |
| SL003173 | Growth-regulated alpha protein | Gro-a | P09341 | 2919 | CXCL1 | 1.512725 | 0.069534 |
| SL002506 | Urokinase plasminogen activator surface receptor | suPAR | Q03405 | 5329 | PLAUR | 1.172726 | 0.070316 |
| SL003340 | Chitinase-3-like protein 1 | YKL-40 | P36222 | 1116 | CHI3L1 | 1.483803 | 0.070355 |
| SL002706 | Low affinity immunoglobulin epsilon Fc receptor | CD23 | P06734 | 2208 | FCER2 | 1.459743 | 0.070778 |
| SL000398 | Cytochrome P450 3A4 | Cytochrome P450 3A4 | P08684 | 1576 | CYP3A4 | -1.190366 | 0.071127 |
| SL009431 | Histidine triad nucleotide-binding protein 1 | HINT1 | P49773 | 3094 | HINT1 | 1.137551 | 0.071142 |
| SL001938 | Inhibin beta A chain | Activin A | P08476 | 3624 | INHBA | 1.229123 | 0.07194 |
| SL005193 | Junctional adhesion molecule B | JAM-B | P57087 | 58494 | JAM2 | 1.112565 | 0.072033 |
| SL001737 | 14-3-3 protein sigma | STRATIFIN | P31947 | 2810 | SFN | 1.154376 | 0.072362 |
| SL004120 | Discoidin domain-containing receptor 2 | Discoidin domain receptor 2 | Q16832 | 4921 | DDR2 | 1.146485 | 0.073575 |
| SL001997 | Interleukin-1 receptor type 1 | IL-1 sRI | P14778 | 3554 | IL1R1 | -1.269947 | 0.074439 |
| SL014188 | Natural killer cell receptor 2B4 | CD244 | Q9BZW8 | 51744 | CD244 | 1.368233 | 0.074746 |
| SL008008 | Arginase-1 | ARGI1 | P05089 | 383 | ARG1 | 1.644117 | 0.07478 |
| SL014029 | Sphingosine kinase 2 | SPHK2 | Q9NRA0 | 56848 | SPHK2 | -1.102216 | 0.075022 |
| SL005361 | Apolipoprotein D | Apo D | P05090 | 347 | APOD | -1.157922 | 0.075185 |
| SL008810 | Neuronal growth regulator 1 | NEGR1 | Q7Z3B1 | 257194 | NEGR1 | -1.14659 | 0.075351 |
| HCE000483 | HCE000483 | HCE000483 | HCE000483 | HCE000483 | HCE000483 | -1.139038 | 0.075628 |
| SL008421 | A disintegrin and metalloproteinase with thrombospondin motifs 1 | ATS1 | Q9UHI8 | 9510 | ADAMTS1 | 1.097228 | 0.076518 |
| SL003191 | Neutrophil-activating peptide 2 | NAP-2 | P02775 | 5473 | PPBP | -1.335043 | 0.076714 |
| SL005201 | Muellerian-inhibiting factor | MIS | P03971 | 268 | AMH | -1.175242 | 0.07677 |
| SL011100 | CD226 antigen | CD226 | Q15762 | 10666 | CD226 | 1.196451 | 0.076942 |
| SL001896 | Clusterin | Clusterin | P10909 | 1191 | CLU | -1.115812 | 0.078723 |
| SL002646 | Matrix metalloproteinase-14 | MMP-14 | P50281 | 4323 | MMP14 | -1.447838 | 0.079684 |
| SL004579 | Macrophage mannose receptor 1 | Macrophage mannose receptor | P22897 | 4360 | MRC1 | 1.160964 | 0.080368 |
| SL003522 | Endoplasmic reticulum resident protein 29 | ERP29 | P30040 | 10961 | ERP29 | -1.203125 | 0.081157 |
| SL000524 | Stromelysin-1 | MMP-3 | P08254 | 4314 | MMP3 | -1.670457 | 0.081369 |
| SL004557 | CD59 glycoprotein | CD59 | P13987 | 966 | CD59 | 1.144034 | 0.083627 |
| SL000338 | Calpastatin | Calpastatin | P20810 | 831 | CAST | 1.470666 | 0.085018 |
| SL000321 | Complement C5b-C6 complex | C5b, 6 Complex | P01031,P13671 | 727 729 | C5 C6 | -1.240973 | 0.085541 |
| SL004154 | Neural cell adhesion molecule L1 | NCAM-L1 | P32004 | 3897 | L1CAM | -1.16453 | 0.086612 |
| SL007642 | Angiopoietin-related protein 4 | ANGL4 | Q9BY76 | 51129 | ANGPTL4 | 1.215728 | 0.086724 |
| SL000006 | Plasminogen activator inhibitor 1 | PAI-1 | P05121 | 5054 | SERPINE1 | 1.450114 | 0.087141 |
| SL003332 | Matrix metalloproteinase-17 | MMP-17 | Q9ULZ9 | 4326 | MMP17 | 1.207915 | 0.087262 |
| SL006917 | Tyrosine-protein kinase Lyn | LYN | P07948 | 4067 | LYN | -1.72094 | 0.087749 |
| SL004060 | Endothelin-converting enzyme 1 | Endothelin-converting enzyme 1 | P42892 | 1889 | ECE1 | -1.196789 | 0.088325 |
| SL007356 | Neurogenic locus notch homolog protein 2 | NOTC2 | Q04721 | 4853 | NOTCH2 | -1.844729 | 0.088621 |
| SL000532 | SPARC | ON | P09486 | 6678 | SPARC | -1.236811 | 0.088625 |
| SL002684 | Macrophage colony-stimulating factor 1 | CSF-1 | P09603 | 1435 | CSF1 | 1.166818 | 0.089083 |
| SL006992 | Matrilin-3 | MATN3 | O15232 | 4148 | MATN3 | 1.138467 | 0.089496 |
| SL006542 | Ficolin-2 | FCN2 | Q15485 | 2220 | FCN2 | -1.125418 | 0.090278 |
| SL000478 | Interleukin-2 | IL-2 | P60568 | 3558 | IL2 | -1.218808 | 0.090537 |
| SL005166 | Galectin-7 | Galectin-7 | P47929 | 3963 | LGALS7 | -1.103772 | 0.090708 |
| SL010500 | Tyrosine-protein kinase Lyn, isoform B | LYNB | P07948 | 4067 | LYN | -1.718801 | 0.09121 |
| SL010464 | Leucine-rich repeats and immunoglobulin-like domains protein 3 | LRIG3 | Q6UXM1 | 121227 | LRIG3 | -1.120434 | 0.092516 |
| SL000139 | Epiregulin | EPI | O14944 | 2069 | EREG | -1.379049 | 0.093374 |
| SL004862 | Programmed cell death 1 ligand 2 | PD-L2 | Q9BQ51 | 80380 | PDCD1LG2 | -1.150929 | 0.093819 |
| SL004152 | Interleukin-18 receptor 1 | IL-18 Ra | Q13478 | 8809 | IL18R1 | 1.22038 | 0.094144 |
| SL000437 | Haptoglobin | Haptoglobin, Mixed Type | P00738 | 3240 | HP | -1.169417 | 0.096065 |
| SL003703 | Bcl-2-related protein A1 | BFL1 | Q16548 | 597 | BCL2A1 | -1.077231 | 0.096199 |
| SL007024 | Gremlin-1 | GREM1 | O60565 | 26585 | GREM1 | 1.139741 | 0.096817 |
| SL006910 | Cathepsin L2 | Cathepsin V | O60911 | 1515 | CTSV | -1.182386 | 0.09701 |
| SL005189 | Interleukin-7 receptor subunit alpha | IL-7 Ra | P16871 | 3575 | IL7R | 1.14243 | 0.099616 |
| SL014468 | SH2 domain-containing protein 1A | SH21A | O60880 | 4068 | SH2D1A | 1.392899 | 0.100463 |
| SL004820 | Phosphoglycerate mutase 1 | Phosphoglycerate mutase 1 | P18669 | 5223 | PGAM1 | -3.132006 | 0.101395 |
| SL000554 | Protein kinase C delta type | PKC-D | Q05655 | 5580 | PRKCD | 1.155724 | 0.101886 |
| SL003041 | Platelet factor 4 | PF-4 | P02776 | 5196 | PF4 | -1.337033 | 0.102994 |
| SL003176 | C-C motif chemokine 1 | I-309 | P22362 | 6346 | CCL1 | -1.085321 | 0.103225 |
| SL018887 | Quinone oxidoreductase-like protein 1 | QORL1 | O95825 | 9946 | CRYZL1 | -1.183504 | 0.103946 |
| SL004708 | Connective tissue-activating peptide III | CTAP-III | P02775 | 5473 | PPBP | -1.346187 | 0.104197 |
| SL003520 | Calreticulin | calreticulin | P27797 | 811 | CALR | 1.229005 | 0.104397 |
| SL004143 | GDNF family receptor alpha-2 | GFRa-2 | O00451 | 2675 | GFRA2 | 1.127584 | 0.105487 |
| SL004142 | Ephrin-B3 | Ephrin-B3 | Q15768 | 1949 | EFNB3 | 1.239207 | 0.105695 |
| SL019978 | Interferon alpha-7 | IFNA7 | P01567 | NaN | IFNA7 | 10.829932 | 0.106598 |
| SL001973 | Mammaglobin-B | Mammaglobin 2 | O75556 | 4246 | SCGB2A1 | -1.062737 | 0.106964 |
| SL012754 | Acid sphingomyelinase-like phosphodiesterase 3a | ASM3A | Q92484 | 10924 | SMPDL3A | 1.17361 | 0.107114 |
| SL004843 | Ephrin-A2 | Ephrin-A2 | O43921 | 1943 | EFNA2 | 1.155667 | 0.10741 |
| SL006091 | Cytoplasmic protein NCK1 | NCK1 | P16333 | 4690 | NCK1 | -1.113996 | 0.108788 |
| SL005156 | Neuroblastoma suppressor of tumorigenicity 1 | DAN | P41271 | 4681 | NBL1 | 1.421933 | 0.109029 |
| HCE004333 | HCE004333 | HCE004333 | HCE004333 | HCE004333 | HCE004333 | -1.124485 | 0.109242 |
| SL008967 | Tyrosine-protein phosphatase non-receptor type substrate 1 | SHPS1 | P78324 | 140885 | SIRPA | 1.276732 | 0.10925 |
| SL008631 | Desmocollin-2 | DSC2 | Q02487 | 1824 | DSC2 | 1.342101 | 0.109333 |
| SL007651 | Fibroblast growth factor 23 | FGF23 | Q9GZV9 | 8074 | FGF23 | 1.476926 | 0.109484 |
| SL010450 | CD48 antigen | CD48 | P09326 | 962 | CD48 | 1.346149 | 0.110049 |
| SL004366 | Tumor necrosis factor receptor superfamily member 12A | TWEAKR | Q9NP84 | 51330 | TNFRSF12A | -2.750451 | 0.110532 |
| SL000695 | Neutrophil gelatinase-associated lipocalin | Lipocalin 2 | P80188 | 3934 | LCN2 | 1.22326 | 0.110684 |
| SL005256 | Histone H1.2 | Histone H1.2 | P16403 | 3006 | HIST1H1C | 1.349964 | 0.110767 |
| SL002785 | N-terminal pro-BNP | N-terminal pro-BNP | P16860 | 4879 | NPPB | 1.270355 | 0.111015 |
| SL004859 | Tumor necrosis factor receptor superfamily member 18 | GITR | Q9Y5U5 | 8784 | TNFRSF18 | -1.424802 | 0.111321 |
| SL004352 | Interleukin-17F | IL-17F | Q96PD4 | 112744 | IL17F | -1.15733 | 0.112384 |
| HCE004331 | HCE004331 | HCE004331 | HCE004331 | HCE004331 | HCE004331 | -1.123752 | 0.11259 |
| SL012517 | R-spondin-4 | RSPO4 | Q2I0M5 | 343637 | RSPO4 | 1.114109 | 0.113287 |
| SL000039 | Interleukin-8 | IL-8 | P10145 | 3576 | CXCL8 | 1.5502 | 0.114091 |
| SL003170 | C-C motif chemokine 26 | Eotaxin-3 | Q9Y258 | 10344 | CCL26 | 1.203028 | 0.114826 |
| SL010519 | Tyrosine-protein kinase ZAP-70 | ZAP70 | P43403 | 7535 | ZAP70 | -1.124005 | 0.114969 |
| SL003192 | Properdin | Properdin | P27918 | 5199 | CFP | -1.060685 | 0.115297 |
| SL011068 | Interleukin-17 receptor C | IL-17 RC | Q8NAC3 | 84818 | IL17RC | 2.940185 | 0.115312 |
| SL006119 | Trefoil factor 3 | TFF3 | Q07654 | 7033 | TFF3 | 1.245027 | 0.115579 |
| SL011769 | Acidic leucine-rich nuclear phosphoprotein 32 family member B | AN32B | Q92688 | 10541 | ANP32B | -1.110513 | 0.117087 |
| SL004908 | Tropomyosin beta chain | Tropomyosin 2 | P07951 | 7169 | TPM2 | -1.141005 | 0.117818 |
| SL004260 | Resistin | resistin | Q9HD89 | 56729 | RETN | 1.218405 | 0.11825 |
| SL003993 | Bone morphogenetic protein 6 | BMP-6 | P22004 | 654 | BMP6 | 1.303659 | 0.118318 |
| SL000597 | Lymphotoxin-alpha | TNF-b | P01374 | 4049 | LTA | 1.133462 | 0.118997 |
| SL007250 | Phosphatidylinositol 4,5-bisphosphate 3-kinase catalytic subunit gamma isoform | PK3CG | P48736 | 5294 | PIK3CG | -1.123836 | 0.121455 |
| SL009324 | Follistatin-related protein 3 | FSTL3 | O95633 | 10272 | FSTL3 | 1.236082 | 0.122643 |
| SL000048 | Vitamin K-dependent protein C | Protein C | P04070 | 5624 | PROC | -1.101147 | 0.122933 |
| SL014896 | Ankyrin-2 | ANK2 | Q01484 | 287 | ANK2 | -1.550827 | 0.123663 |
| SL004139 | Ephrin-A3 | Ephrin-A3 | P52797 | 1944 | EFNA3 | 1.39248 | 0.12511 |
| SL008728 | Neurexin-3-beta | NRX3B | Q9HDB5 | 9369 | NRXN3 | 1.189433 | 0.125377 |
| SL004067 | Group 10 secretory phospholipase A2 | GX | O15496 | 8399 | PLA2G10 | 1.09984 | 0.126395 |
| SL000283 | Beta-2-microglobulin | b2-Microglobulin | P61769 | 567 | B2M | 1.186456 | 0.128924 |
| SL004814 | Coactosin-like protein | Coactosin-like protein | Q14019 | 23406 | COTL1 | -1.102261 | 0.130265 |
| HCE001796 | HCE001796 | HCE001796 | HCE001796 | HCE001796 | HCE001796 | -1.075719 | 0.130859 |
| SL007806 | Interleukin-22 receptor subunit alpha-1 | IL22RA1 | Q8N6P7 | 58985 | IL22RA1 | 1.16632 | 0.132811 |
| SL000450 | 60 kDa heat shock protein, mitochondrial | HSP 60 | P10809 | 3329 | HSPD1 | 1.670418 | 0.134033 |
| SL000045 | Insulin-like growth factor-binding protein 3 | IGFBP-3 | P17936 | 3486 | IGFBP3 | -1.211804 | 0.134428 |
| SL004356 | C-C motif chemokine 4-like | LAG-1 | Q8NHW4 | 388372 | CCL4L1 | -1.241467 | 0.13498 |
| SL012108 | 1-phosphatidylinositol 4,5-bisphosphate phosphodiesterase gamma-1 | PLCG1 | P19174 | 5335 | PLCG1 | -1.06791 | 0.137424 |
| SL004364 | Tumor necrosis factor receptor superfamily member 13B | TACI | O14836 | 23495 | TNFRSF13B | 1.115745 | 0.137431 |
| SL003761 | Phosphatidylinositol 3,4,5-trisphosphate 3-phosphatase and dual-specificity protein phosphatase PTEN | pTEN | P60484 | 5728 | PTEN | 1.285621 | 0.138086 |
| SL007328 | Protein jagged-1 | JAG1 | P78504 | 182 | JAG1 | -1.114936 | 0.139121 |
| SL007280 | Dipeptidyl peptidase 1 | CATC | P53634 | 1075 | CTSC | 2.111889 | 0.139209 |
| SL004697 | Protein E7_HPV16 | HPV E7 Type 16 | P03129 | 1489079 | Human-virus | 1.09815 | 0.139769 |
| SL006694 | Beta-Ala-His dipeptidase | CNDP1 | Q96KN2 | 84735 | CNDP1 | -1.105337 | 0.139897 |
| SL005158 | C-type lectin domain family 4 member M | DC-SIGNR | Q9H2X3 | 10332 | CLEC4M | -1.093465 | 0.140126 |
| SL003178 | Intercellular adhesion molecule 3 | sICAM-3 | P32942 | 3385 | ICAM3 | 1.137302 | 0.140662 |
| SL008609 | Low affinity immunoglobulin gamma Fc region receptor III-B | FCG3B | O75015 | 2215 | FCGR3B | 1.149645 | 0.141209 |
| SL004351 | Interleukin-25 | IL-17E | Q9H293 | 64806 | IL25 | 1.183194 | 0.141542 |
| SL011533 | Small glutamine-rich tetratricopeptide repeat-containing protein alpha | SGTA | O43765 | 6449 | SGTA | -1.275407 | 0.141558 |
| SL000572 | Serum amyloid A-1 protein | SAA | P0DJI8 | 6288 | SAA1 | -1.411763 | 0.142038 |
| SL004146 | Interleukin-1 receptor-like 1 | IL-1 R4 | Q01638 | 9173 | IL1RL1 | 1.349417 | 0.142161 |
| SL018256 | Choline/ethanolamine kinase | CHKB | Q9Y259 | 1120 | CHKB | -1.108884 | 0.142809 |
| SL010505 | Megakaryocyte-associated tyrosine-protein kinase | MATK | P42679 | 4145 | MATK | 1.121781 | 0.143265 |
| SL005846 | Moesin | Moesin | P26038 | 4478 | MSN | -1.368947 | 0.14448 |
| SL007056 | Bone morphogenetic protein 10 | BMP10 | O95393 | 27302 | BMP10 | 1.204767 | 0.145017 |
| SL010617 | Tryptase beta-2 | TPSB2 | P20231 | 64499 | TPSB2 | 1.213585 | 0.146406 |
| SL011510 | Somatostatin-28 | Somatostatin-28 | P61278 | 6750 | SST | -1.095561 | 0.146412 |
| SL004326 | Tumor necrosis factor ligand superfamily member 18 | TNFSF18 | Q9UNG2 | 8995 | TNFSF18 | -1.065115 | 0.146708 |
| SL003739 | Tumor necrosis factor receptor superfamily member 6B | DcR3 | O95407 | 8771 | TNFRSF6B | 1.317228 | 0.148419 |
| SL000466 | Insulin-like growth factor-binding protein 2 | IGFBP-2 | P18065 | 3485 | IGFBP2 | 1.246034 | 0.149458 |
| SL000345 | Cathepsin G | Cathepsin G | P08311 | 1511 | CTSG | 1.176516 | 0.14954 |
| SL004867 | Carbonic anhydrase 3 | Carbonic anhydrase III | P07451 | 761 | CA3 | 2.185381 | 0.149663 |
| SL004646 | Layilin | Layilin | Q6UX15 | 143903 | LAYN | 1.132327 | 0.150981 |
| SL016548 | AMP Kinase (alpha1beta1gamma1) | AMPK a1b1g1 | Q13131 Q9Y478 P54619 | 5562 5564 5571 | PRKAA1 PRKAB1 PRKAG1 | 1.071846 | 0.151897 |
| SL007696 | Complement component C1q receptor | C1QR1 | Q9NPY3 | 22918 | CD93 | 1.131679 | 0.152413 |
| SL005034 | GTP-binding nuclear protein Ran | RAN | P62826 | 5901 | RAN | -1.228454 | 0.152683 |
| SL010390 | Coiled-coil domain-containing protein 80 | URB | Q76M96 | 151887 | CCDC80 | 1.109986 | 0.152956 |
| SL004365 | Tumor necrosis factor ligand superfamily member 12 | TWEAK | O43508 | 8742 | TNFSF12 | -1.089093 | 0.153053 |
| SL004668 | Apolipoprotein E (isoform E3) | Apo E3 | P02649 | 348 | APOE | -1.130652 | 0.153648 |
| SL000584 | Transforming growth factor beta-1 | TGF-b1 | P01137 | 7040 | TGFB1 | 1.13533 | 0.154891 |
| SL010495 | Cyclin-dependent kinase 2:Cyclin-A2 complex | CDK2/cyclin A | P24941 P20248 | 1017 890 | CDK2 CCNA2 | 1.127367 | 0.155641 |
| SL010461 | High affinity immunoglobulin gamma Fc receptor I | FCGR1 | P12314 | 2209 | FCGR1A | 1.132606 | 0.157537 |
| SL002782 | Adrenomedullin | Adrenomedullin | P35318 | 133 | ADM | -1.09279 | 0.157973 |
| SL008378 | Eukaryotic translation initiation factor 4E-binding protein 2 | 4EBP2 | Q13542 | 1979 | EIF4EBP2 | 1.218146 | 0.158452 |
| SL010369 | Carbonic anhydrase 4 | Carbonic Anhydrase IV | P22748 | 762 | CA4 | -1.131702 | 0.159137 |
| SL005308 | Proteasome activator complex subunit 3 | PSME3 | P61289 | 10197 | PSME3 | 1.174549 | 0.160422 |
| SL014008 | Alpha-(1,3)-fucosyltransferase 5 | FUT5 | Q11128 | 2527 | FUT5 | -1.241575 | 0.161855 |
| SL000325 | Complement component C9 | C9 | P02748 | 735 | C9 | -1.069185 | 0.163012 |
| SL004349 | Interferon lambda-2 | IFN-lambda 2 | Q8IZJ0 | 282616 | IFNL2 | 1.134006 | 0.163834 |
| SL000440 | Hemopexin | Hemopexin | P02790 | 3263 | HPX | -1.231101 | 0.164097 |
| SL004010 | Mast/stem cell growth factor receptor Kit | SCF sR | P10721 | 3815 | KIT | -1.165259 | 0.164439 |
| SL003687 | Nucleoside diphosphate kinase A | Nucleoside diphosphate kinase A | P15531 | 4830 | NME1 | -1.137344 | 0.165433 |
| SL004861 | Interleukin-18 receptor accessory protein | IL-18 Rb | O95256 | 8807 | IL18RAP | -1.098013 | 0.167328 |
| SL001721 | Platelet endothelial cell adhesion molecule | PECAM-1 | P16284 | 5175 | PECAM1 | 1.10788 | 0.168428 |
| SL004157 | Tumor necrosis factor receptor superfamily member 10B | TRAIL R2 | O14763 | 8795 | TNFRSF10B | 1.14309 | 0.169164 |
| SL006675 | Cytoskeleton-associated protein 2 | CKAP2 | Q8WWK9 | 26586 | CKAP2 | -1.084008 | 0.169562 |
| HCE004152 | HCE004152 | HCE004152 | HCE004152 | HCE004152 | HCE004152 | -1.101465 | 0.169573 |
| SL005214 | Semaphorin-6A | Semaphorin-6A | Q9H2E6 | 57556 | SEMA6A | -1.130164 | 0.170372 |
| SL000523 | Collagenase 3 | MMP-13 | P45452 | 4322 | MMP13 | 1.222651 | 0.172102 |
| SL011448 | Tumor necrosis factor receptor superfamily member 4 | TNR4 | P43489 | 7293 | TNFRSF4 | 1.083006 | 0.172843 |
| SL016555 | Dual 3',5'-cyclic-AMP and -GMP phosphodiesterase 11A | PDE11 | Q9HCR9 | 50940 | PDE11A | -1.108737 | 0.173861 |
| SL003735 | Tumor necrosis factor ligand superfamily member 9 | 4-1BB ligand | P41273 | 8744 | TNFSF9 | 1.114117 | 0.174945 |
| SL000557 | Protein kinase C zeta type | PKC-Z | Q05513 | 5590 | PRKCZ | 1.160236 | 0.175201 |
| SL003848 | Annexin A5 | Annexin V | P08758 | 308 | ANXA5 | -1.280749 | 0.175853 |
| SL004910 | Fructose-bisphosphate aldolase A | aldolase A | P04075 | 226 | ALDOA | 1.142924 | 0.177113 |
| SL005183 | Interleukin-22 receptor subunit alpha-2 | IL-22BP | Q969J5 | 116379 | IL22RA2 | -1.22631 | 0.178799 |
| SL014288 | Cell surface glycoprotein CD200 receptor 1 | MO2R1 | Q8TD46 | 131450 | CD200R1 | -1.165128 | 0.179818 |
| SL005179 | Interleukin-36 beta | IL-1F8 | Q9NZH7 | 27177 | IL36B | 1.112993 | 0.179994 |
| SL005178 | Interleukin-37 | IL-1F7 | Q9NZH6 | 27178 | IL37 | -1.073616 | 0.180484 |
| SL019979 | Histone H2A type 3 | H2A3 | Q7L7L0 | NaN | HIST3H2A | 1.181987 | 0.180632 |
| SL018509 | R-spondin-3 | RSPO3 | Q9BXY4 | 84870 | RSPO3 | 1.108511 | 0.180652 |
| SL005764 | Scavenger receptor cysteine-rich type 1 protein M130 | sCD163 | Q86VB7 | 9332 | CD163 | 1.227971 | 0.180938 |
| SL004475 | Interferon alpha/beta receptor 1 | IFN-a/b R1 | P17181 | 3454 | IFNAR1 | -1.093237 | 0.180995 |
| SL003680 | Advanced glycosylation end product-specific receptor, soluble | sRAGE | Q15109 | 177 | AGER | 1.335517 | 0.181826 |
| SL006803 | Angiopoietin-related protein 3 | ANGL3 | Q9Y5C1 | 27329 | ANGPTL3 | 1.112194 | 0.181949 |
| SL007022 | Heterogeneous nuclear ribonucleoprotein Q | HNRPQ | O60506 | 10492 | SYNCRIP | -1.09022 | 0.18266 |
| SL004153 | Macrophage colony-stimulating factor 1 receptor | M-CSF R | P07333 | 1436 | CSF1R | -1.088957 | 0.182788 |
| SL018947 | Ubiquitin carboxyl-terminal hydrolase 25 | UBP25 | Q9UHP3 | 29761 | USP25 | -1.129038 | 0.183112 |
| SL003171 | Fibroblast growth factor 4 | FGF-4 | P08620 | 2249 | FGF4 | 1.087646 | 0.183154 |
| SL010501 | Dual specificity mitogen-activated protein kinase kinase 2 | MP2K2 | P36507 | 5605 | MAP2K2 | 1.138734 | 0.18486 |
| SL000479 | Interleukin-3 | IL-3 | P08700 | 3562 | IL3 | 1.129508 | 0.185169 |
| HCE004359 | HCE004359 | HCE004359 | HCE004359 | HCE004359 | HCE004359 | -1.098059 | 0.185447 |
| SL012248 | Focal adhesion kinase 1 | FAK1 | Q05397 | 5747 | PTK2 | 1.214582 | 0.185977 |
| SL008709 | Desmocollin-3 | DSC3 | Q14574 | 1825 | DSC3 | -1.107127 | 0.187517 |
| SL004063 | Fibroblast growth factor receptor 3 | FGFR-3 | P22607 | 2261 | FGFR3 | 1.097333 | 0.188283 |
| SL005205 | Natural cytotoxicity triggering receptor 3 | NKp30 | O14931 | 259197 | NCR3 | -1.142172 | 0.190751 |
| SL007640 | C-type lectin domain family 7 member A | CLC7A | Q9BXN2 | 64581 | CLEC7A | 1.14316 | 0.192005 |
| SL004144 | GDNF family receptor alpha-3 | GFRa-3 | O60609 | 2676 | GFRA3 | 1.128343 | 0.192416 |
| SL006892 | Tyrosine-protein kinase ABL1 | ABL1 | P00519 | 25 | ABL1 | 1.079655 | 0.193463 |
| SL007774 | Protein jagged-2 | JAG2 | Q9Y219 | 3714 | JAG2 | -1.461318 | 0.194293 |
| SL014091 | VPS10 domain-containing receptor SorCS2 | SORC2 | Q96PQ0 | 57537 | SORCS2 | -4.132813 | 0.195483 |
| SL005574 | Aminoacylase-1 | Aminoacylase-1 | Q03154 | 95 | ACY1 | 2.029247 | 0.196212 |
| SL003301 | Ck-beta-8-1 | Ck-b-8-1 | P55773 | 6368 | CCL23 | 1.177837 | 0.196848 |
| SL011535 | RNA-binding protein 39 | RBM39 | Q14498 | 9584 | RBM39 | 1.13285 | 0.198706 |
| SL005159 | Erythropoietin receptor | EPO-R | P19235 | 2057 | EPOR | -1.099818 | 0.199673 |
| SL004304 | Syntaxin-1A | STX1a | Q16623 | 6804 | STX1A | 1.093415 | 0.201215 |
| HCE000342 | HCE000342 | HCE000342 | HCE000342 | HCE000342 | HCE000342 | -1.066349 | 0.203193 |
| SL018921 | HemK methyltransferase family member 2 | HEMK2 | Q9Y5N5 | 29104 | N6AMT1 | -1.143085 | 0.205917 |
| SL004180 | Tumor necrosis factor receptor superfamily member 8 | CD30 | P28908 | 943 | TNFRSF8 | 1.130065 | 0.207704 |
| SL007059 | Lymphocyte antigen 86 | LY86 | O95711 | 9450 | LY86 | -1.047523 | 0.20828 |
| SL004350 | Interleukin-17B | IL-17B | Q9UHF5 | 27190 | IL17B | 1.196784 | 0.209596 |
| SL004298 | Granzyme A | granzyme A | P12544 | 3001 | GZMA | -1.094855 | 0.209888 |
| SL016551 | Casein kinase II 2-alpha':2-beta heterotetramer | CK2-A2:B | P19784 P67870 | 1459 1460 | CSNK2A2 CSNK2B | -1.239818 | 0.210526 |
| SL000570 | Secretin | Secretin | P09683 | 6343 | SCT | -1.08434 | 0.210765 |
| SL004492 | Toll-like receptor 2 | TLR2 | O60603 | 7097 | TLR2 | -1.069273 | 0.21081 |
| SL003220 | C3a anaphylatoxin des Arginine | C3adesArg | P01024 | 718 | C3 | -1.146647 | 0.210881 |
| SL008071 | 14-3-3 protein theta | 14-3-3 protein theta | P27348 | 10971 | YWHAQ | 1.073163 | 0.212635 |
| SL006527 | EGF-containing fibulin-like extracellular matrix protein 1 | FBLN3 | Q12805 | 2202 | EFEMP1 | 1.071049 | 0.213143 |
| PDGFB | Thrombospondin-1 | Thrombospondin-1 | P07996 | 7057 | THBS1 | -1.224467 | 0.213522 |
| SL004652 | Wnt inhibitory factor 1 | WIF-1 | Q9Y5W5 | 11197 | WIF1 | -1.103136 | 0.213966 |
| SL001990 | Interleukin-1 receptor antagonist protein | IL-1Ra | P18510 | 3557 | IL1RN | 1.172734 | 0.214375 |
| SL008023 | Hyaluronan and proteoglycan link protein 1 | HPLN1 | P10915 | 1404 | HAPLN1 | -1.091624 | 0.215982 |
| SL005572 | Gelsolin | Gelsolin | P06396 | 2934 | GSN | 1.117467 | 0.217906 |
| SL002565 | Myc proto-oncogene protein | c-Myc | P01106 | 4609 | MYC | 1.487271 | 0.217919 |
| SL010522 | Cyclin-dependent kinase 8:Cyclin-C complex | CDK8/cyclin C | P49336 P24863 | 1024 892 | CDK8 CCNC | -1.089419 | 0.219744 |
| SL006610 | A disintegrin and metalloproteinase with thrombospondin motifs 13 | ATS13 | Q76LX8 | 11093 | ADAMTS13 | -1.134705 | 0.220384 |
| SL005352 | Fatty acid-binding protein, epidermal | FABPE | Q01469 | 2171 | FABP5 | 1.149908 | 0.220412 |
| SL004297 | Neuregulin-1 | NEUREGULIN-1 | Q02297 | 3084 | NRG1 | 6.332505 | 0.220696 |
| SL008402 | Ly6/PLAUR domain-containing protein 3 | LYPD3 | O95274 | 27076 | LYPD3 | 1.101209 | 0.220906 |
| SL003524 | Protein disulfide-isomerase A3 | Protein disulfide isomerase A3 | P30101 | 2923 | PDIA3 | -1.095369 | 0.221143 |
| SL004872 | Tumor necrosis factor receptor superfamily member EDAR | EDAR | Q9UNE0 | 10913 | EDAR | -1.139082 | 0.222067 |
| SL005157 | CD209 antigen | DC-SIGN | Q9NNX6 | 30835 | CD209 | 1.098723 | 0.222731 |
| SL004725 | Histone acetyltransferase type B catalytic subunit | Hat1 | O14929 | 8520 | HAT1 | 1.159303 | 0.223025 |
| SL004305 | DNA topoisomerase 1 | Topoisomerase I | P11387 | 7150 | TOP1 | 1.210742 | 0.223647 |
| SL005177 | Interleukin-36 alpha | IL-1F6 | Q9UHA7 | 27179 | IL36A | -1.383765 | 0.223994 |
| SL000057 | Thymidine kinase, cytosolic | Thymidine kinase | P04183 | 7083 | TK1 | 1.153401 | 0.224571 |
| SL003679 | Cation-independent mannose-6-phosphate receptor | IGF-II receptor | P11717 | 3482 | IGF2R | -1.07974 | 0.225177 |
| SL000323 | Complement component C7 | C7 | P10643 | 730 | C7 | 1.095799 | 0.225314 |
| SL002762 | Chromogranin-A | CgA | P10645 | 1113 | CHGA | -1.403422 | 0.225815 |
| SL001800 | Tumor necrosis factor receptor superfamily member 1B | TNF sR-II | P20333 | 7133 | TNFRSF1B | 1.165371 | 0.226584 |
| SL010250 | Stress-induced-phosphoprotein 1 | Stress-induced-phosphoprotein 1 | P31948 | 10963 | STIP1 | -1.122683 | 0.22722 |
| SL000508 | Lymphotoxin alpha2:beta1 | Lymphotoxin a2/b1 | P01374, Q06643 | 4049 4050 | LTA LTB | 1.070597 | 0.227242 |
| SL004335 | Fibroblast growth factor 17 | FGF-17 | O60258 | 8822 | FGF17 | 1.080995 | 0.227629 |
| SL004458 | Elafin | Elafin | P19957 | 5266 | PI3 | 1.308879 | 0.229541 |
| SL013989 | R-spondin-2 | RSPO2 | Q6UXX9 | 340419 | RSPO2 | 1.098553 | 0.230464 |
| SL006108 | CD5 antigen-like | CD5L | O43866 | 922 | CD5L | 1.19882 | 0.230491 |
| SL016130 | Transcription factor IIIB 90 kDa subunit | BRF-1 | Q92994 | 2972 | BRF1 | 1.142618 | 0.231539 |
| SL011809 | dCTP pyrophosphatase 1 | XTP3A | Q9H773 | 79077 | DCTPP1 | 1.105393 | 0.231975 |
| SL008504 | N-acetylglucosamine-6-sulfatase | GNS | P15586 | 2799 | GNS | 1.080541 | 0.233095 |
| SL014130 | Carbohydrate sulfotransferase 6 | CHST6 | Q9GZX3 | 4166 | CHST6 | 1.078798 | 0.233566 |
| SL005699 | Apolipoprotein L1 | Apo L1 | O14791 | 8542 | APOL1 | 1.18288 | 0.235085 |
| SL004119 | Epithelial discoidin domain-containing receptor 1 | discoidin domain receptor 1 | Q08345 | 780 | DDR1 | -1.110077 | 0.235602 |
| SL008808 | S-phase kinase-associated protein 1 | SKP1 | P63208 | 6500 | SKP1 | 1.059648 | 0.237603 |
| SL004671 | Tumor necrosis factor receptor superfamily member 13C | BAFF Receptor | Q96RJ3 | 115650 | TNFRSF13C | -1.138695 | 0.238421 |
| SL011628 | Drebrin-like protein | DBNL | Q9UJU6 | 28988 | DBNL | -1.098043 | 0.238697 |
| SL008177 | Complement component 1 Q subcomponent-binding protein, mitochondrial | C1QBP | Q07021 | 708 | C1QBP | -1.054085 | 0.239134 |
| SL010524 | Serine/threonine-protein kinase WNK3 | WNK3 | Q9BYP7 | 65267 | WNK3 | -1.23905 | 0.239852 |
| SL005167 | Galectin-8 | Galectin-8 | O00214 | 3964 | LGALS8 | -2.221708 | 0.240175 |
| SL004683 | Noggin | Noggin | Q13253 | 9241 | NOG | 1.177949 | 0.24028 |
| SL004757 | Vacuolar protein sorting-associated protein VTA1 homolog | DRG-1 | Q9NP79 | 51534 | VTA1 | -1.283777 | 0.24123 |
| SL000441 | Hepatocyte growth factor | HGF | P14210 | 3082 | HGF | 1.123816 | 0.241575 |
| SL004689 | WNT1-inducible-signaling pathway protein 1 | WISP-1 | O95388 | 8840 | WISP1 | -1.110543 | 0.24252 |
| SL000047 | Insulin-like growth factor I | IGF-I | P05019 | 3479 | IGF1 | -1.062448 | 0.242595 |
| SL010460 | Low affinity immunoglobulin gamma Fc region receptor II-b | FCG2B | P31994 | 2213 | FCGR2B | 1.237638 | 0.242832 |
| SL004844 | Ephrin type-A receptor 5 | EphA5 | P54756 | 2044 | EPHA5 | -1.065765 | 0.244009 |
| SL004724 | Histone acetyltransferase KAT6A | MOZ | Q92794 | 7994 | KAT6A | 1.099971 | 0.244603 |
| SL014088 | Fc receptor-like protein 3 | FCRL3 | Q96P31 | 115352 | FCRL3 | 1.075533 | 0.245377 |
| SL004648 | Tumor necrosis factor ligand superfamily member 14 | LIGHT | O43557 | 8740 | TNFSF14 | -1.084786 | 0.245937 |
| SL000445 | Protein Rev_HV2BE | HIV-2 Rev | P18093 | 1724716 | Human-virus | 1.055809 | 0.246309 |
| SL004669 | Apolipoprotein E (isoform E4) | Apo E4 | P02649 | 348 | APOE | -1.092661 | 0.246642 |
| SL000556 | Protein kinase C gamma type | PKC-G | P05129 | 5582 | PRKCG | -1.012096 | 0.24697 |
| SL010348 | Fibronectin Fragment 4 | FN1.4 | P02751 | 2335 | FN1 | 1.113061 | 0.247226 |
| SL007100 | Leukotriene A-4 hydrolase | LKHA4 | P09960 | 4048 | LTA4H | 1.110538 | 0.248885 |
| SL010376 | Membrane metallo-endopeptidase-like 1 | MMEL2 | Q495T6 | 79258 | MMEL1 | 1.451943 | 0.249461 |
| SL000055 | Cadherin-1 | Cadherin E | P12830 | 999 | CDH1 | -1.094038 | 0.249464 |
| SL006132 | Lamin-B1 | Lamin-B1 | P20700 | 4001 | LMNB1 | 1.069366 | 0.25157 |
| SL016549 | AMP Kinase (alpha2beta2gamma1) | AMPK a2b2g1 | P54646 O43741 P54619 | 5563 5565 5571 | PRKAA2 PRKAB2 PRKAG1 | -1.402683 | 0.251799 |
| SL002731 | Tumor necrosis factor receptor superfamily member 6 | Fas, soluble | P25445 | 355 | FAS | 1.181398 | 0.252565 |
| SL007531 | BMP-binding endothelial regulator protein | BMPER | Q8N8U9 | 168667 | BMPER | 1.083752 | 0.252781 |
| SL003302 | C-C motif chemokine 23 | MPIF-1 | P55773 | 6368 | CCL23 | 1.17624 | 0.254837 |
| SL004685 | Persephin | Persephin | O60542 | 5623 | PSPN | -1.10329 | 0.255043 |
| SL007729 | Endoplasmic reticulum aminopeptidase 1 | ARTS1 | Q9NZ08 | 51752 | ERAP1 | 1.088367 | 0.255565 |
| SL004367 | Dickkopf-related protein 1 | DKK1 | O94907 | 22943 | DKK1 | -1.095305 | 0.255582 |
| SL006374 | Cytoplasmic tyrosine-protein kinase BMX | BMX | P51813 | 660 | BMX | -1.083353 | 0.256304 |
| SL004331 | Ciliary neurotrophic factor | CNTF | P26441 | 1270 | CNTF | 1.119313 | 0.257643 |
| SL005115 | Spondin-1 | Spondin-1 | Q9HCB6 | 10418 | SPON1 | 1.094459 | 0.258139 |
| SL005230 | Netrin receptor UNC5C | UNC5H3 | O95185 | 8633 | UNC5C | 1.107555 | 0.258331 |
| SL016557 | 3-hydroxy-3-methylglutaryl-coenzyme A reductase | HMGR | P04035 | 3156 | HMGCR | 1.117679 | 0.259252 |
| SL003800 | Kallikrein-4 | Kallikrein 4 | Q9Y5K2 | 9622 | KLK4 | 1.061703 | 0.260657 |
| SL003329 | C-C motif chemokine 14 | HCC-1 | Q16627 | 6358 | CCL14 | 1.207757 | 0.263533 |
| SL016563 | Mitochondrial glutamate carrier 2 | GHC2 | Q9H1K4 | 83733 | SLC25A18 | 1.059473 | 0.263668 |
| SL011406 | High affinity cAMP-specific 3',5'-cyclic phosphodiesterase 7A | PDE7A | Q13946 | 5150 | PDE7A | 1.061038 | 0.264025 |
| SL004064 | Phospholipase A2 | GIB | P04054 | 5319 | PLA2G1B | -1.418869 | 0.264439 |
| SL004864 | Cadherin-12 | Cadherin-12 | P55289 | 1010 | CDH12 | 1.079026 | 0.266244 |
| SL000164 | Myoglobin | Myoglobin | P02144 | 4151 | MB | 1.651133 | 0.267126 |
| SL004482 | Endoglin | Endoglin | P17813 | 2022 | ENG | -1.113583 | 0.267169 |
| SL000312 | Complement C3 | C3 | P01024 | 718 | C3 | -1.078361 | 0.268131 |
| SL004136 | Tyrosine-protein kinase receptor TYRO3 | Dtk | Q06418 | 7301 | TYRO3 | -1.048564 | 0.268308 |
| SL004183 | Cadherin-3 | P-Cadherin | P22223 | 1001 | CDH3 | -1.112435 | 0.268338 |
| SL004782 | Tumor necrosis factor-inducible gene 6 protein | TSG-6 | P98066 | 7130 | TNFAIP6 | -1.147099 | 0.268467 |
| SL004639 | NT-3 growth factor receptor | TrkC | Q16288 | 4916 | NTRK3 | -1.100287 | 0.268719 |
| SL000343 | Cathepsin B | Cathepsin B | P07858 | 1508 | CTSB | -1.080556 | 0.268851 |
| SL004081 | Decorin | Bone proteoglycan II | P07585 | 1634 | DCN | 1.105812 | 0.269942 |
| SL005164 | Galectin-2 | Galectin-2 | P05162 | 3957 | LGALS2 | 1.132785 | 0.270464 |
| SL003733 | Diablo homolog, mitochondrial | SMAC | Q9NR28 | 56616 | DIABLO | -1.065572 | 0.270519 |
| SL006911 | Serine/threonine-protein kinase Chk1 | CHK1 | O14757 | 1111 | CHEK1 | 1.0695 | 0.270661 |
| SL010516 | Proto-oncogene tyrosine-protein kinase Src | SRCN1 | P12931 | 6714 | SRC | -1.614926 | 0.271432 |
| SL003674 | Bcl-2-like protein 1 | BCL2-like 1 protein | Q07817 | 598 | BCL2L1 | -1.098279 | 0.271727 |
| SL004080 | Bone morphogenetic protein receptor type-1A | BMPR1A | P36894 | 657 | BMPR1A | -1.099901 | 0.27211 |
| SL006998 | 3-phosphoinositide-dependent protein kinase 1 | PDPK1 | O15530 | 5170 | PDPK1 | -1.340966 | 0.274114 |
| SL008931 | CD177 antigen | CD177 | Q8N6Q3 | 57126 | CD177 | 1.285402 | 0.274392 |
| SL000319 | Complement C5 | C5 | P01031 | 727 | C5 | -1.094329 | 0.274627 |
| SL014270 | CMRF35-like molecule 6 | CLM6 | Q08708 | 10871 | CD300C | 1.148021 | 0.274713 |
| SL004726 | CD97 antigen | CD97 | P48960 | 976 | CD97 | -1.0621 | 0.27616 |
| SL000530 | Oncostatin-M | OSM | P13725 | 5008 | OSM | -1.070642 | 0.276337 |
| SL000507 | Lymphotoxin alpha1:beta2 | Lymphotoxin a1/b2 | P01374, Q06643 | 4049 4050 | LTA LTB | -1.058029 | 0.276633 |
| SL010610 | C-type lectin domain family 4 member K | CLC4K | Q9UJ71 | 50489 | CD207 | 1.051315 | 0.278427 |
| SL006923 | TATA-box-binding protein | TBP | P20226 | 6908 | TBP | 1.181288 | 0.279518 |
| SL000252 | Alpha-2-macroglobulin | a2-Macroglobulin | P01023 | 2 | A2M | 1.115457 | 0.279837 |
| SL004148 | Interleukin-12 receptor subunit beta-1 | IL-12 Rb1 | P42701 | 3594 | IL12RB1 | 1.078957 | 0.280288 |
| SL003869 | Growth/differentiation factor 15 | MIC-1 | Q99988 | 9518 | GDF15 | 1.22109 | 0.280432 |
| SL004469 | Amyloid beta A4 protein | amyloid precursor protein | P05067 | 351 | APP | -1.111477 | 0.281692 |
| SL014208 | Cytotoxic and regulatory T-cell molecule | CRTAM | O95727 | 56253 | CRTAM | -1.243594 | 0.282064 |
| SL000541 | Plasminogen | Plasminogen | P00747 | 5340 | PLG | -1.043844 | 0.282827 |
| SL002763 | Kallikrein-11 | Kallikrein 11 | Q9UBX7 | 11012 | KLK11 | 1.218298 | 0.283566 |
| HCE003300 | HCE003300 | HCE003300 | HCE003300 | HCE003300 | HCE003300 | -1.089224 | 0.284219 |
| SL000384 | Cytotoxic T-lymphocyte protein 4 | CTLA-4 | P16410 | 1493 | CTLA4 | -4.658453 | 0.284521 |
| SL010349 | Fibronectin Fragment 3 | FN1.3 | P02751 | 2335 | FN1 | 1.115819 | 0.28481 |
| SL000517 | C-C motif chemokine 13 | MCP-4 | Q99616 | 6357 | CCL13 | 1.125298 | 0.285028 |
| SL007336 | Protein SET | SET | Q01105 | 6418 | SET | -1.092811 | 0.285653 |
| SL004670 | Artemin | Artemin | Q5T4W7 | 9048 | ARTN | 1.721095 | 0.287014 |
| SL005223 | Interleukin-27 receptor subunit alpha | TCCR | Q6UWB1 | 9466 | IL27RA | -1.122066 | 0.287819 |
| SL010619 | Tryptase gamma | TPSG1 | Q9NRR2 | 25823 | TPSG1 | 1.081583 | 0.287843 |
| SL004875 | Interleukin-1 receptor-like 2 | IL-1Rrp2 | Q9HB29 | 8808 | IL1RL2 | 1.053693 | 0.289907 |
| SL007361 | Galectin-10 | LPPL | Q05315 | 1178 | CLC | -1.19859 | 0.289991 |
| SL005228 | NKG2D ligand 2 | ULBP-2 | Q9BZM5 | 80328 | ULBP2 | -1.112089 | 0.289992 |
| SL002783 | Cardiotrophin-1 | Cardiotrophin-1 | Q16619 | 1489 | CTF1 | -1.093007 | 0.291398 |
| SL011509 | Peptide YY | PYY | P10082 | 5697 | PYY | -1.130524 | 0.291606 |
| SL000049 | Vitamin K-dependent protein S | Protein S | P07225 | 5627 | PROS1 | -1.054845 | 0.29323 |
| SL000442 | Somatotropin | HGH | P01241 | 2688 | GH1 | -1.175623 | 0.293395 |
| SL005190 | Leukocyte immunoglobulin-like receptor subfamily B member 1 | ILT-2 | Q8NHL6 | 10859 | LILRB1 | 1.097887 | 0.293534 |
| SL008099 | Macrophage-capping protein | CAPG | P40121 | 822 | CAPG | 1.118967 | 0.294292 |
| SL000566 | Retinol-binding protein 4 | RBP | P02753 | 5950 | RBP4 | -1.034112 | 0.29432 |
| SL004101 | Mothers against decapentaplegic homolog 2 | SMAD2 | Q15796 | 4087 | SMAD2 | -1.278144 | 0.294489 |
| SL000316 | Complement C4 | C4 | P0C0L4, P0C0L5 | 720 721 | C4A C4B | -1.046051 | 0.296493 |
| SL003183 | C-X-C motif chemokine 10 | IP-10 | P02778 | 3627 | CXCL10 | 1.375029 | 0.297232 |
| SL010459 | Low affinity immunoglobulin gamma Fc region receptor II-a | FCG2A | P12318 | 2212 | FCGR2A | 1.300768 | 0.29874 |
| SL004355 | C-C motif chemokine 3-like 1 | LD78-beta | P16619 | 414062 | CCL3L1 | -1.068711 | 0.300293 |
| SL004258 | Adiponectin | Adiponectin | Q15848 | 9370 | ADIPOQ | 1.126975 | 0.302413 |
| SL000655 | Keratin, type I cytoskeletal 18 | Keratin 18 | P05783 | 3875 | KRT18 | -1.098125 | 0.303259 |
| SL000070 | Glypican-3 | Glypican 3 | P51654 | 2719 | GPC3 | -1.08141 | 0.303283 |
| SL006230 | Lumican | Lumican | P51884 | 4060 | LUM | 1.093709 | 0.30334 |
| SL000424 | Fibrinogen | Fibrinogen | P02671 P02675 P02679 | 2243 2244 2266 | FGA FGB FGG | 1.026891 | 0.303901 |
| HCE000414 | HCE000414 | HCE000414 | HCE000414 | HCE000414 | HCE000414 | -1.054237 | 0.303936 |
| SL010518 | Tyrosine-protein kinase Tec | TEC | P42680 | 7006 | TEC | -1.312946 | 0.305052 |
| SL003322 | Vascular endothelial growth factor receptor 3 | VEGF sR3 | P35916 | 2324 | FLT4 | -1.115851 | 0.305487 |
| SL004891 | Heterogeneous nuclear ribonucleoproteins A2/B1 | hnRNP A2/B1 | P22626 | 3181 | HNRNPA2B1 | 1.239155 | 0.305876 |
| SL003690 | Tumor necrosis factor receptor superfamily member 11A | RANK | Q9Y6Q6 | 8792 | TNFRSF11A | -1.179005 | 0.30593 |
| SL011708 | cAMP-regulated phosphoprotein 19 | ARP19 | P56211 | 10776 | ARPP19 | -1.199671 | 0.306063 |
| SL012707 | Proprotein convertase subtilisin/kexin type 9 | PCSK9 | Q8NBP7 | 255738 | PCSK9 | -1.079453 | 0.308304 |
| SL004920 | Cofilin-1 | Cofilin-1 | P23528 | 1072 | CFL1 | -1.167643 | 0.308367 |
| HCE003183 | HCE003183 | HCE003183 | HCE003183 | HCE003183 | HCE003183 | -1.08594 | 0.30861 |
| SL007266 | 26S proteasome non-ATPase regulatory subunit 7 | PSD7 | P51665 | 5713 | PSMD7 | 1.058567 | 0.308652 |
| SL004921 | Nucleoside diphosphate kinase B | NDP kinase B | P22392 | 4831 | NME2 | -1.289266 | 0.309283 |
| SL010374 | Methionine aminopeptidase 1 | METAP1 | P53582 | 23173 | METAP1 | -1.328491 | 0.309316 |
| SL010384 | Testican-1 | Testican-1 | Q08629 | 6695 | SPOCK1 | -1.076702 | 0.310333 |
| SL008916 | Dipeptidyl peptidase 2 | DPP2 | Q9UHL4 | 29952 | DPP7 | -1.0815 | 0.310963 |
| SL004400 | Coagulation factor IXab | Coagulation Factor IXab | P00740 | 2158 | F9 | -1.070342 | 0.31144 |
| SL001943 | Interleukin-6 receptor subunit alpha | IL-6 sRa | P08887 | 3570 | IL6R | -1.113888 | 0.31158 |
| SL009400 | Chordin-like protein 1 | CRDL1 | Q9BU40 | 91851 | CHRDL1 | 1.099811 | 0.314943 |
| SL000138 | Heparin-binding EGF-like growth factor | HB-EGF | Q99075 | 1839 | HBEGF | -1.075835 | 0.315538 |
| SL000515 | C-C motif chemokine 8 | MCP-2 | P80075 | 6355 | CCL8 | 1.12907 | 0.315737 |
| SL000467 | Immunoglobulin G | IgG | P01857 | 3500 3501 3502 3503 50802 3535 | IGHG1 IGHG2 IGHG3 IGHG4 IGK@ IGL@ | 1.06236 | 0.315962 |
| SL005430 | Secreted and transmembrane protein 1 | SECTM1 | Q8WVN6 | 6398 | SECTM1 | 1.062407 | 0.31643 |
| SL000342 | Catalase | Catalase | P04040 | 847 | CAT | 1.203513 | 0.316512 |
| SL004353 | Interleukin-17D | IL-17D | Q8TAD2 | 53342 | IL17D | 1.176212 | 0.317315 |
| SL005266 | Alpha-synuclein | a-Synuclein | P37840 | 6622 | SNCA | -1.171118 | 0.317494 |
| SL008522 | Semaphorin-5A | SEM5A | Q13591 | 9037 | SEMA5A | -1.268757 | 0.31782 |
| SL005206 | Natural cytotoxicity triggering receptor 2 | NKp44 | O95944 | 9436 | NCR2 | 1.056195 | 0.318078 |
| SL015046 | Amphoterin-induced protein 2 | AMGO2 | Q86SJ2 | 347902 | AMIGO2 | 1.152506 | 0.318975 |
| SL004660 | Bone sialoprotein 2 | BSP | P21815 | 3381 | IBSP | 1.351503 | 0.319553 |
| SL004009 | Ras-related C3 botulinum toxin substrate 1 | RAC1 | P63000 | 5879 | RAC1 | -1.200777 | 0.319598 |
| SL003334 | Endothelial monocyte-activating polypeptide 2 | EMAP-2 | Q12904 | 9255 | AIMP1 | -1.075237 | 0.319757 |
| SL010523 | Homeodomain-interacting protein kinase 3 | HIPK3 | Q9H422 | 10114 | HIPK3 | -1.241081 | 0.32072 |
| SL000276 | Apolipoprotein E | Apo E | P02649 | 348 | APOE | -1.146638 | 0.321381 |
| SL004151 | Interleukin-15 receptor subunit alpha | IL-15 Ra | Q13261 | 3601 | IL15RA | 1.081126 | 0.322411 |
| SL012740 | A disintegrin and metalloproteinase with thrombospondin motifs 15 | ATS15 | Q8TE58 | 170689 | ADAMTS15 | 1.058651 | 0.322615 |
| SL010466 | Membrane frizzled-related protein | MFRP | Q9BY79 | 83552 | MFRP | 1.082701 | 0.323616 |
| SL006378 | S-formylglutathione hydrolase | Esterase D | P10768 | 2098 | ESD | 1.234492 | 0.323644 |
| SL004610 | Low-density lipoprotein receptor-related protein 8 | LRP8 | Q14114 | 7804 | LRP8 | 1.108381 | 0.325012 |
| SL005588 | Tyrosine-protein kinase Fer | FER | P16591 | 2241 | FER | -1.389038 | 0.325701 |
| SL001729 | Granulocyte colony-stimulating factor | G-CSF | P09919 | 1440 | CSF3 | 1.14125 | 0.327079 |
| SL010467 | Repulsive guidance molecule A | RGMA | Q96B86 | 56963 | RGMA | -1.093684 | 0.327403 |
| SL005793 | Peptidyl-prolyl cis-trans isomerase F, mitochondrial | Cyclophilin F | P30405 | 10105 | PPIF | -1.532971 | 0.32783 |
| SL009628 | Inhibitor of growth protein 1 | ING1 | Q9UK53 | 3621 | ING1 | -1.144228 | 0.327941 |
| SL007217 | Interferon gamma receptor 2 | INGR2 | P38484 | 3460 | IFNGR2 | -1.079159 | 0.328468 |
| SL004536 | Hepcidin | LEAP-1 | P81172 | 57817 | HAMP | -1.125206 | 0.329665 |
| SL004661 | Aggrecan core protein | Aggrecan | P16112 | 176 | ACAN | 1.084488 | 0.330189 |
| SL001796 | Myeloperoxidase | Myeloperoxidase | P05164 | 4353 | MPO | 1.09093 | 0.330359 |
| SL004869 | Carbonic anhydrase 13 | Carbonic anhydrase XIII | Q8N1Q1 | 377677 | CA13 | -1.388156 | 0.331518 |
| SL012783 | WAP, kazal, immunoglobulin, kunitz and NTR domain-containing protein 1 | WFKN1 | Q96NZ8 | 117166 | WFIKKN1 | -1.116616 | 0.331839 |
| SL005234 | Growth/differentiation factor 9 | GDF-9 | O60383 | 2661 | GDF9 | 1.099232 | 0.332046 |
| SL004594 | Troponin I, fast skeletal muscle | Troponin I, skeletal, fast twitch | P48788 | 7136 | TNNI2 | -1.052741 | 0.332422 |
| SL003774 | Bcl-2-like protein 2 | Apoptosis regulator Bcl-W | Q92843 | 599 | BCL2L2 | 1.41561 | 0.332777 |
| SL010381 | Dickkopf-like protein 1 | Soggy-1 | Q9UK85 | 27120 | DKKL1 | -1.061021 | 0.333046 |
| SL008574 | Osteomodulin | OMD | Q99983 | 4958 | OMD | 1.11239 | 0.333317 |
| SL010508 | Serine/threonine-protein kinase PAK 3 | PAK3 | O75914 | 5063 | PAK3 | -1.471636 | 0.33371 |
| SL007385 | Interleukin-24 | IL24 | Q13007 | 11009 | IL24 | 1.089211 | 0.333949 |
| SL004852 | Programmed cell death 1 ligand 1 | B7-H1 | Q9NZQ7 | 29126 | CD274 | 1.988812 | 0.334514 |
| SL000019 | Apolipoprotein A-I | Apo A-I | P02647 | 335 | APOA1 | 1.186507 | 0.334617 |
| SL009948 | Plexin-B2 | PLXB2 | O15031 | 23654 | PLXNB2 | -1.095912 | 0.334796 |
| SL000449 | DnaJ homolog subfamily B member 1 | HSP 40 | P25685 | 3337 | DNAJB1 | -1.143013 | 0.335388 |
| SL005084 | Periostin | Periostin | Q15063 | 10631 | POSTN | 1.117278 | 0.335546 |
| SL018629 | E3 ubiquitin-protein ligase RNF43 | RNF43 | Q68DV7 | 54894 | RNF43 | 1.115068 | 0.336104 |
| SL003189 | C-C motif chemokine 19 | MIP-3b | Q99731 | 6363 | CCL19 | -1.183478 | 0.336301 |
| SL008837 | CD63 antigen | CD63 | P08962 | 967 | CD63 | -1.069513 | 0.336412 |
| SL010528 | Ubiquitin-fold modifier 1 | UFM1 | P61960 | 51569 | UFM1 | -1.191838 | 0.336736 |
| SL016128 | Thioredoxin domain-containing protein 12 | TXD12 | O95881 | 51060 | TXNDC12 | -1.086548 | 0.337116 |
| SL002078 | TGF-beta receptor type-2 | TGF-b R II | P37173 | 7048 | TGFBR2 | 1.197918 | 0.337913 |
| SL005575 | Galactoside 3(4)-L-fucosyltransferase | Fucosyltransferase 3 | P21217 | 2525 | FUT3 | 1.053932 | 0.338394 |
| SL014108 | Leucine-rich repeat transmembrane neuronal protein 3 | LRRT3 | Q86VH5 | 347731 | LRRTM3 | 1.087908 | 0.338464 |
| SL012822 | Brain-specific serine protease 4 | BSSP4 | Q9GZN4 | 64063 | PRSS22 | 1.073011 | 0.338596 |
| SL006919 | Ribosomal protein S6 kinase alpha-5 | RSK-like protein kinase | O75582 | 9252 | RPS6KA5 | -1.144503 | 0.33863 |
| SL007136 | CCAAT/enhancer-binding protein beta | CEBPB | P17676 | 1051 | CEBPB | 1.10439 | 0.338743 |
| SL004863 | Tumor necrosis factor receptor superfamily member 19 | TAJ | Q9NS68 | 55504 | TNFRSF19 | 1.142392 | 0.340009 |
| SL004650 | Secreted frizzled-related protein 3 | sFRP-3 | Q92765 | 2487 | FRZB | -1.052313 | 0.340127 |
| SL011404 | cAMP-specific 3',5'-cyclic phosphodiesterase 4D | PDE4D | Q08499 | 5144 | PDE4D | -1.074191 | 0.341886 |
| SL000020 | Apolipoprotein B | Apo B | P04114 | 338 | APOB | 1.185527 | 0.342083 |
| SL000527 | Matrix metalloproteinase-9 | MMP-9 | P14780 | 4318 | MMP9 | 1.062522 | 0.342309 |
| SL016553 | cGMP-inhibited 3',5'-cyclic phosphodiesterase A | PDE3A | Q14432 | 5139 | PDE3A | -1.073877 | 0.343948 |
| SL005225 | High affinity nerve growth factor receptor | TrkA | P04629 | 4914 | NTRK1 | 1.061542 | 0.345906 |
| SL004466 | Heparin cofactor 2 | Heparin cofactor II | P05546 | 3053 | SERPIND1 | -1.008146 | 0.347476 |
| SL000638 | Cadherin-2 | Cadherin-2 | P19022 | 1000 | CDH2 | 1.68355 | 0.347593 |
| SL002541 | Tumor necrosis factor ligand superfamily member 11 | sRANKL | O14788 | 8600 | TNFSF11 | 1.589265 | 0.347679 |
| SL002644 | Epidermal growth factor receptor | ERBB1 | P00533 | 1956 | EGFR | -1.07897 | 0.348762 |
| SL004783 | Protein S100-A12 | S100A12 | P80511 | 6283 | S100A12 | 1.250482 | 0.34889 |
| SL001716 | Interleukin-12 | IL-12 | P29459, P29460 | 3592 3593 | IL12A IL12B | -1.110477 | 0.34972 |
| SL014229 | CD83 antigen | CD83 | Q01151 | 9308 | CD83 | -1.093539 | 0.35011 |
| SL004345 | Growth/differentiation factor 11 | GDF-11 | O95390 | 10220 | GDF11 | 1.41142 | 0.351032 |
| SL000565 | Renin | Renin | P00797 | 5972 | REN | -1.315415 | 0.351171 |
| SL011180 | Hexokinase-1 | HXK1 | P19367 | 3098 | HK1 | 1.058782 | 0.351581 |
| SL007153 | Lactoperoxidase | PERL | P22079 | 4025 | LPO | -1.190915 | 0.355725 |
| SL017612 | Heat shock protein HSP 90-alpha/beta | HSP 90a/b | P07900 P08238 | NaN | HSP90AA1 HSP90AB1 | 1.131291 | 0.357577 |
| SL002505 | Atrial natriuretic factor | ANP | P01160 | 4878 | NPPA | 1.090547 | 0.360894 |
| SL002640 | Placenta growth factor | PlGF | P49763 | 5228 | PGF | -1.064958 | 0.363989 |
| SL005488 | SPARC-like protein 1 | SPARCL1 | Q14515 | 8404 | SPARCL1 | 1.087668 | 0.365697 |
| SL000615 | Vasoactive Intestinal Peptide | Vasoactive Intestinal Peptide | P01282 | 7432 | VIP | -1.129289 | 0.366066 |
| SL004438 | Cystatin-M | Cystatin M | Q15828 | 1474 | CST6 | -1.11449 | 0.366577 |
| SL008063 | Low molecular weight phosphotyrosine protein phosphatase | PPAC | P24666 | 52 | ACP1 | -1.106747 | 0.368743 |
| SL017290 | PolyUbiquitin K63-linked | PolyUbiquitin K63 | P0CG48 | 7316 | UBC | 1.274941 | 0.368826 |
| SL000509 | Tumor necrosis factor receptor superfamily member 3 | Lymphotoxin b R | P36941 | 4055 | LTBR | -1.07038 | 0.369172 |
| SL006268 | NSFL1 cofactor p47 | NSF1C | Q9UNZ2 | 55968 | NSFL1C | -1.175708 | 0.369651 |
| SL000247 | 6-phosphogluconate dehydrogenase, decarboxylating | 6-Phosphogluconate dehydrogenase | P52209 | 5226 | PGD | -1.1077 | 0.369893 |
| SL000409 | Mitogen-activated protein kinase 3 | ERK-1 | P27361 | 5595 | MAPK3 | -1.236701 | 0.370039 |
| SL008516 | Cystatin-SA | CYTT | P09228 | 1470 | CST2 | 1.177996 | 0.370127 |
| SL014292 | Sialic acid-binding Ig-like lectin 14 | SIG14 | Q08ET2 | 100049587 | SIGLEC14 | 1.064188 | 0.370614 |
| SL013489 | Protein amnionless | AMNLS | Q9BXJ7 | 81693 | AMN | -1.126083 | 0.370795 |
| SL010496 | Cyclin-dependent kinase 5:Cyclin-dependent kinase 5 activator 1 complex | CDK5/p35 | Q00535 Q15078 | 1020 8851 | CDK5 CDK5R1 | 1.071376 | 0.371296 |
| SL000496 | Lactotransferrin | Lactoferrin | P02788 | 4057 | LTF | 1.125889 | 0.371821 |
| SL000462 | Insulin-like growth factor-binding protein 1 | IGFBP-1 | P08833 | 3484 | IGFBP1 | 1.174477 | 0.37279 |
| SL014488 | Proto-oncogene vav | VAV | P15498 | 7409 | VAV1 | -1.33269 | 0.373856 |
| SL015728 | Glucokinase regulatory protein | GCKR | Q14397 | 2646 | GCKR | 1.056755 | 0.375697 |
| SL002524 | T-cell surface glycoprotein CD4 | sCD4 | P01730 | 920 | CD4 | 1.057093 | 0.377053 |
| SL004118 | Tartrate-resistant acid phosphatase type 5 | TrATPase | P13686 | 54 | ACP5 | -1.083557 | 0.379026 |
| SL007358 | Protein kinase C theta type | KPCT | Q04759 | 5588 | PRKCQ | -1.257468 | 0.37939 |
| SL004644 | Ectodysplasin-A, secreted form | EDA | Q92838 | 1896 | EDA | -1.272011 | 0.379822 |
| SL000426 | Fibronectin | Fibronectin | P02751 | 2335 | FN1 | 1.118265 | 0.380532 |
| SL004342 | Fibroblast growth factor 8 isoform B | FGF-8B | P55075 | 2253 | FGF8 | 1.083068 | 0.381439 |
| SL004296 | Adenylate kinase isoenzyme 1 | Myokinase, human | P00568 | 203 | AK1 | -1.106209 | 0.381481 |
| SL006088 | Sphingosine kinase 1 | Sphingosine kinase 1 | Q9NYA1 | 8877 | SPHK1 | -1.230497 | 0.381553 |
| SL010529 | Ubiquitin-fold modifier-conjugating enzyme 1 | UFC1 | Q9Y3C8 | 51506 | UFC1 | -1.113578 | 0.382629 |
| SL000586 | Thrombin | Thrombin | P00734 | 2147 | F2 | -1.070212 | 0.382753 |
| SL019096 | E3 SUMO-protein ligase PIAS4 | PIAS4 | Q8N2W9 | 51588 | PIAS4 | 1.126459 | 0.383908 |
| SL004248 | Glial cell line-derived neurotrophic factor | GDNF | P39905 | 2668 | GDNF | 1.073746 | 0.384126 |
| SL005155 | Teratocarcinoma-derived growth factor 1 | Cripto | P13385 | 6997 | TDGF1 | -1.454313 | 0.384582 |
| SL005170 | Inducible T-cell costimulator | ICOS | Q9Y6W8 | 29851 | ICOS | 1.066339 | 0.384973 |
| HCE000104 | HCE000104 | HCE000104 | HCE000104 | HCE000104 | HCE000104 | -1.070895 | 0.386831 |
| SL010513 | cAMP-dependent protein kinase catalytic subunit alpha | PRKACA | P17612 | 5566 | PRKACA | -1.222325 | 0.386996 |
| SL010488 | Abelson tyrosine-protein kinase 2 | ABL2 | P42684 | 27 | ABL2 | 1.165084 | 0.387245 |
| SL007281 | Mitogen-activated protein kinase 12 | MK12 | P53778 | 6300 | MAPK12 | -1.064305 | 0.387499 |
| SL008085 | 3-hydroxyisobutyrate dehydrogenase, mitochondrial | 3HIDH | P31937 | 11112 | HIBADH | -1.916217 | 0.388153 |
| SL007122 | Insulin-degrading enzyme | IDE | P14735 | 3416 | IDE | 1.13143 | 0.388303 |
| SL017614 | RAC-alpha/beta/gamma serine/threonine-protein kinase | PKB a/b/g | P31749 P31751 Q9Y243 | 207 208 10000 | AKT1 AKT2 AKT3 | -1.212961 | 0.390252 |
| SL002525 | Complement C2 | C2 | P06681 | 717 | C2 | -1.058848 | 0.390806 |
| SL002561 | Parathyroid hormone-related protein | PTHrP | P12272 | 5744 | PTHLH | 1.188148 | 0.391637 |
| SL019100 | E3 ubiquitin-protein ligase CHIP | CHIP | Q9UNE7 | 10273 | STUB1 | 1.103111 | 0.39408 |
| SL003930 | 15-hydroxyprostaglandin dehydrogenase [NAD(+)] | HPG- | P15428 | 3248 | HPGD | -1.049433 | 0.395238 |
| SL013754 | Ras GTPase-activating protein 1 | RASA1 | P20936 | 5921 | RASA1 | -1.068812 | 0.395588 |
| SL008143 | Ubiquitin-conjugating enzyme E2 N | UBE2N | P61088 | 7334 | UBE2N | -1.113368 | 0.396086 |
| SL005703 | Neurogenic locus notch homolog protein 1 | Notch 1 | P46531 | 4851 | NOTCH1 | -1.063598 | 0.396681 |
| SL000357 | Coagulation factor IX | Coagulation Factor IX | P00740 | 2158 | F9 | -1.05871 | 0.397952 |
| SL003196 | C-C motif chemokine 17 | TARC | Q92583 | 6361 | CCL17 | 1.056995 | 0.39868 |
| SL006805 | Serine/threonine-protein kinase MRCK beta | MRCKB | Q9Y5S2 | 9578 | CDC42BPB | 1.049481 | 0.402586 |
| SL000454 | Heat shock protein HSP 90-beta | HSP 90b | P08238 | 3326 | HSP90AB1 | 1.095548 | 0.407334 |
| SL020172 | Relaxin receptor 1 | RXFP1 | Q9HBX9 | NaN | RXFP1 | -1.112092 | 0.407549 |
| SL014228 | SLAM family member 6 | SLAF6 | Q96DU3 | 114836 | SLAMF6 | 1.040842 | 0.408569 |
| SL007674 | T-lymphocyte surface antigen Ly-9 | LY9 | Q9HBG7 | 4063 | LY9 | -1.07535 | 0.409238 |
| SL003650 | Alpha-enolase | Alpha enolase | P06733 | 2023 | ENO1 | -1.174152 | 0.409279 |
| SL004915 | Chloride intracellular channel protein 1 | NCC27 | O00299 | 1192 | CLIC1 | -1.136537 | 0.409741 |
| SL008759 | Platelet glycoprotein VI | GPVI | Q9HCN6 | 51206 | GP6 | -1.239409 | 0.41039 |
| SL000613 | Urokinase-type plasminogen activator | uPA | P00749 | 5328 | PLAU | 1.076232 | 0.410415 |
| SL000589 | Thyroid Stimulating Hormone | TSH | P01215 P01222 | 1081 12372 | CGA TSHB | -1.178106 | 0.410619 |
| SL006550 | Extracellular matrix protein 1 | ECM1 | Q16610 | 1893 | ECM1 | 1.097172 | 0.410809 |
| SL004752 | Protein FAM107A | DRR1 | O95990 | 11170 | FAM107A | 1.025696 | 0.411262 |
| SL002803 | Ubiquitin carboxyl-terminal hydrolase isozyme L1 | PGP9.5 | P09936 | 7345 | UCHL1 | -1.069723 | 0.411985 |
| SL003803 | Receptor tyrosine-protein kinase erbB-4 | ERBB4 | Q15303 | 2066 | ERBB4 | -1.028799 | 0.413343 |
| SL008331 | Proliferation-associated protein 2G4 | PA2G4 | Q9UQ80 | 5036 | PA2G4 | -1.129757 | 0.414405 |
| SL005730 | cGMP-dependent 3',5'-cyclic phosphodiesterase | cGMP-stimulated PDE | O00408 | 5138 | PDE2A | -1.049695 | 0.414554 |
| SL004339 | Fibroblast growth factor 5 | FGF-5 | P12034 | 2250 | FGF5 | 1.072655 | 0.415029 |
| SL004855 | Contactin-1 | contactin-1 | Q12860 | 1272 | CNTN1 | -1.01541 | 0.415053 |
| SL008486 | Galectin-9 | LEG9 | O00182 | 3965 | LGALS9 | 1.055772 | 0.417744 |
| SL004477 | Protein S100-A9 | calgranulin B | P06702 | 6280 | S100A9 | 1.07973 | 0.418651 |
| SL004704 | COMM domain-containing protein 7 | COMMD7 | Q86VX2 | 149951 | COMMD7 | -1.084705 | 0.420023 |
| SL006922 | DNA repair protein RAD51 homolog 1 | RAD51 | Q06609 | 5888 | RAD51 | 1.301595 | 0.422106 |
| SL000250 | Alpha-2-antiplasmin | a2-Antiplasmin | P08697 | 5345 | SERPINF2 | -1.034949 | 0.423004 |
| SL007680 | Roundabout homolog 2 | ROBO2 | Q9HCK4 | 6092 | ROBO2 | 1.057284 | 0.423168 |
| SL003726 | Serine/threonine-protein kinase Chk2 | Chk2 | O96017 | 11200 | CHEK2 | -1.860474 | 0.424343 |
| SL007869 | Peptidyl-prolyl cis-trans isomerase B | PPIB | P23284 | 5479 | PPIB | 1.057963 | 0.42437 |
| SL003310 | Vascular endothelial growth factor A, isoform 121 | VEGF121 | P15692 | 7422 | VEGFA | 1.061196 | 0.429096 |
| SL009045 | Ectonucleotide pyrophosphatase/phosphodiesterase family member 7 | ENPP7 | Q6UWV6 | 339221 | ENPP7 | 1.50314 | 0.430397 |
| SL008416 | C-type mannose receptor 2 | MRC2 | Q9UBG0 | 9902 | MRC2 | -1.095696 | 0.430458 |
| SL011772 | Pescadillo homolog | PESC | O00541 | 23481 | PES1 | 1.055119 | 0.430653 |
| SL004924 | Eukaryotic translation initiation factor 4H | eIF-4H | Q15056 | 7458 | EIF4H | -1.24953 | 0.431251 |
| SL003320 | Vascular endothelial growth factor D | VEGF-D | O43915 | 2277 | FIGF | 1.060087 | 0.431526 |
| SL009207 | Dynactin subunit 2 | Dynactin subunit 2 | Q13561 | 10540 | DCTN2 | 1.074401 | 0.431659 |
| SL008904 | Lymphatic vessel endothelial hyaluronic acid receptor 1 | LYVE1 | Q9Y5Y7 | 10894 | LYVE1 | 1.105353 | 0.432054 |
| SL005215 | Myeloid cell surface antigen CD33 | Siglec-3 | P20138 | 945 | CD33 | -1.06807 | 0.432061 |
| SL010510 | Serine/threonine-protein kinase PAK 7 | PAK7 | Q9P286 | 57144 | PAK7 | 1.039134 | 0.432542 |
| SL003916 | Kallikrein-12 | kallikrein 12 | Q9UKR0 | 43849 | KLK12 | -1.068919 | 0.432814 |
| SL009089 | Brevican core protein | PGCB | Q96GW7 | 63827 | BCAN | -1.090271 | 0.432997 |
| SL010515 | Ribosomal protein S6 kinase alpha-3 | RPS6KA3 | P51812 | 6197 | RPS6KA3 | -1.17903 | 0.435663 |
| SL016554 | High affinity cGMP-specific 3',5'-cyclic phosphodiesterase 9A | PDE9A | O76083 | 5152 | PDE9A | -1.000257 | 0.435702 |
| SL000678 | Granulysin | Granulysin | P22749 | 10578 | GNLY | -1.104765 | 0.436263 |
| SL005630 | Proteasome subunit alpha type-6 | PSA6 | P60900 | 5687 | PSMA6 | 1.04707 | 0.43629 |
| SL003657 | Calcineurin | Calcineurin | Q08209 P63098 | 5530 5534 | PPP3CA PPP3R1 | -1.146395 | 0.437447 |
| SL000428 | Follicle stimulating hormone | FSH | P01215, P01225 | 1081 2488 | CGA FSHB | 1.08194 | 0.438174 |
| SL000401 | Neutrophil elastase | Elastase | P08246 | 1991 | ELANE | 1.072603 | 0.438857 |
| SL003951 | Brain-derived neurotrophic factor | BDNF | P23560 | 627 | BDNF | 1.086183 | 0.439177 |
| SL006916 | Tyrosine-protein kinase Lck | LCK | P06239 | 3932 | LCK | -1.080136 | 0.439297 |
| SL005236 | Protein NOV homolog | NovH | P48745 | 4856 | NOV | 1.042554 | 0.440119 |
| SL008190 | Spectrin alpha chain, non-erythrocytic 1 | SPTA2 | Q13813 | 6709 | SPTAN1 | 1.062981 | 0.440737 |
| SL004486 | Vascular endothelial growth factor C | VEGF-C | P49767 | 7424 | VEGFC | 1.044318 | 0.441015 |
| SL004714 | Leukemia inhibitory factor receptor | LIF sR | P42702 | 3977 | LIFR | -1.072023 | 0.441776 |
| SL006921 | [Pyruvate dehydrogenase (acetyl-transferring)] kinase isozyme 1, mitochondrial | PDK1 | Q15118 | 5163 | PDK1 | -1.096628 | 0.444964 |
| SL008122 | Dual specificity protein phosphatase 3 | DUS3 | P51452 | 1845 | DUSP3 | -1.208651 | 0.445097 |
| SL006918 | Mitogen-activated protein kinase 1 | MK01 | P28482 | 5594 | MAPK1 | -1.144614 | 0.445111 |
| SL000249 | Alpha-1-antitrypsin | a1-Antitrypsin | P01009 | 5265 | SERPINA1 | -1.074953 | 0.44614 |
| SL004457 | Glia-derived nexin | Protease nexin I | P07093 | 5270 | SERPINE2 | 1.069856 | 0.446355 |
| SL007237 | Dual specificity mitogen-activated protein kinase kinase 4 | MP2K4 | P45985 | 6416 | MAP2K4 | -1.084937 | 0.446536 |
| SL003200 | Angiopoietin-1 receptor, soluble | sTie-2 | Q02763 | 7010 | TEK | -1.07301 | 0.447359 |
| SL000001 | Mucin-1 | MUC1 | P15941 | 4582 | MUC1 | -1.078692 | 0.447422 |
| HCE003167 | HCE003167 | HCE003167 | HCE003167 | HCE003167 | HCE003167 | -1.054874 | 0.448378 |
| SL002722 | ADP-ribosyl cyclase/cyclic ADP-ribose hydrolase 1 | CD38 | P28907 | 952 | CD38 | 1.597164 | 0.448421 |
| SL003300 | C-C motif chemokine 16 | HCC-4 | O15467 | 6360 | CCL16 | 1.164217 | 0.449637 |
| SL008039 | Alcohol dehydrogenase [NADP(+)] | AK1A1 | P14550 | 10327 | AKR1A1 | 1.156555 | 0.449762 |
| SL004156 | Tumor necrosis factor receptor superfamily member 10A | TRAIL R1 | O00220 | 8797 | TNFRSF10A | 1.077669 | 0.450373 |
| SL007033 | Latent-transforming growth factor beta-binding protein 4 | LTBP4 | Q8N2S1 | 8425 | LTBP4 | 1.097137 | 0.451398 |
| SL005102 | Sex hormone-binding globulin | SHBG | P04278 | 6462 | SHBG | 1.100017 | 0.452881 |
| SL010612 | Dickkopf-related protein 4 | Dkk-4 | Q9UBT3 | 27121 | DKK4 | -1.055327 | 0.455388 |
| SL005153 | Ciliary neurotrophic factor receptor subunit alpha | CNTFR alpha | P26992 | 1271 | CNTFR | -1.077544 | 0.457146 |
| SL003198 | Tenascin | Tenascin | P24821 | 3371 | TNC | 1.088676 | 0.458054 |
| SL005358 | Phosphatidylethanolamine-binding protein 1 | prostatic binding protein | P30086 | 5037 | PEBP1 | 1.162678 | 0.458194 |
| SL014470 | Breast cancer anti-estrogen resistance protein 3 | BCAR3 | O75815 | 8412 | BCAR3 | 1.037695 | 0.458933 |
| SL005687 | Eukaryotic translation initiation factor 5A-1 | eIF-5A-1 | P63241 | 1984 | EIF5A | -1.128243 | 0.45939 |
| SL003744 | Galectin-3 | Galectin-3 | P17931 | 3958 | LGALS3 | -1.080691 | 0.459449 |
| SL000481 | Interleukin-5 | IL-5 | P05113 | 3567 | IL5 | -1.142968 | 0.460472 |
| SL000605 | Ubiquitin+1, truncated mutation for UbB | Ubiquitin+1 | P62979 | 6233 | RPS27A | 1.079624 | 0.461007 |
| SL003201 | Vascular endothelial growth factor receptor 2 | VEGF sR2 | P35968 | 3791 | KDR | -1.067967 | 0.461238 |
| SL000347 | Corticosteroid-binding globulin | CBG | P08185 | 866 | SERPINA6 | -1.162782 | 0.462695 |
| SL010373 | Immunoglobulin alpha Fc receptor | FCAR | P24071 | 2204 | FCAR | 1.042956 | 0.463215 |
| SL004938 | Calcium/calmodulin-dependent protein kinase kinase 1 | CaMKK alpha | Q8N5S9 | 84254 | CAMKK1 | -1.047023 | 0.464083 |
| SL000539 | Glucose-6-phosphate isomerase | PHI | P06744 | 2821 | GPI | 1.124507 | 0.464258 |
| SL001691 | Fibroblast growth factor 7 | FGF7 | P21781 | 2252 | FGF7 | -1.110323 | 0.464296 |
| SL003184 | Leptin receptor | sLeptin R | P48357 | 3953 | LEPR | -1.128978 | 0.464669 |
| SL000360 | Coagulation Factor X | Coagulation Factor X | P00742 | 2159 | F10 | -1.031283 | 0.465092 |
| SL004230 | Microtubule-associated protein tau | tau | P10636 | 4137 | MAPT | 1.16945 | 0.465601 |
| SL011808 | Copine-1 | CPNE1 | Q99829 | 8904 | CPNE1 | -1.193633 | 0.466966 |
| SL004853 | ICOS ligand | B7-H2 | O75144 | 23308 | ICOSLG | -1.181969 | 0.467502 |
| SL003280 | High mobility group protein B1 | HMG-1 | P09429 | 3146 | HMGB1 | 1.091728 | 0.469599 |
| SL006629 | NAD-dependent protein deacetylase sirtuin-2 | SIRT2 | Q8IXJ6 | 22933 | SIRT2 | 1.06972 | 0.470625 |
| SL011768 | NudC domain-containing protein 3 | NUDC3 | Q8IVD9 | 23386 | NUDCD3 | 1.267534 | 0.470838 |
| SL007108 | Interferon regulatory factor 1 | IRF1 | P10914 | 3659 | IRF1 | -1.004943 | 0.471258 |
| SL011888 | SPARC-related modular calcium-binding protein 1 | SMOC1 | Q9H4F8 | 64093 | SMOC1 | 1.041965 | 0.471333 |
| SL007274 | Ephrin-B2 | EFNB2 | P52799 | 1948 | EFNB2 | 1.104497 | 0.471606 |
| SL003080 | Macrophage migration inhibitory factor | MIF | P14174 | 4282 | MIF | -1.097248 | 0.473311 |
| SL005222 | Scavenger receptor class F member 2 | SREC-II | Q96GP6 | 91179 | SCARF2 | 1.068608 | 0.474315 |
| SL005169 | Intercellular adhesion molecule 5 | sICAM-5 | Q9UMF0 | 7087 | ICAM5 | 1.035778 | 0.476672 |
| SL006480 | Trypsin-3 | TRY3 | P35030 | 5646 | PRSS3 | -1.068438 | 0.478549 |
| SL002823 | L-Selectin | sL-Selectin | P14151 | 6402 | SELL | 1.069576 | 0.481808 |
| SL000277 | Apolipoprotein E (isoform E2) | Apo E2 | P02649 | 348 | APOE | -1.031727 | 0.481933 |
| SL007207 | Thrombospondin-4 | TSP4 | P35443 | 7060 | THBS4 | 1.08747 | 0.482019 |
| SL000021 | Insulin | Insulin | P01308 | 3630 | INS | -1.081107 | 0.485283 |
| SL000272 | Antithrombin-III | Antithrombin III | P01008 | 462 | SERPINC1 | 1.0125 | 0.485535 |
| SL014096 | Leucine-rich repeat transmembrane neuronal protein 1 | LRRT1 | Q86UE6 | 347730 | LRRTM1 | 1.085431 | 0.485597 |
| SL003918 | Kallikrein-13 | kallikrein 13 | Q9UKR3 | 26085 | KLK13 | -1.122767 | 0.485749 |
| SL004827 | Protein S100-A6 | S100A6 | P06703 | 6277 | S100A6 | 1.064325 | 0.48703 |
| SL007373 | Peptidyl-prolyl cis-trans isomerase D | PPID | Q08752 | 5481 | PPID | -1.159099 | 0.488009 |
| SL010512 | PIK3CA/PIK3R1 | PIK3CA/PIK3R1 | P42336 P27986 | 5290 5295 | PIK3CA PIK3R1 | -1.135433 | 0.488477 |
| SL006777 | Fetuin-B | FETUB | Q9UGM5 | 26998 | FETUB | 1.128036 | 0.489094 |
| SL000024 | Tissue Factor | TF | P13726 | 2152 | F3 | 1.06041 | 0.490686 |
| SL011530 | Dynein light chain roadblock-type 1 | DLRB1 | Q9NP97 | 83658 | DYNLRB1 | -1.128034 | 0.491596 |
| SL004739 | Inter-alpha-trypsin inhibitor heavy chain H4 | ITI heavy chain H4 | Q14624 | 3700 | ITIH4 | 1.045963 | 0.493851 |
| SL003915 | Kallikrein-8 | kallikrein 8 | O60259 | 11202 | KLK8 | 1.098008 | 0.49542 |
| SL003728 | Baculoviral IAP repeat-containing protein 3 | cIAP-2 | Q13489 | 330 | BIRC3 | -1.451279 | 0.495871 |
| SL003324 | Coagulation factor Xa | Coagulation Factor Xa | P00742 | 2159 | F10 | -1.037931 | 0.496364 |
| SL004396 | Signal transducer and activator of transcription 1-alpha/beta | STAT1 | P42224 | 6772 | STAT1 | -1.19788 | 0.496769 |
| SL007003 | Desert hedgehog protein N-product | DHH | O43323 | 50846 | DHH | -1.08706 | 0.497771 |
| SL000038 | C-C motif chemokine 2 | MCP-1 | P13500 | 6347 | CCL2 | -1.6092 | 0.497936 |
| SL002542 | GTPase KRas | K-ras | P01116 | 3845 | KRAS | -1.022555 | 0.498323 |
| SL000358 | Coagulation factor VII | Coagulation Factor VII | P08709 | 2155 | F7 | -1.011913 | 0.498442 |
| SL005212 | Prolactin receptor | Prolactin Receptor | P16471 | 5618 | PRLR | 1.05104 | 0.498559 |
| SL006915 | Interleukin-27 | IL-27 | Q8NEV9 Q14213 | 246778 10148 | IL27 EBI3 | 1.04058 | 0.498912 |
| SL005184 | Interleukin-23 | IL-23 | P29460, Q9NPF7 | 3593 51561 | IL12B IL23A | -1.112864 | 0.500717 |
| SL017529 | Kelch-like ECH-associated protein 1 | KEAP1 | Q14145 | 9817 | KEAP1 | -3.213589 | 0.501606 |
| SL000836 | Hemoglobin | Hemoglobin | P69905, P68871 | 3039 3043 | HBA1 HBB | 1.15152 | 0.502345 |
| SL004676 | Insulin-like growth factor-binding protein 5 | IGFBP-5 | P24593 | 3488 | IGFBP5 | -1.055332 | 0.502956 |
| SL001888 | Antileukoproteinase | SLPI | P03973 | 6590 | SLPI | 1.08539 | 0.503152 |
| SL004765 | MAP kinase-activated protein kinase 3 | MAPKAPK3 | Q16644 | 7867 | MAPKAPK3 | -1.107868 | 0.503183 |
| SL003711 | Caspase-3 | Caspase-3 | P42574 | 836 | CASP3 | -1.196872 | 0.503343 |
| SL003974 | Activated Protein C | Activated Protein C | P04070 | 5624 | PROC | -1.058247 | 0.504175 |
| SL005199 | MHC class I polypeptide-related sequence A | MICA | Q29983 | 4276 | MICA | -1.071362 | 0.504673 |
| SL017106 | Interferon alpha-10 | IFN10 | P01566 | 3446 | IFNA10 | 1.400638 | 0.505202 |
| SL000497 | Laminin | Laminin | P25391, P07942, P11047 | 284217 3912 3915 | LAMA1 LAMB1 LAMC1 | -1.057674 | 0.506193 |
| SL018900 | Formimidoyltransferase-cyclodeaminase | FTCD | O95954 | 10841 | FTCD | 2.158089 | 0.507264 |
| SL005629 | N-acetyl-D-glucosamine kinase | NAGK | Q9UJ70 | 55577 | NAGK | 1.135991 | 0.507425 |
| SL016969 | Cysteine and glycine-rich protein 3 | CSRP3 | P50461 | 8048 | CSRP3 | 1.112328 | 0.508056 |
| SL007888 | Cystatin-S | Cystatin-S | P01036 | 1472 | CST4 | -1.510693 | 0.508847 |
| SL017610 | Gro-beta/gamma | Gro-b/g | P19876 P19875 | NaN | CXCL3 CXCL2 | -1.016948 | 0.509006 |
| SL000598 | Thrombopoietin | Tpo | P40225 | 7066 | THPO | 1.077415 | 0.509143 |
| SL011069 | Serine protease 27 | Marapsin | Q9BQR3 | 83886 | PRSS27 | -1.19928 | 0.509578 |
| SL004914 | Inorganic pyrophosphatase | PPase | Q15181 | 5464 | PPA1 | 1.265714 | 0.509676 |
| SL004347 | Interleukin-22 | IL-22 | Q9GZX6 | 50616 | IL22 | -1.130353 | 0.510152 |
| SL000551 | Protein kinase C alpha type | PKC-A | P17252 | 5578 | PRKCA | -1.238422 | 0.510271 |
| SL003307 | Cytokine receptor common subunit gamma | IL-2 sRg | P31785 | 3561 | IL2RG | -1.125724 | 0.510612 |
| SL004750 | ATP-dependent RNA helicase DDX19B | DEAD-box protein 19B | Q9UMR2 | 11269 | DDX19B | -1.088557 | 0.510705 |
| SL011549 | AT-rich interactive domain-containing protein 3A | ARI3A | Q99856 | 1820 | ARID3A | -1.400145 | 0.513208 |
| SL000658 | Growth arrest-specific protein 1 | GAS1 | P54826 | 2619 | GAS1 | 1.064543 | 0.515396 |
| SL001905 | Mesothelin | Mesothelin | Q13421 | 10232 | MSLN | -1.119697 | 0.515671 |
| SL005675 | ATP synthase subunit beta, mitochondrial | ATP synthase beta chain | P06576 | 506 | ATP5B | 1.188435 | 0.516651 |
| SL000271 | Angiotensinogen | Angiotensinogen | P01019 | 183 | AGT | 1.078276 | 0.517178 |
| SL000645 | Stromelysin-2 | MMP-10 | P09238 | 4319 | MMP10 | 1.169757 | 0.517573 |
| SL017188 | Glycogen synthase kinase-3 alpha/beta | GSK-3 alpha/beta | P49840 P49841 | 2931 2932 | GSK3A GSK3B | -1.171603 | 0.517581 |
| SL010521 | Tyrosine-protein kinase BTK | BTK | Q06187 | 695 | BTK | -1.233125 | 0.519291 |
| SL003722 | RAC-beta serine/threonine-protein kinase | PKB beta | P31751 | 208 | AKT2 | -1.14917 | 0.519985 |
| SL004348 | Interferon lambda-1 | IFN-lambda 1 | Q8IU54 | 282618 | IFNL1 | -1.095978 | 0.520255 |
| SL000088 | Transforming growth factor beta-2 | TGF-b2 | P61812 | 7042 | TGFB2 | 1.031364 | 0.520458 |
| SL000587 | Thyroglobulin | Thyroglobulin | P01266 | 7038 | TG | 1.040849 | 0.521946 |
| SL005168 | Growth hormone receptor | Growth hormone receptor | P10912 | 2690 | GHR | -1.049308 | 0.521959 |
| SL004330 | B-cell receptor CD22 | CD22 | P20273 | 933 | CD22 | 1.025629 | 0.52212 |
| SL010379 | Semaphorin-3A | Semaphorin 3A | Q14563 | 10371 | SEMA3A | -1.027858 | 0.522354 |
| SL003919 | Kallikrein-14 | kallikrein 14 | Q9P0G3 | 43847 | KLK14 | -1.045053 | 0.522355 |
| SL001718 | Interleukin-13 | IL-13 | P35225 | 3596 | IL13 | -1.076655 | 0.522467 |
| SL004716 | Mitogen-activated protein kinase 9 | JNK2 | P45984 | 5601 | MAPK9 | 1.028845 | 0.523317 |
| SL006705 | Prefoldin subunit 5 | PFD5 | Q99471 | 5204 | PFDN5 | 1.061498 | 0.523533 |
| SL012561 | Regenerating islet-derived protein 4 | REG4 | Q9BYZ8 | 83998 | REG4 | -1.099079 | 0.526644 |
| SL001791 | Osteocalcin | Osteocalcin | P02818 | 632 | BGLAP | -1.053281 | 0.526693 |
| SL011202 | Alpha-soluble NSF attachment protein | SNAA | P54920 | 8775 | NAPA | -1.133884 | 0.527429 |
| SL000299 | Fibroblast growth factor 1 | b-ECGF | P05230 | 2246 | FGF1 | -1.012537 | 0.527794 |
| SL010504 | MAP kinase-activated protein kinase 5 | MAPK5 | Q8IW41 | 8550 | MAPKAPK5 | 1.396248 | 0.528807 |
| SL006713 | Collectin-11 | Collectin Kidney 1 | Q9BWP8 | 78989 | COLEC11 | 1.087166 | 0.530146 |
| SL002792 | beta-adrenergic receptor kinase 1 | BARK1 | P25098 | 156 | ADRBK1 | -1.174508 | 0.530211 |
| SL004141 | Ephrin-A5 | Ephrin-A5 | P52803 | 1946 | EFNA5 | 1.054052 | 0.530813 |
| SL001880 | Interferon beta | IFN-b | P01574 | 3456 | IFNB1 | 1.120216 | 0.531559 |
| SL020171 | HERV-H LTR-associating protein 2 | HHLA2 | Q9UM44 | NaN | HHLA2 | -1.858114 | 0.531628 |
| SL005195 | Lymphocyte activation gene 3 protein | LAG-3 | P18627 | 3902 | LAG3 | -1.066432 | 0.53164 |
| SL004795 | 3-hydroxyacyl-CoA dehydrogenase type-2 | ERAB | Q99714 | 3028 | HSD17B10 | -1.263148 | 0.533279 |
| SL005220 | Sonic hedgehog protein | Sonic Hedgehog | Q15465 | 6469 | SHH | -1.054685 | 0.533929 |
| SL004649 | Tumor necrosis factor ligand superfamily member 4 | OX40 Ligand | P23510 | 7292 | TNFSF4 | 1.084983 | 0.53527 |
| SL000448 | Heat shock protein beta-1 | HSP 27 | P04792 | 3315 | HSPB1 | -1.189092 | 0.535292 |
| SL004346 | Interleukin-20 | IL-20 | Q9NYY1 | 50604 | IL20 | 1.024135 | 0.536201 |
| SL002704 | Pleiotrophin | PTN | P21246 | 5764 | PTN | 1.030211 | 0.537373 |
| SL008936 | Neural cell adhesion molecule L1-like protein | CHL1 | O00533 | 10752 | CHL1 | -1.071179 | 0.537403 |
| SL005185 | Interleukin-23 receptor | IL-23 R | Q5VWK5 | 149233 | IL23R | -1.068253 | 0.537834 |
| SL017189 | Kininogen-1 | Kininogen, HMW | P01042 | 3827 | KNG1 | 1.08784 | 0.537965 |
| SL004515 | Peptidoglycan recognition protein 1 | PGRP-S | O75594 | 8993 | PGLYRP1 | -2.131452 | 0.540063 |
| SL000396 | Cytochrome c | Cytochrome c | P99999 | 54205 | CYCS | -1.066204 | 0.541766 |
| SL004692 | Cytokine receptor-like factor 1:Cardiotrophin-like cytokine factor 1 Complex | CLF-1/CLC Complex | O75462 Q9UBD9 | 9244 23529 | CRLF1 CLCF1 | 1.145928 | 0.543576 |
| SL004078 | Bone morphogenetic protein 7 | BMP-7 | P18075 | 655 | BMP7 | -1.102045 | 0.544711 |
| SL001717 | Interleukin-10 | IL-10 | P22301 | 3586 | IL10 | -1.144521 | 0.54539 |
| SL008822 | Adhesion G protein-coupled receptor E2 | EMR2 | Q9UHX3 | 30817 | ADGRE2 | -1.036311 | 0.545636 |
| SL004484 | Pulmonary surfactant-associated protein D | SP-D | P35247 | 6441 | SFTPD | 1.197932 | 0.546787 |
| SL000542 | Integrin alpha-IIb: beta-3 complex | gpIIbIIIa | P08514 P05106 | 3674 3690 | ITGA2B ITGB3 | 1.050303 | 0.548136 |
| SL007620 | Interleukin-12 receptor subunit beta-2 | IL-12 RB2 | Q99665 | 3595 | IL12RB2 | 1.0142 | 0.548404 |
| SL005694 | Peroxiredoxin-6 | Peroxiredoxin-6 | P30041 | 9588 | PRDX6 | -1.045231 | 0.548498 |
| SL003653 | Phosphoglycerate kinase 1 | phosphoglycerate kinase 1 | P00558 | 5230 | PGK1 | -1.115392 | 0.549343 |
| SL002650 | Pyruvate kinase PKM | M2-PK | P14618 | 5315 | PKM2 | -1.199895 | 0.550037 |
| SL003331 | Matrix metalloproteinase-16 | MMP-16 | P51512 | 4325 | MMP16 | 1.053116 | 0.550119 |
| SL004269 | Appetite-regulating hormone | ghrelin | Q9UBU3 | 51738 | GHRL | 1.001759 | 0.550199 |
| SL008466 | 3-hydroxyanthranilate 3,4-dioxygenase | 3HAO | P46952 | 23498 | HAAO | 1.008869 | 0.550545 |
| SL006914 | Glucocorticoid receptor | Glucocorticoid receptor | P04150 | 2908 | NR3C1 | -1.267121 | 0.550609 |
| SL010503 | MAP kinase-activated protein kinase 2 | MAPK2 | P49137 | 9261 | MAPKAPK2 | -1.181671 | 0.551536 |
| SL006522 | Galectin-3-binding protein | LG3BP | Q08380 | 3959 | LGALS3BP | 1.091824 | 0.552695 |
| SL000324 | Complement component C8 | C8 | P07357,P07358,P07360 | 731 732 733 | C8A C8B C8G | -1.02689 | 0.554757 |
| SL010328 | Mediator of RNA polymerase II transcription subunit 1 | MED-1 | Q15648 | 5469 | MED1 | -1.041217 | 0.555337 |
| SL012395 | Netrin-1 | NET1 | O95631 | 9423 | NTN1 | -1.133392 | 0.555346 |
| SL005181 | Interleukin-20 receptor subunit alpha | IL-20 Ra | Q9UHF4 | 53832 | IL20RA | 1.441555 | 0.556754 |
| SL011232 | Hsp90 co-chaperone Cdc37 | CDC37 | Q16543 | 11140 | CDC37 | -1.228345 | 0.558221 |
| SL000592 | Metalloproteinase inhibitor 2 | TIMP-2 | P16035 | 7077 | TIMP2 | 1.055502 | 0.558922 |
| SL004637 | Macrophage-stimulating protein receptor | MSP R | Q04912 | 4486 | MST1R | 1.03136 | 0.559496 |
| SL017528 | Ribonuclease H1 | RNase H1 | O60930 | 246243 | RNASEH1 | -1.655942 | 0.560102 |
| SL000104 | Apoptosis regulator Bcl-2 | Bcl-2 | P10415 | 596 | BCL2 | 1.034915 | 0.561542 |
| SL005213 | Tumor necrosis factor receptor superfamily member 19L | RELT | Q969Z4 | 84957 | RELT | 1.083519 | 0.562155 |
| SL004866 | Carbonic anhydrase 1 | Carbonic anhydrase I | P00915 | 759 | CA1 | 1.096913 | 0.56341 |
| SL013872 | Low-density lipoprotein receptor-related protein 1B | LRP1B | Q9NZR2 | 53353 | LRP1B | -1.032964 | 0.564437 |
| SL006114 | Tyrosine-protein kinase transmembrane receptor ROR1 | ROR1 | Q01973 | 4919 | ROR1 | -1.062974 | 0.565023 |
| SL010368 | Alpha-L-iduronidase | IDUA | P35475 | 3425 | IDUA | -1.020987 | 0.568697 |
| SL007173 | Granulins | GRN | P28799 | 2896 | GRN | 1.037117 | 0.569659 |
| SL000076 | Cyclin-dependent kinase inhibitor 1B | p27Kip1 | P46527 | 1027 | CDKN1B | -1.063122 | 0.570869 |
| SL013928 | Peptidyl-prolyl cis-trans isomerase E | PPIE | Q9UNP9 | 10450 | PPIE | -1.110543 | 0.570926 |
| SL010388 | Trypsin-2 | Trypsin 2 | P07478 | 5645 | PRSS2 | -1.542533 | 0.571036 |
| SL010616 | Tyrosine-protein phosphatase non-receptor type 11 | SHP-2 | Q06124 | 5781 | PTPN11 | 1.065898 | 0.571249 |
| SL000414 | Complement factor B | Factor B | P00751 | 629 | CFB | -1.030215 | 0.57268 |
| SL003166 | CD166 antigen | ALCAM | Q13740 | 214 | ALCAM | 1.048489 | 0.573656 |
| SL001998 | Tissue factor pathway inhibitor | TFPI | P10646 | 7035 | TFPI | 1.042175 | 0.574226 |
| SL007631 | Sclerostin | SOST | Q9BQB4 | 50964 | SOST | 1.115293 | 0.574628 |
| SL004125 | Insulin receptor | IR | P06213 | 3643 | INSR | -1.065635 | 0.575303 |
| SL004359 | Neurotrophin-3 | Neurotrophin-3 | P20783 | 4908 | NTF3 | 1.050436 | 0.575768 |
| SL000027 | Prostaglandin G/H synthase 2 | COX-2 | P35354 | 5743 | PTGS2 | 1.076662 | 0.576014 |
| SL004751 | Heme oxygenase 2 | HO-2 | P30519 | 3163 | HMOX2 | 1.045653 | 0.576473 |
| SL002688 | Osteopontin | Osteopontin | P10451 | 6696 | SPP1 | 1.060073 | 0.576519 |
| SL000560 | P-selectin | P-Selectin | P16109 | 6403 | SELP | 1.032253 | 0.578474 |
| SL003308 | Interleukin-4 receptor subunit alpha | IL-4 sR | P24394 | 3566 | IL4R | -1.095851 | 0.579432 |
| SL000406 | Eotaxin | Eotaxin | P51671 | 6356 | CCL11 | -1.117621 | 0.580482 |
| SL005200 | MHC class I polypeptide-related sequence B | MICB | Q29980 | 4277 | MICB | -1.158368 | 0.582035 |
| SL000525 | Matrilysin | MMP-7 | P09237 | 4316 | MMP7 | 1.057427 | 0.582526 |
| SL002528 | Phospholipase A2, membrane associated | NPS-PLA2 | P14555 | 5320 | PLA2G2A | 1.037051 | 0.582646 |
| SL000313 | C3a anaphylatoxin | C3a | P01024 | 718 | C3 | -1.064976 | 0.582693 |
| SL010372 | Enteropeptidase | Enterokinase | P98073 | 5651 | TMPRSS15 | 1.028757 | 0.582999 |
| SL003309 | Lipopolysaccharide-binding protein | LBP | P18428 | 3929 | LBP | 1.016621 | 0.583103 |
| SL005231 | Netrin receptor UNC5D | UNC5H4 | Q6UXZ4 | 137970 | UNC5D | 1.070187 | 0.583461 |
| SL000461 | Immunoglobulin E | IgE | P01854 | 3497 50802 3535 | IGHE IGK@ IGL@ | 1.666662 | 0.584442 |
| SL005210 | Neuronal cell adhesion molecule | Nr-CAM | Q92823 | 4897 | NRCAM | 1.057997 | 0.585065 |
| SL005160 | Endothelial cell-selective adhesion molecule | ESAM | Q96AP7 | 90952 | ESAM | 1.04156 | 0.585333 |
| SL007272 | Hexokinase-2 | HXK2 | P52789 | 3099 | HK2 | -1.030228 | 0.585536 |
| SL008072 | Collagen alpha-1(VIII) chain | CO8A1 | P27658 | 1295 | COL8A1 | -1.766545 | 0.585561 |
| SL011071 | Platelet-derived growth factor C | PDGF-CC | Q9NRA1 | 56034 | PDGFC | -1.034305 | 0.585733 |
| SL008865 | Proteasome subunit alpha type-2 | PSA2 | P25787 | 5683 | PSMA2 | 1.033376 | 0.586425 |
| SL006197 | Mitochondrial import inner membrane translocase subunit TIM14 | DnaJ homolog | Q96DA6 | 131118 | DNAJC19 | 1.027338 | 0.588203 |
| SL004511 | Bactericidal permeability-increasing protein | BPI | P17213 | 671 | BPI | 1.053896 | 0.589537 |
| SL000468 | Immunoglobulin M | IgM | P01871 | 3507 3512 50802 3535 | IGHM IGJ IGK@ IGL@ | 1.188374 | 0.589657 |
| SL000550 | Plasma serine protease inhibitor | PCI | P05154 | 5104 | SERPINA5 | -1.003123 | 0.589821 |
| SL005392 | Arylsulfatase A | Arylsulfatase A | P15289 | 410 | ARSA | 1.098832 | 0.590541 |
| SL000254 | Serum albumin | Albumin | P02768 | 213 | ALB | -1.097359 | 0.591287 |
| SL014071 | Leucine-rich repeat transmembrane protein FLRT1 | FLRT1 | Q9NZU1 | 23769 | FLRT1 | -1.040229 | 0.592252 |
| SL004635 | Tumor necrosis factor ligand superfamily member 8 | CD30 Ligand | P32971 | 944 | TNFSF8 | -1.067691 | 0.593362 |
| SL000308 | Plasma protease C1 inhibitor | C1-Esterase Inhibitor | P05155 | 710 | SERPING1 | -1.055463 | 0.593873 |
| SL003643 | Glutathione S-transferase P | Glutathione S-transferase Pi | P09211 | 2950 | GSTP1 | 1.034213 | 0.594147 |
| SL002662 | Coagulation Factor XI | Coagulation Factor XI | P03951 | 2160 | F11 | 1.007937 | 0.595238 |
| SL001947 | Melanoma-derived growth regulatory protein | MIA | Q16674 | 8190 | MIA | -1.078098 | 0.595872 |
| SL003658 | Aflatoxin B1 aldehyde reductase member 2 | Aflatoxin B1 aldehyde reductase | O43488 | 8574 | AKR7A2 | -1.16338 | 0.599002 |
| SL001795 | Interleukin-1 beta | IL-1b | P01584 | 3553 | IL1B | 1.042957 | 0.600123 |
| SL004781 | Tyrosine-protein kinase CSK | CSK | P41240 | 1445 | CSK | -1.170198 | 0.600362 |
| SL008414 | Ephrin type-B receptor 6 | EphB6 | O15197 | 2051 | EPHB6 | 1.04378 | 0.601126 |
| SL004654 | Granzyme H | Granzyme H | P20718 | 2999 | GZMH | 1.016655 | 0.604012 |
| SL004901 | Protein disulfide-isomerase | Protein disulfide-isomerase | P07237 | 5034 | P4HB | -1.052801 | 0.604979 |
| SL006406 | Plexin-C1 | PLXC1 | O60486 | 10154 | PLXNC1 | -1.048265 | 0.607066 |
| SL000382 | Creatine kinase M-type:Creatine kinase B-type heterodimer | CK-MB | P12277 P06732 | 1152 1158 | CKB CKM | -1.112055 | 0.608046 |
| SL004301 | X-ray repair cross-complementing protein 6 | Ku70 | P12956 | 2547 | XRCC6 | 1.061699 | 0.609547 |
| SL010927 | Importin subunit beta-1 | IMB1 | Q14974 | 3837 | KPNB1 | 1.066734 | 0.609992 |
| SL008835 | Asialoglycoprotein receptor 1 | ASGR1 | P07306 | 432 | ASGR1 | 1.163791 | 0.610288 |
| SL006189 | Kinesin-like protein KIF23 | KIF23 | Q02241 | 9493 | KIF23 | 1.06551 | 0.610405 |
| SL004672 | Tumor necrosis factor receptor superfamily member 17 | BCMA | Q02223 | 608 | TNFRSF17 | -1.074381 | 0.610528 |
| SL011532 | Persulfide dioxygenase ETHE1, mitochondrial | ETHE1 | O95571 | 23474 | ETHE1 | -1.032365 | 0.612716 |
| SL016550 | Casein kinase II 2-alpha:2-beta heterotetramer | CK2-A1:B | P68400 P67870 | 1457 1460 | CSNK2A1 CSNK2B | -1.110319 | 0.613095 |
| SL004133 | Bone morphogenetic protein receptor type-2 | BMP RII | Q13873 | 659 | BMPR2 | -1.048121 | 0.613133 |
| SL008588 | SLAM family member 5 | SLAF5 | Q9UIB8 | 8832 | CD84 | 1.051074 | 0.613947 |
| SL004137 | Ephrin type-A receptor 1 | EphA1 | P21709 | 2041 | EPHA1 | -1.066618 | 0.614759 |
| SL011631 | Nascent polypeptide-associated complex subunit alpha | NACA | Q13765 | 4666 | NACA | 1.044823 | 0.614886 |
| SL004337 | Fibroblast growth factor 19 | FGF-19 | O95750 | 9965 | FGF19 | -1.021438 | 0.614923 |
| SL001720 | Vascular cell adhesion protein 1 | VCAM-1 | P19320 | 7412 | VCAM1 | 1.110873 | 0.616126 |
| SL016567 | Mitogen-activated protein kinase kinase kinase 7:TGF-beta-activated kinase 1 and MAP3K7-binding protein 1 fusion | TAK1-TAB1 | O43318 Q15750 | 6885 10454 | MAP3K7 TAB1 | -1.064444 | 0.616296 |
| SL007673 | Netrin-4 | NET4 | Q9HB63 | 59277 | NTN4 | -1.065313 | 0.616495 |
| SL007311 | Synaptosomal-associated protein 25 | SNP25 | P60880 | 6616 | SNAP25 | 1.036213 | 0.617093 |
| SL003305 | Interleukin-2 receptor subunit alpha | IL-2 sRa | P01589 | 3559 | IL2RA | -1.004135 | 0.617527 |
| SL007756 | Growth/differentiation factor 2 | GDF2 | Q9UK05 | 2658 | GDF2 | -1.03843 | 0.617649 |
| SL004925 | Anterior gradient protein 2 homolog | AGR2 | O95994 | 10551 | AGR2 | -1.021473 | 0.618533 |
| SL002695 | Cytosolic non-specific dipeptidase | Glutamate carboxypeptidase | Q96KP4 | 55748 | CNDP2 | 1.027152 | 0.618792 |
| SL000427 | Fractalkine | Fractalkine/CX3CL-1 | P78423 | 6376 | CX3CL1 | 1.036824 | 0.619007 |
| SL004605 | 40S ribosomal protein SA | 40S ribosomal protein SA | P08865 | 3921 | RPSA | -1.043672 | 0.621555 |
| SL004415 | Angiotensin-converting enzyme 2 | ACE2 | Q9BYF1 | 59272 | ACE2 | -1.05007 | 0.622761 |
| SL011630 | Seizure 6-like protein 2 | SE6L2 | Q6UXD5 | 26470 | SEZ6L2 | -1.082275 | 0.623065 |
| SL005250 | Chymase | Chymase | P23946 | 1215 | CMA1 | 1.050397 | 0.623857 |
| SL008381 | Cathepsin F | CATF | Q9UBX1 | 8722 | CTSF | -1.016509 | 0.625189 |
| SL000003 | Angiogenin | Angiogenin | P03950 | 283 | ANG | -1.005653 | 0.627652 |
| SL002755 | Pappalysin-1 | PAPP-A | Q13219 | 5069 | PAPPA | -1.068014 | 0.628108 |
| SL000526 | Neutrophil collagenase | MMP-8 | P22894 | 4317 | MMP8 | 1.11313 | 0.628471 |
| SL007471 | Collectin-12 | COLEC12 | Q5KU26 | 81035 | COLEC12 | 1.026298 | 0.628802 |
| SL010465 | Matrilin-2 | MATN2 | O00339 | 4147 | MATN2 | 1.038868 | 0.630952 |
| SL003193 | C-C motif chemokine 21 | 6Ckine | O00585 | 6366 | CCL21 | -1.011177 | 0.632348 |
| SL000470 | Interleukin-11 | IL-11 | P20809 | 3589 | IL11 | -1.000633 | 0.63261 |
| SL003199 | Tyrosine-protein kinase receptor Tie-1, soluble | sTie-1 | P35590 | 7075 | TIE1 | 1.038825 | 0.632706 |
| SL003177 | Intercellular adhesion molecule 2 | sICAM-2 | P13598 | 3384 | ICAM2 | -1.044099 | 0.632954 |
| SL000009 | Receptor tyrosine-protein kinase erbB-2 | ERBB2 | P04626 | 2064 | ERBB2 | -1.023559 | 0.633532 |
| SL014294 | Ephrin type-A receptor 10 | EPHAA | Q5JZY3 | 284656 | EPHA10 | 1.030944 | 0.63373 |
| SL010451 | Cryptic protein | CFC1 | P0CG37 | 55997 | CFC1 | -1.030481 | 0.634184 |
| SL004856 | Desmoglein-1 | Desmoglein-1 | Q02413 | 1828 | DSG1 | -1.021182 | 0.63604 |
| SL010491 | Calcium/calmodulin-dependent protein kinase type II subunit alpha | CAMK2A | Q9UQM7 | 815 | CAMK2A | -1.072906 | 0.637513 |
| SL005493 | Serum paraoxonase/arylesterase 1 | paraoxonase 1 | P27169 | 5444 | PON1 | -1.026105 | 0.637639 |
| SL000136 | Amphiregulin | AREG | P15514 | 374 | AREG | -1.103356 | 0.637679 |
| SL008360 | Leucine-rich repeat transmembrane protein FLRT2 | FLRT2 | O43155 | 23768 | FLRT2 | 1.041085 | 0.638489 |
| SL001731 | Gamma-enolase | NSE | P09104 | 2026 | ENO2 | -1.036267 | 0.638577 |
| SL008157 | Ubiquitin-conjugating enzyme E2 L3 | UB2L3 | P68036 | 7332 | UBE2L3 | -1.078848 | 0.639091 |
| SL000458 | Interferon gamma receptor 1 | IFN-g R1 | P15260 | 3459 | IFNGR1 | 1.028548 | 0.63982 |
| SL006912 | Tyrosine-protein kinase Fgr | FGR | P09769 | 2268 | FGR | -1.141502 | 0.640849 |
| SL004686 | Tumor necrosis factor ligand superfamily member 15 | TNFSF15 | O95150 | 9966 | TNFSF15 | 1.035633 | 0.6412 |
| SL005372 | Sorting nexin-4 | Sorting nexin 4 | O95219 | 8723 | SNX4 | -1.189295 | 0.641868 |
| SL005161 | Fibroblast growth factor 12 | FGF-12 | P61328 | 2257 | FGF12 | 1.043556 | 0.641976 |
| SL003066 | Pigment epithelium-derived factor | PEDF | P36955 | 5176 | SERPINF1 | 1.034215 | 0.642102 |
| SL004768 | Allograft inflammatory factor 1 | AIF1 | P55008 | 199 | AIF1 | 1.07166 | 0.64252 |
| SL007145 | Nuclear receptor subfamily 1 group D member 1 | NR1D1 | P20393 | 9572 | NR1D1 | 1.031547 | 0.642932 |
| SL008773 | CD109 antigen | CD109 | Q6YHK3 | 135228 | CD109 | -1.058132 | 0.64545 |
| SL007453 | Mitogen-activated protein kinase 11 | MK11 | Q15759 | 5600 | MAPK11 | 1.011387 | 0.646652 |
| SL003304 | Insulin-like growth factor 1 receptor | IGF-I sR | P08069 | 3480 | IGF1R | -1.039066 | 0.649217 |
| SL003792 | Growth factor receptor-bound protein 2 | GRB2 adapter protein | P62993 | 2885 | GRB2 | -1.162515 | 0.64965 |
| SL000346 | Cathepsin H | Cathepsin H | P09668 | 1512 | CTSH | -1.117995 | 0.65026 |
| SL002081 | Cadherin-5 | Cadherin-5 | P33151 | 1003 | CDH5 | -1.044246 | 0.65072 |
| SL000485 | Interleukin-9 | IL-9 | P15248 | 3578 | IL9 | 1.057818 | 0.652059 |
| SL000521 | Interstitial collagenase | MMP-1 | P03956 | 4312 | MMP1 | -1.114074 | 0.653195 |
| SL003655 | Transketolase | Transketolase | P29401 | 7086 | TKT | -1.05165 | 0.65342 |
| SL009412 | Dickkopf-related protein 3 | DKK3 | Q9UBP4 | 27122 | DKK3 | 1.068445 | 0.65545 |
| SL010502 | Mitogen-activated protein kinase 8 | MK08 | P45983 | 5599 | MAPK8 | -1.07934 | 0.658414 |
| SL000052 | Troponin T, cardiac muscle | Troponin T | P45379 | 7139 | TNNT2 | 1.056872 | 0.658603 |
| SL017611 | 14-3-3 protein family | 14-3-3 | P31946, P62258, P61981, Q04917, P27348, P63104, P31947 | 7529, 7531, 7532, 7533, 10971, 7534, 2810 | YWHAB,YWHAE,YWHAG,YWHAH,YWHAQ,YWHAZ,SFN | 1.045597 | 0.65896 |
| SL000480 | Interleukin-4 | IL-4 | P05112 | 3565 | IL4 | 1.016685 | 0.659489 |
| SL000134 | Hepatocyte growth factor receptor | Met | P08581 | 4233 | MET | 1.040408 | 0.662487 |
| SL005258 | Serine/threonine-protein kinase PLK1 | PLK-1 | P53350 | 5347 | PLK1 | -1.001303 | 0.662705 |
| SL007242 | Dual specificity mitogen-activated protein kinase kinase 3 | MP2K3 | P46734 | 5606 | MAP2K3 | -1.058941 | 0.662999 |
| SL000251 | Alpha-2-HS-glycoprotein | a2-HS-Glycoprotein | P02765 | 197 | AHSG | 1.016378 | 0.663521 |
| SL010391 | WAP, Kazal, immunoglobulin, Kunitz and NTR domain-containing protein 2 | WFKN2 | Q8TEU8 | 124857 | WFIKKN2 | 1.047814 | 0.66538 |
| SL011499 | Pancreatic hormone | PH | P01298 | 5539 | PPY | -1.189398 | 0.666237 |
| SL000553 | Protein kinase C beta type (splice variant beta-II) | PKC-B-II | P05771 | 5579 | PRKCB | -1.139839 | 0.666787 |
| SL000474 | Interleukin-16 | IL-16 | Q14005 | 3603 | IL16 | 1.026113 | 0.668793 |
| SL000581 | Superoxide dismutase [Cu-Zn] | SOD | P00441 | 6647 | SOD1 | -1.051809 | 0.670708 |
| SL004742 | Afamin | Afamin | P43652 | 173 | AFM | 1.014315 | 0.671636 |
| SL000522 | Macrophage metalloelastase | MMP-12 | P39900 | 4321 | MMP12 | 1.042165 | 0.673086 |
| SL011529 | Ribosome maturation protein SBDS | SBDS | Q9Y3A5 | 51119 | SBDS | -1.064469 | 0.673102 |
| SL015510 | Ubiquitin-like protein ISG15 | UCRP | P05161 | 9636 | ISG15 | 1.039601 | 0.673938 |
| SL002621 | Midkine | Midkine | P21741 | 4192 | MDK | 1.028249 | 0.675247 |
| SL006523 | Lactadherin | MFGM | Q08431 | 4240 | MFGE8 | -1.016001 | 0.675565 |
| SL004589 | Azurocidin | Azurocidin | P20160 | 566 | AZU1 | -1.026816 | 0.675637 |
| SL005191 | Leukocyte immunoglobulin-like receptor subfamily B member 2 | ILT-4 | Q8N423 | 10288 | LILRB2 | 1.101431 | 0.67853 |
| SL000456 | Complement C3b, inactivated | iC3b | P01024 | 718 | C3 | -1.006267 | 0.678823 |
| SL004097 | Mothers against decapentaplegic homolog 3 | SMAD3 | P84022 | 4088 | SMAD3 | 1.091287 | 0.679409 |
| SL000540 | Plasmin | Plasmin | P00747 | 5340 | PLG | -1.063708 | 0.679799 |
| SL014148 | Roundabout homolog 3 | ROBO3 | Q96MS0 | 64221 | ROBO3 | 1.018315 | 0.680734 |
| SL000588 | Thyroid peroxidase | TMA | P07202 | 7173 | TPO | 1.017263 | 0.681368 |
| SL000130 | G2/mitotic-specific cyclin-B1 | Cyclin B1 | P14635 | 891 | CCNB1 | 1.012325 | 0.68274 |
| SL012148 | Peroxisomal targeting signal 1 receptor | PEX5 | P50542 | 5830 | PEX5 | -1.14116 | 0.683567 |
| SL017328 | Myosin-binding protein C, slow-type | MYPC1 | Q00872 | 4604 | MYBPC1 | 1.256947 | 0.684269 |
| SL000408 | Erythropoietin | Epo | P01588 | 2056 | EPO | 1.129217 | 0.685469 |
| SL008614 | Ephrin-B1 | EFNB1 | P98172 | 1947 | EFNB1 | -1.018033 | 0.686681 |
| SL008623 | Contactin-2 | CNTN2 | Q02246 | 6900 | CNTN2 | 1.009853 | 0.686857 |
| SL018971 | E3 ubiquitin-protein ligase ZNRF3 | ZNRF3 | Q9ULT6 | 84133 | ZNRF3 | 1.027499 | 0.688203 |
| SL010530 | OCIA domain-containing protein 1 | OCAD1 | Q9NX40 | 54940 | OCIAD1 | 1.036816 | 0.690609 |
| SL007310 | Ras-related C3 botulinum toxin substrate 3 | RAC3 | P60763 | 5881 | RAC3 | 1.025831 | 0.690846 |
| SL008158 | Histone H3.1 | H31 | P68431 | 8350 | HIST1H3A | 1.059483 | 0.692013 |
| SL001945 | E-selectin | sE-Selectin | P16581 | 6401 | SELE | 1.09471 | 0.692821 |
| SL014129 | Heparan-sulfate 6-O-sulfotransferase 1 | H6ST1 | O60243 | 9394 | HS6ST1 | -1.035941 | 0.694902 |
| SL004838 | Leukocyte surface antigen CD47 | CD47 | Q08722 | 961 | CD47 | -1.035581 | 0.695626 |
| SL004340 | Fibroblast growth factor 6 | FGF-6 | P10767 | 2251 | FGF6 | -1.0474 | 0.697061 |
| SL001890 | Tumor-associated calcium signal transducer 2 | GA733-1 protein | P09758 | 4070 | TACSTD2 | -1.097154 | 0.697302 |
| SL014209 | Killer cell lectin-like receptor subfamily F member 1 | KLRF1 | Q9NZS2 | 51348 | KLRF1 | 1.004707 | 0.697322 |
| SL004140 | Ephrin-A4 | Ephrin-A4 | P52798 | 1945 | EFNA4 | 1.049617 | 0.698062 |
| SL016928 | SLAM family member 7 | SLAF7 | Q9NQ25 | 57823 | SLAMF7 | -1.117094 | 0.700072 |
| SL003326 | C-X-C motif chemokine 11 | I-TAC | O14625 | 6373 | CXCL11 | 1.153525 | 0.701183 |
| SL011400 | Calcium/calmodulin-dependent 3',5'-cyclic nucleotide phosphodiesterase 1A | PDE1A | P54750 | 5136 | PDE1A | -1.01422 | 0.701631 |
| SL000622 | Coagulation Factor V | Coagulation Factor V | P12259 | 2153 | F5 | -1.067468 | 0.702712 |
| SL003323 | C-C motif chemokine 18 | PARC | P55774 | 6362 | CCL18 | 1.109434 | 0.702806 |
| SL010493 | Calcium/calmodulin-dependent protein kinase type II subunit delta | CAMK2D | Q13557 | 817 | CAMK2D | -1.116291 | 0.70284 |
| SL003862 | CD40 ligand | CD40 ligand, soluble | P29965 | 959 | CD40LG | 1.026491 | 0.704482 |
| SL011709 | Pyridoxal phosphate phosphatase | PLPP | Q96GD0 | 57026 | PDXP | -1.095252 | 0.705518 |
| SL005685 | Elongation factor 1-beta | EF-1-beta | P24534 | 1933 | EEF1B2 | -1.070069 | 0.706638 |
| SL000004 | Fibroblast growth factor 2 | bFGF | P09038 | 2247 | FGF2 | 1.07498 | 0.706651 |
| SL007228 | Protein kinase C iota type | KPCI | P41743 | 5584 | PRKCI | -1.036185 | 0.707823 |
| SL000506 | Luteinizing hormone | Luteinizing hormone | P01215, P01229 | 1081 3972 | CGA LHB | -1.131299 | 0.708385 |
| SL009629 | Methyl-CpG-binding domain protein 4 | MBD4 | O95243 | 8930 | MBD4 | -1.066522 | 0.70849 |
| SL001728 | Platelet-derived growth factor receptor alpha | PDGFRA | P16234 | 5156 | PDGFRA | -1.885745 | 0.709624 |
| SL005229 | NKG2D ligand 3 | ULBP-3 | Q9BZM4 | 79465 | ULBP3 | -1.026223 | 0.709957 |
| SL005261 | Tyrosine-protein phosphatase non-receptor type 2 | TCPTP | P17706 | 5771 | PTPN2 | 1.003214 | 0.711745 |
| SL000053 | Tissue-type plasminogen activator | tPA | P00750 | 5327 | PLAT | 1.102277 | 0.71189 |
| SL000674 | Follistatin | FST | P19883 | 10468 | FST | -1.041682 | 0.713036 |
| SL011405 | cGMP-specific 3',5'-cyclic phosphodiesterase | PDE5A | O76074 | 8654 | PDE5A | 1.010005 | 0.71511 |
| SL000616 | Vitronectin | Vitronectin | P04004 | 7448 | VTN | -1.038074 | 0.715242 |
| SL007747 | Serine/threonine-protein kinase TBK1 | TBK1 | Q9UHD2 | 29110 | TBK1 | -1.006124 | 0.715906 |
| SL004823 | Peptidyl-prolyl cis-trans isomerase A | Cyclophilin A | P62937 | 5478 | PPIA | -1.071703 | 0.717305 |
| SL011528 | 40S ribosomal protein S7 | RS7 | P62081 | 6201 | RPS7 | 1.102421 | 0.718032 |
| SL014268 | OX-2 membrane glycoprotein | OX2G | P41217 | 4345 | CD200 | -1.04062 | 0.718631 |
| SL004145 | Tumor necrosis factor receptor superfamily member 14 | HVEM | Q92956 | 8764 | TNFRSF14 | -1.611485 | 0.718977 |
| SL010517 | Serine/threonine-protein kinase 16 | STK16 | O75716 | 8576 | STK16 | 1.024895 | 0.71909 |
| SL001766 | Human Chorionic Gonadotropin | HCG | P01215,P01233 | 1081 1082 | CGA CGB | 1.998289 | 0.719334 |
| SL009951 | Protein Wnt-7a | WNT7A | O00755 | 7476 | WNT7A | 1.057391 | 0.71947 |
| SL002519 | Receptor tyrosine-protein kinase erbB-3 | ERBB3 | P21860 | 2065 | ERBB3 | 1.012588 | 0.719489 |
| SL010462 | Ficolin-1 | FCN1 | O00602 | 2219 | FCN1 | -1.010242 | 0.720199 |
| SL005188 | Interleukin-5 receptor subunit alpha | IL-5 Ra | Q01344 | 3568 | IL5RA | -1.070785 | 0.72138 |
| SL010471 | Testican-2 | Testican-2 | Q92563 | 9806 | SPOCK2 | -1.025169 | 0.722299 |
| SL001753 | Sialoadhesin | Sialoadhesin | Q9BZZ2 | 6614 | SIGLEC1 | 2.262808 | 0.722359 |
| SL000017 | von Willebrand factor | vWF | P04275 | 7450 | VWF | 1.056583 | 0.723016 |
| SL008113 | ATP synthase subunit O, mitochondrial | ATPO | P48047 | 539 | ATP5O | -1.662387 | 0.723141 |
| SL003328 | Complement factor I | Factor I | P05156 | 3426 | CFI | -1.009575 | 0.723772 |
| SL000131 | Proliferating cell nuclear antigen | PCNA | P12004 | 5111 | PCNA | 1.021562 | 0.724646 |
| SL003349 | Hypoxia-inducible factor 1-alpha | HIF-1a | Q16665 | 3091 | HIF1A | -1.024865 | 0.725787 |
| SL009202 | Junctional adhesion molecule-like | JAML1 | Q86YT9 | 120425 | AMICA1 | -2.340505 | 0.728614 |
| SL004821 | Protein S100-A4 | S100A4 | P26447 | 6275 | S100A4 | 1.028931 | 0.729316 |
| SL013165 | Non-histone chromosomal protein HMG-14 | HMGN1 | P05114 | 3150 | HMGN1 | -1.032663 | 0.729367 |
| SL004253 | Estradiol 17-beta-dehydrogenase 1 | 17-beta-HSD 1 | P14061 | 3292 | HSD17B1 | 1.001127 | 0.729752 |
| SL008611 | N-acylethanolamine-hydrolyzing acid amidase | ASAHL | Q02083 | 27163 | NAAA | -1.023708 | 0.73057 |
| SL003188 | C-X-C motif chemokine 9 | MIG | Q07325 | 4283 | CXCL9 | -1.049731 | 0.731882 |
| SL009790 | Small nuclear ribonucleoprotein F | RUXF | P62306 | 6636 | SNRPF | 1.032066 | 0.733953 |
| SL007025 | Tyrosine-protein kinase JAK2 | JAK2 | O60674 | 3717 | JAK2 | 1.007496 | 0.73462 |
| SL005194 | Junctional adhesion molecule C | JAM-C | Q9BX67 | 83700 | JAM3 | -1.057766 | 0.734824 |
| SL014735 | Inosine-5'-monophosphate dehydrogenase 1 | IMDH1 | P20839 | 3614 | IMPDH1 | 1.027454 | 0.735141 |
| SL007284 | Cysteine-rich secretory protein 3 | CRIS3 | P54108 | 10321 | CRISP3 | -1.360337 | 0.735604 |
| SL004812 | Triosephosphate isomerase | Triosephosphate isomerase | P60174 | 7167 | TPI1 | -1.0261 | 0.736882 |
| SL005187 | Interleukin-3 receptor subunit alpha | IL-3 Ra | P26951 | 3563 | IL3RA | -1.102542 | 0.737672 |
| SL000007 | Estrogen receptor | ER | P03372 | 2099 | ESR1 | 1.023203 | 0.738088 |
| SL010492 | Calcium/calmodulin-dependent protein kinase type II subunit beta | CAMK2B | Q13554 | 816 | CAMK2B | -1.081279 | 0.739069 |
| SL005725 | GRB2-related adapter protein 2 | GRB2-related adapter protein 2 | O75791 | 9402 | GRAP2 | -1.13397 | 0.739745 |
| SL004134 | CD27 antigen | CD27 | P26842 | 939 | CD27 | 1.092076 | 0.740135 |
| SL003461 | Corticotropin | ACTH | P01189 | 5443 | POMC | -1.10686 | 0.741533 |
| SL004329 | Growth/differentiation factor 5 | BMP-14 | P43026 | 8200 | GDF5 | 1.032564 | 0.741601 |
| SL007179 | Ephrin type-B receptor 2 | EPHB2 | P29323 | 2048 | EPHB2 | -1.027911 | 0.742014 |
| SL004723 | Histone deacetylase 8 | HDAC8 | Q9BY41 | 55869 | HDAC8 | -1.033614 | 0.74275 |
| SL007151 | Glycerol-3-phosphate dehydrogenase [NAD(+)], cytoplasmic | GPDA | P21695 | 2819 | GPD1 | 1.193452 | 0.743043 |
| SL005197 | Lysosome membrane protein 2 | LIMP II | Q14108 | 950 | SCARB2 | -1.076872 | 0.743109 |
| SL004327 | Tumor necrosis factor ligand superfamily member 13B | BAFF | Q9Y275 | 10673 | TNFSF13B | -1.026162 | 0.743316 |
| SL004871 | Tumor necrosis factor receptor superfamily member 21 | DR6 | O75509 | 27242 | TNFRSF21 | 1.022378 | 0.744417 |
| SL007327 | Oxidized low-density lipoprotein receptor 1 | OLR1 | P78380 | 4973 | OLR1 | -1.013674 | 0.744518 |
| SL010454 | Contactin-4 | Contactin-4 | Q8IWV2 | 152330 | CNTN4 | -1.023282 | 0.744541 |
| SL002508 | Interleukin-18-binding protein | IL-18 BPa | O95998 | 10068 | IL18BP | -1.017191 | 0.744697 |
| SL014308 | Fibroblast growth factor 8 isoform A | FGF-8A | P55075 | 2253 | FGF8 | -1.12373 | 0.744753 |
| SL004208 | Annexin A1 | annexin I | P04083 | 301 | ANXA1 | 1.076699 | 0.744974 |
| SL008909 | Legumain | LGMN | Q99538 | 5641 | LGMN | -1.022426 | 0.745116 |
| SL010490 | Calcium/calmodulin-dependent protein kinase type 1D | CAMK1D | Q8IU85 | 57118 | CAMK1D | -1.024647 | 0.745153 |
| SL004306 | SUMO-conjugating enzyme UBC9 | UBC9 | P63279 | 7329 | UBE2I | -1.044189 | 0.745937 |
| SL000403 | Endostatin | Endostatin | P39060 | 80781 | COL18A1 | 1.024509 | 0.747136 |
| SL006029 | Chitotriosidase-1 | Chitotriosidase-1 | Q13231 | 1118 | CHIT1 | 1.045999 | 0.747182 |
| SL003704 | BH3-interacting domain death agonist | BID | P55957 | 637 | BID | -1.814138 | 0.748656 |
| SL004336 | Fibroblast growth factor 18 | FGF-18 | O76093 | 8817 | FGF18 | -1.011335 | 0.748679 |
| SL009791 | Heterogeneous nuclear ribonucleoprotein A/B | hnRNP A/B | Q99729 | 3182 | HNRNPAB | 1.072758 | 0.749419 |
| SL004015 | Carboxypeptidase B2 | TAFI | Q96IY4 | 1361 | CPB2 | -1.006639 | 0.7501 |
| SL010499 | Tyrosine-protein kinase HCK | HCK | P08631 | 3055 | HCK | -1.027176 | 0.750939 |
| SL009768 | Chromobox protein homolog 5 | CBX5 | P45973 | 23468 | CBX5 | -1.036466 | 0.751159 |
| SL010514 | Protein-tyrosine kinase 6 | PTK6 | Q13882 | 5753 | PTK6 | -1.149639 | 0.751938 |
| SL004008 | Myeloblastin | Proteinase-3 | P24158 | 5657 | PRTN3 | -1.00969 | 0.752107 |
| SL000582 | Baculoviral IAP repeat-containing protein 5 | Survivin | O15392 | 332 | BIRC5 | -1.02412 | 0.752525 |
| SL005196 | Limbic system-associated membrane protein | LSAMP | Q13449 | 4045 | LSAMP | -1.048201 | 0.754479 |
| SL008176 | Proteasome activator complex subunit 1 | PSME1 | Q06323 | 5720 | PSME1 | -1.033167 | 0.754996 |
| SL012538 | Cerebral dopamine neurotrophic factor | ARMEL | Q49AH0 | 441549 | CDNF | 1.020624 | 0.755888 |
| SL009792 | Adenylosuccinate lyase | PUR8 | P30566 | 158 | ADSL | -1.122115 | 0.755941 |
| SL010830 | Protein 4.1 | 41 | P11171 | 2035 | EPB41 | 1.080119 | 0.756288 |
| SL010393 | Kremen protein 2 | KREM2 | Q8NCW0 | 79412 | KREMEN2 | 1.009973 | 0.756711 |
| SL011629 | AH receptor-interacting protein | AIP | O00170 | 9049 | AIP | -1.067901 | 0.757533 |
| SL009868 | FACT complex subunit SSRP1 | SSRP1 | Q08945 | 6749 | SSRP1 | 1.026698 | 0.758063 |
| SL000460 | Immunoglobulin D | IgD | P01880 | 3495 50802 3535 | IGHD IGK@ IGL@ | -1.463946 | 0.758285 |
| SL005202 | Hepatocyte growth factor-like protein | MSP | P26927 | 4485 | MST1 | -1.058076 | 0.760633 |
| SL008102 | Malate dehydrogenase, cytoplasmic | MDHC | P40925 | 4190 | MDH1 | 1.128475 | 0.764259 |
| SL004642 | Disintegrin and metalloproteinase domain-containing protein 9 | ADAM 9 | Q13443 | 8754 | ADAM9 | -1.12349 | 0.764647 |
| SL003793 | Dual specificity mitogen-activated protein kinase kinase 1 | MEK1 | Q02750 | 5604 | MAP2K1 | 1.030733 | 0.765383 |
| SL005165 | Galectin-4 | Galectin-4 | P56470 | 3960 | LGALS4 | -1.085083 | 0.765718 |
| SL000062 | Prostate-specific antigen | PSA | P07288 | 354 | KLK3 | 1.354138 | 0.766804 |
| SL009349 | Follistatin-related protein 1 | FSTL1 | Q12841 | 11167 | FSTL1 | 1.032243 | 0.766838 |
| SL000314 | Complement C3b | C3b | P01024 | 718 | C3 | 1.106445 | 0.767353 |
| SL004829 | Protein S100-A7 | S100A7 | P31151 | 6278 | S100A7 | 1.316206 | 0.769216 |
| SL010973 | Proteasome subunit alpha type-1 | PSA1 | P25786 | 5682 | PSMA1 | -1.777996 | 0.769271 |
| SL018587 | Immunoglobulin superfamily containing leucine-rich repeat protein 2 | ISLR2 | Q6UXK2 | 57611 | ISLR2 | 1.00094 | 0.769742 |
| SL000280 | Aspartate aminotransferase, cytoplasmic | GOT1 | P17174 | 2805 | GOT1 | 1.070513 | 0.769904 |
| SL012774 | Cysteine-rich with EGF-like domain protein 1 | CREL1 | Q96HD1 | 78987 | CRELD1 | -1.002481 | 0.770486 |
| SL000573 | Serum amyloid P-component | SAP | P02743 | 325 | APCS | 1.034629 | 0.771029 |
| SL014289 | Killer cell immunoglobulin-like receptor 3DS1 | KI3S1 | Q14943 | 3813 | KIR3DS1 | -1.028825 | 0.77259 |
| SL003168 | C-C motif chemokine 27 | CTACK | Q9Y4X3 | 10850 | CCL27 | -1.048826 | 0.774759 |
| SL004070 | Ubiquitin | Ubiquitin | P62979 | 6233 | RPS27A | -1.076707 | 0.776094 |
| SL004759 | Eukaryotic translation initiation factor 5 | eIF-5 | P55010 | 1983 | EIF5 | 1.014307 | 0.776365 |
| SL010468 | RGM domain family member B | RGMB | Q6NW40 | 285704 | RGMB | 1.024861 | 0.777212 |
| SL003710 | Caspase-2 | Caspase-2 | P42575 | 835 | CASP2 | 1.012413 | 0.777265 |
| SL010494 | Cyclin-dependent kinase 1:G2/mitotic-specific cyclin-B1 complex | CDK1/cyclin B | P06493 P14635 | 983 891 | CDC2 CCNB1 | 1.014071 | 0.778571 |
| SL001902 | Basal Cell Adhesion Molecule | BCAM | P50895 | 4059 | BCAM | 1.0107 | 0.779419 |
| SL000415 | Complement factor H | Factor H | P08603 | 3075 | CFH | -1.000355 | 0.779771 |
| SL004147 | Interleukin-10 receptor subunit beta | IL-10 Rb | Q08334 | 3588 | IL10RB | 1.028164 | 0.780606 |
| SL004489 | Toll-like receptor 4 | TLR4 | O00206 | 7099 | TLR4 | -1.110362 | 0.780653 |
| SL003167 | C-X-C motif chemokine 13 | BLC | O43927 | 10563 | CXCL13 | -1.01951 | 0.78144 |
| SL002522 | Retinoblastoma-associated protein | Rb | P06400 | 5925 | RB1 | 1.008704 | 0.781855 |
| SL001726 | Granulocyte-macrophage colony-stimulating factor | GM-CSF | P04141 | 1437 | CSF2 | -1.256993 | 0.78247 |
| SL003104 | C-C motif chemokine 24 | Eotaxin-2 | O00175 | 6369 | CCL24 | 1.008345 | 0.782803 |
| SL004940 | Tyrosine-protein phosphatase non-receptor type 6 | PTP-1C | P29350 | 5777 | PTPN6 | -1.093216 | 0.782904 |
| SL010463 | Glypican-2 | GPC2 | Q8N158 | 221914 | GPC2 | 1.007429 | 0.783472 |
| SL000519 | C-C motif chemokine 3 | MIP-1a | P10147 | 6348 | CCL3 | -1.019284 | 0.784459 |
| SL016828 | Epidermal growth factor receptor variant III | EGFRvIII | P00533 | NaN | EGFR | -1.026228 | 0.785358 |
| SL001999 | E3 ubiquitin-protein ligase Mdm2 | MDM2 | Q00987 | 4193 | MDM2 | -1.336012 | 0.787024 |
| SL004815 | 14-3-3 protein beta/alpha | 14-3-3 protein beta/alpha | P31946 | 7529 | YWHAB | -1.071499 | 0.788268 |
| SL004984 | 14-3-3 protein epsilon | 14-3-3E | P62258 | 7531 | YWHAE | 1.024577 | 0.788365 |
| SL006830 | Complement factor H-related protein 5 | complement factor H-related 5 | Q9BXR6 | 81494 | CFHR5 | -1.000654 | 0.788756 |
| SL000133 | C-C motif chemokine 20 | MIP-3a | P78556 | 6364 | CCL20 | -1.102741 | 0.790216 |
| SL003872 | Interleukin-6 receptor subunit beta | gp130, soluble | P40189 | 3572 | IL6ST | -1.035987 | 0.790753 |
| SL004720 | Calcineurin subunit B type 1 | Calcineurin B a | P63098 | 5534 | PPP3R1 | -1.01438 | 0.790946 |
| SL003785 | Glyceraldehyde-3-phosphate dehydrogenase | GAPDH, liver | P04406 | 2597 | GAPDH | -1.054477 | 0.792875 |
| SL000383 | Creatine kinase M-type | CK-MM | P06732 | 1158 | CKM | 1.08405 | 0.793375 |
| SL004149 | Interleukin-13 receptor subunit alpha-1 | IL-13 Ra1 | P78552 | 3597 | IL13RA1 | 1.040396 | 0.794744 |
| SL000601 | Serotransferrin | Transferrin | P02787 | 7018 | TF | -1.003916 | 0.795128 |
| SL000022 | D-dimer | D-dimer | P02671 P02675 P02679 | 2243 2244 2266 | FGA FGB FGG | -1.023907 | 0.796513 |
| SL008590 | Olfactomedin-4 | Olfactomedin-4 | Q6UX06 | 10562 | OLFM4 | -1.026685 | 0.797636 |
| SL003646 | Tropomyosin alpha-4 chain | Tropomyosin 4 | P67936 | 7171 | TPM4 | -1.131049 | 0.799089 |
| SL002071 | Gastrin-releasing peptide | Gastrin-releasing peptide | P07492 | 2922 | GRP | 1.033619 | 0.799271 |
| SL008945 | Protein-glutamine gamma-glutamyltransferase E | TGM3 | Q08188 | 7053 | TGM3 | -1.710962 | 0.799958 |
| SL000528 | NADPH--cytochrome P450 reductase | NADPH-P450 Oxidoreductase | P16435 | 5447 | POR | -1.003535 | 0.800168 |
| SL000089 | Transforming growth factor beta-3 | TGF-b3 | P10600 | 7043 | TGFB3 | 1.01786 | 0.800712 |
| SL004334 | Fibroblast growth factor 16 | FGF-16 | O43320 | 8823 | FGF16 | -1.047209 | 0.800766 |
| SL003700 | Bcl2-associated agonist of cell death | BAD | Q92934 | 572 | BAD | -1.069767 | 0.801472 |
| SL008372 | Leucine-rich repeat transmembrane protein FLRT3 | FLRT3 | Q9NZU0 | 23767 | FLRT3 | 1.023153 | 0.802531 |
| SL004737 | Tropomyosin alpha-1 chain | Tropomyosin 1 alpha chain | P09493 | 7168 | TPM1 | -1.00686 | 0.802717 |
| SL004333 | Fibroblast growth factor 10 | FGF-10 | O15520 | 2255 | FGF10 | -1.011211 | 0.802855 |
| SL012469 | Glypican-5 | GPC5 | P78333 | 2262 | GPC5 | -1.040671 | 0.803938 |
| SL007049 | Cystatin-F | CYTF | O76096 | 8530 | CST7 | -1.174792 | 0.804357 |
| SL004126 | Tumor necrosis factor receptor superfamily member 9 | 4-1BB | Q07011 | 3604 | TNFRSF9 | -1.909105 | 0.804867 |
| SL010613 | Interleukin-17 receptor D | IL-17 RD | Q8NFM7 | 54756 | IL17RD | -1.013873 | 0.805425 |
| SL003341 | Fibrinogen gamma chain | Fibrinogen g-chain dimer | P02679 | 2266 | FGG | -1.00837 | 0.807761 |
| SL000339 | Carbonic anhydrase 2 | carbonic anhydrase II | P00918 | 760 | CA2 | -1.109825 | 0.808776 |
| SL000344 | Cathepsin D | Cathepsin D | P07339 | 1509 | CTSD | -1.139784 | 0.811981 |
| SL002036 | Fibroblast growth factor receptor 4 | FGFR4 | P22455 | 2264 | FGFR4 | -1.078329 | 0.811994 |
| SL006544 | Transforming growth factor-beta-induced protein ig-h3 | BGH3 | Q15582 | 7045 | TGFBI | 1.041392 | 0.81381 |
| SL003770 | Secreted frizzled-related protein 1 | SARP-2 | Q8N474 | 6422 | SFRP1 | 1.045236 | 0.814388 |
| SL000318 | Complement C4b | C4b | P0C0L4 P0C0L5 | 720 721 | C4A C4B | 1.06195 | 0.814439 |
| SL011211 | Eukaryotic translation initiation factor 4 gamma 2 | IF4G2 | P78344 | 1982 | EIF4G2 | -1.097096 | 0.814905 |
| SL004845 | Ephrin type-B receptor 4 | EphB4 | P54760 | 2050 | EPHB4 | 1.010848 | 0.815149 |
| SL007804 | Biglycan | BGN | P21810 | 633 | BGN | 1.056841 | 0.81523 |
| SL004849 | X-linked interleukin-1 receptor accessory protein-like 2 | IL-1 sR9 | Q9NP60 | 26280 | IL1RAPL2 | -1.057976 | 0.815618 |
| SL005437 | Matrix extracellular phosphoglycoprotein | MEPE | Q9NQ76 | 56955 | MEPE | -1.013385 | 0.820669 |
| SL008591 | Trefoil factor 1 | TFF1 | P04155 | 7031 | TFF1 | 1.09966 | 0.821478 |
| SL002539 | Tumor necrosis factor receptor superfamily member 11B | OPG | O00300 | 4982 | TNFRSF11B | -1.018974 | 0.821769 |
| SL016566 | Serine/threonine-protein kinase 17B | DRAK2 | O94768 | 9262 | STK17B | 1.030807 | 0.823047 |
| SL004591 | Granulocyte colony-stimulating factor receptor | G-CSF-R | Q99062 | 1441 | CSF3R | -1.019846 | 0.825119 |
| SL006920 | Mitogen-activated protein kinase 14 | MAPK14 | Q16539 | 1432 | MAPK14 | -1.041525 | 0.825908 |
| SL000310 | Complement C1r subcomponent | C1r | P00736 | 715 | C1R | 1.041696 | 0.826428 |
| SL004733 | Low-density lipoprotein receptor-related protein 1, soluble | sLRP1 | Q07954 | 4035 | LRP1 | -1.02067 | 0.829792 |
| SL004209 | Annexin A2 | annexin II | P07355 | 302 | ANXA2 | -1.030838 | 0.830288 |
| SL004626 | A disintegrin and metalloproteinase with thrombospondin motifs 5 | ADAMTS-5 | Q9UNA0 | 11096 | ADAMTS5 | 1.008026 | 0.830945 |
| SL003327 | Complement factor D | Factor D | P00746 | 1675 | CFD | -1.017224 | 0.831152 |
| SL003863 | Kallikrein-5 | kallikrein 5 | Q9Y337 | 25818 | KLK5 | -1.356402 | 0.831537 |
| SL000640 | Nidogen-1 | Nidogen | P14543 | 4811 | NID1 | -1.009167 | 0.832678 |
| SL004932 | Peroxiredoxin-5, mitochondrial | Peroxiredoxin-5 | P30044 | 25824 | PRDX5 | 1.021345 | 0.833703 |
| SL008059 | 40S ribosomal protein S3 | RS3 | P23396 | 6188 | RPS3 | -1.031352 | 0.834814 |
| SL006448 | Histidine-rich glycoprotein | HRG | P04196 | 3273 | HRG | 1.080853 | 0.839162 |
| SL018938 | Small ubiquitin-related modifier 3 | SUMO3 | P55854 | 6613 | SUMO3 | -1.065725 | 0.840255 |
| SL005059 | Transforming growth factor beta receptor type 3 | TGF-b R III | Q03167 | 7049 | TGFBR3 | 1.013418 | 0.842122 |
| SL017424 | Platelet receptor Gi24 | GI24 | Q9H7M9 | NaN | C10orf54 | -1.00272 | 0.843276 |
| SL010520 | Aurora kinase B | AURKB | Q96GD4 | 9212 | AURKB | 1.00601 | 0.843559 |
| SL006970 | Delta-like protein 1 | DLL1 | O00548 | 28514 | DLL1 | -1.020445 | 0.845214 |
| SL012881 | Glypican-6 | GPC6 | Q9Y625 | 10082 | GPC6 | -1.01512 | 0.846727 |
| SL003187 | C-C motif chemokine 22 | MDC | O00626 | 6367 | CCL22 | 1.068834 | 0.847374 |
| SL004636 | Receptor-type tyrosine-protein kinase FLT3 | Flt-3 | P36888 | 2322 | FLT3 | -1.023741 | 0.847963 |
| SL000420 | Ferritin | Ferritin | P02794 P02792 | 2495 2512 | FTH1 FTL | -1.042169 | 0.848085 |
| SL000248 | Alpha-1-antichymotrypsin | a1-Antichymotrypsin | P01011 | 12 | SERPINA3 | -1.079697 | 0.848308 |
| SL003970 | Parathyroid hormone | PTH | P01270 | 5741 | PTH | -1.263491 | 0.848389 |
| SL003182 | Integrin alpha-V: beta-5 complex | Integrin aVb5 | P06756, P18084 | 3685 3693 | ITGAV ITGB5 | 1.015953 | 0.848608 |
| SL007953 | Pyridoxal kinase | PDXK | O00764 | 8566 | PDXK | 1.001527 | 0.851226 |
| SL011616 | Eukaryotic initiation factor 4A-III | IF4A3 | P38919 | 9775 | EIF4A3 | -1.014043 | 0.853788 |
| SL004687 | Thymic stromal lymphopoietin | TSLP | Q969D9 | 85480 | TSLP | -1.012105 | 0.855496 |
| SL005688 | 14-3-3 protein zeta/delta | 14-3-3 protein zeta/delta | P63104 | 7534 | YWHAZ | -1.086578 | 0.856913 |
| SL005209 | Neurogenic locus notch homolog protein 3 | Notch-3 | Q9UM47 | 4854 | NOTCH3 | 1.012606 | 0.857926 |
| SL004804 | Cell adhesion molecule 3 | Nectin-like protein 1 | Q8N126 | 57863 | CADM3 | 1.044623 | 0.858668 |
| SL014093 | Ectonucleoside triphosphate diphosphohydrolase 3 | ENTP3 | O75355 | 956 | ENTPD3 | -1.060061 | 0.859639 |
| SL000124 | 72 kDa type IV collagenase | MMP-2 | P08253 | 4313 | MMP2 | -1.001591 | 0.860281 |
| SL004580 | Macrophage scavenger receptor types I and II | Macrophage scavenger receptor | P21757 | 4481 | MSR1 | -1.017776 | 0.860757 |
| SL000002 | Vascular endothelial growth factor A | VEGF | P15692 | 7422 | VEGFA | 1.025981 | 0.862229 |
| SL010455 | Contactin-5 | Contactin-5 | O94779 | 53942 | CNTN5 | -1.030109 | 0.863239 |
| SL016148 | gp41 C34 peptide, HIV | C34 gp41 HIV Fragment | Q70626 | NaN | Human-virus | 1.047321 | 0.863633 |
| SL006476 | Glycylpeptide N-tetradecanoyltransferase 1 | NMT1 | P30419 | 4836 | NMT1 | -1.041082 | 0.864234 |
| SL005679 | Translationally-controlled tumor protein | TCTP | P13693 | 7178 | TPT1 | -1.037032 | 0.865933 |
| SL004338 | Fibroblast growth factor 20 | FGF-20 | Q9NP95 | 26281 | FGF20 | 1.010972 | 0.871335 |
| SL003197 | C-C motif chemokine 25 | TECK | O15444 | 6370 | CCL25 | -1.073705 | 0.873028 |
| SL002517 | Tumor necrosis factor | TNF-a | P01375 | 7124 | TNF | -1.004439 | 0.876207 |
| SL010509 | Serine/threonine-protein kinase PAK 6 | PAK6 | Q9NQU5 | 56924 | PAK6 | -1.09771 | 0.878051 |
| SL004805 | Cell adhesion molecule 1 | Nectin-like protein 2 | Q9BY67 | 23705 | CADM1 | -1.002375 | 0.87964 |
| SL004271 | Glial fibrillary acidic protein | GFAP | P14136 | 2670 | GFAP | -1.0477 | 0.881366 |
| SL017289 | PolyUbiquitin K48-linked | PolyUbiquitin K48 | P0CG47 | 7314 | UBB | 1.022213 | 0.882312 |
| SL011770 | Leucine carboxyl methyltransferase 1 | LCMT1 | Q9UIC8 | 51451 | LCMT1 | -1.009199 | 0.883033 |
| SL008933 | Protein deglycase DJ-1 | PARK7 | Q99497 | 11315 | PARK7 | -1.074723 | 0.884606 |
| SL000455 | Transcription factor AP-1 | c-Jun | P05412 | 3725 | JUN | -1.007836 | 0.885037 |
| SL000137 | Betacellulin | BTC | P35070 | 685 | BTC | 1.002824 | 0.890441 |
| SL003849 | Fibroblast growth factor 9 | FGF9 | P31371 | 2254 | FGF9 | -1.016555 | 0.890821 |
| SL010371 | Ectonucleoside triphosphate diphosphohydrolase 1 | CD39 | P49961 | 953 | ENTPD1 | -1.013615 | 0.892113 |
| SL001897 | Kunitz-type protease inhibitor 2 | SPINT2 | O43291 | 10653 | SPINT2 | -1.028338 | 0.892608 |
| SL007223 | B-cell lymphoma 6 protein | BCL6 | P41182 | 604 | BCL6 | 1.190407 | 0.895293 |
| SL000590 | Thyroxine-binding globulin | Thyroxine-Binding Globulin | P05543 | 6906 | SERPINA7 | 1.001982 | 0.895357 |
| SL012769 | Protein FAM3D | FAM3D | Q96BQ1 | 131177 | FAM3D | -1.06861 | 0.895787 |
| SL006397 | Neuropilin-1 | NRP1 | O14786 | 8829 | NRP1 | -1.000743 | 0.896104 |
| SL004354 | Interleukin-19 | IL-19 | Q9UHD0 | 29949 | IL19 | 1.015657 | 0.89633 |
| SL011049 | Mannan-binding lectin serine protease 1 | MASP3 | P48740 | 5648 | MASP1 | -1.007096 | 0.8983 |
| SL003362 | Complement C3d fragment | C3d | P01024 | 718 | C3 | 1.031234 | 0.898968 |
| SL005217 | Sialic acid-binding Ig-like lectin 6 | Siglec-6 | O43699 | 946 | SIGLEC6 | -1.004328 | 0.899871 |
| SL006924 | Agouti-related protein | ART | O00253 | 181 | AGRP | -1.023355 | 0.900262 |
| SL010449 | Carbonic anhydrase-related protein 10 | Carbonic Anhydrase X | Q9NS85 | 56934 | CA10 | -1.012763 | 0.9008 |
| SL004467 | Protein Z-dependent protease inhibitor | protein Z inhibitor | Q9UK55 | 51156 | SERPINA10 | 1.022899 | 0.901096 |
| SL000377 | Creatine kinase B-type | CK-BB | P12277 | 1152 | CKB | 1.049394 | 0.902393 |
| SL007324 | Casein kinase II subunit alpha | CSK21 | P68400 | 1457 | CSNK2A1 | -1.030322 | 0.902772 |
| SL004851 | Serine/threonine-protein kinase receptor R3 | ALK-1 | P37023 | 94 | ACVRL1 | -1.240488 | 0.903404 |
| SL014269 | Killer cell immunoglobulin-like receptor 3DL2 | KI3L2 | P43630 | 3812 | KIR3DL2 | -1.159387 | 0.9036 |
| SL007121 | Cathepsin E | CATE | P14091 | 1510 | CTSE | -1.121335 | 0.905641 |
| SL004858 | GDNF family receptor alpha-1 | GFRa-1 | P56159 | 2674 | GFRA1 | 1.012566 | 0.907747 |
| SL010489 | Calcium/calmodulin-dependent protein kinase type 1 | CAMK1 | Q14012 | 8536 | CAMK1 | -1.033123 | 0.907882 |
| SL003990 | Fibroblast growth factor receptor 2 | FGFR-2 | P21802 | 2263 | FGFR2 | -1.819349 | 0.908036 |
| SL008703 | Carboxypeptidase E | CBPE | P16870 | 1363 | CPE | -1.026713 | 0.908277 |
| SL005172 | Insulin-like growth factor-binding protein 6 | IGFBP-6 | P24592 | 3489 | IGFBP6 | 1.00136 | 0.909059 |
| SL004066 | Group IIE secretory phospholipase A2 | GIIE | Q9NZK7 | 30814 | PLA2G2E | -1.012546 | 0.909091 |
| SL013240 | Adapter molecule crk | CRK | P46108 | 1398 | CRK | 1.013617 | 0.911537 |
| SL000510 | Lysozyme C | Lysozyme | P61626 | 4069 | LYZ | -1.01057 | 0.91163 |
| SL000300 | Beta-endorphin | b-Endorphin | P01189 | 5443 | POMC | -1.016392 | 0.912008 |
| SL000633 | Tumor necrosis factor ligand superfamily member 6, soluble form | Fas ligand, soluble | P48023 | 356 | FASLG | 1.040771 | 0.913221 |
| SL018548 | Alpha-1-antichymotrypsin complex | alpha-1-antichymotrypsin complex | P01011 | NaN | SERPINA3 | 1.005473 | 0.91401 |
| SL003648 | Rab GDP dissociation inhibitor beta | Rab GDP dissociation inhibitor beta | P50395 | 2665 | GDI2 | 1.024006 | 0.915851 |
| SL008956 | Arylsulfatase B | ARSB | P15848 | 411 | ARSB | 1.011022 | 0.916162 |
| SL009210 | Pro-opiomelanocortin | Corticotropin-lipotropin | P01189 | 5443 | POMC | 1.008546 | 0.918147 |
| SL004919 | Peroxiredoxin-1 | Peroxiredoxin-1 | Q06830 | 5052 | PRDX1 | -1.024969 | 0.919379 |
| SL002922 | Intercellular adhesion molecule 1 | sICAM-1 | P05362 | 3383 | ICAM1 | -1.01656 | 0.919416 |
| SL003169 | C-X-C motif chemokine 5 | ENA-78 | P42830 | 6374 | CXCL5 | -1.002609 | 0.920047 |
| SL003764 | Neural cell adhesion molecule 1, 120 kDa isoform | NCAM-120 | P13591 | 4684 | NCAM1 | -1.023798 | 0.920112 |
| SL000158 | Glutamate carboxypeptidase 2 | PSMA | Q04609 | 2346 | FOLH1 | -1.034428 | 0.920769 |
| SL000558 | Prothrombin | Prothrombin | P00734 | 2147 | F2 | 1.035642 | 0.920994 |
| SL004712 | Stromal cell-derived factor 1 | SDF-1 | P48061 | 6387 | CXCL12 | 1.02017 | 0.921213 |
| SL008644 | ADP-ribosyl cyclase/cyclic ADP-ribose hydrolase 2 | BST1 | Q10588 | 683 | BST1 | -1.054894 | 0.921857 |
| SL011498 | Pituitary adenylate cyclase-activating polypeptide 38 | PACAP-38 | P18509 | 116 | ADCYAP1 | -1.02043 | 0.922436 |
| SL005171 | Insulin-like growth factor-binding protein 4 | IGFBP-4 | P22692 | 3487 | IGFBP4 | 1.019234 | 0.925246 |
| SL013570 | Dual specificity tyrosine-phosphorylation-regulated kinase 3 | DYRK3 | O43781 | 8444 | DYRK3 | -1.012822 | 0.926265 |
| SL004718 | Importin subunit alpha-1 | Karyopherin-a2 | P52292 | 3838 | KPNA2 | -1.006862 | 0.928332 |
| SL003440 | Platelet-activating factor acetylhydrolase | PAFAH | Q13093 | 7941 | PLA2G7 | 1.016724 | 0.928705 |
| SL007752 | Death-associated protein kinase 2 | DAPK2 | Q9UIK4 | 23604 | DAPK2 | 1.003493 | 0.929089 |
| SL004343 | Fms-related tyrosine kinase 3 ligand | Flt3 ligand | P49771 | 2323 | FLT3LG | -1.232246 | 0.929238 |
| SL007560 | Stabilin-2 | STAB2 | Q8WWQ8 | 55576 | STAB2 | -1.009357 | 0.929867 |
| SL014248 | Semaphorin-6B | SEM6B | Q9H3T3 | 10501 | SEMA6B | 1.029616 | 0.930476 |
| SL009328 | Chorionic somatomammotropin hormone | CSH | P0DML2 P0DML3 | 1442 1443 | CSH1 CSH2 | -1.007669 | 0.931047 |
| SL012168 | Protein lin-7 homolog B | LIN7B | Q9HAP6 | 64130 | LIN7B | 1.014089 | 0.931332 |
| SL011073 | Xaa-Pro aminopeptidase 1 | XPNPEP1 | Q9NQW7 | 7511 | XPNPEP1 | 1.035464 | 0.931399 |
| SL005226 | Cytokine receptor-like factor 2 | TSLP R | Q9HC73 | 64109 | CRLF2 | -1.02781 | 0.932308 |
| SL010470 | Semaphorin-3E | Semaphorin 3E | O15041 | 9723 | SEMA3E | 1.020873 | 0.932427 |
| SL001713 | Interleukin-17A | IL-17 | Q16552 | 3605 | IL17A | 1.008255 | 0.93282 |
| SL014469 | SHC-transforming protein 1 | SHC1 | P29353 | 6464 | SHC1 | -1.022815 | 0.933269 |
| SL005174 | Interleukin-17 receptor B | IL-17B R | Q9NRM6 | 55540 | IL17RB | 1.311219 | 0.933301 |
| SL008193 | Nidogen-2 | NID2 | Q14112 | 22795 | NID2 | 1.014268 | 0.935867 |
| SL000145 | Interleukin-1 receptor type 2 | IL-1 sRII | P27930 | 7850 | IL1R2 | 1.018188 | 0.936028 |
| SL004673 | Cathepsin S | Cathepsin S | P25774 | 1520 | CTSS | 1.006329 | 0.936186 |
| SL004811 | Transgelin-2 | Transgelin-2 | P37802 | 8407 | TAGLN2 | -1.007022 | 0.936527 |
| SL000305 | beta-nerve growth factor | b-NGF | P01138 | 4803 | NGF | -1.132091 | 0.936635 |
| SL000433 | Glucagon | Glucagon | P01275 | 2641 | GCG | 1.023655 | 0.937277 |
| SL012698 | Killer cell immunoglobulin-like receptor 2DL4 | KI2L4 | Q99706 | 3805 | KIR2DL4 | 1.046686 | 0.937879 |
| SL005221 | Scavenger receptor class F member 1 | SREC-I | Q14162 | 8578 | SCARF1 | -1.018505 | 0.938113 |
| SL004698 | Protein E7_HPV18 | HPV E7 Type18 | P06788 | 1489089 | Human-virus | -1.009379 | 0.938345 |
| SL004332 | Prokineticin-1 | EG-VEGF | P58294 | 84432 | PROK1 | -1.015359 | 0.939187 |
| SL007429 | Transmembrane glycoprotein NMB | GPNMB | Q14956 | 10457 | GPNMB | -1.013835 | 0.940384 |
| SL003303 | C-C motif chemokine 28 | CCL28 | Q9NRJ3 | 56477 | CCL28 | 1.003779 | 0.941518 |
| SL000337 | Calpain I | Calpain I | P07384 P04632 | 823 826 | CAPN1 CAPNS1 | -1.053021 | 0.941707 |
| SL000011 | alpha-Fetoprotein | AFP | P02771 | 174 | AFP | -1.012384 | 0.942107 |
| SL000545 | Plasma kallikrein | Prekallikrein | P03952 | 3818 | KLKB1 | 1.069201 | 0.943227 |
| SL006993 | Mitogen-activated protein kinase 13 | MK13 | O15264 | 5603 | MAPK13 | -1.084648 | 0.944947 |
| SL013490 | Brother of CDO | BOC | Q9BWV1 | 91653 | BOC | 1.008974 | 0.945678 |
| SL002702 | Serine/threonine-protein kinase pim-1 | PIM1 | P11309 | 5292 | PIM1 | -1.277942 | 0.945734 |
| SL005207 | Natural cytotoxicity triggering receptor 1 | NKp46 | O76036 | 9437 | NCR1 | -1.06715 | 0.946091 |
| SL002086 | Ficolin-3 | Ficolin-3 | O75636 | 8547 | FCN3 | -1.043266 | 0.946163 |
| SL008380 | Cathepsin Z | CATZ | Q9UBR2 | 1522 | CTSZ | 1.039662 | 0.946474 |
| SL010457 | Delta-like protein 4 | DLL4 | Q9NR61 | 54567 | DLL4 | 1.011909 | 0.948193 |
| SL011508 | Pituitary adenylate cyclase-activating polypeptide 27 | PACAP-27 | P18509 | 116 | ADCYAP1 | 1.534135 | 0.948755 |
| SL013548 | Interleukin-34 | IL-34 | Q6ZMJ4 | 146433 | IL34 | 1.013058 | 0.949075 |
| SL003190 | C-C motif chemokine 15 | MIP-5 | Q16663 | 6359 | CCL15 | 1.012048 | 0.950475 |
| SL011588 | 40S ribosomal protein S3a | RS3A | P61247 | 6189 | RPS3A | -1.061694 | 0.951122 |
| SL005227 | NKG2D ligand 1 | ULBP-1 | Q9BZM6 | 80329 | ULBP1 | 1.004024 | 0.951377 |
| SL007306 | Protein FAM3B | FAM3B | P58499 | 54097 | FAM3B | -1.018309 | 0.95244 |
| SL004556 | Complement decay-accelerating factor | DAF | P08174 | 1604 | CD55 | 1.016979 | 0.952696 |
| SL005508 | Carbonic anhydrase 9 | Carbonic anhydrase 9 | Q16790 | 768 | CA9 | -1.015259 | 0.952786 |
| SL009054 | Neurexin-1-beta | NRX1B | P58400 | 9378 | NRXN1 | -1.015278 | 0.95292 |
| SL004299 | Baculoviral IAP repeat-containing protein 7 Isoform beta | Livin B | Q96CA5 | 79444 | BIRC7 | -1.0495 | 0.953579 |
| SL004850 | Interleukin-17 receptor A | IL-17 sR | Q96F46 | 23765 | IL17RA | -1.025167 | 0.956181 |
| SL005152 | Retinoic acid receptor responder protein 2 | TIG2 | Q99969 | 5919 | RARRES2 | 1.000927 | 0.957791 |
| SL007221 | Signal transducer and activator of transcription 3 | STAT3 | P40763 | 6774 | STAT3 | -1.052518 | 0.960743 |
| SL005357 | Lithostathine-1-alpha | PSP | P05451 | 5967 | REG1A | -1.091588 | 0.961463 |
| SL004747 | Apolipoprotein M | ApoM | O95445 | 55937 | APOM | -1.029425 | 0.961528 |
| SL004516 | Mannose-binding protein C | MBL | P11226 | 4153 | MBL2 | -1.072312 | 0.961715 |
| SL004760 | Platelet-activating factor acetylhydrolase IB subunit beta | PAFAH beta subunit | P68402 | 5049 | PAFAH1B2 | -1.026921 | 0.965374 |
| SL007070 | Low-density lipoprotein receptor | LDLR | P01130 | 3949 | LDLR | -1.031708 | 0.965481 |
| SL005218 | Sialic acid-binding Ig-like lectin 7 | Siglec-7 | Q9Y286 | 27036 | SIGLEC7 | 1.021555 | 0.965985 |
| SL003738 | T-lymphocyte activation antigen CD80 | B7 | P33681 | 941 | CD80 | -1.006512 | 0.966708 |
| SL004128 | Activin receptor type-1B | Activin RIB | P36896 | 91 | ACVR1B | -1.029742 | 0.968135 |
| SL014684 | 2'-5'-oligoadenylate synthase 1 | OAS1 | P00973 | 4938 | OAS1 | -1.055066 | 0.9688 |
| SL004068 | Granzyme B | Granzyme B | P10144 | 3002 | GZMB | -1.020532 | 0.968804 |
| SL007261 | Methionine aminopeptidase 2 | AMPM2 | P50579 | 10988 | METAP2 | -1.046225 | 0.971528 |
| SL018625 | Toll-like receptor 4:Lymphocyte antigen 96 complex | TLR4:MD-2 complex | O00206 Q9Y6Y9 | 7099 23643 | TLR4 LY96 | -1.017067 | 0.977131 |
| SL006698 | Ligand-dependent nuclear receptor corepressor-like protein | transcription factor MLR1, isoform CRA_b | Q8N3X6 | 254251 | LCORL | -1.030176 | 0.977171 |
| SL005208 | Reticulon-4 receptor | Nogo Receptor | Q9BZR6 | 65078 | RTN4R | -1.002768 | 0.977177 |
| SL004182 | Calcium-dependent phospholipase A2 | GV | P39877 | 5322 | PLA2G5 | -1.00948 | 0.977925 |
| SL018891 | Down syndrome cell adhesion molecule | DSCAM | O60469 | 1826 | DSCAM | -1.031365 | 0.978481 |
| SL003060 | Fibroblast growth factor receptor 1 | bFGF-R | P11362 | 2260 | FGFR1 | -1.0175 | 0.978986 |
| SL000142 | Thymidylate synthase | TS | P04818 | 7298 | TYMS | -1.0271 | 0.981092 |
| SL009216 | Aromatic-L-amino-acid decarboxylase | dopa decarboxylase | P20711 | 1644 | DDC | -1.008702 | 0.981738 |
| SL005219 | Sialic acid-binding Ig-like lectin 9 | Siglec-9 | Q9Y336 | 27180 | SIGLEC9 | -1.075082 | 0.982046 |
| SL007229 | Signal transducer and activator of transcription 6 | STAT6 | P42226 | 6778 | STAT6 | 1.0111 | 0.982396 |
| SL006460 | Platelet glycoprotein Ib alpha chain | GP1BA | P07359 | 2811 | GP1BA | -1.001935 | 0.982451 |
| SL004876 | Kallistatin | Kallistatin | P29622 | 5267 | SERPINA4 | 1.023709 | 0.983208 |
| SL010928 | Inosine-5'-monophosphate dehydrogenase 2 | IMDH2 | P12268 | 3615 | IMPDH2 | -1.025574 | 0.985007 |
| SL000670 | Glutathione S-transferase A3 | GSTA3 | Q16772 | 2940 | GSTA3 | -1.000822 | 0.986137 |
| SL003186 | Lymphotactin | Lymphotactin | P47992 | 6375 | XCL1 | -1.974213 | 0.988167 |
| SL002602 | Trefoil factor 2 | Trefoil factor 2 | Q03403 | 7032 | TFF2 | 1.033892 | 0.988358 |
| SL007295 | Cadherin-15 | CAD15 | P55291 | 1013 | CDH15 | -1.03309 | 0.988777 |
| SL003672 | Extracellular superoxide dismutase [Cu-Zn] | SOD3 | P08294 | 6649 | SOD3 | -1.133554 | 0.989594 |
| SL012188 | Epidermal growth factor receptor substrate 15-like 1 | EP15R | Q9UBC2 | 58513 | EPS15L1 | -1.004999 | 0.990114 |
| SL014048 | Collagen alpha-1(XXIII) chain | CONA1 | Q86Y22 | 91522 | COL23A1 | -1.016089 | 0.990299 |
| SL007871 | UMP-CMP kinase | Cytidylate kinase | P30085 | 51727 | CMPK1 | -1.002604 | 0.990475 |
| SL005204 | NKG2-D type II integral membrane protein | NKG2D | P26718 | 22914 | KLRK1 | -1.007188 | 0.990896 |
| SL006512 | Hepatocyte growth factor activator | HGFA | Q04756 | 3083 | HGFAC | -1.009177 | 0.992193 |
| SL004155 | Platelet-derived growth factor receptor beta | PDGF Rb | P09619 | 5159 | PDGFRB | -1.023892 | 0.993739 |
| SL007502 | Carbohydrate sulfotransferase 15 | ST4S6 | Q7LFX5 | 51363 | CHST15 | 1.005904 | 0.993741 |
| SL009988 | Disintegrin and metalloproteinase domain-containing protein 12 | ADAM12 | O43184 | 8038 | ADAM12 | -1.437047 | 0.994636 |
| SL009213 | Lysosomal protective protein | Cathepsin A | P10619 | 5476 | CTSA | 1.02612 | 0.994821 |
| SL002075 | Interferon alpha-2 | IFN-aA | P01563 | 3440 | IFNA2 | 1.007061 | 0.995464 |
| SL008178 | Dermatopontin | DERM | Q07507 | 1805 | DPT | -1.010696 | 0.996258 |
| SL014070 | SLIT and NTRK-like protein 5 | SLIK5 | O94991 | 26050 | SLITRK5 | 1.017372 | 0.996498 |
| SL000074 | Cellular tumor antigen p53 | p53 | P04637 | 7157 | TP53 | -1.005974 | 0.999728 |
| SL004643 | Angiopoietin-4 | Angiopoietin-4 | Q9Y264 | 51378 | ANGPT4 | -1.006717 | 0.999936 |

**Table S4a.** Spearman correlation coefficients for PREOP.

|  | **HAMP** | **IGFBP2** | **LCN2** | **TIMP1** | **ANGPT2** | **CHI3L1** | **CCL5** | **THBS1** | **IL6** | **NFL** | **PDGFB** | **TNFRSF1A** |
| --- | --- | --- | --- | --- | --- | --- | --- | --- | --- | --- | --- | --- |
| **HAMP** |  |  |  |  |  |  |  |  |  |  |  |  |
| **IGFBP2** | -0.08 |  |  |  |  |  |  |  |  |  |  |  |
| **LCN2** | 0.03 | **0.36** |  |  |  |  |  |  |  |  |  |  |
| **TIMP1** | 0.05 | **0.30** | **0.55** |  |  |  |  |  |  |  |  |  |
| **ANGPT2** | 0.06 | **0.24** | **0.26** | **0.35** |  |  |  |  |  |  |  |  |
| **CHI3L1** | -0.10 | **0.19** | **0.41** | **0.37** | **0.33** |  |  |  |  |  |  |  |
| **CCL5** | 0.13 | -0.02 | 0.04 | **0.33** | -0.10 | -0.13 |  |  |  |  |  |  |
| **THBS1** | 0.17 | -0.08 | 0.11 | **0.31** | -0.12 | -0.08 | **0.91** |  |  |  |  |  |
| **IL6** | 0.12 | **0.34** | **0.29** | **0.56** | **0.28** | **0.27** | 0.16 | 0.12 |  |  |  |  |
| **NFL** | 0.03 | **0.53** | **0.48** | **0.42** | **0.20** | **0.30** | -0.10 | -0.10 | **0.46** |  |  |  |
| **PDGFB** | **0.23** | -0.10 | 0.13 | **0.33** | -0.11 | -0.14 | **0.83** | **0.88** | 0.11 | -0.07 |  |  |
| **TNFRSF1A** | -0.13 | **0.33** | **0.49** | **0.57** | 0.16 | **0.40** | -0.10 | -0.08 | **0.46** | **0.56** | -0.11 |  |

The correlation coefficient higher than 0.9 is marked in red, lower than 0.9 and higher than 0.7 in orange, and lower than 0.7 and higher than 0.5 in yellow. The coefficient with the p-value < 0.05 is in bold text. ANGPT2; angiopoietin-2; CCL5: C-C motif chemokine 5; CHI3L1: chitinase-3-like protein 1; HAMP: hepcidin; IGFBP2: insulin-like growth factor-binding protein 2; IL6: interleukin-6; LCN2: lipocalin-2; NFL: neurofilament light chain; PDGFB: platelet-derived growth factor subunit B; THBS1: thrombospondin-1; TIMP1: metalloproteinase inhibitor 1; TNFRSF1A: tumor necrosis factor receptor superfamily member 1A.

**Table S4b.** Spearman correlation coefficients for POD2.

| 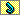 | **HAMP** | **IGFBP2** | **LCN2** | **TIMP1** | **ANGPT2** | **CHI3L1** | **CCL5** | **THBS1** | **IL6** | **NFL** | **PDGFB** | **TNFRSF1A** |
| --- | --- | --- | --- | --- | --- | --- | --- | --- | --- | --- | --- | --- |
| **HAMP** |  |  |  |  |  |  |  |  |  |  |  |  |
| **IGFBP2** | -0.16 |  |  |  |  |  |  |  |  |  |  |  |
| **LCN2** | 0.14 | **0.46** |  |  |  |  |  |  |  |  |  |  |
| **TIMP1** | -0.05 | **0.50** | **0.69** |  |  |  |  |  |  |  |  |  |
| **ANGPT2** | 0.07 | 0.07 | 0.08 | **0.20** |  |  |  |  |  |  |  |  |
| **CHI3L1** | -0.11 | **0.33** | **0.56** | **0.61** | 0.06 |  |  |  |  |  |  |  |
| **CCL5** | 0.10 | **-0.25** | -0.05 | -0.07 | -0.11 | -0.12 |  |  |  |  |  |  |
| **THBS1** | 0.07 | **-0.21** | 0.01 | -0.04 | -0.10 | -0.08 | **0.95** |  |  |  |  |  |
| **IL6** | **0.25** | 0.04 | **0.48** | **0.50** | 0.13 | **0.45** | -0.08 | 0.00 |  |  |  |  |
| **NFL** | -0.08 | **0.56** | **0.44** | **0.46** | 0.01 | **0.22** | **-0.23** | -0.17 | **0.22** |  |  |  |
| **PDGFB** | 0.00 | -0.17 | 0.00 | 0.00 | -0.10 | -0.08 | **0.85** | **0.93** | 0.02 | -0.11 |  |  |
| **TNFRSF1A** | 0.04 | **0.59** | **0.79** | **0.77** | 0.13 | **0.52** | -0.14 | -0.07 | **0.47** | **0.60** | -0.03 |  |

The correlation coefficient higher than 0.9 is marked in red, lower than 0.9 and higher than 0.7 in orange, and lower than 0.7 and higher than 0.5 in yellow. The coefficient with the p-value < 0.05 is in bold text. ANGPT2; angiopoietin-2; CCL5: C-C motif chemokine 5; CHI3L1: chitinase-3-like protein 1; HAMP: hepcidin; IGFBP2: insulin-like growth factor-binding protein 2; IL6: interleukin-6; LCN2: lipocalin-2; NFL: neurofilament light chain; PDGFB: platelet-derived growth factor subunit B; THBS1: thrombospondin-1; TIMP1: metalloproteinase inhibitor 1; TNFRSF1A: tumor necrosis factor receptor superfamily member 1A.
